# Ethnic and region-specific genetic risk variants of stroke and its comorbid conditions can define the variations in the burden of stroke and its phenotypic traits

**DOI:** 10.1101/2023.12.05.23299453

**Authors:** Rashmi Sukumaran, Achuthsankar S. Nair, Moinak Banerjee

**Author notes:** Corresponding author: Dr. Moinak Banerjee, Laboratory of Human Molecular Genetics Rajiv Gandhi Centre of Biotechnology (RGCB), Thycaud Post, Poojappura, Thiruvananthapuram, Kerala 695014. India.

## Abstract

Burden of stroke differs by region, which could be attributed to differences in comorbid conditions and ethnicity. Genomewide variation acts as a proxy marker for ethnicity, and comorbid conditions. We present an integrated approach to understand this variation by considering prevalence and mortality rates of stroke and its comorbid risk for 204 countries from 2009 to 2019, and GWAS risk variant for all these conditions. Global and regional trend analysis of rates using linear regression, correlation and proportion analysis, signify ethnogeographic differences. Interestingly, the comorbid conditions that act as risk drivers for stroke differed by regions, with more of metabolic risk in America and Europe, in contrast to high SBP in Asian and African regions. GWAS risk loci of stroke and its comorbid conditions indicate distinct population stratification for each of these conditions, signifying for population specific risk. Unique and shared genetic risk variants for stroke, and its comorbid and followed up with ethnic specific variation can help in determining regional risk drivers for stroke. Unique ethnic specific risk variants and their distinct patterns of Linkage Disequilibrium further uncover the drivers for phenotypic variation. Therefore, identifying population and comorbidity specific risk variants might help in defining the threshold for risk, and aid in developing population specific prevention strategies for stroke.

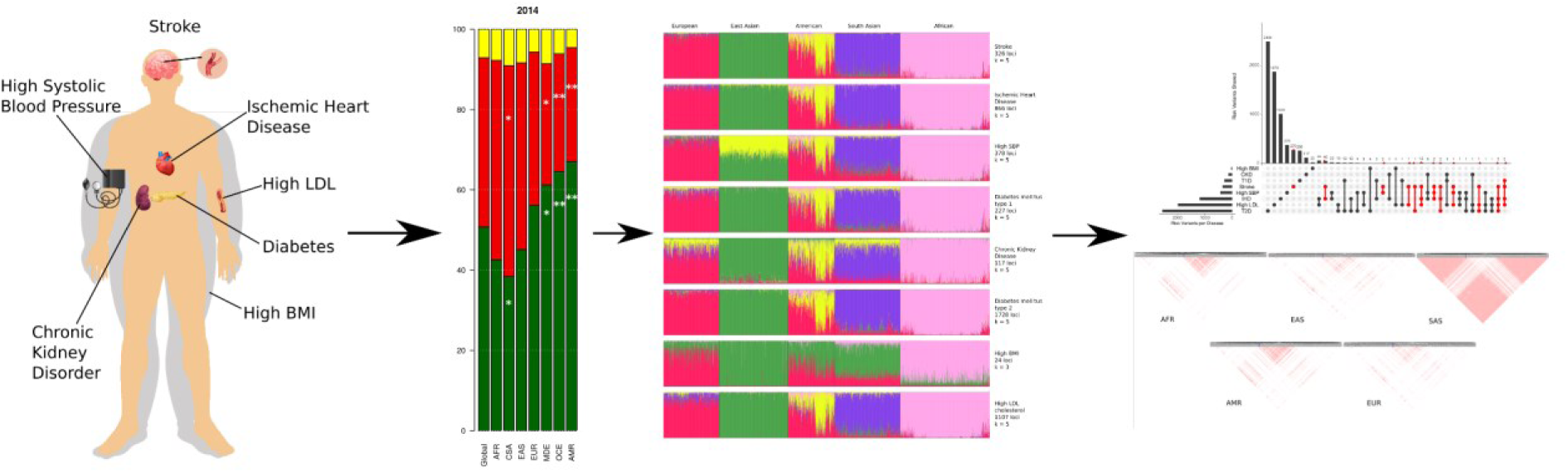
Graphical Abstract.

## INTRODUCTION

Stroke affects over 101 million people worldwide and is ranked the second most fatal disease in the world, with 6.5 million deaths in 2019^1^. Comorbid conditions of stroke are critical contributors to burden of stroke and the duration of the comorbid conditions can further determine the severity of stroke risk or mortality. Prevalence for comorbid conditions range from 43% to 94% and estimates can go as high as 99% above 66 years of age^2^. Prevalence and mortality risk in stroke has often been evaluated from socio-economic viewpoint, but it is also critical to understand the differences in drivers such as comorbid conditions. It is the accumulated risk of comorbid conditions that enhances the risk of stroke further. Are these comorbid conditions differentially impacted by socio-economic factors and ethnogeographic factors. This was clearly evident in COVID era, when COVID-19 differentially impacted the risk of stroke, possibly due to its differential influence on the comorbidities of stroke.

Mortality in Stroke, its subtypes and their comorbid conditions have a strong ethnic bias^3,4,5^. Genetics act as surrogate markers for ethnogeographic indices. It is important to understand which comorbid conditions are influenced by socio-economic indices, and how they impact the risk of stroke and their underlying genetic basis. A Danish study reported the effect sizes of association with comorbid conditions for stroke to have 15% higher mortality risk in presence of diabetes mellitus with end-organ damage, 20% for peripheral vascular disease, 25% for chronic pulmonary disease, 35% for congestive heart failure and atrial flutter, 45% for moderate to severe renal disease, and 1.8– to 2.4-fold for mild to severe liver disease^6^. A UK Biobank study on stroke multimorbidity reported 1.5x higher risk of mortality in those with 2 additional comorbidities and a ≈2.5× higher risk of mortality in those with ≥5 comorbidities over seven years^7^. Thus, the differential impact of comorbid or multimorbid conditions contributing to the additive effect of illness burden needs to be addressed from an ethnogenetic perspective. Devising an appropriate strategy for prevention of stroke burden, needs a careful evaluation of the underlying genetic signature for each of these comorbid conditions and distinguishing their ethnic bias.

The objective of the study was to understand what determines the differences in stroke burden around the globe. Variations in burden of stroke could be influenced by comorbid conditions, and incidentally both stroke and its comorbid conditions can be influenced by ethnogeographic factors and genetics can act as a stable proxy marker for all. To resolve this, we considered the prevalence and mortality of a total of eleven disease conditions, consisting of stroke and its comorbid conditions, across different continents and ethnicities from 2009 to 2019. The disease conditions were further stratified as per their ethnogeographic locations and their genetic risk variants extrapolated from GWAS data. This study would provide insights on the regional patterns of the burden of stroke and its comorbid conditions, and help in resolving it from an ethnic and genetic viewpoint. These insights would further aid in developing and strategizing regional and ethnic specific needs for prevention of the risk of comorbid conditions and stroke.

## RESULTS

### Global mortality, incidence and prevalence rates of stroke and its comorbid conditions

Globally, stroke ranks as the second most fatal disease in 2019 (84.2/100000, 95%UI 76.8-90.2) among the eight diseases analyzed, preceded by ischemic heart disease (IHD; 117.9/100000, 95%UI 107.8-125.9) as shown in Figure 1-figure supplement 1 and Table S1. High systolic blood pressure (high SBP) ranks as the most fatal comorbid disease condition with an age-standardized mortality rate (ASMR) of 138.9/100000 [95%UI 121.3-155.7] among all conditions. Within stroke subtypes, ischemic stroke ranks highest globally with ASMR of 43.5/100000 [95%UI 39.08-46.8], followed by intracerebral hemorrhage (ICH) at 36.0/100000 [95%UI 32.9-38.7] and subarachnoid hemorrhage (SAH) at 4.7/100000 [95%UI 4.1-5.2]. The ranking of the ASMRs of the diseases follow the same trend throughout the last decade with minor exceptions, where high body-mass index (high BMI) improved its rank in 2014 by swapping with high low-density lipoprotein cholesterol (high LDL), and type 1 diabetes (T1D) dethroned chronic kidney disease (CKD) in 2019.

**Figure 1.**
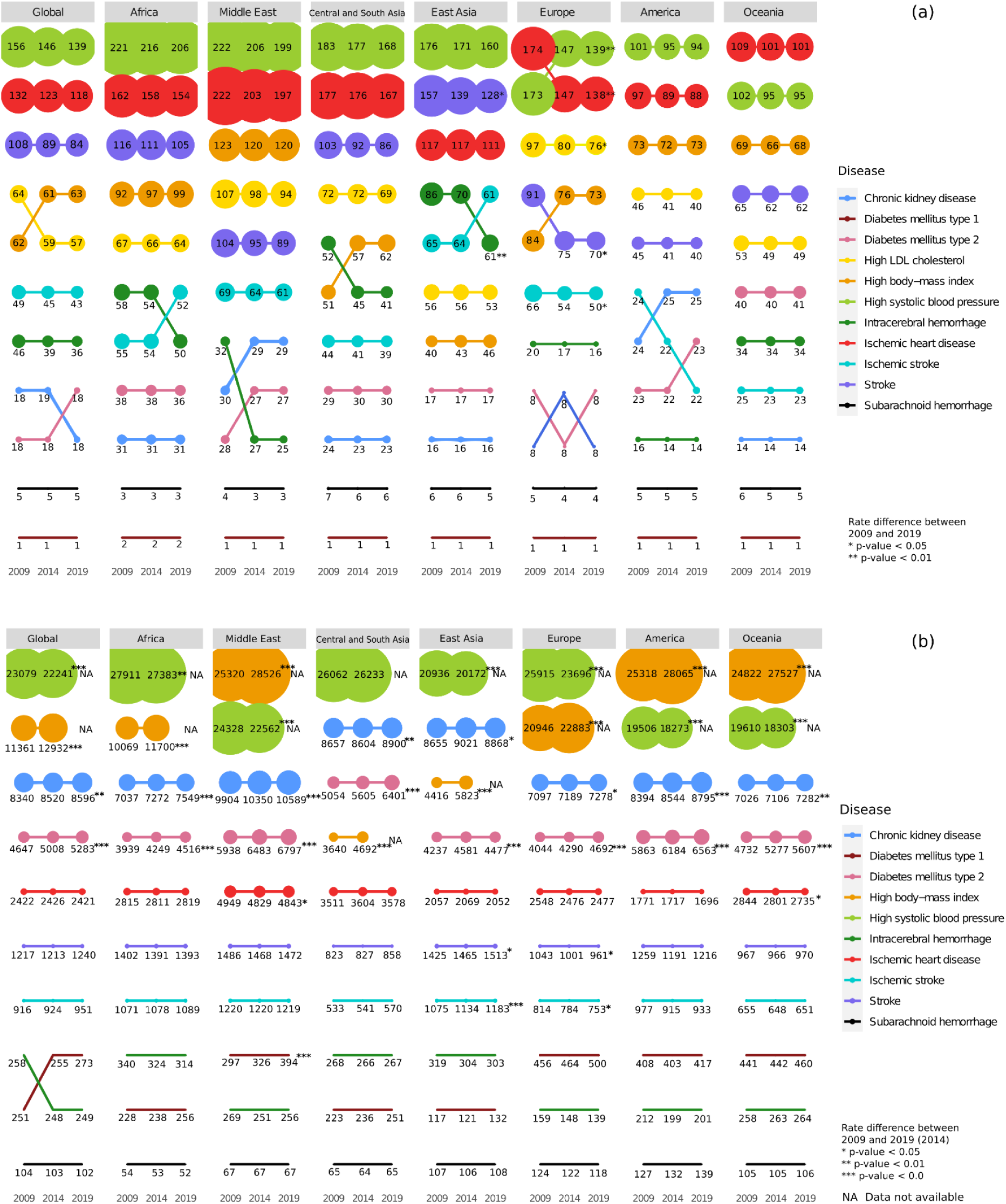
Regional (a) mortality rates and (b) prevalence rates for Stroke, its subtypes and comorbid factors. The figure shows the age-standardized mortality and prevalence rates per 100,000 people for Stroke, its subtypes and its comorbid factors in 2009, 2014 and 2019 in the geographical regions. The size of the points indicates the rate and position indicates rank.

Global trends in crude and age-standardized incidence rates (ASIRs) show that stroke incidence ranks fourth, while IHD is on top followed by T2D and CKD (Figure 1-figure supplement 1, Table S1). Crude incidence rates of stroke and its subtypes increased in the last decade but ASIRs decreased, with exception of ischemic stroke, where an increase in ASIR was observed. Among other diseases, IHD, type 2 diabetes (T2D) and T1D show a continuous increase in the last decade, with T2D (280.1/100000, 95%UI 258.8-303.9) surpassing IHD (274.0/100000, 95%UI 242.9-306.4) in 2019 in crude rates. The global trend between crude and age-standardized prevalence rates (ASPRs) revealed that the ranking of stroke remains at sixth position throughout the years (Figure 1-figure supplement 1, Table S1). Among stroke subtypes, ischemic stroke ranks highest. The comorbid conditions, high SBP and high BMI, ranks first and second followed by CKD and T2D, and interestingly all rank above stroke. We also observe that though there is a continuous increase in prevalence rates in the last decade, the ranking of stroke or its comorbid conditions does not change over the years, with the exception of T1D ASPRs overtaking ICH. We were keen to understand if these global trends of ASMR and ASPR are influenced by region and ethnicity.

### Ethno-regional differences in mortality and prevalence of stroke and its major comorbid conditions

We observed interesting patterns of ASMRs of stroke, its subtypes and its major comorbidities across different regions over the years as shown in Figure 1a and Figure 1-figure supplement 2, Table 1 and S2. When assessed in terms of ranks, high SBP is the most fatal condition followed by IHD in all regions, except Oceania where IHD and high SBP swap ranks. Africa (206.2/100000, 95%UI 177.4-234.2) and Middle East (198.6/100000, 95%UI 162.8-234.4) have the highest ASMR for high SBP, even though they rank as only the third and sixth most populous continents (Appendix Figure 1), respectively. Both high SBP (−0.64% to –2.25% EAPC) and IHD (−0.45% to –1.17% EAPC) show a decreasing trend for ASMRs in all regions. However, only Europe shows a significant decrease for high SBP (−2.26%; P=0.009) and IHD (−2.37%; P=0.006) in the decade. Stroke has a decreasing trend for ASMRs with East Asia and Europe showing a significant decrease of –2.2% (P=0.021) and –2.6% (P=0.03), respectively. Stroke has the highest mortality in East Asia (127.1/100000, 95%UI 104.9-150.5 in 2019), and is the only region that ranks stroke higher than IHD. Though Europe, Middle East and Central & South Asia have ASMRs similar to global rates for stroke, Central & South Asia ranks stroke as the third most fatal factor, while America, Europe and Middle East ranks it fifth. Oceania (62.1/100000, 95%UI 34.1-90.2) and America (40.3/100000, 95%UI 36.2-43.1) have lowest rates for stroke in 2019.

**Table 1.** EAPC from 2009 to 2019 of age-standardized mortality rates of Stroke and its comorbid conditions. ASMR per 100,000 for Stroke and its comorbid conditions for 2009 and 2019, as well as EAPC from 2009 to 2019 is show. 95% uncertainty interval is shown in paranthesis, statistically significant intervals are highlighted in bold.

|  | Chronic Kidney disease |  |  | High BMI |  |  |
| --- | --- | --- | --- | --- | --- | --- |
| Region | 2019 | 2009 | 2009-2019 EAPC | 2019 | 2009 | 2009-2019 EAPC |
| Global | 18.29<br>(19.6, 16.72) | 18.41<br>(19.22, 17.1) | -0.07<br>(-4.32, 4.37) | 62.59<br>(89.13, 39.92) | 61.66<br>(90.2, 37.44) | 0.16<br>(-2.2, 2.57) |
| Africa | 30.58<br>(27.11, 34.05) | 31.18<br>(28.37, 33.99) | -0.12<br>(-3.41, 3.27) | 98.83<br>(64.77, 136.6) | 91.64<br>(58.6, 130.04) | 0.88<br>(-1.02, 2.82) |
| Central & South Asia | 22.74<br>(17.68, 27.81) | 23.72<br>(18.93, 28.5) | 0.24<br>(-4.07, 3.73) | 62.07<br>(40.97, 83.17) | 51.31<br>(30.2, 72.47) | 2.0<br>(-0.5, 4.56) |
| East Asia | 15.8<br>(11.17, 20.42) | 16.22<br>(12.1, 20.34) | -0.38<br>(-4.9, 4.36) | 46.41<br>(36.68, 56.14) | 39.76<br>(32.9, 46.61) | 1.45<br>(-1.39, 4.38) |
| Europe | 8.18<br>(7.06, 9.3) | 8.19<br>(7.1, 9.29) | 0.11<br>(-6.17, 6.81) | 73.34<br>(60.31, 86.36) | 84.45<br>(69.2, 99.71) | -1.42<br>(-3.49, 0.7) |
| Middle East | 28.68<br>(22.76, 34.59) | 29.75<br>(22.4, 37.14) | -0.46<br>(-3.85, 3.06) | 120.14<br>(100.3, 139.9) | 122.6<br>(98.3, 146.9) | -0.23<br>(-1.91, 1.48) |
| Oceania | 13.71<br>(9.67, 17.74) | 13.6<br>(9.16, 18.04) | 0.17<br>(-4.76, 5.36) | 67.52<br>(41.1, 93.93) | 69.2<br>(42.2, 96.19) | -0.28<br>(-2.53, 2.02) |
| America | 24.95<br>(22.85, 26.81) | 23.83<br>(22.22, 24.68) | 0.48<br>(-3.24, 4.34) | 72.83<br>(48.62, 97.9) | 73.34<br>(49.02, 99.92) | -0.01<br>(-2.17, 2.23) |
|  | High LDL |  |  | High SBP |  |  |
| Region | 2019 | 2009 | 2009-2019 EAPC | 2019 | 2009 | 2009-2019 EAPC |
| Global | 56.51<br>(73.62, 41.83) | 63.9<br>(82.9, 47.71) | -1.29<br>(-3.65, 1.13) | 138.88<br>(155.7, 121.3) | 156.22<br>(174.7, 137.3) | -1.23<br>(-2.74, 0.31) |
| Africa | 64.39<br>(45.03, 86.13) | 67.15<br>(48.42, 88.17) | -0.34<br>(-2.6, 1.98) | 206.18<br>(177.4, 234.2) | 221.26<br>(196.2, 247.2) | -0.64<br>(-1.9, 0.63) |
| Central & South Asia | 68.91<br>(45.31, 92.51) | 72.3<br>(45.6, 98.99) | -0.46<br>(-2.64, 1.77) | 167.69<br>(129.1, 206.3) | 183.48<br>(140.3, 226.6) | -0.84<br>(-2.23, 0.57) |
| East Asia | 53.44<br>(46.16, 60.71) | 56.11<br>(48.82, 63.4) | -0.68<br>(-3.15, 1.82) | 160.36<br>(130.6, 190.1) | 176.4<br>(146.7, 206.1) | -1.1<br>(-2.51, 0.33) |
| Europe | 75.53<br>(57.71, 93.34) | 96.66<br>(74.4, 119.0) | **-2.53**<br>**(-4.51, -0.51)** | 138.88<br>(111.6, 166.2) | 173.48<br>(139.1, 207.9) | **-2.26**<br>**(-3.73, -0.76)** |
| Middle East | 94.43<br>(75.03, 113.8) | 107.43<br>(88.6, 126.3) | -1.3<br>(-3.14, 0.57) | 198.61<br>(162.8, 234.4) | 222.34<br>(187.2, 257.5) | -1.16<br>(-2.43, 0.13) |
| Oceania | 48.92<br>(33.56, 64.27) | 53.34<br>(39.4, 67.27) | -0.92<br>(-3.51, 1.74) | 94.63<br>(62.2, 127.06) | 102.14<br>(70.5, 133.76) | -0.81<br>(-2.68, 1.09) |
| America | 40.44<br>(30.0, 52.53) | 45.86<br>(34.22, 59.38) | -1.24<br>(-4.05, 1.64) | 93.91<br>(80.49, 105.8) | 101.2<br>(87.28, 114.1) | -0.7<br>(-2.57, 1.21) |
|  | Ischemic heart disease |  |  | Intracerebral hemorrhage |  |  |
| Region | 2019 | 2009 | 2009-2019 EAPC | 2019 | 2009 | 2009-2019 EAPC |
| Global | 117.95<br>(125.9, 107.8) | 131.77<br>(138.3, 122.4) | -1.17<br>(-2.81, 0.5) | 36.04<br>(38.67, 32.98) | 45.92<br>(48.45, 43.07) | -2.53<br>(-5.37, 0.39) |
| Africa | 154.04<br>(132.6, 175.4) | 162.34<br>(146.4, 180.7) | -0.45<br>(-1.91, 1.04) | 49.82<br>(43.07, 57.02) | 58.06<br>(51.19, 65.11) | -1.45<br>(-3.93, 1.09) |
| Central & South Asia | 167.39<br>(115.8, 219.0) | 176.75<br>(118.3, 235.2) | -0.51<br>(-1.91, 0.92) | 41.45<br>(32.34, 50.55) | 51.57<br>(37.19, 65.94) | -2.13<br>(-4.8, 0.62) |
| East Asia | 110.93 | 117.17 | -0.76 | 61.2 | 85.51 | **-3.54** |
|  | (94.7,127.17) | (100.7,133.7) | (-2.47, 0.97) | (49.06,73.33) | (70.7,100.37) | **(-5.65, -1.38)** |
| Europe | 138.19<br>(105.5,170.8) | 174.31<br>(134.8,213.8) | **-2.37**<br>**(-3.85, -0.88)** | 15.7<br>(12.25,19.15) | 20.31<br>(15.85,24.77) | -2.46<br>(-6.77, 2.04) |
| Middle East | 197.4<br>(154.9,239.9) | 221.61<br>(178.7,264.5) | -1.14<br>(-2.41, 0.16) | 24.94<br>(15.45,34.42) | 31.66<br>(19.43,43.89) | -2.445<br>(-5.8, 1.03) |
| Oceania | 100.75<br>(65.96,135.6) | 109.14<br>(77.5,140.7) | -0.86<br>(-2.67, 0.99) | 34.32<br>(12.57,56.07) | 34.25<br>(12.46,56.04) | -0.056<br>(-3.2, 3.19) |
| America | 87.51<br>(79.07,93.31) | 97.5<br>(89.02,102.0) | -1.06<br>(-2.98, 0.91) | 14.06<br>(12.97,15.03) | 15.95<br>(15.05,16.68) | -1.21<br>(-5.93, 3.74) |
|  | **Ischemic stroke** |  |  | **Subarachnoid hemorrhage** |  |  |
| **Region** | **2019** | **2009** | **2009-2019**<br>**EAPC** | **2019** | **2009** | **2009-2019**<br>**EAPC** |
| Global | 43.5<br>(46.8, 39.08) | 49.24<br>(52.21,44.75) | -1.3<br>(-3.99, 1.47) | 4.66<br>(5.17, 4.13) | 5.34<br>(6.2, 4.57) | -1.29<br>(-9.21, 7.33) |
| Africa | 52.05<br>(45.89,58.59) | 54.74<br>(48.9, 60.76) | -0.43<br>(-2.94, 2.14) | 2.73<br>(1.62, 4.97) | 3.27<br>(1.9, 5.95) | -1.71<br>(-11.75, 9.48) |
| Central & South Asia | 39.2<br>(28.7, 49.75) | 44.32<br>(31.11,57.53) | -0.99<br>(-3.88, 1.95) | 5.77<br>(4.42, 7.12) | 7.15<br>(5.05, 9.26) | -2.16<br>(-9.13, 5.34) |
| East Asia | 61.33<br>(50.37,72.28) | 64.96<br>(54.8, 75.11) | -0.734<br>(-3.04, 1.61) | 5.14<br>(4.66, 5.62) | 6.05<br>(5.45, 6.65) | -1.66<br>(-9.11, 6.4) |
| Europe | 50.36<br>(38.22,62.51) | 65.67<br>(49.49,81.85) | **-2.72**<br>**(-5.14, -0.25)** | 4.16<br>(3.55, 4.78) | 4.84<br>(4.08, 5.6) | -1.3<br>(-9.74, 7.93) |
| Middle East | 61.43<br>(49.71,73.15) | 68.72<br>(57.62,79.81) | -1.17<br>(-3.44, 1.15) | 2.91<br>(2.0, 3.83) | 3.74<br>(2.82, 4.67) | -2.39<br>(-11.73,8.14) |
| Oceania | 22.56<br>(17.61,27.52) | 24.92<br>(20.87,28.98) | -1.04<br>(-4.81, 2.89) | 5.27<br>(3.32, 7.21) | 5.61<br>(3.57, 7.64) | -0.64<br>(-8.31, 7.68) |
| America | 21.69<br>(18.8, 23.4) | 23.96<br>(21.31,25.49) | -0.99<br>(-4.83, 2.99) | 4.57<br>(4.16, 4.9) | 4.81<br>(4.47, 5) | -0.45<br>(-8.72, 8.55) |
|  | **Stroke** |  |  | **Type 2 diabetes** |  |  |
| **Region** | **2019** | **2009** | **2009-2019**<br>**EAPC** | **2019** | **2009** | **2009-2019**<br>**EAPC** |
| Global | 84.19<br>(90.15,76.76) | 100.5<br>(105.4,93.24) | -1.85<br>(-3.76, 0.1) | 18.49<br>(19.66,17.18) | 18.23<br>(18.97,17.12) | 0.21<br>(-4.07, 4.69) |
| Africa | 104.6<br>(93.07,116.9) | 116.07<br>(104.7,127.9) | -0.97<br>(-2.71, 0.81) | 36.45<br>(33.0, 40.01) | 37.5<br>(34.54, 40.3) | -0.24<br>(-3.24, 2.85) |
| Central & South Asia | 86.42<br>(67.06,105.8) | 103.04<br>(75.1,130.94) | -1.63<br>(-3.53, 0.3) | 29.61<br>(24.29,34.93) | 28.54<br>(23.45,33.63) | 0.62<br>(-2.83, 4.17) |
| East Asia | 127.66<br>(104.9,150.5) | 156.52<br>(132.0,181.0) | **-2.2**<br>**(-3.73, -0.64)** | 17.45<br>(9.53, 25.37) | 16.52<br>(9.67, 23.37) | 0.59<br>(-3.85, 5.24) |
| Europe | 70.22<br>(54.79,85.65) | 90.82<br>(70.3,111.34) | **-2.6**<br>**(-4.65, -0.5)** | 8.26<br>(7.03, 9.48) | 8.46<br>(6.95, 9.96) | 0.09<br>(-6.19, 6.78) |
| Middle East | 89.28<br>(70.7,107.87) | 104.12<br>(84.6,123.63) | -1.59<br>(-3.44, 0.3) | 27.43<br>(19.25, 35.6) | 28.38<br>(18.53,38.22) | -0.38<br>(-3.86, 3.22) |
| Oceania | 62.15<br>(34.12,90.18) | 64.78<br>(37.63,91.92) | -0.47<br>(-2.79, 1.9) | 41.29<br>(15.02,67.55) | 40.0<br>(14.19,65.81) | 0.27<br>(-2.65, 3.27) |
| America | 40.33<br>(36.24, 43.1) | 44.72<br>(40.98,46.98) | -1.01<br>(-3.83, 1.9) | 22.55<br>(20.65,24.08) | 22.89<br>(21.35,23.69) | -0.08<br>(-3.96, 3.97) |
|  | **Type 1 diabetes** |  |  |  |  |  |
| **Region** | **2019** | **2009** | **2009-2019**<br>**EAPC** | - | - | - |
| Global | 0.98<br>(1.17, 0.85) | 1.03<br>(1.22, 0.88) | -0.45<br>(-17.5,20.07) | - | - | - |
| Africa | 1.59<br>(1.24, 1.92) | 1.69<br>(1.29, 2.14) | -0.57<br>(-14.01, 14.9) | - | - | - |
| Central & South Asia | 1.34<br>(1.09, 1.6) | 1.4<br>(1.09, 1.71) | -0.33<br>(-14.93, 16.8) | - | - | - |
| East Asia | 0.91<br>(0.33, 1.49) | 0.98<br>(0.39, 1.56) | -1.17<br>(-18.6, 20.01) | - | - | - |
| Europe | 0.51<br>(0.42, 0.61) | 0.59<br>(0.49, 0.69) | -1.57<br>(-24.09, 27.6) | - | - | - |
| Middle East | 1.05<br>(0.8, 1.3) | 1.27<br>(0.97, 1.57) | -2.26<br>(-17.89, 16.3) | - | - | - |
| Oceania | 1.06<br>(0.52, 1.6) | 1.06<br>(0.53, 1.59) | -0.26<br>(-16.9, 19.74) | - | - | - |
| America | 1.04<br>(0.8, 1.29) | 1.1<br>(0.87, 1.37) | -0.27<br>(-16.89, 19.7) | - | - | - |

Among the stroke subtypes, ICH and ischemic stroke show maximum ethnogenetic differences in mortality rates (14.1/100000 to 61.4/100000) and ranking (4th to 9th) in 2019. While ICH shows a significant decrease in ASMR in East Asia (−3.53%, P=0.009), ischemic stroke shows a significant decrease in Europe (−2.37%, P=0.06). High BMI and high LDL rank in the top five but their mortality rates differed across all regions, with the highest rates for both in the Middle East. Only Europe shows a significant decrease in high LDL (−2.53%, P=0.03) over the decade. The Middle East has the highest ASMRs due to IHD and high SBP, followed by Africa, Central & South Asia, East Asia and Europe, all having rates higher than global. All continents have similar mortality rates for T2D and CKD across the years, except Oceania, where the T2D rate is nearly three times CKD rate. Africa has the highest mortality rate for T1D (1.59/100000, 95%UI 1.2-1.9).

ASPRs also showed an interesting pattern of distribution, and, in contrast to mortality, showed an increase over the decade (Figure 1b and Figure 1-figure supplement 3, Table 2 and S3). Highest ASPRs were observed for high SBP across all regions, except America, Middle East and Oceania, where high BMI has most prevalence. While EAPC of prevalence of high SBP showed significant decrease in all regions (−0.38% to –1.77%), except Central & South Asia, high BMI showed a significant increase (1.77% to 5.6%) in all (Table 2). The ASPR ranking of CKD and T2D rose to top five, in sharp contrast to their ASMR rankings. Prevalence of CKD (EAPC 0.24% to 0.7%) and T2D (EAPC 0.6% to 2.18%) is significantly increasing in all regions. For all other diseases, the pattern of ranking and rates across regions were stable with minor exceptions. Stroke ranks sixth for ASPRs in all regions, and it is interesting to note that ASPRs of all the comorbid conditions, except T1D, rank above stroke. While Europe shows a significant decline in ASPRs of stroke (−0.9%, P=0.008) and ischemic stroke (−0.85%, P=0.03) across the years, East Asia shows a significant increase for stroke (0.7%, P=0.02) and ischemic stroke (1.09%, P=<0.001). Globally, T1D swapped its prevalence ranking with ICH in 2014, largely influenced by the significant increase in prevalence in Middle East (2.83%, P=<0.001). However, the highest prevalence of T1D is in Europe and Oceania. The prevalence of IHD has remained nearly constant in all continents in the last decade, except Oceania (−0.43%), America (−0.49%) and Middle East (−0.27%) which shows a significant decrease. In 2019, Middle East has the highest prevalence for IHD (4843.02/100000, 95%UI 4243.02-5442.58), while America (1695.6/100000, 95%UI 1530.8-1871.9) has the lowest ASPR, less than half the rate of the Middle East. We were further keen to understand if these regional differences in mortality and prevalence rates also reflect a socio-economic bias and if so, does it reflect in a category of comorbid conditions.

**Table 2.**
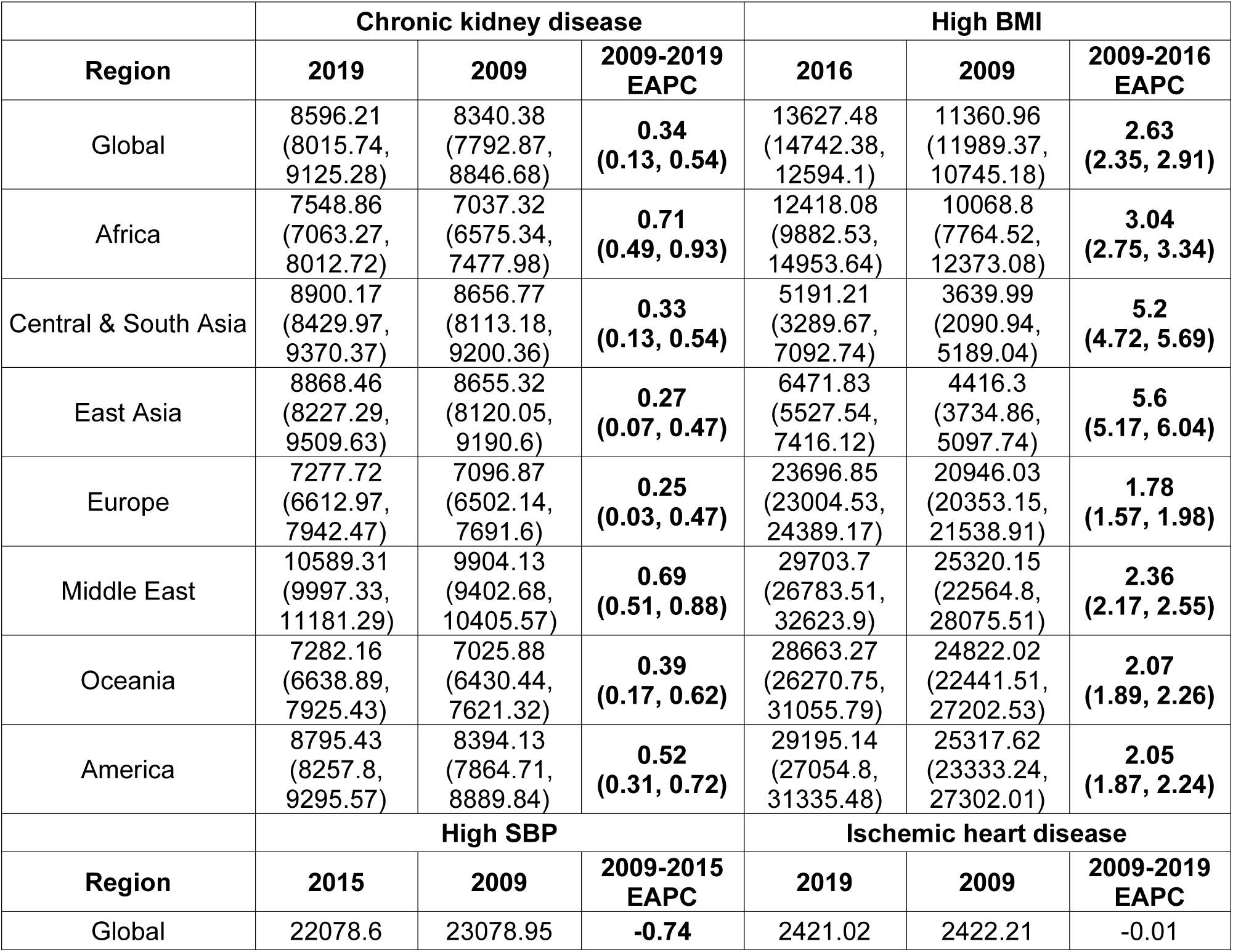
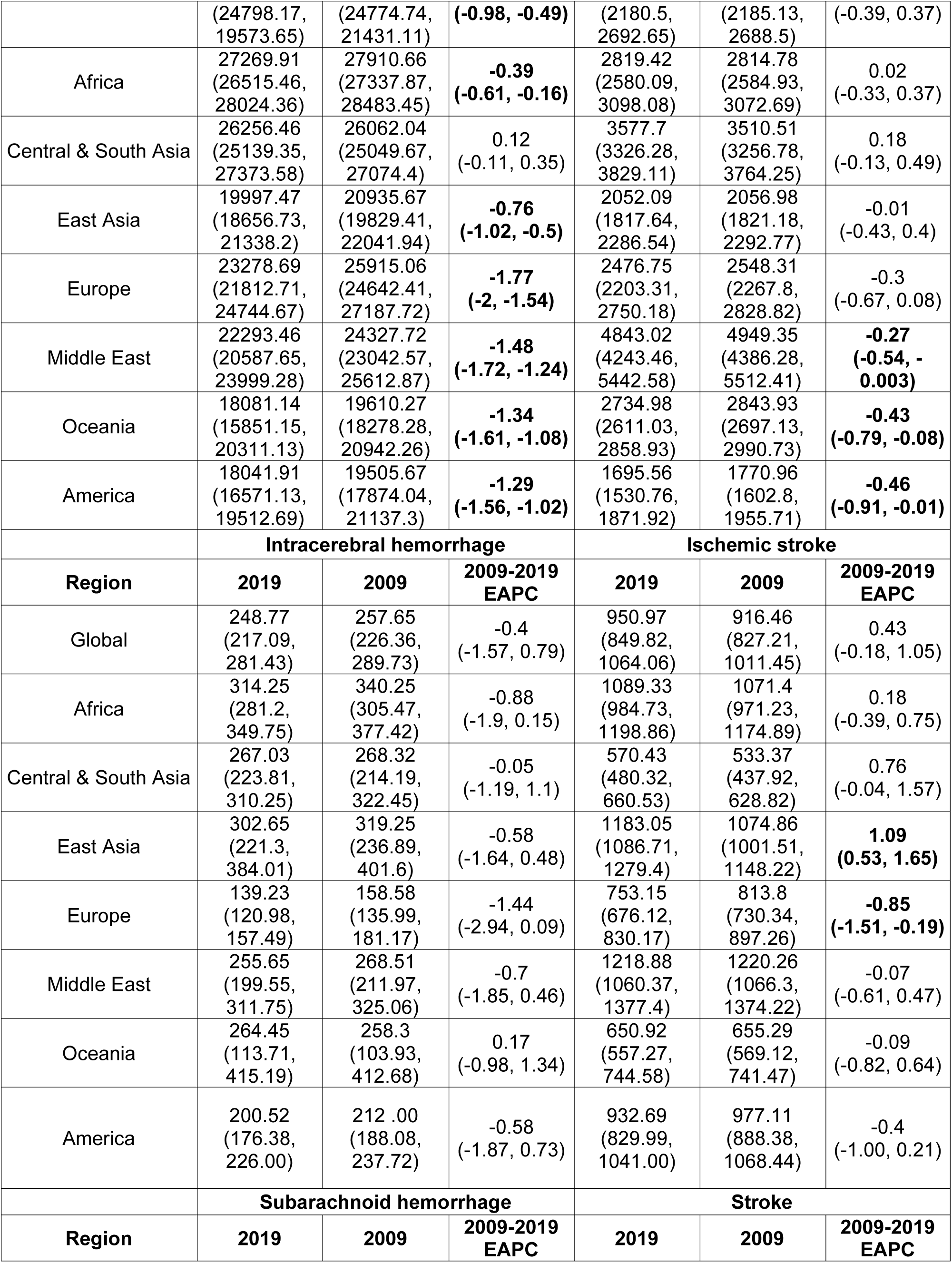
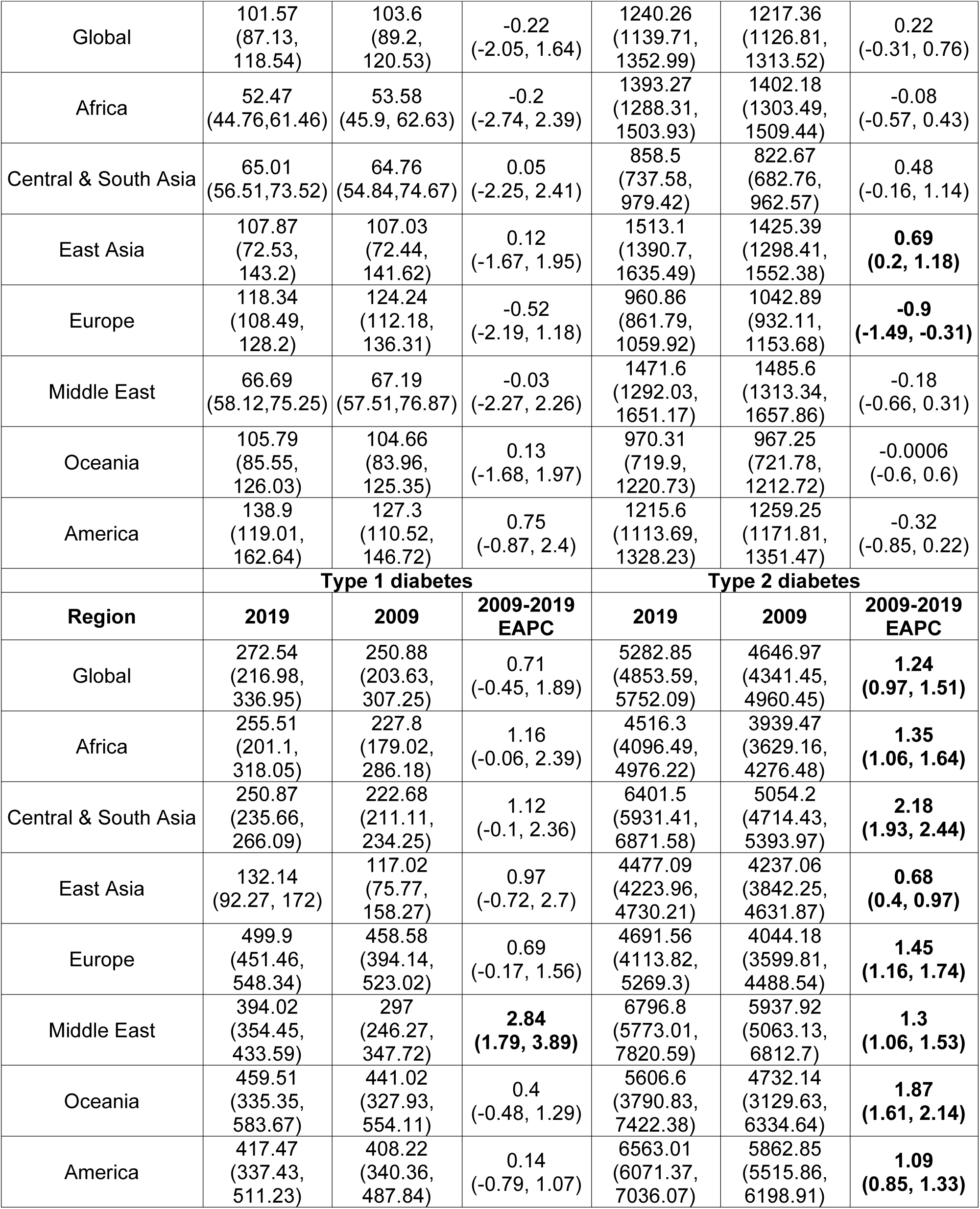
EAPC from 2009 to 2019 of age-standardized prevalence rates of Stroke and its comorbid conditions. ASPR per 100,000 for Stroke and its comorbid conditions for 2009 and 2019, as well as EAPC from 2009 to 2019 is show. 95% uncertainty interval is shown in paranthesis, statistically significant intervals are highlighted in bold.

### Contribution of metabolic risk and hypertension in stroke based on ethnogenetic locations

When the prevalence and mortality rates of stroke and its comorbidities were grouped into three groups, namely strokes, metabolic disorders and high SBP, we find that out of the three proportional mortalities shown in figure 2, strokes group has the highest proportion (37.1%-47.2%) across all years and regions, except Oceania and America, where instead, metabolic disorders have the highest proportion (39.1%-42.2%) that is significantly higher compared to global proportion (Table S4). East Asia has the highest proportional mortality for strokes among all regions in all three years (44.7%-47.2%). This was in sharp contrast to the prevalence proportion of strokes (4.6%-9.1%), which was the least among the three groups, with the highest proportional prevalence for strokes being 8.9%-9.1% in Central & South Asia. The proportional mortality for high SBP (22.0%-30.5%) is very similar across the regions. Metabolic disorders have significantly higher proportional prevalence compared to global in Middle East, Oceania and America, the highest being in America (63.9%, Table S5). Asian and African regions have lowest proportional prevalence for metabolic disorders, with Central & South Asia having significantly lower proportions. However, these regions have the highest prevalence proportion for high SBP (46.6% – 54.3%), with Central & South Asia having a significantly higher proportion compared to global. On the other hand, the Middle East, Oceania and America have significantly lower proportional prevalence compared to global. We were further keen to understand the correlation among ASMR and ASPR of comorbid conditions among ethnogeographic regions.

**Figure 2.**
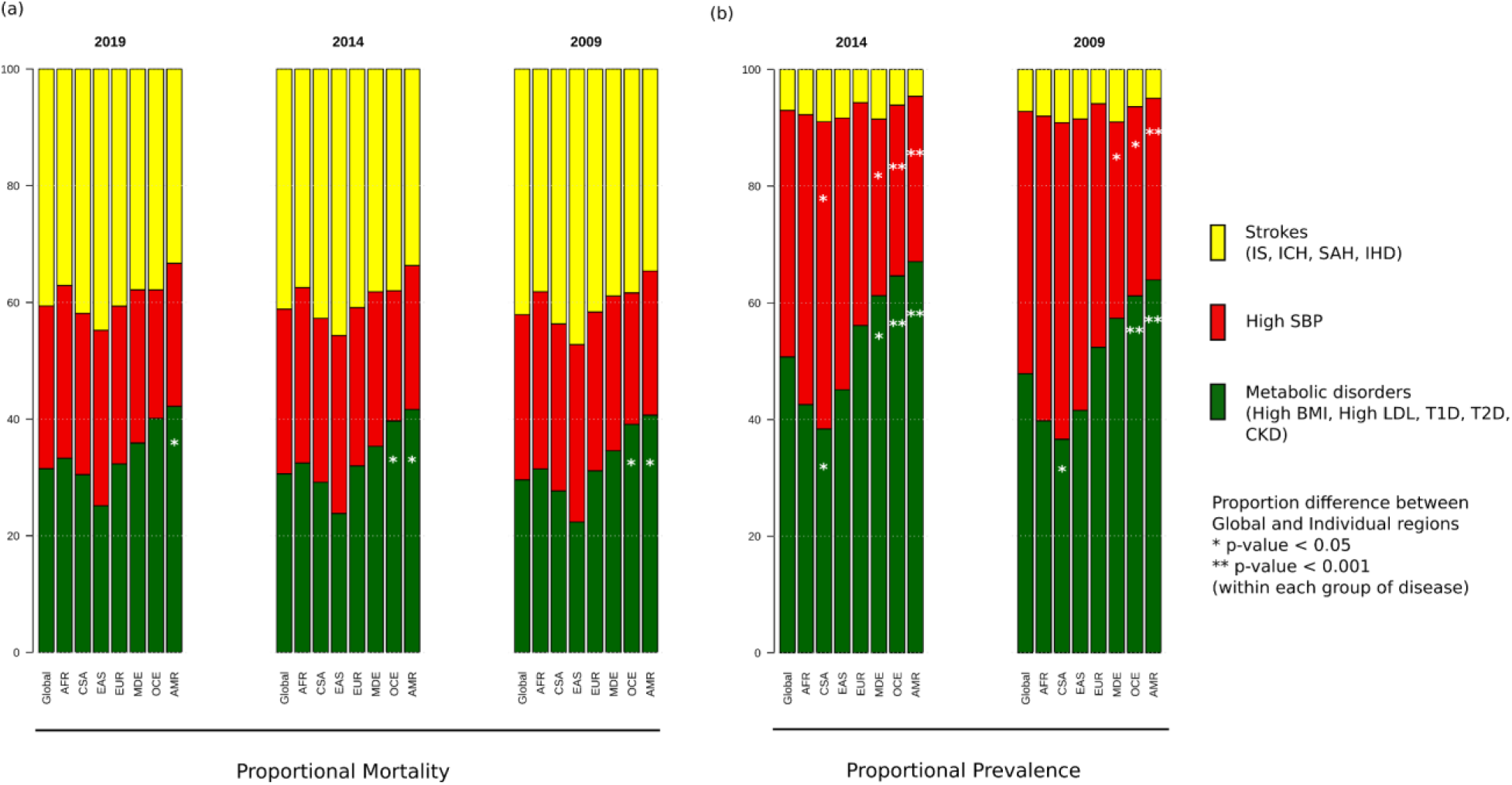
Proportional (a) mortality rates and (b) prevalence rates for Strokes, High SBP and Metabolic disorders. The figure shows the proportional mortality and prevalence for all strokes in yellow (ischemic stroke, intracerebral hemorrhage, subarachnoid hemorrhage and ischemic heart disease), high systolic blood pressure in red, and metabolic disorders in green (high BMI, high LDL, diabetes mellitus type 1 and 2, chronic kidney disorder). CSA – Central and South Asia, AFR – Africa, EAS – East Asia, EUR – Europe, MDE – Middle East, OCE – Oceania, AMR – America.

### Correlation among prevalence and mortality rates based on ethnogeographic region

Correlation between ASMRs and ASPRs for stroke and all comorbid conditions across each ethnogeographic location is shown in figure 3. High SBP prevalence and mortality show a strong positive correlation in Central & South Asia but a strong negative correlation in the Middle East. The prevalence and mortality of high BMI has a strong negative correlation in Central & South Asia and Middle East populations, but a strong positive correlation in East Asia. Prevalence of CKD has negative correlation with mortality rates in East Asia. Prevalence of T2D has negative correlation with mortality rates in Oceania and positive correlation in America. It is interesting to note that there was not much of a correlation in mortality and prevalence rates for most of the conditions. For overall stroke, though minor correlations between various ethnicities are seen, this becomes alarmingly clear in the stroke subtypes. The correlation matrix of mortality and prevalence rates of stroke and its comorbidities does reflect strong ethnogeographic distinctions, which formed the basis of further investigation on the genetic basis of stroke and its comorbidities.

**Figure 3:**
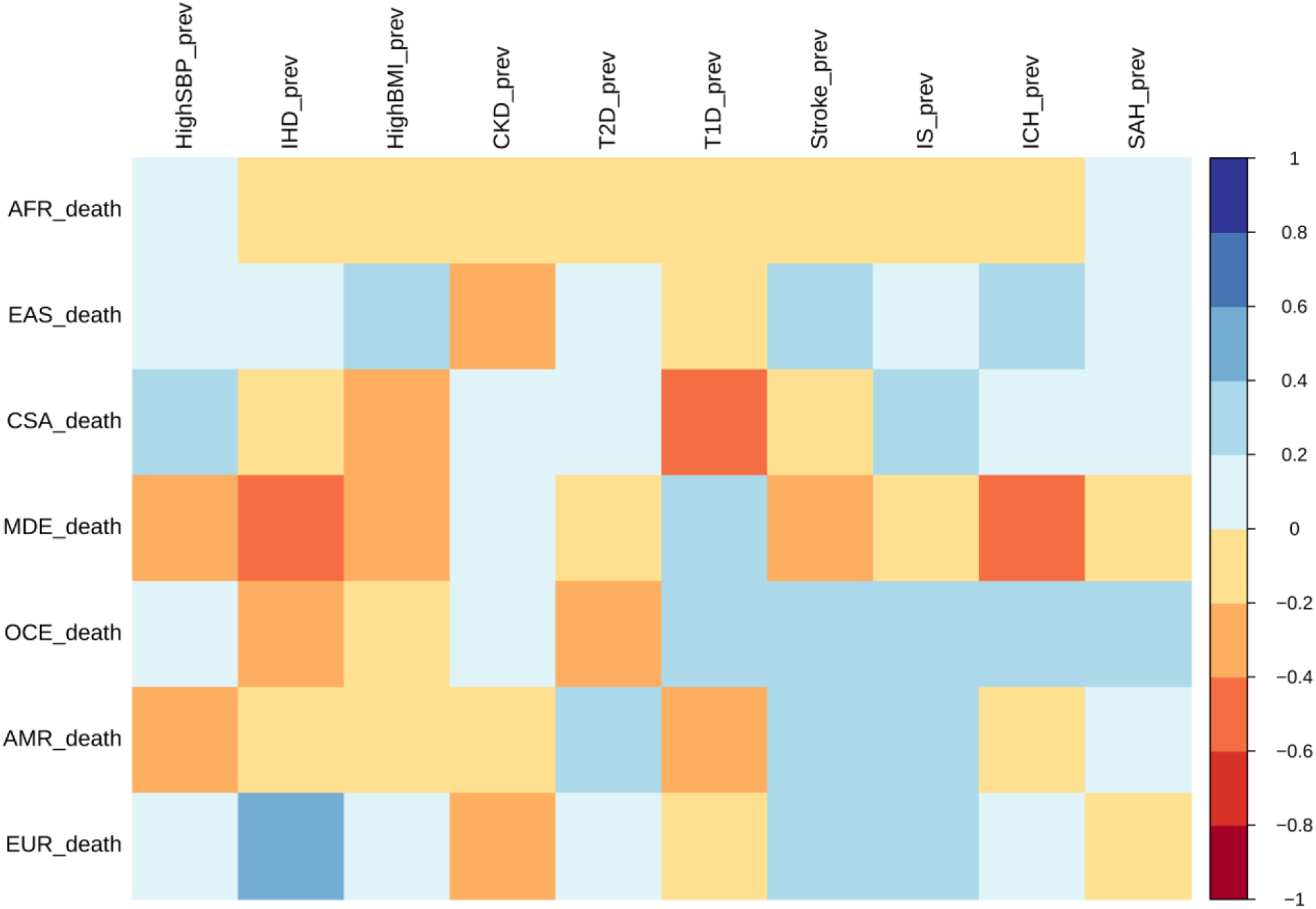
Correlation of mortality versus prevalence rates of Stroke, its subtypes and comorbid factors. The figure shows the Pearson correlation coefficients of age-standardized mortality rates versus prevalence rates across different continents for 2014. CSA – Central and South Asia, AFR – Africa, EAS – East Asia, EUR – Europe, MDE – Middle East, OCE – Oceania, AMR – America.

### Ethnogeographic stratification of stroke and its comorbidities based on GWAS data

To resolve the ethnogeographic distinctions for stroke and its major comorbid conditions based on their genetic risk, we considered all GWAS loci for stroke and its major comorbid conditions, and subjected it to stratification analysis. From the GWAS loci, we observed a distinct population structure that distinguished ethnogeographic populations based on their genetic signatures (Figure 4). For all diseases, except high BMI, the individuals clustered into five groups, each corresponding with the five super-populations from 1000 Genome project namely, African, East Asian, South Asian, European and American. For high BMI, the individuals clustered into three groups corresponding to African, East Asian, European. Though broad clustering of ancestral populations among the diseases looks similar, the proportions of ancestral populations in certain diseases vary greatly. Among Stroke, IHD and T2D risk variants, the populations structured in a similar way, while for T1D, CKD and LDL the patterns were slightly different in European and South Asian ancestry. Whereas for high SBP the major fluctuations were observed in South Asian and East Asian populations. This stratification was further explored using population-based clustering, where a similar pattern was observed in PCA plots for stroke and its comorbid conditions (Figure 5). African population seems to be a distinct outlier for most diseases, and the East Asian comes a distant second in the cluster pattern.

**Figure 4:**
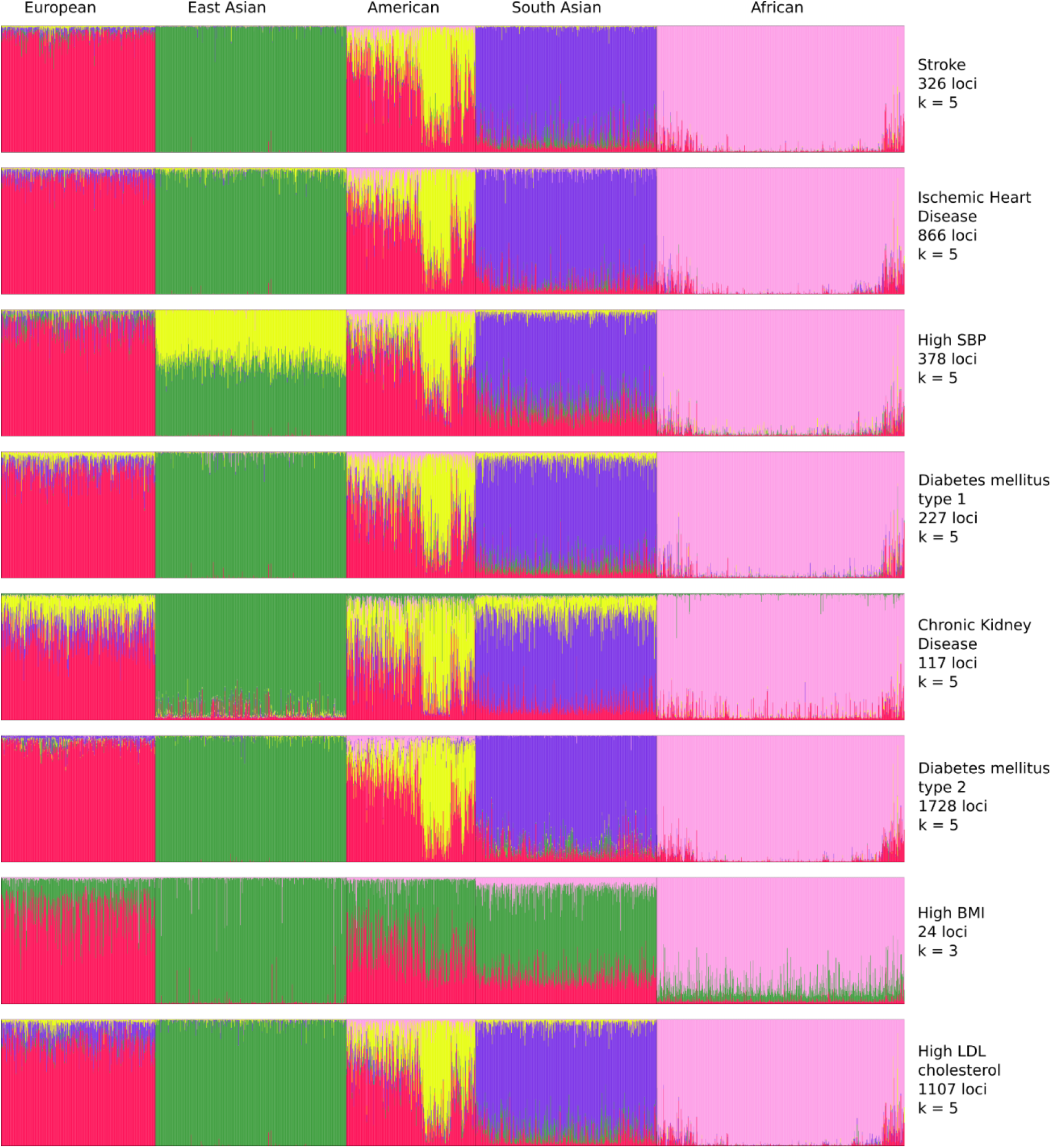
Population structure of risk variants for Stroke and its comorbid factors using a model-based clustering. The proportion of ancestral populations was estimated from the genotype of risk variants of each disease in unrelated individuals from the 1000 Genomes project using a model-based clustering approach. The individuals were represented by their super-population in 1000 Genomes (African, East Asian, South Asian, European and American). The estimated ancestral populations clustered into either 5 clusters or 3 clusters (in case of BMI) represented by the different colors.

**Figure 5:**
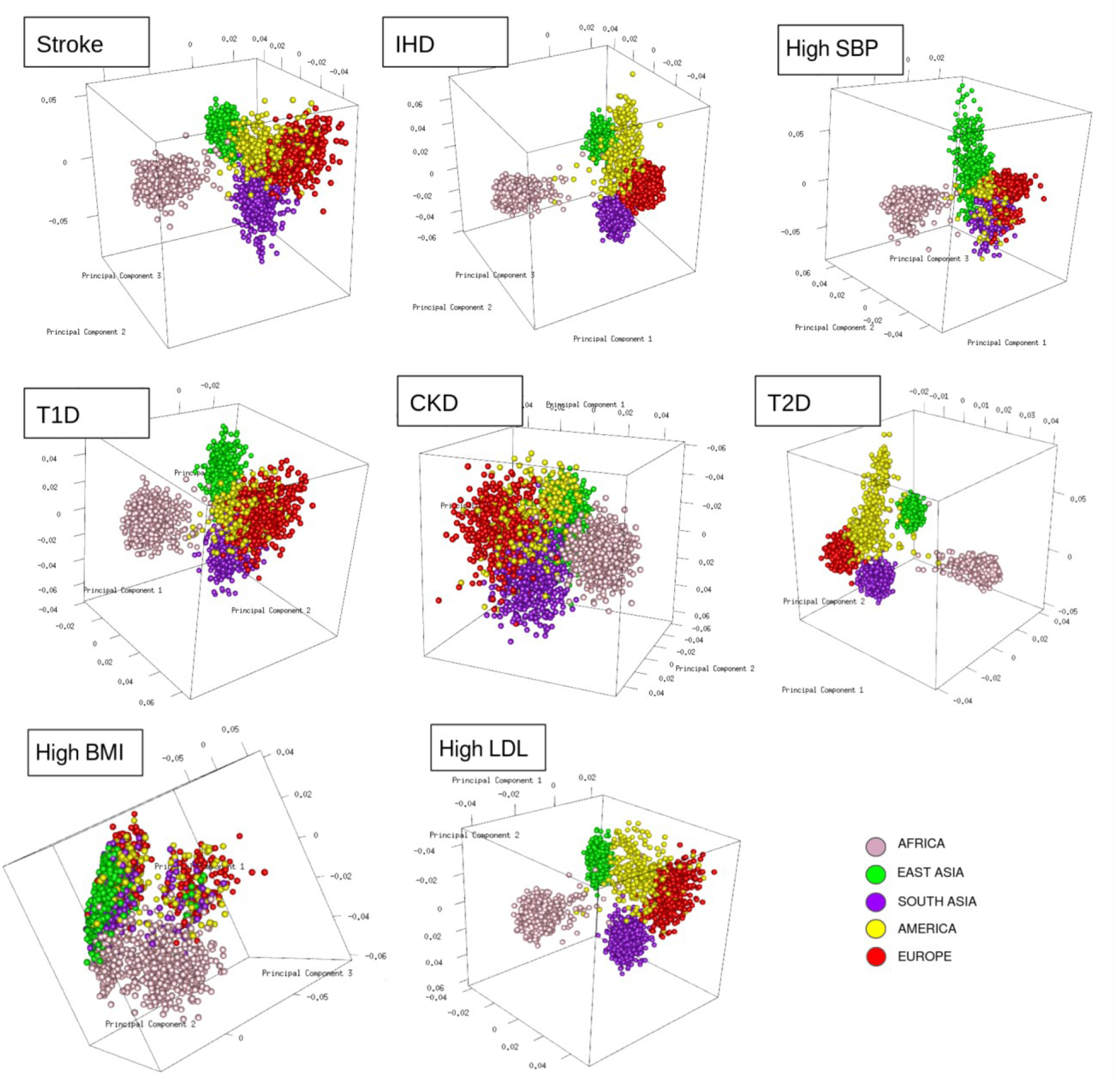
Clustering of risk variants for Stroke and its comorbid factors using a PCA. The eigenvectors were estimated from the genotype of risk variants of each disease in unrelated individuals from the 1000 Genomes project using PCA clustering. The individuals were represented by their super-population in 1000 Genomes (African, East Asian, South Asian, European and American) shown in five different colors.

Similar patterns were observed in stratification analysis after grouping the GWAS loci of stroke and its comorbidities into three groups as before, namely strokes, metabolic disorders and high SBP. The African, East Asian and South Asian populations had distinct structure in all three groups (Figure 4-figure supplement 1). These observations do indicate that the underlying genetic factors of stroke and its comorbid factors can be the real indicators of ethnogeographic patterns of risk for stroke and its comorbid conditions. However, we were further keen to understand the extent of shared and unique individual risk variants across stroke and its comorbid condition and how these unique or shared variants can help in distinguishing their relevance across ethnicities.

### Shared and unique risk variants of stroke among the different ethnogenetic regions

The unique and shared individual variants across stroke and its comorbid conditions were identified from the GWAS data irrespective of ethnicity. We find the majority of the risk variants were unique to a disease condition, however, several risk variants were also seen to be shared with stroke and other comorbid conditions as seen in Figure 6. We were further keen to have a deeper insight into the distribution of risk variants in stroke across ethnicities. Stroke has only 55% of the risk variants common to all the five populations as seen in Figure 7 and Table S6. Two groups of populations share the most number of variants, namely, the Africa-America-Europe-South Asia (6% of variants shared) group and the East Asia-America-Europe-South Asia (4%) group. Africa has the highest number of unique variants for stroke (6%), followed by Europe (3%).

**Figure 6:**
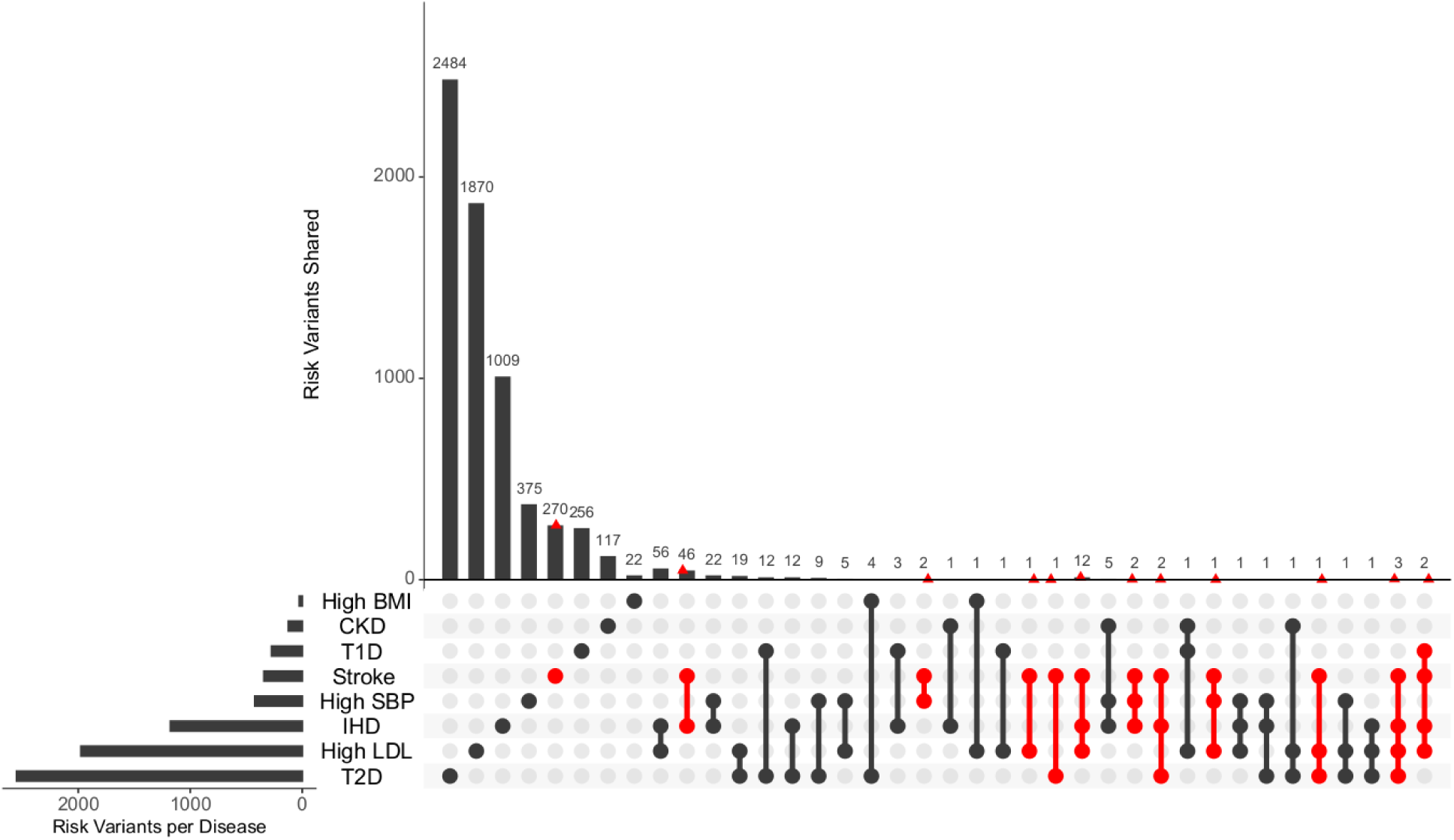
Distribution of unique and shared risk variants in stroke and its comorbid conditions. The barplot shows the number of GWAS risk variants unique for stroke and its comorbidities, as well as number of GWAS risk variants shared among the different diseases. The intersection of diseases is indicated below. Intersections containing stroke are highlighted in red. The total number of GWAS risk variants considered are stroke (366), IHD (1137), High SBP (419), T2D (2227), T1D (269), CKD (125), High BMI (27) and High LDL (1734).

**Figure 7:**
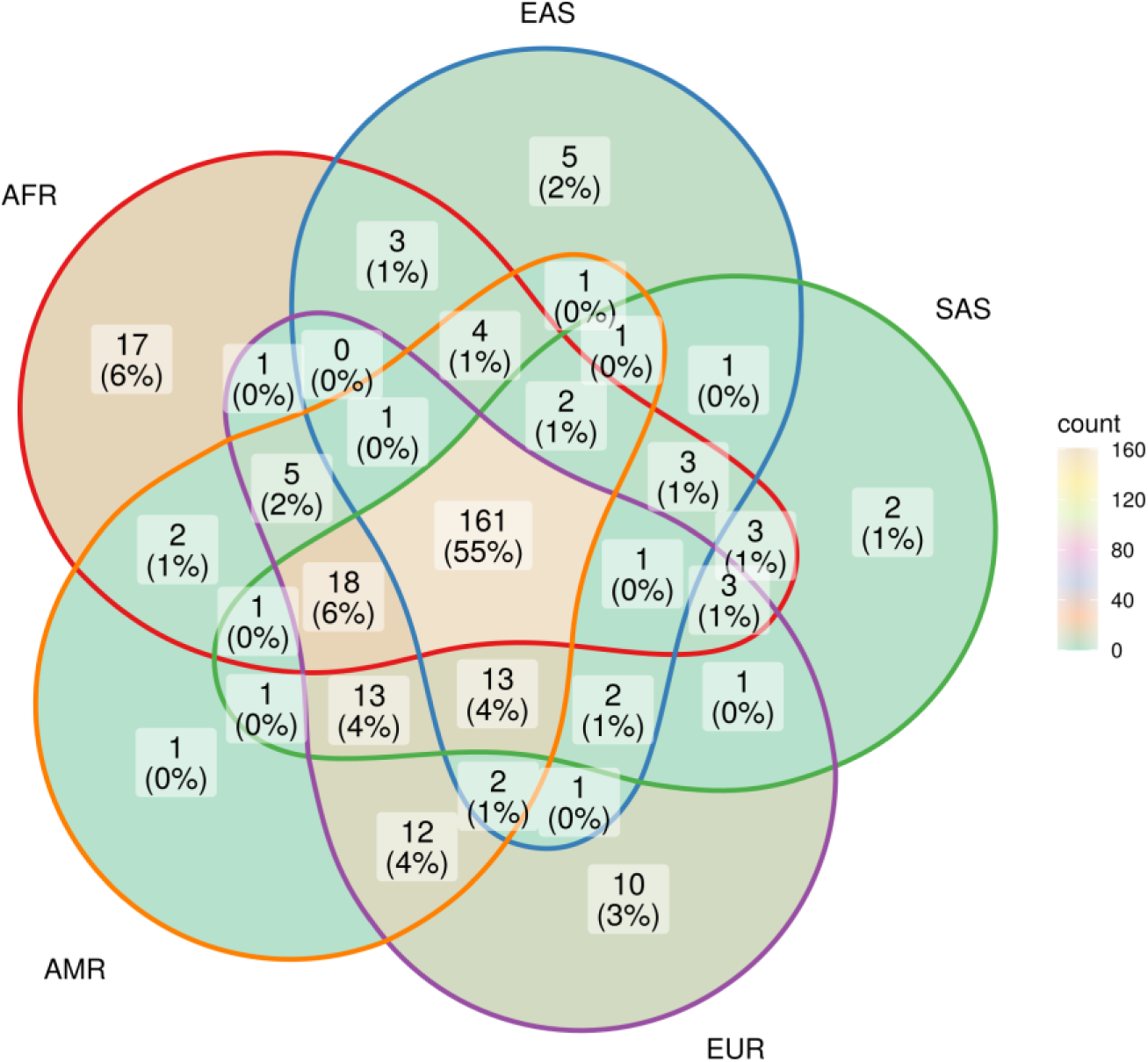
Risk variants of Stroke shared among super-populations. The GWAS risk variants for stroke (total 291) shared among the five super-populations in 1000 Genomes (African, East Asian, South Asian, European and American). A risk variant was considered to be present in a population if the alternate allele frequency in 1000 Genomes was greater than or equal to 0.05.

South Asia has two unique variants for stroke, rs528002287 and rs148010464, which maps to genes PCSK6 and the intergenic region of PLA2G4A/LINC01036, respectively. The variants rs528002287 and rs148010464 are low frequency variants in South Asia with a MAF of 0.053 and 0.054, respectively. We were further keen to understand how these unique variants in South Asia are tagged to the nearby variants of different frequencies in the different ethnicities. Can the LD patterns of rare allele and common allele help in distinguishing the ethnogeographic distinction in phenotype variation. In comparison, the linkage disequilibrium (LD) pattern of low frequency variants and common variants across ethnicities showed contrasting patterns. The LD plots between the unique variants and low frequency variants tagged to it clearly demonstrated a unique LD pattern in South Asia, compared to other populations (Figure 8). Contrastingly, the LD plots of common variants tagged to the risk variant of stroke unique to South Asia showed similar LD patterns among all populations (Figure 8-figure supplement 1). These differences might also reflect unique or distinct phenotypic differences among ethnicities for risk in stroke.

**Figure 8:**
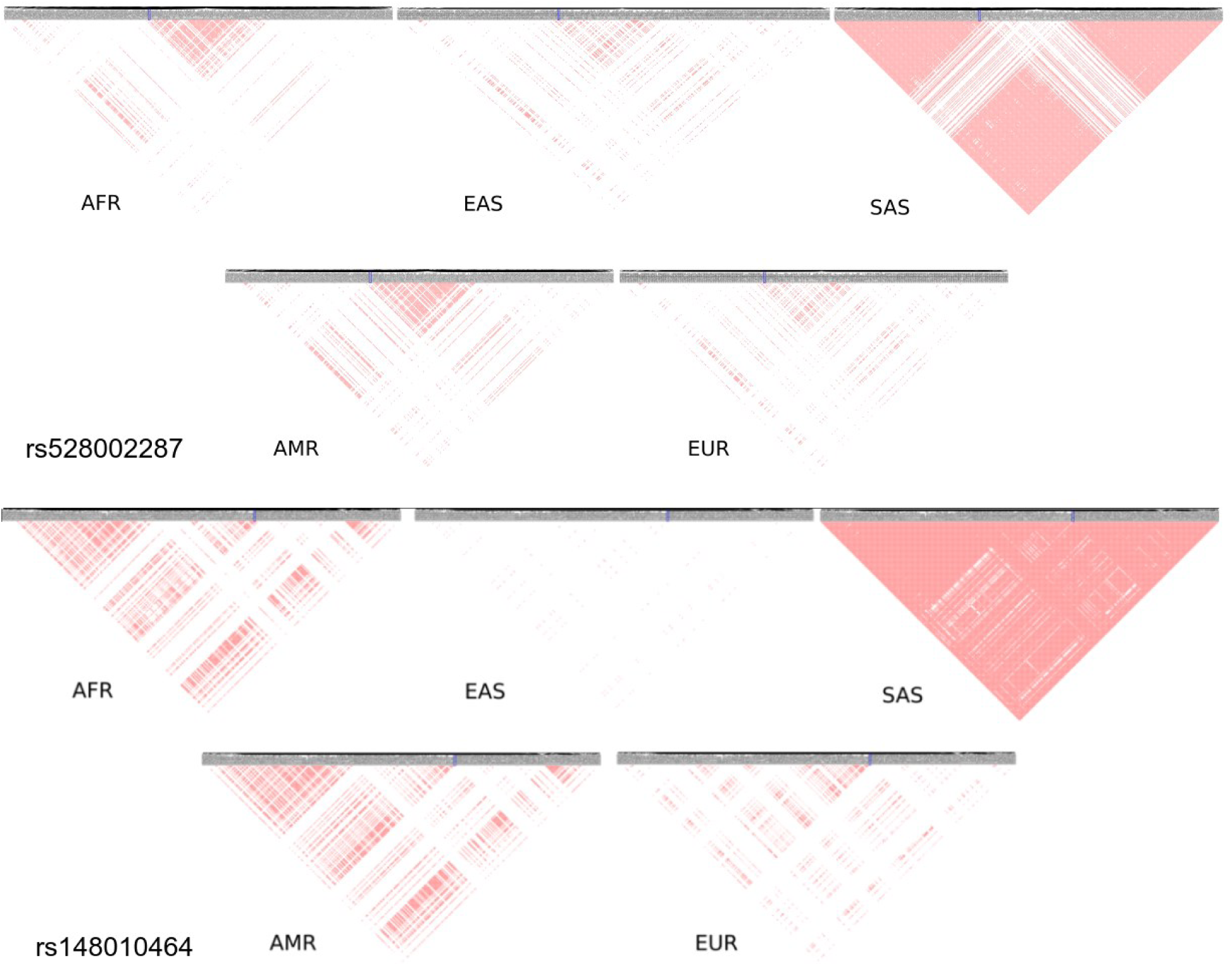
Linkage disequilibrium of low frequency variants proxy to risk variants of stroke unique in South Asian population. Proxy variants with frequency less than 0.1 in 1MB region flanking the stroke risk variants unique in South Asia (a) rs528002287 and (b) rs148010464. The risk variants are marked in blue box. LD plots of variants in different 1000 Genome super-populations (AFR-Africa, EAS-East Asia, SAS – South Asia, AMR – America, EUR – Europe) are shown.

## DISCUSSION

To the best of our knowledge, this is the first study that explored the burden of stroke and its comorbid conditions across regions, stratifying and distinguishing their unique features based on their genetic background extrapolated from the GWAS risk loci. The dynamics of different rates of stroke, its subtypes and comorbid factors do reflect ethnogeographic differences. Globally, the prevalence and incidence rates of stroke have increased, while mortality rates decreased with minor shifts in ranking in the last decade. Interestingly, the incidence and prevalence rank of stroke rates remain the same globally, but for mortality it ranks third, preceded only by IHD and high SBP. While the global stroke prevalence is nearly 15 times its mortality rate, prevalence of comorbid conditions such as high SBP, high BMI, CKD, T2D are alarmingly 150– to 500-fold higher than their mortality rates. These comorbid conditions can drastically affect the outcome of stroke. Interestingly, these disparities in rates get further widened when evaluated from an ethnogeographic perspective. The age-standardized prevalence of stroke in 2019 ranges from lowest in Central & South Asia (858.5/100000, 95%UI 737.6-979.4) to highest in East Asia (1513.1/100000, 95%UI 1390.7-1635.5), in contrast to mortality rates, lowest in America (40.3/100000, 95%UI 36.2-43.1) and highest in East Asia (127.7/100000, 95%UI 104.9-150.5).

The rates of stroke, its subtypes and comorbid conditions do correlate to some extent but their ranking varies significantly. In terms of ranking, stroke ranks sixth in prevalence across all ethnogeographic locations, but ranks second in mortality in East Asia and third in Central & South Asia and Africa. We find that among the considered comorbid conditions, some rank above stroke in both incidence and prevalence, however in terms of mortality, stroke ranks highest with exception to high SBP. Similarly, the ranking of the comorbid conditions also varies when the global population is stratified based on ethnogeographic locations. In the last decade, there has been tremendous development in the healthcare industry globally, but this is not reflected in the mortality or prevalence data from 2009-2019. Ranking of comorbid conditions by the rates is very crucial to identify ethnic-specific comorbid risk that can be helpful in guiding and managing stroke risk.

The changing dynamics of stroke or its comorbid conditions can be attributed to a multitude of factors. Often the global burden of stroke has been discussed from the point of view of socio-economic parameters. Studies indicate that half of the stroke-related deaths are attributable to poor management of modifiable risk factors^8,9^. However, we observe that different socio-economic regions are driven by different risk factors. Considering that Europe, America, Oceania and the Middle East represent high socio-economic regions, the comorbid conditions that drive prevalence and mortality rates here seem to be more of metabolic in nature, while for South Asians, high SBP is the prominent factor. It is evident from the correlation between prevalence and mortality for stroke, its subtypes and comorbid conditions that there is an ethnic and comorbid specific correlation, which possibly does not reflect a clear socio-economic distinction. The comorbid conditions for stroke subtypes also differ to a large extent. Diabetes is a comorbid condition for stroke but not for SAH, and this risk is ethnic-specific^10,11^. Therefore, it is very pertinent to understand the stroke risk from an ethnic view point, beyond the boundaries of socio-economic criteria, as the drivers of comorbid risk and ethnicity rely on genetic and epigenetic components. Studies reported reduction in life expectancy in 31 of 37 high-income countries, deduced to be due to COVID-19^12^. However, it would be unfair to ignore the comorbid conditions which could also be the critical determinants for reduced life expectancy in these countries.

Stroke has a complex etiology, which is further influenced by its comorbid conditions and this impacts its phenotypic variability. A strong genetic risk drives both stroke and its comorbid conditions. Genetic risk variants for diabetes, cardiovascular disease, diabetic retinopathies and nephropathies, hypertension, inflammation and kidney diseases have been reportedly shown to have strong ethnogenetic variation^13^. Implications of ethnogenetic differences were evident when GWAS genes for stroke and all studies of comorbid conditions were used to stratify the 1000 genome super-populations. We observed that the GWAS risk loci for stroke and its comorbid conditions like high BMI, high LDL, high systolic BP, type 2 diabetes, type 1 diabetes, and chronic kidney disease could stratify the super-populations based on its ethnogenetic considerations. Fluctuations in genetic structure of stroke and its comorbid conditions signify the impact of ethnic variations on mortality and prevalence rates. Stroke accounts for approximately 20% of deaths in diabetics^14,15^. Diabetics and pre-diabetics, and the duration of diabetes have been reported to have increased risk of stroke, which gets aggravated in African-Americans^14,15^. As stroke and its comorbid conditions are heavily influenced by lifestyle, high-income countries showed evidence of metabolic disorders being the major cause of concern for both prevalence and mortality. Similar observation in a UK biobank cohort study on stroke suggests that genetic and lifestyle factors were independently associated with incident stroke, which emphasizes the benefit of entire populations adhering to a healthy lifestyle, independent of genetic risk^16^.

Metabolic risk variations could also be a reflection of their underlying genetic differences. Significant differences among ethnicities in metabolism of various macromolecules have been reported^17^. Genetic variations demonstrate inter-ethnic differences in LDL levels resulting in differential impact on dyslipidemia^18^. A meta-analysis on European, East Asian and African-American ethnicities revealed that common variants of *CDH13* and *ADIPOQ* regulate adiponectin levels, an important component of BMI indicator^19^. *ALDH2*504Lys* allele has been reported to be associated with high BMI, increased tolerance of alcohol, high SBP, and decreased high-density lipoprotein in East Asians^20,21^. MEGASTROKE consortium on stroke and its comorbid factors reported five variants associated with blood pressure, two with LDL cholesterol and reported that all stroke subtypes were associated with a Genetic Risk Score for high SBP^22^. A study on IHD, T2D in European populations with different health-care systems, and local population substructures reported Polygenic risk score with similar accuracy across Europeans, to a lesser extent to South and East Asian populations, and very poor transferability for Africans^23^.

Even stroke subtypes show a strong ethnic variation^22,24^. While *COL4A1* and *COL4A2* were common denominators for most of the stroke subtypes, high-density lipoprotein was inversely associated with small vessel stroke^25^. The INTERSTROKE study on stroke subtypes reported that among comorbid factors, high SBP was significantly associated with ICH and diabetes, cardiac issues, and apolipoproteins with ischemic stroke^26^. Ischemic stroke associated with ApoB/ApoA1 ratio had higher (67.6%) population attributable fraction (PAF) in Southeast Asia compared to Western Europe, North America, and Australia (24.8%), while ischemic stroke associated with atrial fibrillation had lower PAF in South Asia (3.1%) compared to the rest (17.1%)^26^. Emerging studies like novel ethnic specific genetic variants, such as SUMOylation pathway in Indians^27^, *SFXN4* and *TMEM108* in Africans,^28^ indicate the involvement of different pathways among different ethnicities in stroke. The A allele of c.*84G>A loci in *CETP* gene was found to be a risk factor for IHD in South Asians^29^. High SBP was found to be a risk factor in all major stroke subtypes except lobar ICH^30^. Therefore, identifying differential risk in different ethnicities for stroke and its subtypes, and its impact on comorbid conditions might also indicate different treatment modalities which can minimize adverse metabolic side effects.

Identifying the pattern of genetic variation is critical in distinguishing stroke and its endophenotypic variations. The risk variants rs528002287 (locus 15q26.3) in *PCSK6* and rs148010464 (locus 1q31.1) an intergenic variant in *PLA2G4A/LINC01036* for stroke were unique to South Asia, and were found to be associated with cardioembolic stroke and small vessel stroke in South Asians^27^. A recent INTERSTROKE study reported the association of short sleep duration with increased risk for stroke to be highest in the South Asian ethnicity (OR 9.13, 95% CI 5.86-14.66)^31^. Decreased sleep quantity and quality has been reported to increase blood pressure^32^, prevalence for which was found to be highest for South Asians in our study. These observations are interesting as *PCSK6* is known to regulate sodium homeostasis and thereby maintain diastolic blood pressure.^33,34^ Reports also indicate 2.34 fold difference in the expression of *PCSK6* during maintenance phase of hypertension^35^. PCSK6 has also been reported to be involved in processing of precursors of Melanin Concentrating hormone (MCH) under certain conditions^36^. MCH is known to play a central role in promoting and stabilizing sleep^37,38^. In insomnia patients, *PLA2G4A* was reported to be upregulated by 1.88 fold after improvement in sleep^39^. In sleep deprived mice, glycolytic pathway and lipid metabolism was upregulated and expression of *PLA2G4A* was downregulated^40^. These observations are interesting as *PLA2G4A* is known to play a role in the metabolism of phospholipids, production of lipid mediators and the release of arachidonic acid^41,42^. Arachidonic acid is involved in signaling pathways of metabolic processes like release of insulin and glucose disposal^43–45^. *PLA2G4A* also plays a role in the production of pro-thrombotic TXA2 and thus, inhibition of *PLA2G4A* can reduce platelet aggregation and thromboembolism^46^. Thus, the risk of unique genetic variants in *PCSK6* and *PLA2G4A* in South Asian ancestry may indicate a unique endophenotype for stroke, which might also indicate the influence of underlying risk variants for comorbid conditions, e.g. *PLA2G4A* in metabolic processes. GWAS has yielded numerous common risk alleles that are associated with various human phenotypes^47,48^. Since the rationale for GWAS is the ‘common disease, common variant’ hypothesis, it has been able to identify only those common variants with a moderate effect on the associated trait and thus, these identified variants only explain a small proportion of the heritability of the trait. One of the major conclusions from the 1000 Genomes Project was that most variations in the human genome are rare and unique to specific sub-populations^49,50^. From an evolutionary point of view, alleles with strong effects that are detrimental will be controlled by purifying selection keeping its frequency low. Hence, rare and low frequency variants might be variants with large effects on traits^51^. Examples of genes like *ABCA1*, *PCSK9* and *LDLR*^52,53^, which carries both common variants with moderate effects as well as rare variants with large effect for lipid levels indicate that genes can contain both types of variants associated with a complex trait. Using this logic, we were keen to identify the influence of common and rare variants in stroke and its comorbid conditions and their pattern of LD in distinguishing ethnic specific risk.

While we observe the majority of the GWAS variants associated with stroke are common variants, a minority of these are low frequency variants with below 10% frequency. The alternate alleles of these variants were found to be present only in specific super-populations of 1000 Genomes, while the variants are monoallelic in the other super-populations. This difference among populations gets further enhanced when we look at the LD patterns of these low frequency variants with other low frequency variants which are in proxy. Distinct LD blocks with these unique rare variants present in one super-population were seen to be absent in other populations. On the other hand, LD patterns of common variants (frequency greater than 10%) in the same regions show similar LD patterns for all populations. Thus, the rare variant hypothesis could explain a significant proportion of the differences in burden of stroke seen across populations. Such genes identified could be possible candidate genes for identifying rare and low frequency variants that could play a role in the heritability of stroke and its comorbid factors.

## CONCLUSIONS

The dynamics of incidence, prevalence and mortality rates of stroke and its subtypes along with its comorbid risk factors reflect strong ethnogeographic differences. Our work highlights that these ethnogeographic differences for stroke and its comorbid conditions need to be evaluated and stratified based on their ethnogenetic background. Genetic variables should be considered as primary evidence as they define the threshold for biochemical, metabolomic or epigenetic variables. The different socio-economic regions are driven by different risk factors of stroke and low frequency variants could be playing a role in the differences in burden of stroke seen in the different regions. Identifying population specific unique variants for stroke and its comorbid conditions might refine the drivers for endophenotypic variations for stroke risk. We would like to suggest that integrating public health genomics and articulating it with comorbid conditions for stroke should be considered crucial irrespective of the economic status, as both lower and higher socio-economic regions have different drivers of stroke risk.

## METHODS

### Data sources

We obtained age-standardized incidence rates (ASIRs), age-standardized prevalence rates (ASPRs) and age-standardized mortality rates (ASMRs) for a total of eight diseases and three disease conditions in 204 countries, for the years 2009 to 2019 using the GBD Results Tool (vizhub.healthdata.org/gbd-results/)^54^ and from NCD Risk Factor Collaboration (NCD-RisC, ncdrisc.org/) study^55,56^ in November 2021. GBD codes for the selected diseases were B.2.2 (Ischemic heart disease, IHD), B.2.3 (Stroke), B.2.3.1 (Ischemic stroke), B.2.3.2 (Intracerebral hemorrhage, ICH), B.2.3.3 (Subarachnoid hemorrhage, SAH), B.8.1.1 (Diabetes mellitus type 1, T1D), B.8.1.2 (Diabetes mellitus type 2, T2D) and B.8.2 (Chronic kidney disease, CKD). Three disease conditions, which are also comorbid factors of stroke, were also selected, high systolic blood pressure (SBP) (>110-115 mmHg), high body mass index (BMI > 23.0 kg/m^2^), and high LDL cholesterol. For these, ASMRs in 204 countries, as well as global rates for 2009 to 2019 were obtained using the GBD Results Tool, and age-standardized prevalence percentages in 204 countries for the years 2009 to 2016 (data for 2017 to 2019 was not available) were obtained using NCD-Risc website (ncdrisc.org/)^55,56^. Global crude rates and age-standardized incidence, prevalence and mortality rates were obtained from GBD and NCD for all the diseases and disease conditions (Figure 1-figure supplement 1, Table S1). GBD 2019 and NCD-RisC study compiled with the GATHER Guidelines.

### Spatio-temporal trend analysis and estimated annual percent change

The 204 countries were grouped into eight geographic regions namely Global, America, Europe, Middle East, Africa, Central Asia & South Asia, East Asia, and Oceania, based on ethnicity (Table S7, Appendix Figure 1). The prevalence percentages were converted into ASPRs for each disease condition. ASMRs and ASPRs for each region were obtained using Bayesian model averaging of linear regression models with Markov chain Monte Carlo sampling. BIC was the model selection criteria. Poisson distribution with global rates as lambda was the prior distribution on the models. The population of each country was used as weights. 10000 draws from the posterior distribution of model parameters were used to obtain the point estimates (mean of the draws) and 95% uncertainty intervals (2.5th to 97.5th percentiles of the posterior distributions) of mortality and prevalence rates for each region for years 2009 to 2019. Rate estimates presented in this paper are age-standardized rates per 100,000 population.

The ASMRs and ASPRs thus obtained for the different diseases in each region were subjected to a temporal rank analysis for 2009, 2014, and 2019 using custom perl scripts and change in the trend was plotted as a bump plot. The size and position of the points indicates the rate and rank respectively. To quantify the temporal trends, estimated annual percentage change (EAPC) was modeled using Poisson regression using a generalized linear model for the log-transformed rates: *log(y)= β0+ β1×1 + β2×2 + ….+βpxp*, where y is the age-standardized rate, x_i_ are the calendar years, *β*_i_ are the rate trends. Under the assumption of linearity of log of age-standardized rate with time, EAPC =100* exp(β^) – 1. The 95% uncertainty interval is calculated as CI(EAPC) = β + (Z_(1-α)/2_) x SE, where α is the confidence level, SE is the standard error of β^.

EAPC for ASMRs and ASPRs were calculated for the time period 2009 to 2019 (Table 1 and 2). EAPC for high SBP prevalence was calculated from 2009 to 2015 and for high BMI prevalence from 2009 to 2016. EAPC was considered statistically significant if the uncertainty interval of EAPC did not cross zero. Statistical significance of spatio-temporal difference in rates was calculated using chi-square test by assuming the rate to be under Poisson distribution. 2019 ASMRs (or ASPRs) in each region were compared with global ASMR (or ASPR) to compare change over locations and with 2009 ASMRs (ASPRs) to compare change over time. 2014 ASPRs were used when 2019 data was not available. P-values (two-sided) for ASMRs and ASPRs are shown in Table S2 and S3, respectively. Chi-square tests were done using Open Source Epidemiologic Statistics for Public Health (www.openepi.com). The relation between ASMRs and ASPRs of each region was measured using Pearson correlation. For correlation, we considered the data of 2014 due to the complete spectrum of data availability. All analysis, unless specified, was done using R Statistical Software (version 4.1.2)^57^ with packages *BAS*^58^, *Rcan*^59^, *corrplot*^60^, *dplyr* and *ggplot2*.

### Proportional mortality and prevalence

The diseases were classified into three categories as stroke (ischemic stroke, ICH, SAH and IHD), metabolic disorders (High BMI, High LDL, T2D, CKD, and T1D) and high SBP. To determine the proportion of each category in a region, total ASMRs were scaled to 100 for all three years. The same was done for ASPRs (high LDL cholesterol data was not available). Statistical significance of difference in proportions was calculated using a one-sample test for binomial proportion using normal-theory method. The proportion mortality in each category was compared in a pairwise manner with global as well other regional proportions to calculate the two-sided p-value. The same was done for proportional prevalence. P-values for proportional mortality and prevalence are shown in Tables S4 and S5, respectively. Proportion comparisons were done using Open Source Epidemiologic Statistics for Public Health (www.openepi.com/Menu/OE_Menu.htm).

### Population Structure Analysis

For evaluating the ethnogenetic perspective of stroke and its comorbid conditions we considered the risk variants associated with each disease. The risk variants were obtained from GWAS Catalog (www.ebi.ac.uk/gwas/home), during the period November 2021 to August 2022. The trait IDs used to retrieve data from GWAS Catalog for the different diseases were EFO_0000712 (Stroke), EFO_0001645 (IHD), EFO_0000537 (High SBP), MONDO_0005148 (T2D), MONDO_0005147 (T1D), EFO_0003884 (CKD), EFO_0007041 (High BMI) and EFO_0004611 (High LDL). The position of the risk variants in GRCh37 assembly was obtained, and variants less than 10000 bp apart were excluded using custom perl scripts. The total number of risk variants for each disease thus obtained are shown in Table S8. The genotype of identified biallelic autosomal SNPs in unrelated individuals was extracted from the 1000 Genome VCF files (www.internationalgenome.org)^53^. The proportion of ancestral populations in each individual was estimated from their genotype using a model-based clustering approach^61^. The admixture model with correlated allele frequencies was specified to cluster the individuals into either five clusters or three clusters (in case of BMI). The genotype was converted to eigenvectors using principal component analysis^62,63^.

### Shared and Unique risk variants among ethnicities

The number of GWAS risk variants for stroke shared among, as well as, unique to the five super-populations in 1000 Genomes (African, East Asian, South Asian, European and American) was obtained (Figure 6 and Table S6). A risk variant was considered to be present in a population if the alternate allele frequency in 1000 Genomes was greater than or equal to 0.05. For each GWAS risk loci, the gene(s) the variant maps to was obtained and shared genes among the populations was also estimated (Table S6).

### Calculation of Linkage Disequilibrium (LD) of risk variants of stroke in South Asia

Proxy variants (R2 > 0.01) in the region –/+ 500 Kb of the risk variant of stroke present in the South Asian population was obtained from LDlink^64^. Among the proxy variants, variants with minor allele frequency (MAF) less than 0.1 were selected along with the risk variant as low frequency variants, and variants with MAF greater than 0.1 were termed as common variants. LD between two alleles A and B is quantified using the coefficient of linkage disequilibrium DAB calculated using the equation DAB = pAB-pApB, where pi represents the frequency of the allele i or haplotype i. To be able to compare the level of LD between different pairs of alleles, D is normalized as follows: D’ = D/ Dmax, where Dmax = max{-pApB, –(1-pA)(1-pB)} when D < 0 and Dmax = min{pB(1-pA), pA(1-pB)} when D > 0^65^. Estimates of D’ were calculated separately for low frequency variants and common variants, and plotted using the R library *gaston*^66^.

## Data Availability

The datasets analysed during the current study are available in the GBD database, at https://vizhub.healthdata.org/gbd-results/?params=gbd-api-2019-permalink/fe8b05e8222bcf3ec3af555762006f2a. All other materials including computer code will be available upon request. Please contact corresponding author for the same.

## ACKNOWLEDGEMENTS

RS acknowledges the support of Kerala State Council for Science, Technology and Environment, (KSCSTE) for providing the research fellowship. RS and ASN acknowledge the SIUCEB support at the Department of Computational Biology and Bioinformatics, University of Kerala for providing the necessary facilities to carry out the work. MB acknowledges the Dept. of Biotechnology for providing intra-mural support to carry out the work.

## AUTHOR CONTRIBUTIONS

RS, ASN and MB conceptualized and designed the workflow, and wrote the manuscript. RS performed the trend and statistical analyses, principal component analysis and population structure analysis, and generated figures and tables. ASN and MB provided overall guidance and direction.

## MATERIALS & CORRESPONDENCE

The datasets analyzed during the current study are available in the GBD database, at https://vizhub.healthdata.org/gbd-results/?params=gbd-api-2019-permalink/fe8b05e8222bcf3ec3af555762006f2a.

All other materials including computer code will be available upon request. Please contact the corresponding author for the same.

## Supplementary Files

Supplementary File 1

Table S1 Global incidence, mortality and prevalence rates for Stroke, its subtypes and comorbid factors.

Supplementary File 2

Table S2. Spatio-temporal comparison of mortality rates of Stroke, its subtypes and comorbid factors.

Table S3. Spatio-temporal comparison of prevalence rates of Stroke, its subtypes and comorbid factors.

Supplementary File 3

Table S4. Comparison of proportional mortality of Stroke, HSBP and Metabolic conditions.

Table S5. Comparison of proportional prevalence of Stroke, HSBP and Metabolic conditions.

Supplementary File 4

Table S6. Variants and genes associated with Stroke and its comorbid factors that are shared among populations, as well as unique to populations.

Supplementary File 5

Table S7. Continent Regions.

Supplementary File 6

Table S8. Number of final risk variants for each disease.

Supplementary File 7

Figure 1-figure supplement 1. Global incidence, mortality and prevalence rates for Stroke, its subtypes and comorbid factors

Figure 1-figure supplement 2. Boxplots of point estimates and uncertainty intervals of age-standardised mortality rates of (A) stroke, its subtypes and (B) comorbid conditions across regions from 2009-2019

Figure 1-figure supplement 3. Boxplots of point estimates and uncertainty intervals of age-standardised prevalence rates of (A) stroke, its subtypes and (B) comorbid conditions across regions from 2009-2019

Figure 4-figure supplement 1. Population structure of risk variants for Strokes, High SBP and Metabolic disorders using a model-based clustering

Figure 8-figure supplement 1. Linkage disequilibrium of common variants proxy to risk variants of stroke unique in South Asian population.

Appendix Figure 1. Population across different continents in 2009, 2014 and 2019

## REFERENCES

1. GBD 2019 Stroke Collaborators. Global, regional, and national burden of stroke and its risk factors, 1990-2019: a systematic analysis for the Global Burden of Disease Study 2019. Lancet Neurol. 2021 Oct;20(10):795–820. doi: 10.1016/S1474-4422(21)00252-0. Epub 2021 Sep 3. PMID: 34487721.

2. Gallacher KI, Jani BD, Hanlon P, Nicholl BI, Mair FS. Multimorbidity in Stroke. Stroke. 2019 Jul;50(7):1919–1926. doi: 10.1161/STROKEAHA.118.020376. Epub 2019 Apr 11. PMID: 31233391.

3. Tarko L, Costa L, Galloway A, Ho YL, Gagnon D, et al. Racial and Ethnic Differences in Short– and Long-term Mortality by Stroke Type. Neurology. 2022 Jun 13;98(24):e2465–e2473. doi: 10.1212/WNL.0000000000200575. PMID: 35649728.

4. Gardener H, Sacco RL, Rundek T, Battistella V, Cheung YK, et al. Race and Ethnic Disparities in Stroke Incidence in the Northern Manhattan Study. Stroke. 2020 Apr;51(4):1064–1069. doi: 10.1161/STROKEAHA.119.028806. Epub 2020 Feb 12. PMID: 32078475.

5. Mkoma, G.F., Johnsen, S.P., Iversen, H.K., Andersen, G., Nørredam, M.L.. Ethnic differences in incidence and mortality of stroke in Denmark, European Journal of Public Health, Volume 30, Issue Supplement_5, September 2020, ckaa165.346, 10.1093/eurpub/ckaa165.346.

6. Schmidt M, Jacobsen JB, Johnsen SP, Bøtker HE, Sørensen HT. Eighteen-year trends in stroke mortality and the prognostic influence of comorbidity. Neurology. 2014 Jan 28;82(4):340–50. doi: 10.1212/WNL.0000000000000062. Epub 2013 Dec 20. PMID: 24363134.

7. Gallacher, K. I., McQueenie, R., Nicholl, B., Jani, B. D., Lee, D., et al. (2018). Risk factors and mortality associated with multimorbidity in people with stroke or transient ischaemic attack: a study of 8,751 UK Biobank participants. Journal of comorbidity, 8(1), 1–8. 10.15256/joc.2018.8.129

8. Avan A, Digaleh H, Di Napoli M, Stranges S, Behrouz R, et al. Socioeconomic status and stroke incidence, prevalence, mortality, and worldwide burden: an ecological analysis from the Global Burden of Disease Study 2017. BMC Med. 2019 Oct 24;17(1):191. doi: 10.1186/s12916-019-1397-3. PMID: 31647003.

9. Baatiema L, de-Graft Aikins A, Sarfo FS, Abimbola S, Ganle JK, et al. Improving the quality of care for people who had a stroke in a low-/middle-income country: A qualitative analysis of health-care professionals’ perspectives. Health Expect. 2020 Apr;23(2):450–460. doi:10.1111/hex.13027.PMID:31967387.

10. Lindgren AE, Kurki MI, Riihinen A, Koivisto T, Ronkainen A, et al. Type 2 diabetes and risk of rupture of saccular intracranial aneurysm in eastern Finland. Diabetes Care. 2013 Jul;36(7):2020–6. doi: 10.2337/dc12-1048. PMID: 23536581.

11. Koshy L, Easwer HV, Premkumar S, Alapatt JP, Pillai AM, et al. Risk factors for aneurysmal subarachnoid hemorrhage in an Indian population. Cerebrovasc Dis. 2010 Feb;29(3):268–74. doi: 10.1159/000275501. Epub 2010 Jan 15. PMID: 20090318.

12. Islam N, Jdanov DA, Shkolnikov VM, Khunti K, Kawachi I, et al. Effects of covid-19 pandemic on life expectancy and premature mortality in 2020: time series analysis in 37 countries. BMJ. 2021 Nov 3;375:e066768. doi: 10.1136/bmj-2021-066768. PMID: 34732390.

13. Shoily SS, Ahsan T, Fatema K, Sajib AA. Common genetic variants and pathways in diabetes and associated complications and vulnerability of populations with different ethnic origins. Sci Rep. 2021 Apr 5;11(1):7504. doi: 10.1038/s41598-021-86801-2. PMID: 33820928; PMCID: PMC8021559.

14. Banerjee C, Moon YP, Paik MC, Rundek T, Mora-McLaughlin C, et al. Duration of diabetes and risk of ischemic stroke: the Northern Manhattan Study. Stroke. 2012 May;43(5):1212–7. doi: 10.1161/STROKEAHA.111.641381. Epub 2012 Mar 1. PMID: 22382158.

15. Boehme AK, Esenwa C, Elkind MS. Stroke Risk Factors, Genetics, and Prevention. Circ Res. 2017 Feb 3;120(3):472–495. doi: 10.1161/CIRCRESAHA. 116.308398. PMID: 28154098.

16. Rutten-Jacobs LC, Larsson SC, Malik R, Rannikmäe K; MEGASTROKE consortium; et al. Genetic risk, incident stroke, and the benefits of adhering to a healthy lifestyle: cohort study of 306 473 UK Biobank participants. BMJ. 2018 Oct 24;363:k4168. doi: 10.1136/bmj.k4168. PMID: 30355576.

17. Vasishta S, Ganesh K, Umakanth S, Joshi MB. Ethnic disparities attributed to the manifestation in and response to type 2 diabetes: insights from metabolomics. Metabolomics. 2022 Jun 28;18(7):45. doi: 10.1007/s11306-022-01905-8. PMID: 35763080.

18. Klarin D, Damrauer SM, Cho K, Sun YV, Teslovich TM, et al. Genetics of blood lipids among ∼300,000 multi-ethnic participants of the Million Veteran Program. Nat Genet. 2018 Nov;50(11):1514–1523. doi: 10.1038/s41588-018-0222-9. Epub 2018 Oct 1. PMID: 30275531.

19. Dastani Z, Hivert MF, Timpson N, Perry JR, Yuan X, et al. Novel loci for adiponectin levels and their influence on type 2 diabetes and metabolic traits: a multi-ethnic meta-analysis of 45,891 individuals. PLoS Genet. 2012;8(3):e1002607 doi: 10.1371/journal.pgen.1002607. Epub 2012 Mar 29. PMID: 22479202.

20. Takeuchi F, Isono M, Nabika T, Katsuya T, Sugiyama T, et al. Confirmation of ALDH2 as a Major locus of drinking behavior and of its variants regulating multiple metabolic phenotypes in a Japanese population. Circ J. 2011;75(4):911–8. doi: 10.1253/circj.cj-10-0774. Epub 2011 Mar 1. PMID: 21372407.

21. Xu F, Chen Y, Lv R, Zhang H, Tian H, et al. ALDH2 genetic polymorphism and the risk of type II diabetes mellitus in CAD patients. Hypertens Res. 2010 Jan;33(1):49–55. doi: 10.1038/hr.2009.178. Epub 2009 Oct 30. PMID: 19876063.

22. Malik R, Chauhan G, Traylor M, Sargurupremraj M, Okada Y, et al. Multiancestry genome-wide association study of 520,000 subjects identifies 32 loci associated with stroke and stroke subtypes. Nat Genet. 2018 Apr;50(4):524–537. doi: 10.1038/s41588-018-0058-3. Epub 2018 Mar 12.

23. Mars N, Kerminen S, Feng YA, Kanai M, Läll K, et al. Genome-wide risk prediction of common diseases across ancestries in one million people. Cell Genom. 2022 Apr 13;2(4): None. doi: 10.1016/j.xgen.2022.100118. PMID: 35591975.

24. Hu Y, Haessler JW, Manansala R, Wiggins KL, Moscati A, et al. Whole-Genome Sequencing Association Analyses of Stroke and Its Subtypes in Ancestrally Diverse Populations From Trans-Omics for Precision Medicine Project. Stroke. 2022 Mar;53(3):875–885. doi: 10.1161/STROKEAHA.120.031792. Epub 2021 Nov 3. PMID: 34727735.

25. Meschia JF. Effects of Genetic Variants on Stroke Risk. Stroke. 2020 Mar;51(3):736–741. doi: 10.1161/STROKEAHA.119.024158. Epub 2020 Feb 12. PMID: 32078489.

26. O’Donnell MJ, Chin SL, Rangarajan S, Xavier D, Liu L, et al. Global and regional effects of potentially modifiable risk factors associated with acute stroke in 32 countries (INTERSTROKE): a case-control study. Lancet. 2016 Aug 20;388(10046):761–75. doi: 10.1016/S0140-6736(16)30506-2. Epub 2016 Jul 16. PMID: 27431356.

27. Kumar A, Chauhan G, Sharma S, Dabla S, Sylaja PN, et al. Association of SUMOylation Pathway Genes With Stroke in a Genome-Wide Association Study in India. Neurology. 2021 Jul 27;97(4):e345–e356. doi: 10.1212/WNL.0000000000012258. Epub 2021 May 24. PMID: 34031191.

28. Keene KL, Hyacinth HI, Bis JC, Kittner SJ, Mitchell BD, et al. Genome-Wide Association Study Meta-Analysis of Stroke in 22 000 Individuals of African Descent Identifies Novel Associations With Stroke. Stroke. 2020 Aug;51(8):2454–2463. doi: 10.1161/STROKEAHA.120.029123. Epub 2020 Jul 22. PMID: 32693751; PMCID: PMC7387190.

29. Ganesan M, Nizamuddin S, Katkam SK, Kumaraswami K, Hosad UK, et al. c.*84G>A Mutation in CETP Is Associated with Coronary Artery Disease in South Indians. PLoS One. 2016 Oct 21;11(10):e0164151. doi: 10.1371/journal.pone. 0164151. PMID: 27768712.

30. Georgakis MK, Gill D, Webb AJS, Evangelou E, Elliott P, et al. Genetically determined blood pressure, antihypertensive drug classes, and risk of stroke subtypes. Neurology. 2020 Jul 28;95(4):e353–e361. doi: 10.1212/WNL.0000000000009814. Epub 2020 Jul 1. PMID: 32611631.

31. Mc Carthy CE, Yusuf S, Judge C, Alvarez-Iglesias A, Hankey GJ, et al. Sleep Patterns and the Risk of Acute Stroke: Results From the INTERSTROKE International Case-Control Study. Neurology. 2023 May 23;100(21):e2191–e2203. doi: 10.1212/WNL.0000000000207249. Epub 2023 Apr 5. PMID: 37019662.

32. Sabanayagam C, Shankar A. Sleep duration and cardiovascular disease: results from the National Health Interview Survey. Sleep. 2010 Aug;33(8):1037–42. doi: 10.1093/sleep/33.8.1037. PMID: 20815184.

33. Li JP, Wang XB, Chen CZ, Xu X, Hong XM, et al. The association between paired basic amino acid cleaving enzyme 4 gene haplotype and diastolic blood pressure. Chin Med J (Engl). 2004 Mar;117(3):382–8. PMID: 15043778.

34. Chen S, Cao P, Dong N, Peng J, Zhang C, et al. PCSK6-mediated corin activation is essential for normal blood pressure. Nat Med. 2015 Sep;21(9):1048–53. doi: 10.1038/nm.3920. Epub 2015 Aug 10. PMID: 26259032.

35. Marques FZ, Campain AE, Yang YH, Morris BJ. Meta-analysis of genome-wide gene expression differences in onset and maintenance phases of genetic hypertension. Hypertension. 2010 Aug;56(2):319–24. doi: 10.1161/HYPERTENSIONAHA.110.155366. Epub 2010 Jun 28.

36. Viale A, Ortola C, Hervieu G, Furuta M, Barbero P, et al. Cellular localization and role of prohormone convertases in the processing of pro-melanin concentrating hormone in mammals. J Biol Chem. 1999 Mar 5;274(10):6536–45. doi: 10.1074/jbc.274.10.6536. PMID: 10037747.

37. Jego S, Glasgow SD, Herrera CG, Ekstrand M, Reed SJ, et al. Optogenetic identification of a rapid eye movement sleep modulatory circuit in the hypothalamus. Nat Neurosci. 2013 Nov;16(11):1637–43. doi: 10.1038/nn.3522. Epub 2013 Sep 22. PMID: 24056699.

38. Konadhode RR, Pelluru D, Blanco-Centurion C, Zayachkivsky A, Liu M, et al. Optogenetic stimulation of MCH neurons increases sleep. J Neurosci. 2013 Jun 19;33(25):10257–63. doi: 10.1523/JNEUROSCI.1225-13.2013. PMID: 23785141.

39. Livingston WS, Rusch HL, Nersesian PV, Baxter T, Mysliwiec V, et al. Improved Sleep in Military Personnel is Associated with Changes in the Expression of Inflammatory Genes and Improvement in Depression Symptoms. Front Psychiatry. 2015 Apr 30;6:59. doi: 10.3389/fpsyt.2015.00059. PMID: 25983695.

40. Hinard V, Mikhail C, Pradervand S, Curie T, Houtkooper RH, et al. Key electrophysiological, molecular, and metabolic signatures of sleep and wakefulness revealed in primary cortical cultures. J Neurosci. 2012 Sep 5;32(36):12506–17. doi: 10.1523/JNEUROSCI.2306-12.2012. PMID: 22956841.

41. Burke JE, Dennis EA. Phospholipase A2 biochemistry. Cardiovasc Drugs Ther. 2009 Feb;23(1):49–59. doi: 10.1007/s10557-008-6132-9. Epub 2008 Oct 18. PMID: 18931897.

42. Shimizu T, Ohto T, Kita Y. Cytosolic phospholipase A2: biochemical properties and physiological roles. IUBMB Life. 2006 May-Jun;58(5-6):328–33. doi: 10.1080/15216540600702289. PMID: 16754327.

43. Chen M, Yang Z, Naji A, Wolf BA. Identification of calcium-dependent phospholipase A2 isoforms in human and rat pancreatic islets and insulin secreting beta-cell lines. Endocrinology. 1996 Jul;137(7):2901–9. doi: 10.1210/endo. 137.7.8770912. PMID: 8770912.

44. Nugent C, Prins JB, Whitehead JP, Wentworth JM, Chatterjee VK, et al. Arachidonic acid stimulates glucose uptake in 3T3-L1 adipocytes by increasing GLUT1 and GLUT4 levels at the plasma membrane. Evidence for involvement of lipoxygenase metabolites and peroxisome proliferator-activated receptor gamma. J Biol Chem. 2001 Mar 23;276(12):9149–57. doi: 10.1074/jbc.M009817200. Epub 2000 Dec 21. PMID: 11124961.

45. Wolford JK, Konheim YL, Colligan PB, Bogardus C. Association of a F479L variant in the cytosolic phospholipase A2 gene (PLA2G4A) with decreased glucose turnover and oxidation rates in Pima Indians. Mol Genet Metab. 2003 May;79(1):61–6. doi: 10.1016/s1096-7192(03)00051-9. PMID: 12765847.

46. Murakami M, Taketomi Y, Miki Y, Sato H, Hirabayashi T, et al. Recent progress in phospholipase A₂ research: from cells to animals to humans. Prog Lipid Res. 2011 Apr;50(2):152–92. doi: 10.1016/j.plipres.2010.12.001. Epub 2010 Dec 24. PMID: 21185866.

47. Manolio TA, Collins FS, Cox NJ, Goldstein DB, Hindorff LA, et al. Finding the missing heritability of complex diseases. Nature. 2009 Oct 8;461(7265):747–53. doi: 10.1038/nature08494. PMID: 19812666.

48. McCarthy MI, Hirschhorn JN. Genome-wide association studies: potential next steps on a genetic journey. Hum Mol Genet. 2008 Oct 15;17(R2):R156–65. doi: 10.1093/hmg/ddn289. PMID: 18852205.

49. 1000 Genomes Project Consortium; Abecasis GR, Altshuler D, Auton A, Brooks LD, et al. A map of human genome variation from population-scale sequencing. Nature. 2010 Oct 28;467(7319):1061–73. doi: 10.1038/nature09534.

50. 1000 Genomes Project Consortium; Abecasis GR, Auton A, Brooks LD, DePristo MA, et al. An integrated map of genetic variation from 1,092 human genomes. Nature 2012 Nov 1;491(7422):56–65. doi: 10.1038/nature11632. PMID: 23128226.

51. Bodmer W, Bonilla C. Common and rare variants in multifactorial susceptibility to common diseases. Nat Genet. 2008 Jun;40(6):695–701. doi: 10.1038/ng.f.136. PMID: 18509313.

52. Kathiresan S, Willer CJ, Peloso GM, Demissie S, Musunuru K, et al. Common variants at 30 loci contribute to polygenic dyslipidemia. Nat Genet. 2009 Jan;41(1):56–65. doi: 10.1038/ng.291. Epub 2008 Dec 7. PMID: 19060906.

53. Lusis AJ, Pajukanta P. A treasure trove for lipoprotein biology. Nat Genet. 2008 Feb;40(2):129–30. doi: 10.1038/ng0208-129. PMID: 18227868.

54. Global Burden of Disease Collaborative Network. Global Burden of Disease Study 2019 (GBD 2019) Results. Seattle, United States: Institute for Health Metrics and Evaluation (IHME), 2020. URL: https://vizhub.healthdata.org/gbd-results/. Date accessed: November 2021

55. NCD Risk Factor Collaboration (NCD-RisC). Worldwide trends in blood pressure from 1975 to 2015: a pooled analysis of 1479 population-based measurement studies with 19·1 million participants. Lancet. 2017 Jan 7;389(10064):37–55. doi: 10.1016/S0140-6736(16)31919-5. Epub 2016 Nov 16.

56. NCD Risk Factor Collaboration (NCD-RisC). Worldwide trends in body-mass index, underweight, overweight, and obesity from 1975 to 2016: a pooled analysis of 2416 population-based measurement studies in 128·9 million children, adolescents, and adults. Lancet. 2017 Dec 16;390(10113):2627–2642. doi: 10.1016/S0140-6736(17)32129-3. Epub 2017 Oct 10. PMID: 29029897; PMCID: PMC5735219.

57. R Core Team (2021). R: A language and environment for statistical computing. R Foundation for Statistical Computing, Vienna, Austria. URL: https://www.R-project.org/.

58. Clyde, Merlise (2022) BAS: Bayesian Variable Selection and Model Averaging using Bayesian Adaptive Sampling, R package version 1.6.4.

59. Laversanne M, Vignat J (2020). Rcan: Cancer Registry Data Analysis and Visualisation. R package version 1.3.82, URL: https://CRAN.R-project.org/package=Rcan.

60. Taiyun Wei and Viliam Simko (2021). ’corrplot’: Visualization of a Correlation Matrix, R package version 0.92. URL: https://github.com/taiyun/corrplot.

61. Pritchard JK, Stephens M, Donnelly P. Inference of population structure using multilocus genotype data. Genetics. 2000 Jun;155(2):945–59. doi: 10.1093/genetics/155.2.945. PMID: 10835412; PMCID: PMC1461096.

62. PLINK [v1.9], Shaun Purcell, Christopher Chang, URL: www.cog-genomics.org/plink/1.9/

63. Chang CC, Chow CC, Tellier LC, Vattikuti S, Purcell SM, et al. Second-generation PLINK: rising to the challenge of larger and richer datasets. Gigascience. 2015 Feb 25;4:7. doi: 10.1186/s13742-015-0047-8. PMID: 25722852; PMCID: PMC4342193.

64. Machiela MJ, Chanock SJ. LDlink: a web-based application for exploring population-specific haplotype structure and linking correlated alleles of possible functional variants. Bioinformatics. 2015 Nov 1;31(21):3555–7. doi: 10.1093/bioinformatics/btv402. Epub 2015 Jul 2. PMID: 26139635; PMCID: PMC4626747.

65. Lewontin RC. The Interaction of Selection and Linkage. I. General Considerations; Heterotic Models. Genetics. 1964 Jan;49(1):49–67. doi: 10.1093/genetics/49.1.49. PMID: 17248194; PMCID: PMC1210557.

66. Perdry H, Dandine-Roulland C (2022). gaston: Genetic Data Handling (QC, GRM, LD, PCA) & Linear Mixed Models. R package version 1.5.9, URL: https://CRAN.R-project.org/package=gaston.

