## Supplementary file 1 for "Ethnic and region-specific genetic risk variants of stroke and its comorbid conditions can define the variations in the burden of stroke and its phenotypic traits"

**Table S1. Global incidence, mortality and prevalence rates for Stroke, its subtypes and comorbid factors.** The crude and age-standardized rates for (a) incidence, (b) mortality and (c) prevalence for the global region Stroke, its subtypes and comorbid factors in 2009, 2014 and 2019.

| **a. Global crude and age-standardized incidence rates for Stroke, its subtypes and comorbid factors in 2009, 2014 and 2019.** | | | | | | |
| --- | --- | --- | --- | --- | --- | --- |
| **Disease** | **Crude Incidence Rate per 100,000 (95% uncertainty interval)** | | | **Age-standardized Incidence Rate per 100,000 (95% uncertainty interval)** | | |
|  | **2009** | **2014** | **2019** | **2009** | **2014** | **2019** |
| High SBP | - | - | - | - | - | - |
| Ischemic Heart Disease | 242.09 (215.97,  268.51) | 257.79 (232.03,  284.56) | 274.04 (242.96,  306.36) | 274.95 (246.52,  305.45) | 269.16 (243.38,  296.56) | 262.39 (233.25,  293.26) |
| Stroke | 139.55 (128.19,  152.97) | 146.23 (133.59,  160.82) | 157.99 (142.71,  175.63) | 154.41 (141.63,  168.98) | 150.46 (137.65,  165.65) | 150.77 (136.52,  167.46) |
| High BMI | - | - | - | - | - | - |
| High LDL | - | - | - | - | - | - |
| Ischemic stroke | 81.77 (71.5,93.9) | 88.63 (77.3,102.2) | 98.62 (84.9,115.8) | 91.87 (80.4,105.4) | 92.17 (80.5,106.3) | 94.51 (81.9,110.8) |
| Intracerebral hemorrhage | 43.59 (38.7,49.2) | 43.16 (38.41,48.6) | 44.06 (38.39,50.5) | 47.36 (41.9,53.49) | 43.78 (38.87,49.3) | 41.81 (36.5,47.88) |
| Type 2 Diabetes | 234.22 (219.82,  249.47) | 257.42 (241.01,  274.98) | 280.07 (258.75,  303.89) | 236.01 (221.74,  251.19) | 247.96 (232.5,  264.75) | 259.94 (240.35,  281.44) |
| Chronic kidney disease | 199.1 (184.44,  215.14) | 220.52 (204.21,  238.1) | 245.39 (226.9,  265.18) | 219.29 (203.34,  236.21) | 226.42 (209.55,  244.19) | 233.65 (216.56,  252.31) |
| Subarachnoid hemorrhage | 14.2 (12.3,16.4) | 14.44 (12.5,16.73) | 15.31 (13,17.97) | 15.17 (13.18,17.5) | 14.51 (12.6,16.75) | 14.46 (12.3,16.94) |
| Type 1 diabetes | 6.82 (5.6,8.32) | 6.94 (5.67,8.48) | 7.36 (5.99,9.01) | 6.83 (5.62,8.32) | 7.06 (5.78,8.63) | 7.6 (6.18,9.32) |

| **b. Global crude and age-standardized mortality rates for Stroke, its subtypes and comorbid factors in 2009, 2014 and 2019.** | | | | | | |
| --- | --- | --- | --- | --- | --- | --- |
| **Disease** | **Crude Mortality Rate per 100,000**  **(95% uncertainty interval)** | | | **Age-standardized Mortality Rate per 100,000 (95% uncertainty interval)** | | |
|  | **2009** | **2014** | **2019** | **2009** | **2014** | **2019** |
| High SBP | 130.23 (115.66,  144.51) | 133.3 (118.27,  147.5) | 140.17 (122.96,  156.78) | 156.22 (137.3,  174.67) | 145.74 (128.45,  162.06) | 138.88 (121.25,  155.73) |
| Ischemic Heart Disease | 108.46 (101.92,  113.35) | 111.51 (104.09,  116.59) | 118.1 (108.51,  125.93) | 131.77 (122.35,  138.33) | 123.16 (113.8,  129.11) | 117.95 (107.83,  125.92) |
| Stroke | 83.62 (78.3,87.45) | 81.49 (76.06,85.6) | 84.69 (77.48,90.7) | 100.5 (93.2,105.4) | 89.29 (82.44,93.9) | 84.19 (76.76,90.15) |
| High BMI | 53.52 (32.8,77.72) | 58.02 (36.4,83.35) | 64.87 (41.66,91.9) | 61.66 (37.4,90.18) | 61.36 (38.1,89.03) | 62.59 (39.92,89.13) |
| High LDL | 53.17 (40.66,67.3) | 54.04 (41.3,68.89) | 56.83 (42.7,73.04) | 63.9 (47.7,82.87) | 59.07 (44.01,76.9) | 56.51 (41.83,73.62) |
| Ischemic stroke | 38.94 (35.78,41.3) | 39.56 (36.1,42.06) | 42.56 (38.43,45.7) | 49.24 (44.75,52.2) | 45.1 (40.84,47.9) | 43.5 (39.08,46.77) |
| Intracerebral hemorrhage | 39.92 (37.6,42.09) | 37.19 (34.97,39.3) | 37.3 (34.2,40.06) | 45.92 (43.07,48.5) | 39.23 (36.75,41.5) | 36.04 (32.98,38.67) |
| Type 2 Diabetes | 15.58 (14.7,16.18) | 17.07 (16.04,17.8) | 19.04 (17.7,20.24) | 18.23 (17.1,18.97) | 18.24 (16.9,19.02) | 18.49 (17.18,19.66) |
| Chronic kidney disease | 15.57 (14.6,16.22) | 17.04 (15.87,17.8) | 18.45 (16.98,19.7) | 18.41 (17.1,19.22) | 18.56 (17.16,19.5) | 18.29 (16.72,19.55) |
| Subarachnoid hemorrhage | 4.75 (4.05,5.56) | 4.75 (4.14,5.39) | 4.82 (4.27,5.38) | 5.34 (4.57,6.2) | 4.96 (4.34,5.61) | 4.66 (4.13,5.17) |
| Type 1 diabetes | 0.99 (0.84,1.16) | 0.98 (0.84,1.16) | 1.01 (0.88,1.21) | 1.03 (0.88,1.22) | 0.98 (0.84,1.17) | 0.98 (0.85,1.17) |

| **c. Global crude and age-standardized prevalence rates for Stroke, its subtypes and comorbid factors in 2009, 2014 and 2019.** | | | | | | |
| --- | --- | --- | --- | --- | --- | --- |
| **Disease** | **Crude Prevalence Rate per 100,000 (95% uncertainty interval)** | | | **Age-standardized Prevalence Rate per 100,000 (95%** **uncertainty interval)** | | |
|  | **2009** | **2014** | **2019** | **2009** | **2014** | **2019** |
| High SBP | - | - | - | 23078.9 (21431.1,  24774.7) | 22241.3 (19927.5,  24730.8) | - |
| Ischemic Heart Disease | 2156.58 (1949.69,  2395.37) | 2341.63 (2124.85,  2596.83) | 2548.89 (2296.47,  2836.86) | 2422.21 (2185.13,  2688.5) | 2425.63 (2202.93,  2689.22) | 2421.02 (2180.5,  2692.65) |
| Stroke | 1128.24 (1042.55,  1213.15) | 1198.27 (1118.54,  1278.91) | 1311.47 (1204.68,  1428.46) | 1217.36 (1126.81,  1313.52) | 1213.25 (1133.43,  1298.29) | 1240.26 (1139.71,  1352.99) |
| High BMI | - | - | - | 11360.9 (10745.2,  11989.4) | 12932.5 (12055.8,  13852.7) | - |
| High LDL | - | - | - | - | - | - |
| Ischemic stroke | 837.47 (756.23,  923.39) | 901.26 (820.79,  982.86) | 997.65 (889.92,  1117.39) | 916.46 (827.21,  1011.45) | 923.67 (840.53,  1005.76) | 950.97 (849.82,  1064.06) |
| Intracerebral hemorrhage | 248.13 (217.4,  279.92) | 252.48 (226.3,  280.51) | 267.06 (232.84,  302.66) | 257.65 (226.36,  289.73) | 247.88 (222.07,  274.81) | 248.77 (217.09,  281.43) |
| Type 2 Diabetes | 4372.47 (4084.66,  4674.09) | 5034.53 (4692.9,  5390.88) | 5659.56 (5196.06,  6165.05) | 4646.97 (4341.45,  4960.45) | 5007.94 (4674.29,  5361.89) | 5282.85 (4853.59,  5752.09) |
| Chronic kidney disease | 7905.83 (7355.51,  8435.14) | 8487.63 (7905.89,  9019.4) | 9011.92 (8401.27,  9577.82) | 8340.38 (7792.87,  8846.68) | 8519.77 (7956.29,  9037.24) | 8596.21 (8015.74,  9125.28) |
| Subarachnoid hemorrhage | 99.5 (85.71,  115.56) | 103.7 (90.52,  119.03) | 108.52 (92.9,  127.09) | 103.6 (89.2,  120.53) | 102.57 (89.53,  117.81) | 101.57 (87.13,  118.54) |
| Type 1 diabetes | 249.48 (201.82,  306.1) | 259.98 (208.66,  320.11) | 283.93 (225.92,  349.86) | 250.88 (203.63,  307.25) | 255.31 (205.21,  313.96) | 272.54 (216.98,  336.95) |
