## Supplementary file 2 for "Ethnic and region-specific genetic risk variants of stroke and its comorbid conditions can define the variations in the burden of stroke and its phenotypic traits"

**Table S2. Spatio-temporal comparison of mortality rates of Stroke, its subtypes and comorbid factors.** The 2019 ASMR for stroke, its subtypes and its comorbid factors in each region was compared with (a) global mortality rates and (b) mortality rates in 2009. CKD – chronic kidney disease, ICH – intracerebral hemorrhage, IHD – ischemic heart disease, IS – ischemic stroke, SAH – subarachnoid hemorrhage, T1D – type 1 diabetes, T2D – type 2 diabetes.

| a. The 2019 ASMR of each disease in a region was compared with the global 2019 ASMR and p-value obtained using chi-square test. The variables are assumed to be in Poisson distribution. | | | | | | | | | | | | | | | | |
| --- | --- | --- | --- | --- | --- | --- | --- | --- | --- | --- | --- | --- | --- | --- | --- | --- |
| Region | | | **CKD** | | | **P-value** | | **High BMI** | | | **P-value** | | **High LDL** | | | **P-value** |
| Global | | | 18.29 | | |  | | 62.59 | | |  | | 56.51 | | |  |
| Africa | | | 30.58 | | | 0.004051 | | 98.83 | | | <0.001 | | 64.39 | | | 0.2948 |
| Central and South Asia | | | 22.74 | | | 0.2975 | | 62.07 | | | 0.9468 | | 68.91 | | | 0.09916 |
| East Asia | | | 15.80 | | | 0.5599 | | 46.41 | | | 0.04079 | | 53.44 | | | 0.6825 |
| Europe | | | 8.18 | | | 0.01808 | | 73.34 | | | 0.1745 | | 75.53 | | | 0.01144 |
| Middle East | | | 28.68 | | | 0.01515 | | 120.14 | | | <0.001 | | 94.43 | | | <0.001 |
| Oceania | | | 13.71 | | | 0.284 | | 67.52 | | | 0.5337 | | 48.92 | | | 0.3122 |
| America | | | 24.95 | | | 0.1195 | | 72.83 | | | 0.1958 | | 40.44 | | | 0.03248 |
| Region | | | **High SBP** | | | **P-value** | | **ICH** | | | **P-value** | | **IHD** | | | **P-value** |
| Global | | | 138.88 | | |  | | 36.04 | | |  | | 117.95 | | |  |
| Africa | | | 206.18 | | | <0.001 | | 49.82 | | | 0.02168 | | 154.04 | | | <0.001 |
| Central and South Asia | | | 167.69 | | | 0.01449 | | 41.45 | | | 0.3674 | | 167.39 | | | <0.001 |
| East Asia | | | 160.36 | | | 0.06838 | | 61.20 | | | <0.001 | | 110.93 | | | 0.518 |
| Europe | | | 138.88 | | | 0.9999 | | 15.70 | | | <0.001*** | | 138.19 | | | 0.0624 |
| Middle East | | | 198.61 | | | <0.001 | | 24.94 | | | 0.06453 | | 197.40 | | | <0.001 |
| Oceania | | | 94.63 | | | <0.001*** | | 34.32 | | | 0.775 | | 100.75 | | | 0.1133 |
| America | | | 93.91 | | | <0.001 | | 14.06 | | | <0.001 | | 87.51 | | | 0.005066 |
| Region | | | **IS** | | | **P-value** | | **SAH** | | | **P-value** | | **Stroke** | | | **P-value** |
| Global | | | 43.50 | | |  | | 4.66 | | |  | | 84.19 | | |  |
| Africa | | | 52.05 | | | 0.1945 | | 2.73 | | | 0.663 | | 104.60 | | | 0.02613 |
| Central and South Asia | | | 39.20 | | | 0.5152 | | 5.77 | | | 0.4775 | | 86.42 | | | 0.8081 |
| East Asia | | | 61.33 | | | 0.006866 | | 5.14 | | | 0.8247 | | 127.66 | | | <0.001 |
| Europe | | | 50.36 | | | 0.2978 | | 4.16 | | | 0.8188 | | 70.22 | | | 0.128 |
| Middle East | | | 61.43 | | | 0.006551 | | 2.91 | | | 0.6203 | | 89.28 | | | 0.579 |
| Oceania | | | 22.56 | | | 0.001502 | | 5.27 | | | 0.7775 | | 62.15 | | | 0.0163 |
| America | | | 21.69 | | | <0.001 | | 4.57 | | | 0.1221 | | 40.33 | | | <0.001 |
| Region | | | **T1D** | | | **P-value** | | **T2D** | | | **P-value** | |  | | |  |
| Global | | | 0.98 | | |  | | 18.49 | | |  | |  | | |  |
| Africa | | | 1.59 | | | 0.7342 | | 36.45 | | | <0.001 | |  | | |  |
| Central and South Asia | | | 1.34 | | | 0.7105 | | 29.61 | | | 0.009683 | |  | | |  |
| East Asia | | | 0.91 | | | 0.9452 | | 17.45 | | | 0.8093 | |  | | |  |
| Europe | | | 0.51 | | | 0.6404 | | 8.26 | | | 0.01733 | |  | | |  |
| Middle East | | | 1.05 | | | 0.9416 | | 27.43 | | | 0.03761 | |  | | |  |
| Oceania | | | 1.06 | | | 0.9317 | | 41.29 | | | <0.001 | |  | | |  |
| America | | | 1.04 | | | 0.9501 | | 22.55 | | | 0.3454 | |  | | |  |
|  | b. The 2019 ASMR of each disease in a region was compared with the 2009 ASMR in the region and p-value obtained using chi-square test. The variables are assumed to be in Poisson distribution. | | | | | | | | | | | | | | | |
|  |  | **CKD** | | | | | **High BMI** | | | | | **High LDL** | | | | |
|  | **Region** | **2019** | | **2009** | **P-value** | | **2019** | | **2009** | **P-value** | | **2019** | | **2009** | **P-value** | |
|  | Global | 18.29 | | 18.41 | 0.9778 | | 62.59 | | 61.66 | 0.9051 | | 56.51 | | 63.90 | 0.3259 | |
|  | Africa | 30.58 | | 31.18 | 0.9136 | | 98.83 | | 91.64 | 0.4527 | | 64.39 | | 67.15 | 0.7309 | |
|  | Central & South Asia | 22.74 | | 23.72 | 0.838 | | 62.07 | | 51.31 | 0.1334 | | 68.91 | | 72.30 | 0.683 | |
|  | East Asia | 15.80 | | 16.22 | 0.9158 | | 46.41 | | 39.76 | 0.2916 | | 53.44 | | 56.11 | 0.7151 | |
|  | Europe | 8.18 | | 8.19 | 0.9962 | | 73.34 | | 84.45 | 0.2264 | | 75.53 | | 96.66 | 0.03157 | |
|  | Middle East | 28.68 | | 29.75 | 0.8413 | | 120.14 | | 122.60 | 0.8228 | | 94.43 | | 107.43 | 0.181 | |
|  | Oceania | 13.71 | | 13.60 | 0.977 | | 67.52 | | 69.20 | 0.8395 | | 48.92 | | 53.34 | 0.5451 | |
|  | America | 24.95 | | 23.83 | 0.819 | | 72.83 | | 73.34 | 0.9521 | | 40.44 | | 45.86 | 0.3938 | |
|  |  | **High SBP** | | | | | **ICH** | | | | | **IHD** | | | | |
|  | **Region** | **2019** | | **2009** | **P-value** | | **2019** | | **2009** | **P-value** | | **2019** | | **2009** | **P-value** | |
|  | Global | 138.88 | | 156.22 | 0.1411 | | 36.04 | | 45.92 | 0.09968 | | 117.95 | | 131.77 | 0.2288 | |
|  | Africa | 206.18 | | 221.26 | 0.2935 | | 49.82 | | 58.06 | 0.2427 | | 154.04 | | 162.34 | 0.5037 | |
|  | Central & South Asia | 167.69 | | 183.48 | 0.2229 | | 41.45 | | 51.57 | 0.1159 | | 167.39 | | 176.75 | 0.4692 | |
|  | East Asia | 160.36 | | 176.40 | 0.2051 | | 61.20 | | 85.51 | 0.00854 | | 110.93 | | 117.17 | 0.5539 | |
|  | Europe | 138.88 | | 173.49 | 0.0086 | | 15.70 | | 20.31 | 0.3061 | | 138.19 | | 174.31 | 0.00622 | |
|  | Middle East | 198.61 | | 222.34 | 0.09216 | | 24.94 | | 31.66 | 0.1782 | | 197.40 | | 221.61 | 0.08477 | |
|  | Oceania | 94.63 | | 102.14 | 0.4574 | | 34.32 | | 34.25 | 0.9901 | | 100.75 | | 109.14 | 0.4221 | |
|  | America | 93.91 | | 101.20 | 0.4516 | | 14.06 | | 15.95 | 0.6142 | | 87.51 | | 97.50 | 0.2858 | |
|  |  | **IS** | | | | | **SAH** | | | | | **Stroke** | | | | |
|  | **Region** | **2019** | | **2009** | **P-value** | | **2019** | | **2009** | **P-value** | | **2019** | | **2009** | **P-value** | |
|  | Global | 43.50 | | 49.24 | 0.3836 | | 4.66 | | 5.34 | 0.751 | | 84.19 | | 100.50 | 0.07539 | |
|  | Africa | 52.05 | | 54.74 | 0.71 | | 2.73 | | 3.27 | 0.7428 | | 104.60 | | 116.07 | 0.2621 | |
|  | Central & South Asia | 39.20 | | 44.32 | 0.4143 | | 5.77 | | 7.15 | 0.5641 | | 86.42 | | 103.04 | 0.07384 | |
|  | East Asia | 61.33 | | 64.96 | 0.643 | | 5.14 | | 6.05 | 0.6866 | | 127.66 | | 156.52 | 0.02105 | |
|  | Europe | 50.36 | | 65.67 | 0.05896 | | 4.16 | | 4.84 | 0.7583 | | 70.22 | | 90.82 | 0.0307 | |
|  | Middle East | 61.43 | | 68.72 | 0.3525 | | 2.91 | | 3.74 | 0.6273 | | 89.28 | | 104.12 | 0.1163 | |
|  | Oceania | 22.56 | | 24.92 | 0.6362 | | 5.27 | | 5.61 | 0.8858 | | 62.15 | | 64.78 | 0.7439 | |
|  | America | 21.69 | | 23.96 | 0.6261 | | 4.57 | | 4.81 | 0.912 | | 40.33 | | 44.72 | 0.4887 | |
|  |  | **T1D** | | | | | **T2D** | | | | |  | | | | |
|  | **Region** | **2019** | | **2009** | **P-value** | | **2019** | | **2009** | **P-value** | |  | |  |  | |
|  | Global | 0.98 | | 1.03 | 0.9554 | | 18.49 | | 18.23 | 0.9513 | |  | |  |  | |
|  | Africa | 1.59 | | 1.69 | 0.9319 | | 36.45 | | 37.50 | 0.8615 | |  | |  |  | |
|  | Central & South Asia | 1.34 | | 1.40 | 0.9591 | | 29.61 | | 28.54 | 0.8405 | |  | |  |  | |
|  | East Asia | 0.91 | | 0.98 | 0.9416 | | 17.45 | | 16.52 | 0.819 | |  | |  |  | |
|  | Europe | 0.51 | | 0.59 | 0.9228 | | 8.26 | | 8.46 | 0.9455 | |  | |  |  | |
|  | Middle East | 1.05 | | 1.27 | 0.8297 | | 27.43 | | 28.38 | 0.856 | |  | |  |  | |
|  | Oceania | 1.08 | | 1.08 | 0.9993 | | 43.95 | | 42.38 | 0.8091 | |  | |  |  | |
|  | America | 1.04 | | 1.10 | 0.9513 | | 22.55 | | 22.89 | 0.9421 | |  | |  |  | |

**Table S3. Spatio-temporal** **comparison of prevalence rates of Stroke, its subtypes and comorbid factors.** The 2019 ASPR for stroke, its subtypes and its comorbid factors in each region was compared with (a) global prevalence rates and (b) prevalence rates in 2009. Where 2019 rates were not available, 2014 rates were used instead. CKD–chronic kidney disease, ICH–intracerebral hemorrhage, IHD–ischemic heart disease, IS–ischemic stroke, SAH–subarachnoid hemorrhage, T1D–type 1 diabetes, T2D– type 2 diabetes.

| a. The 2019 ASPR of each disease in a region was compared with the global 2019 ASPR and p-value obtained using chi-square test. The variables are assumed to be in Poisson distribution. | | | | | | | | | | | | | | | |
| --- | --- | --- | --- | --- | --- | --- | --- | --- | --- | --- | --- | --- | --- | --- | --- |
| Region | | **CKD** | | **P-value** | | | **High BMI** | | **P-value** | | | **High LDL** | | **P-value** | |
| Global | | 8596.21 | |  | | | 12932.47 | |  | | | Data NA | | | |
| Africa | | 7548.86 | | <0.001 | | | 11699.59 | | <0.001 | | |  |  |  |  |
| Central and South Asia | | 8900.16 | | 0.001044 | | | 4691.78 | | <0.001 | | |  |  |  |  |
| East Asia | | 8868.46 | | 0.003321 | | | 5822.84 | | <0.001 | | |  |  |  |  |
| Europe | | 7277.72 | | <0.001 | | | 22882.85 | | <0.001 | | |  |  |  |  |
| Middle East | | 10589.31 | | <0.001 | | | 28526.37 | | <0.001 | | |  |  |  |  |
| Oceania | | 7282.16 | | <0.001 | | | 27527.29 | | <0.001 | | |  |  |  |  |
| America | | 8795.43 | | 0.03166 | | | 28064.95 | | <0.001 | | |  |  |  |  |
| Region | | **High SBP** | | **P-value** | | | **ICH** | | **P-value** | | | **IHD** | | **P-value** | |
| Global | | 22241.26 | |  | | | 248.77 | |  | | | 2421.02 | |  | |
| Africa | | 27382.96 | | <0.001 | | | 314.25 | | <0.001 | | | 2819.42 | | <0.001 | |
| Central and South Asia | | 26232.76 | | <0.001 | | | 267.03 | | 0.2469 | | | 3577.70 | | <0.001 | |
| East Asia | | 20171.79 | | <0.001 | | | 302.65 | | <0.001 | | | 2052.09 | | <0.001 | |
| Europe | | 23696.26 | | <0.001 | | | 139.23 | | <0.001 | | | 2476.74 | | 0.2574 | |
| Middle East | | 22561.63 | | 0.0317 | | | 255.65 | | 0.6627 | | | 4843.02 | | <0.001 | |
| Oceania | | 18302.80 | | <0.001 | | | 264.45 | | 0.3202 | | | 2734.98 | | <0.001 | |
| America | | 18273.49 | | <0.001 | | | 200.52 | | 0.002218 | | | 1695.56 | | <0.001 | |
| Region | | **IS** | | **P-value** | | | **SAH** | | **P-value** | | | **Stroke** | | **P-value** | |
| Global | | 950.97 | |  | | | 101.57 | |  | | | 1240.26 | |  | |
| Africa | | 1089.33 | | <0.001 | | | 52.47 | | <0.001 | | | 1393.27 | | <0.001 | |
| Central and South Asia | | 570.43 | | <0.001 | | | 65.01 | | <0.001 | | | 858.50 | | <0.001 | |
| East Asia | | 1183.05 | | <0.001 | | | 107.87 | | 0.5319 | | | 1513.10 | | <0.001 | |
| Europe | | 753.15 | | <0.001 | | | 118.34 | | 0.09603 | | | 960.86 | | <0.001 | |
| Middle East | | 1218.88 | | <0.001 | | | 66.69 | | <0.001 | | | 1471.60 | | <0.001 | |
| Oceania | | 650.92 | | <0.001 | | | 105.79 | | 0.6752 | | | 970.31 | | <0.001 | |
| America | | 932.69 | | 0.5532 | | | 138.90 | | <0.001 | | | 1215.60 | | 0.4838 | |
| Region | | **T1D** | | **P-value** | | | **T2D** | | **P-value** | | |  | | | |
| Global | | 272.54 | |  | | | 5282.85 | |  | | |  |  |  |  |
| Africa | | 255.51 | | 0.3023 | | | 4516.30 | | <0.001 | | |  |  |  |  |
| Central and South Asia | | 250.87 | | 0.1893 | | | 6401.50 | | <0.001 | | |  |  |  |  |
| East Asia | | 132.14 | | <0.001 | | | 4477.09 | | <0.001 | | |  |  |  |  |
| Europe | | 499.90 | | <0.001 | | | 4691.56 | | <0.001 | | |  |  |  |  |
| Middle East | | 394.02 | | <0.001 | | | 6796.80 | | <0.001 | | |  |  |  |  |
| Oceania | | 459.51 | | <0.001 | | | 5606.60 | | <0.001 | | |  |  |  |  |
| America | | 417.47 | | <0.001 | | | 6563.01 | | <0.001 | | |  |  |  |  |
| b. The 2019 (or 2014) ASPR of each disease in a region was compared with the 2009 ASPR in the region and p-value obtained using chi-square test. The variables are assumed to be in Poisson distribution. | | | | | | | | | | | | | | | |
|  | **CKD** | | | | | **High BMI** | | | | | **High LDL** | | | | |
| **Region** | **2019** | | **2009** | | **P-value** | **2014** | | **2009** | | **P-value** | **Data NA** | | | | |
| Global | 8596.2 | | 8340.4 | | 0.00508 | 12932.4 | | 11360.9 | | <0.001 |  | |  | |  |
| Africa | 7548.9 | | 7037.3 | | <0.001 | 11699.5 | | 10068.7 | | <0.001 |  | |  | |  |
| Central & South Asia | 8900.2 | | 8656.8 | | 0.00889 | 4691.77 | | 3639.98 | | <0.001 |  | |  | |  |
| East Asia | 8868.5 | | 8655.3 | | 0.02196 | 5822.84 | | 4416.30 | | <0.001 |  | |  | |  |
| Europe | 7277.7 | | 7096.9 | | 0.03181 | 22882.8 | | 20946.0 | | <0.001 |  | |  | |  |
| Middle East | 10589.3 | | 9904.1 | | <0.001 | 28526.3 | | 25320.1 | | <0.001 |  | |  | |  |
| Oceania | 7282.2 | | 7025.9 | | 0.00223 | 27527.2 | | 24822.0 | | <0.001 |  | |  | |  |
| America | 8795.4 | | 8394.1 | | <0.001 | 28064.9 | | 25317.6 | | <0.001 |  | |  | |  |
|  | **High SBP** | | | | | **ICH** | | | | | **IHD** | | | | |
| **Region** | **2014** | | **2009** | | **P-value** | **2019** | | **2009** | | **P-value** | **2019** | | **2009** | | **P-value** |
| Global | 22241.2 | | 23078.9 | | <0.001 | 248.8 | | 257.6 | | 0.5802 | 2421.0 | | 2422.2 | | 0.9807 |
| Africa | 27382.9 | | 27910.6 | | 0.00158 | 314.2 | | 340.2 | | 0.1424 | 2819.4 | | 2814.8 | | 0.9302 |
| Central & South Asia | 26232.7 | | 26062.0 | | 0.2903 | 267.0 | | 268.3 | | 0.9373 | 3577.7 | | 3510.5 | | 0.2568 |
| East Asia | 20171.7 | | 20935.6 | | <0.001 | 302.7 | | 319.2 | | 0.3402 | 2052.1 | | 2057.0 | | 0.9142 |
| Europe | 23696.2 | | 25915.0 | | <0.001 | 139.2 | | 158.6 | | 0.1011 | 2476.7 | | 2548.3 | | 0.1504 |
| Middle East | 22561.6 | | 24327.7 | | <0.001 | 255.6 | | 268.5 | | 0.4324 | 4843.0 | | 4949.3 | | 0.1307 |
| Oceania | 18302.7 | | 19610.2 | | <0.001 | 264.5 | | 258.3 | | 0.7021 | 2735.0 | | 2843.9 | | 0.04106 |
| America | 18273.4 | | 19505.6 | | <0.001 | 200.5 | | 212.0 | | 0.4302 | 1695.6 | | 1771.0 | | 0.07318 |
|  | **IS** | | | | | **SAH** | | | | | **Stroke** | | | | |
| **Region** | **2019** | | **2009** | | **P-value** | **2019** | | **2009** | | **P-value** | **2019** | | **2009** | | **P-value** |
| Global | 951.0 | | 916.5 | | 0.2543 | 101.6 | | 103.6 | | 0.8419 | 1240.3 | | 1217.4 | | 0.5116 |
| Africa | 1089.3 | | 1071.4 | | 0.5838 | 52.5 | | 53.6 | | 0.8786 | 1393.3 | | 1402.2 | | 0.8115 |
| Central & South Asia | 570.4 | | 533.4 | | 0.1086 | 65.0 | | 64.8 | | 0.9743 | 858.5 | | 822.7 | | 0.2116 |
| East Asia | 1183.1 | | 1074.9 | | <0.001 | 107.9 | | 107.0 | | 0.9355 | 1513.1 | | 1425.4 | | 0.02018 |
| Europe | 753.1 | | 813.8 | | 0.0271 | 118.3 | | 124.2 | | 0.5874 | 960.9 | | 1042.9 | | 0.00813 |
| Middle East | 1218.9 | | 1220.3 | | 0.9686 | 66.7 | | 67.2 | | 0.9507 | 1471.6 | | 1485.6 | | 0.7164 |
| Oceania | 650.9 | | 655.3 | | 0.8645 | 105.8 | | 104.7 | | 0.9119 | 970.3 | | 967.2 | | 0.9215 |
| America | 932.7 | | 977.1 | | 0.1457 | 138.9 | | 127.3 | | 0.3038 | 1215.6 | | 1259.3 | | 0.2187 |
|  | **T1D** | | | | | **T2D** | | | | |  | | | | |
| **Region** | **2019** | | **2009** | | **P-value** | **2019** | | **2009** | | **P-value** |  | | | | |
| Global | 272.5 | | 250.9 | | 0.1714 | 5282.9 | | 4647.0 | | <0.001 |  |  |  |  |  |
| Africa | 255.5 | | 227.8 | | 0.06632 | 4516.3 | | 3939.5 | | <0.001 |  |  |  |  |  |
| Central & South Asia | 250.9 | | 222.7 | | 0.05885 | 6401.5 | | 5054.2 | | <0.001 |  |  |  |  |  |
| East Asia | 132.1 | | 117.0 | | 0.1622 | 4477.1 | | 4237.1 | | <0.001 |  |  |  |  |  |
| Europe | 499.9 | | 458.6 | | 0.05367 | 4691.6 | | 4044.2 | | <0.001 |  |  |  |  |  |
| Middle East | 394.0 | | 297.0 | | <0.001 | 6796.8 | | 5937.9 | | <0.001 |  |  |  |  |  |
| Oceania | 459.5 | | 441.0 | | 0.3785 | 5606.6 | | 4732.1 | | <0.001 |  |  |  |  |  |
| America | 417.5 | | 408.2 | | 0.647 | 6563.0 | | 5862.8 | | <0.001 |  |  |  |  |  |
