## Supplementary file 3 for "Ethnic and region-specific genetic risk variants of stroke and its comorbid conditions can define the variations in the burden of stroke and its phenotypic traits"

**Table S4. Comparison of proportional mortality of Stroke, HSBP and Metabolic conditions.** The proportional mortality for stroke, HSBP and metabolic conditions in each region was compared with global proportional mortality as well as regional proportional mortality in a pairwise manner for 2009 and 2014. P-value (2-sided) obtained using one-sample test for binomial proportion using normal-theory method.

| **P-values for metabolic proportional mortality in 2009** | | | | | | | | |
| --- | --- | --- | --- | --- | --- | --- | --- | --- |
|  | Proportional mortality | **Global** | **AFR** | **CSA** | **EAS** | **EUR** | **MDE** | **OCE** |
| **Global** | 29.58 |  |  |  |  |  |  |  |
| **Africa** | 31.44 | 0.6839 |  |  |  |  |  |  |
| **Central & South Asia** | 27.68 | 0.6758 | 0.4172 |  |  |  |  |  |
| **East Asia** | 22.35 | 0.1132 | 0.05028 | 0.2343 |  |  |  |  |
| **Europe** | 31.14 | 0.7333 | 0.948 | 0.4387 | 0.03496 |  |  |  |
| **Middle East** | 34.56 | 0.2759 | 0.5023 | 0.124 | 0.00339 | 0.4605 |  |  |
| **Oceania** | 39.10 | 0.03721 | 0.09935 | 0.0107 | 5.87E-05 | 0.08584 | 0.3401 |  |
| **America** | 40.70 | 0.01494 | 0.04633 | 0.003619 | 1.07E-05 | 0.03911 | 0.197 | 0.7432 |
| **P-values for High SBP proportional mortality in 2009** | | | | | | | | |
|  | Proportional mortality | **Global** | **AFR** | **CSA** | **EAS** | **EUR** | **MDE** | **OCE** |
| **Global** | 28.32 |  |  |  |  |  |  |  |
| **Africa** | 30.36 | 0.6503 |  |  |  |  |  |  |
| **Central & South Asia** | 28.64 | 0.9418 | 0.7093 |  |  |  |  |  |
| **East Asia** | 30.43 | 0.6386 | 0.9873 | 0.6926 |  |  |  |  |
| **Europe** | 27.24 | 0.8107 | 0.4972 | 0.7555 | 0.4874 |  |  |  |
| **Middle East** | 26.55 | 0.6949 | 0.4073 | 0.6429 | 0.3987 | 0.8772 |  |  |
| **Oceania** | 22.53 | 0.1994 | 0.08884 | 0.1766 | 0.0861 | 0.2909 | 0.3634 |  |
| **America** | 24.66 | 0.4167 | 0.215 | 0.3778 | 0.2094 | 0.5623 | 0.6684 | 0.6115 |
| **P-values for Strokes proportional mortality in 2009** | | | | | | | | |
|  | Proportional mortality | **Global** | **AFR** | **CSA** | **EAS** | **EUR** | **MDE** | **OCE** |
| **Global** | 42.10 |  |  |  |  |  |  |  |
| **Africa** | 38.20 | 0.4295 |  |  |  |  |  |  |
| **Central & South Asia** | 43.68 | 0.7489 | 0.2593 |  |  |  |  |  |
| **East Asia** | 47.21 | 0.3003 | 0.06353 | 0.4761 |  |  |  |  |
| **Europe** | 41.62 | 0.9232 | 0.4809 | 0.6784 | 0.2628 |  |  |  |
| **Middle East** | 38.90 | 0.5161 | 0.8862 | 0.3346 | 0.09559 | 0.5797 |  |  |
| **Oceania** | 38.37 | 0.4501 | 0.9717 | 0.2845 | 0.0765 | 0.5094 | 0.9146 |  |
| **America** | 34.65 | 0.1313 | 0.4652 | 0.06867 | 0.01185 | 0.1572 | 0.384 | 0.4442 |
| **P-values for Metabolic proportional mortality in 2014** | | | | | | | | |
|  | Proportional mortality | **Global** | **AFR** | **CSA** | **EAS** | **EUR** | **MDE** | **OCE** |
| **Global** | 30.64 |  |  |  |  |  |  |  |
| **Africa** | 32.47 | 0.6902 |  |  |  |  |  |  |
| **Central & South Asia** | 29.16 | 0.7491 | 0.4794 |  |  |  |  |  |
| **East Asia** | 23.83 | 0.1395 | 0.06477 | 0.2403 |  |  |  |  |
| **Europe** | 31.97 | 0.7726 | 0.9141 | 0.5369 | 0.05594 |  |  |  |
| **Middle East** | 35.32 | 0.3099 | 0.5437 | 0.1757 | 0.006987 | 0.4728 |  |  |
| **Oceania** | 39.69 | 0.04966 | 0.1236 | 0.0206 | 0.000197 | 0.09801 | 0.3608 |  |
| **America** | 41.66 | 0.01683 | 0.04989 | 0.00598 | 2.85E-05 | 0.03778 | 0.1847 | 0.687 |
| **P-values for High SBP proportional mortality in 2014** | | | | | | | | |
|  | Proportional mortality | **Global** | **AFR** | **CSA** | **EAS** | **EUR** | **MDE** | **OCE** |
| **Global** | 28.22 |  |  |  |  |  |  |  |
| **Africa** | 30.05 | 0.6851 |  |  |  |  |  |  |
| **Central & South Asia** | 28.11 | 0.9794 | 0.6719 |  |  |  |  |  |
| **East Asia** | 30.50 | 0.6123 | 0.9208 | 0.5938 |  |  |  |  |
| **Europe** | 27.17 | 0.8148 | 0.53 | 0.8348 | 0.4688 |  |  |  |
| **Middle East** | 26.47 | 0.6973 | 0.4354 | 0.7162 | 0.3812 | 0.8755 |  |  |
| **Oceania** | 22.29 | 0.1877 | 0.09073 | 0.1959 | 0.07452 | 0.273 | 0.3435 |  |
| **America** | 24.64 | 0.4265 | 0.2385 | 0.4411 | 0.2031 | 0.5703 | 0.6786 | 0.5722 |
| **P-values for Strokes proportional mortality in 2014** | | | | | | | | |
|  | Proportional mortality | **Global** | **AFR** | **CSA** | **EAS** | **EUR** | **MDE** | **OCE** |
| **Global** | 41.14 |  |  |  |  |  |  |  |
| **Africa** | 37.48 | 0.4567 |  |  |  |  |  |  |
| **Central & South Asia** | 42.73 | 0.7465 | 0.2778 |  |  |  |  |  |
| **East Asia** | 45.67 | 0.3573 | 0.09057 | 0.5524 |  |  |  |  |
| **Europe** | 40.86 | 0.9549 | 0.4844 | 0.7056 | 0.3344 |  |  |  |
| **Middle East** | 38.21 | 0.5516 | 0.8796 | 0.3608 | 0.1343 | 0.5896 |  |  |
| **Oceania** | 38.02 | 0.5262 | 0.9105 | 0.3411 | 0.1247 | 0.5634 | 0.969 |  |
| **America** | 33.70 | 0.1305 | 0.4351 | 0.0679 | 0.01626 | 0.1451 | 0.3532 | 0.3733 |
| **P-values for Metabolic proportional mortality in 2019** | | | | | | | | |
|  | Proportional mortality | **Global** | **AFR** | **CSA** | **EAS** | **EUR** | **MDE** | **OCE** |
| **Global** | 31.51 |  |  |  |  |  |  |  |
| **Africa** | 33.28 | 0.7028 |  |  |  |  |  |  |
| **Central & South Asia** | 30.47 | 0.8229 | 0.5506 |  |  |  |  |  |
| **East Asia** | 25.14 | 0.1708 | 0.08429 | 0.2476 |  |  |  |  |
| **Europe** | 32.32 | 0.8616 | 0.8381 | 0.6878 | 0.09831 |  |  |  |
| **Middle East** | 35.89 | 0.3448 | 0.5788 | 0.2382 | 0.01321 | 0.4441 |  |  |
| **Oceania** | 40.11 | 0.06392 | 0.147 | 0.03609 | 0.00056 | 0.09549 | 0.3792 |  |
| **America** | 42.19 | 0.02151 | 0.05874 | 0.01089 | 8.57E-05 | 0.03484 | 0.1897 | 0.6724 |
| **P-values for High SBP proportional mortality in 2019** | | | | | | | | |
|  | Proportional mortality | **Global** | **AFR** | **CSA** | **EAS** | **EUR** | **MDE** | **OCE** |
| **Global** | 27.89 |  |  |  |  |  |  |  |
| **Africa** | 29.60 | 0.7044 |  |  |  |  |  |  |
| **Central & South Asia** | 27.66 | 0.9591 | 0.6722 |  |  |  |  |  |
| **East Asia** | 30.09 | 0.6246 | 0.914 | 0.5878 |  |  |  |  |
| **Europe** | 27.07 | 0.8536 | 0.5795 | 0.8937 | 0.5099 |  |  |  |
| **Middle East** | 26.24 | 0.7116 | 0.4617 | 0.7495 | 0.4009 | 0.8517 |  |  |
| **Oceania** | 22.01 | 0.1892 | 0.09634 | 0.2059 | 0.07799 | 0.2547 | 0.3362 |  |
| **America** | 24.48 | 0.4471 | 0.2628 | 0.4772 | 0.2217 | 0.5612 | 0.6906 | 0.5496 |
| **P-values for Strokes proportional mortality in 2019** | | | | | | | | |
|  | Proportional mortality | **Global** | **AFR** | **CSA** | **EAS** | **EUR** | **MDE** | **OCE** |
| **Global** | 40.60 |  |  |  |  |  |  |  |
| **Africa** | 37.13 | 0.4793 |  |  |  |  |  |  |
| **Central & South Asia** | 41.87 | 0.796 | 0.3261 |  |  |  |  |  |
| **East Asia** | 44.77 | 0.3961 | 0.1137 | 0.557 |  |  |  |  |
| **Europe** | 40.62 | 0.9971 | 0.4698 | 0.7997 | 0.404 |  |  |  |
| **Middle East** | 37.87 | 0.5782 | 0.8776 | 0.4175 | 0.1654 | 0.5757 |  |  |
| **Oceania** | 37.88 | 0.58 | 0.8756 | 0.4189 | 0.1662 | 0.5775 | 0.9979 |  |
| **America** | 33.33 | 0.1388 | 0.4322 | 0.08348 | 0.02145 | 0.1379 | 0.3494 | 0.3481 |

**Table S5. Comparison of proportional prevalence of Stroke, HSBP and Metabolic conditions.** The proportional prevalence for stroke, HSBP and metabolic conditions in each region was compared with global proportional prevalence as well as regional proportional Prevalence in a pairwise manner for 2009 and 2014. P-value (2-sided) obtained using one-sample test for binomial proportion using normal-theory method.

| **P-values for metabolic proportional prevalence in 2009** | | | | | | | | |
| --- | --- | --- | --- | --- | --- | --- | --- | --- |
|  | Proportional Prevalence | **Global** | **AFR** | **CSA** | **EAS** | **EUR** | **MDE** | **OCE** |
| **Global** | 47.88 |  |  |  |  |  |  |  |
| **Africa** | 39.79 | 0.1054 |  |  |  |  |  |  |
| **Central & South Asia** | 36.60 | 0.02399 | 0.5148 |  |  |  |  |  |
| **East Asia** | 41.57 | 0.2066 | 0.7162 | 0.3025 |  |  |  |  |
| **Europe** | 52.40 | 0.365 | 0.009966 | 0.001037 | 0.02793 |  |  |  |
| **Middle East** | 57.35 | 0.05798 | 0.000334 | 1.66E-05 | 0.001366 | 0.322 |  |  |
| **Oceania** | 61.20 | 0.007669 | 1.22E-05 | 3.29E-07 | 6.81E-05 | 0.07824 | 0.4364 |  |
| **America** | 63.89 | 0.001345 | 8.45E-07 | <1E-07 | 5.9E-06 | 0.0214 | 0.1857 | 0.58 |
| **P-values for High SBP proportional prevalence in 2009** | | | | | | | | |
|  | Proportional Prevalence | **Global** | **AFR** | **CSA** | **EAS** | **EUR** | **MDE** | **OCE** |
| **Global** | 44.92 |  |  |  |  |  |  |  |
| **Africa** | 52.20 | 0.1431 |  |  |  |  |  |  |
| **Central & South Asia** | 54.28 | 0.05982 | 0.6775 |  |  |  |  |  |
| **East Asia** | 49.94 | 0.3126 | 0.6507 | 0.3837 |  |  |  |  |
| **Europe** | 41.73 | 0.521 | 0.03595 | 0.01173 | 0.1004 |  |  |  |
| **Middle East** | 33.65 | 0.02349 | 0.000204 | 3.47E-05 | 0.001121 | 0.1015 |  |  |
| **Oceania** | 32.42 | 0.01195 | 7.45E-05 | 1.14E-05 | 0.000457 | 0.05902 | 0.7938 |  |
| **America** | 31.17 | 0.005709 | 2.55E-05 | 3.49E-06 | 0.000174 | 0.0323 | 0.5996 | 0.7901 |
| **P-values for Strokes proportional prevalence in 2009** | | | | | | | | |
|  | Proportional Prevalence | **Global** | **AFR** | **CSA** | **EAS** | **EUR** | **MDE** | **OCE** |
| **Global** | 7.20 |  |  |  |  |  |  |  |
| **Africa** | 8.01 | 0.7558 |  |  |  |  |  |  |
| **Central & South Asia** | 9.12 | 0.4588 | 0.6823 |  |  |  |  |  |
| **East Asia** | 8.49 | 0.6187 | 0.8589 | 0.8272 |  |  |  |  |
| **Europe** | 5.87 | 0.6063 | 0.4311 | 0.2592 | 0.3474 |  |  |  |
| **Middle East** | 8.99 | 0.4869 | 0.7144 | 0.9674 | 0.8546 | 0.183 |  |  |
| **Oceania** | 6.38 | 0.752 | 0.5503 | 0.3426 | 0.4504 | 0.8264 | 0.361 |  |
| **America** | 4.93 | 0.3804 | 0.2577 | 0.1462 | 0.2022 | 0.6907 | 0.1555 | 0.5529 |
| **P-values for Metabolic proportional prevalence in 2014** | | | | | | | | |
|  | Proportional Prevalence | **Global** | **AFR** | **CSA** | **EAS** | **EUR** | **MDE** | **OCE** |
| **Global** | 50.74 |  |  |  |  |  |  |  |
| **Africa** | 42.57 | 0.1024 |  |  |  |  |  |  |
| **Central & South Asia** | 38.39 | 0.01356 | 0.3984 |  |  |  |  |  |
| **East Asia** | 45.11 | 0.2603 | 0.6077 | 0.1674 |  |  |  |  |
| **Europe** | 56.12 | 0.2811 | 0.006117 | 0.000267 | 0.02683 |  |  |  |
| **Middle East** | 61.23 | 0.03582 | 0.000161 | 2.66E-06 | 0.001195 | 0.3036 |  |  |
| **Oceania** | 64.59 | 0.005576 | 8.42E-06 | <1E-07 | 9.01E-05 | 0.0879 | 0.49 |  |
| **America** | 67.04 | 0.001109 | 7.45E-07 | <1E-07 | 1.04E-05 | 0.02782 | 0.233 | 0.6087 |
| **P-values for High SBP proportional prevalence in 2014** | | | | | | | | |
|  | Proportional Prevalence | **Global** | **AFR** | **CSA** | **EAS** | **EUR** | **MDE** | **OCE** |
| **Global** | 42.24 |  |  |  |  |  |  |  |
| **Africa** | 49.69 | 0.1313 |  |  |  |  |  |  |
| **Central & South Asia** | 52.63 | 0.03541 | 0.5568 |  |  |  |  |  |
| **East Asia** | 46.55 | 0.3824 | 0.5302 | 0.2236 |  |  |  |  |
| **Europe** | 38.19 | 0.4122 | 0.02141 | 0.003825 | 0.09354 |  |  |  |
| **Middle East** | 30.24 | 0.01512 | 9.99E-05 | 7.31E-06 | 0.001073 | 0.1018 |  |  |
| **Oceania** | 29.30 | 0.008794 | 4.53E-05 | 2.97E-06 | 0.000542 | 0.06728 | 0.8377 |  |
| **America** | 28.36 | 0.004958 | 1.99E-05 | 1.17E-06 | 0.000265 | 0.04309 | 0.6826 | 0.8368 |
| **P-values for Strokes proportional prevalence in 2014** | | | | | | | | |
|  | Proportional Prevalence | **Global** | **AFR** | **CSA** | **EAS** | **EUR** | **MDE** | **OCE** |
| **Global** | 7.02 |  |  |  |  |  |  |  |
| **Africa** | 7.74 | 0.7801 |  |  |  |  |  |  |
| **Central & South Asia** | 8.98 | 0.4451 | 0.6432 |  |  |  |  |  |
| **East Asia** | 8.34 | 0.6072 | 0.8222 | 0.8235 |  |  |  |  |
| **Europe** | 5.69 | 0.6007 | 0.4427 | 0.2499 | 0.3375 |  |  |  |
| **Middle East** | 8.53 | 0.5556 | 0.7667 | 0.8763 | 0.9445 | 0.2195 |  |  |
| **Oceania** | 6.11 | 0.72 | 0.5419 | 0.3158 | 0.4199 | 0.8556 | 0.3858 |  |
| **America** | 4.60 | 0.3424 | 0.2399 | 0.1256 | 0.1761 | 0.6383 | 0.1592 | 0.3424 |
