## Supplementary file 4 for "Ethnic and region-specific genetic risk variants of stroke and its comorbid conditions can define the variations in the burden of stroke and its phenotypic traits"

**Table S6.** Variants and genes associated with Stroke and its comorbid factors that are shared among populations, as well as unique to populations. The number and percentage of GWAS risk variants which are shared among the different super-populations Africa (AFR), East Asia (EAS), South Asia (SAS), Europe (EUR) and America (AMR) for (a) Stroke, (b) IHD, (c) High BP, (d) T2D, (e) T1D, (f) CKD, (g) High BMI and (h) High LDL are given. The risk variant is considered to be present in a population if the alternate allele frequency in 1000 Genomes project is above 0.05. The number of genes that variants map to are also given. For each disease, the populations that share the variant, the rsID, the genes that the variant maps to as well as location are given.

| **a. GWAS Risk variants of Stroke unique to and shared among populations** | | | | | |
| --- | --- | --- | --- | --- | --- |
| **Intersection of populations** | **RSID** | **Gene mapped to** | **Intersection of populations** | **RSID** | **Gene mapped to** |
| AFR:EAS:EUR:AMR | rs2267739 | ADCYAP1R1 | AFR:EAS:SAS:EUR | rs799165 | VPS37D, MLXIPL |
| SAS:AMR | rs117688302 | PARVA | SAS:EUR | rs78037902 | - |
| AFR | rs11202867 | ANKRD22 | SAS | rs148010464 | PLA2G4A, LINC01036 |
|  | rs113952126 | - |  | rs2023938 | HDAC9 |
|  | rs114527838 | FAT2, SPARC |  | rs528002287 | PCSK6 |
|  | rs11862102 | - | SAS:EUR:AMR | rs11065979 | BRAP, ATXN2-AS |
|  | rs11864414 | LINC02181 |  | rs73236850 | TBC1D1 |
|  | rs12291066 | MICAL2 |  | rs11556924 | ZC3HC1 |
|  | rs12310617 | TSPAN9, TEAD4 |  | rs12196992 | LINC01016 |
|  | rs16867253 | GRHL1 |  | rs12204590 | FOXF2-DT, LINC01394 |
|  | rs16962778 | MBTPS1 |  | rs17696736 | NAA25 |
|  | rs1869717 | MAML3 |  | rs2084898 | TRIM29 |
|  | rs6891305 | LMAN2 |  | rs60942712 | ICE2P2, GAPDHP50 |
|  | rs72976591 | KALRN |  | rs62083469 | RPL12P40 |
|  | rs7342953 | RGS9 |  | rs62367577 | RNU6-727P |
|  | rs73923591 | RN7SL340P, UQCRFS1-DT |  | rs11065987 | BRAP, ATXN2-AS |
|  | rs7610618 | FKBP1AP4, RPL32P8 |  | rs80194260 | CPSF2, RNU6-366P |
|  | rs768606 | LINC00709 |  | rs77707008 | DLG2 |
|  | rs8082812 | AKR1B1P6, THEMIS3P | AFR:AMR1 | rs7081476 | FAM238C, ANKRD26 |
| EAS:AMR | rs199722446 | POMZP3 | AFR:AMR2 | rs7811443 | AMPH |
| AFR:EAS | rs11217335 | USP2-AS1, KRT8P7 | EUR | rs71654438 | LINC01650, LINC01760 |
|  | rs17140821 | FERD3L |  | rs112455974 | DLGAP2 |
|  | rs55682257 | DKK2 |  | rs13114738 | SLC39A8 |
| AFR:EAS:AMR | rs13116934 | C4orf54, DAPP1 |  | rs17827444 | ADCY2 |
|  | rs34727243 | CLYBL |  | rs35276016 | LINC02828 |
|  | rs906689 | ATPSCKMT |  | rs10455872 | LPA |
|  | rs9324898 | ARHGAP26 |  | rs72983521 | MMP12, MMP3 |
| AFR:EAS:SAS | rs17167021 | LINC02113, GUSBP8 |  | rs71654444 | LINC02607, LINC01761 |
|  | rs72749603 | CDH6 |  | rs79820282 | RGS4, RGS5 |
|  | rs7937106 | ALKBH8, CWF19L2 |  | rs79861969 | Y_RNA |
| AFR:EAS:SAS:AMR | rs2417957 | SLCO1B1 | EUR:AMR | rs12369179 | ZCCHC8 |
|  | rs6046851 | RALGAPA2 |  | rs17463708 | NFXL1 |
| EAS:EUR | rs4471613 | ALDH1A2 |  | rs17490626 | TSPAN15 |
| EAS:SAS:AMR | rs9345396 | - |  | rs188186531 | NBEAL1 |
| EAS1 | rs72614300 | TEK |  | rs34324219 | TCN1 |
|  | rs74095939 | KLF12 |  | rs78268213 | NTS, MGAT4C |
|  | rs75473520 | TCEANC2 |  | rs61949042 | MGAT4C |
|  | rs77237189 | LINC01781 |  | rs704341 | PTPRG |
|  | rs78868369 | - |  | rs72934535 | NBEAL1 |
| EAS:EUR:AMR1 | rs116482258 | CYP4F44P |  | rs7582720 | WDR12 |
|  | rs17201317 | MTND1P5, ATP6V1C1 |  | rs4959130 | FOXF2-DT, LINC01394 |
| EAS:SAS1 | rs12420422 | BSX, RPL34P23 |  | rs76906076 | TRPC6 |
|  | rs57496717 | HIP1 | AMR | rs4905014 | ITPK1 |
| EAS:SAS:EUR | rs17347800 | HDAC9 | AFR:EUR | rs10132309 | MEG8 |
|  | rs2074755 | BAZ1B | AFR:SAS | rs6759518 | SLC30A3 |
| EAS:SAS:EUR:AMR | rs10486776 | RPL36AP26 |  | rs13354619 | RNU6-909P |
|  | rs1122608 | SMARCA4 |  | rs114233315 | TUBGCP3, ATP11AUN |
|  | rs12037987 | WNT2B | AFR:EUR:AMR | rs16939239 | - |
|  | rs4867766 | LINC01411, SUMO2P6 |  | rs184708393 | RFX3-AS1, GLIS3 |
|  | rs12190287 | TCF21 |  | rs55634002 | - |
|  | rs12413409 | CNNM2 |  | rs73031894 | - |
|  | rs17275724 | GATB |  | rs9936995 | C16orf95 |
|  | rs174547 | FADS1, FADS2 |  | rs59447555 | AK5 |
|  | rs2025481 | ENOX1 | AFR:SAS:EUR | rs11596587 | HK1 |
|  | rs2293576 | SLC39A13 |  | rs73504401 | - |
|  | rs2822388 | FRG2MP, ANKRD20A18P |  | rs9899728 | RNU6-362P, MRPL58 |
|  | rs4782918 | WFDC1 |  |  |  |
|  | rs12118362 | NBPF3 |  |  |  |
| AFR:SAS:EUR:AMR | | | | | |
| rs10017904 | CAMK2D | rs17114036 | PLPP3 | rs562338 | TDRD15, APOB |
| rs10029218 | CAMK2D | rs11984041 | HDAC9 | rs6463230 | RBAK-RBAKDN, RNF216P1 |
| rs2005108 | RNU7-159P, BOLA3P1 | rs2287921 | RASIP1 | rs6647 | SERPINA1 |
| rs12122341 | LINC01765 | rs42039 | CDK6 | rs7086422 | PRKG1 |
| rs12124533 | LINC01765 | rs4506565 | TCF7L2 | rs71524263 | HDAC9 |
| rs1564060 | MTND4P33 | rs4714955 | PHACTR1 | rs938291 | SP3, RPL5P7 |
| rs492602 | FUT2 |  |  |  |  |
| AFR:EAS:SAS:EUR:AMR | | | | | |
| rs11604904 | MICAL2 | rs1005224 | TTLL5 | rs1333048 | CDKN2B-AS1 |
| rs225132 | ERRFI1 | rs10057967 | POC5 | rs1333049 | CDKN2B-AS1 |
| rs11681884 | IL1RN, RNU6-1180P | rs1465330 | ZDHHC22, TMEM63C | rs13401697 | LINC02572, PLAC9P1 |
| rs11846182 | DIO2-AS1 | rs10241902 | DLX6-AS1 | rs1364044 | ADAMTS12 |
| rs11697087 | PHF20 | rs10401969 | SUGP1 | rs1449514 | DOCK10 |
| rs1052053 | PMF1-BGLAP, PMF1 | rs1046040 | GPX4, POLR2E | rs11694469 | EVA1A, EVA1A-AS |
| rs11867415 | PRPF8 | rs10863936 | DTL | rs1564454 | FARP1 |
| rs16851055 | SPSB4 | rs10820405 | LINC01492 | rs161802 | PARK7 |
| rs12425791 | NINJ2-AS1, LINC02455 | rs12445022 | JPH3, ZCCHC14 | rs16896398 | TTBK1, SLC22A7 |
| rs1801222 | CUBN | rs114209171 | FUNDC2 | rs11957829 | ZNF474-AS1 |
| rs12646447 | LINC01438 | rs11553746 | ACP1 | rs1697421 | NBPF3, ALPL |
| rs12476527 | KCNK3 | rs13143308 | LINC01438 | rs1728918 | PPM1G, NRBP1 |
| rs12579302 | ATP2B1 | rs13168506 | TGFBI | rs17612742 | EDNRA |
| rs12611820 | - | rs1333047 | CDKN2B-AS1 | rs1799983 | NOS3 |
| rs1937787 | HMGB1P18, HNRNPA1P64 | rs2219939 | ADAMTS7, CHRNB4 | rs2456519 | RPSAP55, ONECUT1 |
| rs12932445 | ZFHX3 | rs222826 | RNU7-2P | rs12449964 | SMCR2, PEMT |
| rs12936587 | SMCR2, PEMT | rs2229383 | ILF3 | rs2084637 | MIR100HG |
| rs12939005 | TOM1L1 | rs2235797 | ST13P19, HHAT | rs2107595 | TWIST1, HDAC9 |
| rs615756 | LINC01234 | rs2238151 | ALDH2 | rs2169955 | - |
| rs11681377 | VDAC2P5, SNORA74 | rs2295786 | SH3PXD2A, STN1 | rs2200733 | LINC01438, PITX2 |
| rs62312664 | MMRN1 | rs2290911 | SH3YL1 | rs10179686 | TMEM163 |
| rs339800 | - | rs35436 | - | rs599839 | CELSR2, PSRC1 |
| rs248812 | PAFAH1B2P2, RMST | rs3790099 | GNAO1 | rs60216804 | OR4D2, OR4D1 |
| rs2594966 | ATG7 | rs3803802 | DNAH2 | rs60401382 | HTRA1 |
| rs261332 | LIPC-AS1, LIPC, ALDH1A2 | rs4304924 | RPL21P111, OBI1 | rs2866611 | ADI1P1, ZHX3 |
| rs2632512 | RNF43, TSPOAP1-AS1 | rs4420638 | APOC1, APOC1P1 | rs4424353 | ARMC8P1, IGFBPL1 |
| rs4686799 | KNG1 | rs4684776 | ATG7 | rs62314748 | LINC02511 |
| rs2980853 | TRIB1 | rs5744680 | POLK | rs629301 | CELSR2 |
|  |  | rs59640442 | - | rs632728 | PRRC2A |
| rs3184504 | ATXN2, SH2B3 | rs247617 | HERPUD1, CETP | rs638704 | GORAB, PRRX1 |
| rs34311906 | RPL7AP30, ANK2 | rs4803750 | BCL3, CEACAM16-AS1 | rs6533530 | MIR297, LINC01438 |
| rs34978547 | - | rs469568 | ADAMTS2 | rs6695915 | PTPRF |
| rs35323955 | - | rs4704221 | CERT1 | rs6825454 | FGB, FGA |
| rs4444072 | GALNT18 | rs4710305 | - | rs780094 | GCKR |
| rs4444878 | F11-AS1 | rs4727833 | CAV2 | rs6843082 | LINC01438 |
| rs460976 | MX1 | rs4737316 | EYA1 |  |  |
| rs4621303 | ULK4 | rs478442 | TDRD15 | rs6967981 | - |
| rs4674746 | KCNE4 | rs8003055 | VRTN | rs7714373 | LHFPL2 |
| rs6841581 | EDNRA, PRMT5P1 | rs4932370 | RN7SL363P, FURIN | rs6891174 | BNIP1, RPL7AP33 |
| rs8103309 | SMARCA4 | rs536348 | LINC01558, LINC02487 | rs781542 | SPINK2 |
| rs879324 | ZFHX3 | rs556621 | HCG27 | rs783396 | CRYBG1 |
| rs74154533 | NEURL1 | rs56393506 | PLG, LPA | rs7859727 | CDKN2B-AS1 |
| rs7506045 | IMPA2 | rs7103059 | TRPC2 | rs880315 | CASZ1 |
| rs75731302 | - | rs710446 | KNG1 | rs899997 | CHRNB4 |
| rs760762 | PLCG1 | rs7115242 | SIK3 | rs903817 | LINC02743 |
| rs7632505 | SEMA5B | rs7117121 | CNTN5 | rs940274 | ABCA12 |
| rs7654093 | FGG, LRAT | rs7126770 | TRPC6 | rs9348394 | LNC-LBCS |
| rs765547 | - | rs7156510 | MTND4P33 | rs9351814 | LINC01626 |
| rs9939609 | FTO | rs7190256 | ZFHX3 | rs9797861 | SLC44A2 |
| rs7705819 | RNU6-500P | rs9526212 | LRCH1 | rs9515201 | COL4A2 |
| rs9536591 | LINC00558 | rs7304841 | PDE3A | rs9521634 | COL4A1 |
| rs7703051 | ANKRD31, HMGCR | rs9295674 | H2BP5, SLC17A2 | rs9924207 | LINC02164, ENPP7P14 |
| rs964184 | ZPR1 | rs720470 | KCNJ6, DYRK1A | rs9958650 | ZBTB14, EPB41L3 |
| **b. GWAS Risk variants of IHD unique to and shared among populations** | | | | | |
| **Intersection** | **RSID** | **Gene mapped to** | **Intersection** | **RSID** | **Gene mapped to** |
| AFR:EAS:EUR | rs7412 | APOE | AMR | rs114799463 | HNRNPA1P67, RNU4ATAC9P |
| AFR | rs12310617 | TSPAN9, TEAD4 |  | rs12658928 | NPR3 |
|  | rs16867253 | GRHL1 |  | rs140570886 | LPA |
|  | rs1869717 | MAML3 |  | rs144648928 | FGGY, LINC02884 |
|  | rs571622299 | KCNB2 |  | rs180986320 | FGD6 |
|  | rs6997330 | LPL |  | rs41272114 | LPA |
|  | rs7011846 | LPL |  | rs4905014 | ITPK1 |
|  | rs8082812 | AKR1B1P6, THEMIS3P |  | rs72823390 | FBXL20 |
| AFR:AMR1 | rs7081476 | FAM238C, ANKRD26 |  | rs79576311 | SILC1 |
|  | rs980831 | FLT4 | EAS:EUR | rs11631816 | HCN4 |
| AFR:EAS | rs11606719 | BCL9L | EAS | rs11066015 | ACAD10 |
|  | rs17140821 | FERD3L |  | rs11066280 | HECTD4 |
| AFR:EAS:EUR:AMR | rs4076834 | ABCG8 |  | rs11235604 | ATG16L2 |
|  | rs4456565 | BCAS3 |  | rs75419803 | RNU6-334P |
|  | rs7212798 | BCAS3 |  | rs139382189 | FBXO21 |
|  | rs7214227 | BCAS3 |  | rs671 | ALDH2 |
|  | rs8080784 | BCAS3 |  | rs73018809 | NUP210 |
| AFR:EAS:SAS | rs141622900 | APOC1P1, APOC1 |  | rs12569313 | C1orf220, RNA5SP69 |
|  | rs180803 | POM121L9P, BCRP1 |  | rs77202337 | - |
|  | rs190712692 | APOC1P1, APOC1 |  | rs79105258 | CUX2 |
|  | rs28607113 | MCTP2, H3P40 | EAS:AMR | rs11066707 | KCTD10 |
|  | rs5760293 | POM121L9P, SPECC1L |  | rs1558803 | UBE3B |
|  | rs7775726 | NUS1 |  | rs3798220 | LPA |
|  | rs73386640 | BET1L |  | rs74323928 | RASAL3, WIZ |
|  | rs6004124 | SPECC1L-ADORA2A, SPECC1L |  | rs75160195 | LINC02381, SMUG1 |
|  | rs9624456 | SPECC1L-ADORA2A, SPECC1L |  | rs79946623 | - |
| AFR:EAS:SAS:AMR | rs73392700 | SIRT3 | EAS:EUR:AMR | rs17165136 | THSD7A |
|  | rs75887718 | OPCML |  | rs35093463 | ABCA1 |
|  | rs77628708 | TRIM5, TRIM6-TRIM34 |  | rs72805949 | RPL17P35 |
|  | rs9493740 | LINC01312, TARID | EAS:SAS | rs11065783 | LINC01405 |
| EAS:SAS:AMR | rs192256050 | TMEM132D |  | rs12420422 | BSX, RPL34P23 |
|  | rs3796562 | GUCY1B1 |  | rs12469758 | NUTF2P8 |
| AFR:EAS:SAS:EUR | rs11167260 | PROCR | EAS:SAS:EUR | rs17577085 | SPRY4-AS1 |
|  | rs2683043 | CYP4F3 |  | rs17678683 | LINC01412 |
|  | rs3782889 | MYL2 |  | rs2074755 | BAZ1B |
|  | rs799165 | VPS37D, MLXIPL |  | rs2623168 | CDH17, GEM |
| SAS:EUR | rs147802096 | Y_RNA, RNU1-88P |  | rs769446 | APOE, TOMM40 |
|  | rs17458018 | FN1 | EUR | rs10455872 | LPA |
|  | rs41311933 | C5 |  | rs113114656 | SMAD2 |
|  | rs73082363 | QRICH1 |  | rs113716316 | AHDC1 |
| SAS:AMR | rs12214416 | LPAL2 |  | rs11670056 | ELL |
| SAS | rs1014675 | LINC00485 |  | rs11752218 | RAB23, PRIM2 |
|  | rs11571537 | ATF3 |  | rs13114738 | SLC39A8 |
|  | rs73596816 | LPA |  | rs28641753 | - |
| AFR:EUR:AMR | rs10111944 | NARS1P2 |  | rs34891485 | DHX58 |
|  | rs10961206 | - |  | rs3918226 | NOS3 |
|  | rs11206510 | BSND, PCSK9 |  | rs4252185 | PLG |
|  | rs11610422 | SINHCAF, MREGP1 |  | rs55730499 | LPA |
|  | rs1346132 | HNRNPA1P67, RNU4ATAC9P |  | rs71566846 | RAB23, PRIM2 |
|  | rs35465346 | LDLRAD2, USP48 |  | rs73104091 | SAMMSON |
|  | rs6460942 | VWDE |  | rs74617384 | LPA |
|  | rs6984210 | BMP1 | AFR:SAS | rs112001543 | XKR4 |
|  | rs75331444 | ABCG8 |  | rs12344245 | FAM189A2 |
|  | rs9501744 | GMDS, FOXC1 |  | rs6759518 | SLC30A3 |
| AFR:SAS:AMR | rs7305553 | LINC02882 | AFR:SAS:EUR | rs1657346 | LINC02881 |
|  |  |  |  | rs78349783 | NOX3 |
| EAS:SAS:EUR:AMR | | | | | |
| rs1034246 | PTK7 | rs12520174 | - | rs13195460 | HECA |
| rs10456100 | KCNK5 | rs12788925 | MMP13, RNU7-159P | rs1334894 | FKBP5 |
| rs10488763 | LINC02732 | rs12826942 | PPHLN1 | rs13734 | RRBP1 |
| rs10883832 | NT5C2 | rs12413409 | CNNM2 | rs1378942 | CSK |
| rs10940235 | EMB | rs27906 | - | rs140244541 | WDR12, CARF |
| rs10947789 | KCNK5 | rs2879627 | PRKG1 | rs1405130 | LINC01173 |
| rs10953541 | BCAP29, DUS4L-BCAP29 | rs332827 | NFIA | rs16844401 | HGFAC |
| rs10981012 | SHOC1 | rs35006 | ARL15 | rs16917237 | BDNF, BDNF-AS |
| rs10985348 | DAB2IP | rs35224956 | MARK3 | rs174547 | FADS2, FADS1 |
| rs11107909 | FGD6 | rs360798 | EHBP1 | rs17843797 | UMPS |
| rs11191447 | AS3MT, BORCS7-ASMT | rs36096196 | SKI, MORN1 | rs1800978 | ABCA1 |
| rs11204666 | MCL1 | rs3740390 | AS3MT, BORCS7-ASMT | rs1887318 | JCAD |
| rs1122608 | SMARCA4 | rs3741782 | CORO1C | rs1926032 | CNNM2 |
| rs113244441 | TULP1, FKBP5 | rs3827066 | ZNF335 | rs197907 | LINC01974, WNT9B |
| rs11552449 | DCLRE1B | rs4678145 | UMPS | rs2048327 | SLC22A3 |
| rs11601507 | TRIM5 | rs4794213 | MBTD1 | rs2306363 | SIPA1 |
| rs11698996 | GGT7 | rs4836390 | SLC27A6 | rs2505083 | JCAD |
| rs11760633 | MAD1L1, ELFN1 | rs55714120 | PHB | rs26079 | PARP8 |
| rs117742247 | ATP2B1 | rs56307353 | CSNK1G2 | rs60154123 | ST13P19, HHAT |
| rs12149545 | HERPUD1, CETP | rs56336142 | SAYSD1, KCNK5 | rs60787346 | HSF2BP |
| rs12190287 | TCF21 | rs564398 | CDKN2B-AS1 | rs61797063 | NGF-AS1 |
| rs12315434 | R3HDM2 | rs59372292 | HNRNPM | rs62076439 | ZNF652 |
| rs9532984 | DGKH | rs852787 | DAB1 | rs67807996 | VPS45, OTUD7B |
| rs9985766 | DCLK2 | rs885150 | DAB2IP | rs68170813 | DUS4L-BCAP29, BCAP29 |
| rs7767084 | LPA | rs933899 | NR2C1 | rs72675530 | RPSAP20 |
| rs78031527 | C4orf50, JAKMIP1 | rs9349379 | PHACTR1 | rs72689147 | GUCY1A1 |
| rs78368565 | COL4A2 | rs9354144 | - | rs7269833 | LINC01522, RNU7-92P |
| rs7484541 | R3HDM2 | rs9381634 | PHACTR1 | rs72874178 | - |
| rs76954792 | RNU6-1134P, MIR365B | rs7502499 | PHB | rs73155085 | LINC02557, PCAT14 |
| EUR:AMR | | | | | |
| rs112941079 | SEMA5A | rs17263917 | TAS2R1, SNHG18 | rs6435169 | WDR12 |
| rs114123510 | WDR12, CARF | rs17608766 | GOSR2 | rs6725887 | WDR12 |
| rs115654617 | NBEAL1 | rs17616661 | KANK2 | rs6728861 | WDR12 |
| rs116161686 | ST13P19, HHAT | rs2107732 | CCM2 | rs72926772 | WDR12, CARF |
| rs12369179 | ZCCHC8 | rs28451064 | LINC00310, KCNE2 | rs72934535 | NBEAL1 |
| rs144961578 | MTRNR2L9 | rs61741262 | TNS1 | rs73143584 | ZBTB46, ZBTB46-AS1 |
| rs147055617 | PLPP3 | rs61879614 | TUB | rs73225842 | BMP1 |
| rs17206992 | GALNT13 | rs62390615 | FOXC1, GMDS | rs7582720 | WDR12 |
| SAS:EUR:AMR | | | | | |
| rs10950702 | GRIFIN, CHST12 | rs12423664 | FBRSL1 | rs10951983 | DAGLB, RAC1 |
| rs35099186 | ST3GAL4 | rs12866570 | COL4A1 | rs3864814 | KIFBP |
| rs11065987 | ATXN2-AS, BRAP | rs273909 | MIR3936HG, SLC22A4 | rs73045269 | CCDC97, TGFB1 |
| rs113756303 | PPP2R2A | rs17228058 | SMAD3 | rs56170783 | PLPP3 |
| rs11556924 | ZC3HC1 | rs17416285 | PLPP3 | rs6813378 | HGFAC |
| rs11617955 | COL4A1 | rs17609940 | ANKS1A | rs72664335 | PLPP3 |
| rs11806316 | NGF-AS1 | rs17696736 | NAA25 | rs72743461 | SMAD3 |
| rs12136712 | NGF-AS1 | rs2008603 | PREX1 | rs56062135 | SMAD3 |
| rs12205331 | ANKS1A | rs13065626 | STAG1 | rs9337951 | JCAD |
| AFR:SAS:EUR:AMR | | | | | |
| rs10495907 | PLEKHH2, RN7SKP66 | rs34372836 | TEX41 | rs688034 | SEZ6L |
| rs112009435 | MAP1S, FCHO1 | rs35879803 | ZNF827 | rs6922269 | MTHFD1L |
| rs11673093 | EXOC3L2, MARK4 | rs3732837 | MRAS | rs7110894 | OR52B6, HNRNPA1P53 |
| rs12144309 | RSBN1 | rs4472337 | UHRF1BP1 | rs7251815 | MAP1S |
| rs12264252 | TSPAN14 | rs4506565 | TCF7L2 | rs72945132 | FADD, PPFIA1 |
| rs12526453 | PHACTR1 | rs4634932 | PLPP3 | rs73015714 | MAP1S, FCHO1 |
| rs12740374 | CELSR2 | rs4643373 | IGF2BP1 | rs73059933 | ZNF787 |
| rs12753816 | NME7 | rs4714955 | PHACTR1 | rs7528419 | CELSR2 |
| rs17035270 | TET2 | rs4919044 | EXOC6 | rs7604735 | TEX41 |
| rs17114036 | PLPP3 | rs55791371 | SMARCA4 | rs7623687 | RHOA |
| rs17114046 | PLPP3 | rs56125973 | SMARCA4 | rs7901695 | TCF7L2 |
| rs17228212 | SMAD3 | rs56289821 | SMARCA4 | rs7933887 | ST3GAL4 |
| rs1746049 | LINC02881 | rs56313611 | PREX1, ARFGEF2 | rs7947761 | ARHGAP42 |
| rs17517928 | FN1 | rs56348932 | PLPP3 | rs8061878 | RNA5SP430, TERF2IP |
| rs17680741 | TSPAN14 | rs5943057 | CHRDL1 | rs898123 | PAIP2B, MPHOSPH10 |
| rs2306374 | MRAS | rs61194703 | SMARCA4, LDLR | rs9379774 | CARMIL1 |
| rs2493298 | PRDM16 | rs62125344 | - | rs9469890 | UHRF1BP1 |
| rs2590942 | RNU6-1246P, RPL31P12 | rs62473335 | PTPRN2 | rs9818870 | MRAS |
| rs2616407 | LNX1 | rs6511720 | LDLR | rs9906944 | IGF2BP1 |
| rs2814944 | IFITM3P3, ILRUN | rs663640 | RNU4-17P | rs9970807 | PLPP3 |
| rs2814993 | ILRUN | rs6740322 | THADA | rs9982601 | KCNE2, LINC00310 |
| AFR:EAS:SAS:EUR:AMR | | | | | |
| rs10006310 | ZNF827 | rs10811650 | CDKN2B-AS1 | rs11191416 | PFN1P11 |
| rs10041378 | LINC02063 | rs10811652 | CDKN2B-AS1 | rs11191559 | NT5C2 |
| rs10055631 | MAST4 | rs10811654 | CDKN2B-AS1 | rs11204085 | RPL30P9 |
| rs10056811 | HMGCR, ANKRD31 | rs10818576 | DAB2IP | rs11204892 | LINGO4, TDRKH-AS1 |
| rs10057967 | POC5 | rs10826753 | JCAD | rs11207415 | LINC01358, FGGY-DT |
| rs10063823 | - | rs10840293 | SWAP70 | rs11226017 | MIR4693 |
| rs1008898 | C1GALT1 | rs10841443 | LINC02398 | rs11226018 | MIR4693 |
| rs10093110 | ZFPM2-AS1, ZFPM2 | rs10857147 | FGF5, PRDM8 | rs11226029 | MIR4693, DYNC2H1 |
| rs10106652 | RPL30P9 | rs10892564 | ARHGEF12 | rs11230728 | LRRC10B |
| rs10121140 | NFIB | rs10909862 | ACTRT2 | rs112370447 | COG5 |
| rs10128951 | SCARB1 | rs10930114 | CTNNA2 | rs11238907 | - |
| rs10131519 | EML1 | rs10933436 | INPP5D | rs113075218 | SRFBP1 |
| rs10131894 | EIF2B2, PGF | rs10938397 | THAP12P9, PRDX4P1 | rs113230003 | PGPEP1 |
| rs10139550 | HHIPL1 | rs10940455 | PARP8 | rs1133773 | FAM242C, FLJ12825 |
| rs12436072 | HHIPL1 | rs10947786 | KCNK5, SAYSD1 | rs1135889 | FBF1 |
| rs10237377 | PARP12 | rs10948713 | TMEM14A, TRAM2-AS1 | rs11509880 | TMEM106B |
| rs10249651 | CFTR, ASZ1 | rs10985343 | DAB2IP | rs1154988 | PCCB, MSL2 |
| rs10267593 | MAD1L1 | rs11000448 | OIT3 | rs11613352 | R3HDM2 |
| rs10268797 | MAD1L1 | rs11022738 | ARNTL | rs11637783 | MORF4L1 |
| rs10401969 | SUGP1 | rs11024074 | PLEKHA7 | rs11650066 | ASIC2 |
| rs10406816 | TGFB1 | rs11042937 | IRAG1, Y_RNA | rs11655024 | BCAS3 |
| rs10420373 | KLF2-DT | rs11057401 | CCDC92 | rs11677932 | COL6A3 |
| rs1050362 | DHX38 | rs11057830 | SCARB1 | rs11685304 | RNU6-440P, FZD7 |
| rs1050382 | PECAM1 | rs11057840 | SCARB1 | rs11690462 | SELENOI |
| rs10512861 | DNAJC13, ACAD11 | rs11066028 | ALDH2 | rs1169288 | HNF1A-AS1, HNF1A |
| rs1051860 | ARID4A | rs11072811 | MORF4L1 | rs11713141 | MRAS |
| rs1057040 | USP36 | rs11080107 | ABHD15-AS1, ANKRD13B | rs11718165 | BSN |
| rs1058588 | VAMP8 | rs11099097 | PRDM8, FGF5 | rs11718455 | RNU6-367P |
| rs10733376 | CDKN2B-AS1 | rs11099493 | RASGEF1B | rs11723436 | MAD2L1, LINC02502 |
| rs10734649 | SBF2-AS1, LINC02709 | rs11107903 | FGD6 | rs11809443 | GUK1, FAM96AP2 |
| rs10738607 | CDKN2B-AS1 | rs11124924 | APOB | rs11810571 | TDRKH |
| rs10755578 | LPA | rs11133381 | CLOCK, TMEM165 | rs11823828 | OR52E4, TRIM5 |
| rs10757274 | CDKN2B-AS1 | rs111684993 | INO80, CIBAR1P1 | rs11830157 | KSR2 |
| rs10774625 | ATXN2 | rs11170820 | FLJ12825, FAM242C | rs11838267 | C1S |
| rs1077534 | FGD5 | rs11172113 | LRP1 | rs11838776 | COL4A2 |
| rs10778546 | BTBD11 | rs12541758 | - | rs11840502 | PDS5B |
| rs10755578 | LPA | rs1254531 | - | rs1185460 | VPS11 |
| rs10793514 | LINC00841 | rs12594508 | LINC00923 | rs1193069 | COL4A1 |
| rs10797374 | ACTRT2 | rs1260387 | RPRD2 | rs11947277 | DOK7 |
| rs10806235 | LINC02542 | rs12619842 | - | rs11968400 | SNHG32 |
| rs12154802 | TBX20 | rs12625329 | RGS19 | rs11977187 | DUS4L, DUS4L-BCAP29 |
| rs12193307 | TARID | rs12645070 | SPINK2, REST | rs12019546 | MCF2L |
| rs12194592 | TARID | rs12658752 | ERAP2 | rs12028817 | ATP1B1 |
| rs12200560 | FHL5, RPS7P8 | rs12663498 | PLEKHG1 | rs12129500 | IL6R |
| rs12202017 | LINC01312, TARID | rs12675257 | RPS4XP10, GEM | rs12133641 | IL6R |
| rs1223583 | PLCE1 | rs12713421 | MIR4432HG | rs1214752 | ABCC10 |
| rs1231206 | SMG6 | rs12714757 | MITF | rs12579302 | ATP2B1 |
| rs12414777 | CNNM2 | rs12740679 | MRPS21, CIART | rs12440045 | RTF1, ITPKA |
| rs12524865 | TARID | rs12765878 | STN1 | rs12801636 | PCNX3 |
| rs12444314 | FOXL1, LINC02188 | rs12893887 | HHIPL1 | rs10186133 | RNU6-1180P, IL1F10 |
| rs12449964 | SMCR2, PEMT | rs12897 | FNDC3B | rs12936587 | SMCR2, PEMT |
| rs12469628 | ARHGAP15 | rs12905312 | CHD2 | rs12976411 | DPY19L3-DT |
| rs12482425 | - | rs1291621 | SCAF11 | rs12980942 | TGFB1 |
| rs12493885 | ARHGEF26 | rs12930452 | CFDP1 | rs12999907 | - |
| rs12495221 | RN7SL664P, CDC25A | rs13102675 | - | rs13003675 | GIGYF2 |
| rs12500824 | SHROOM3 | rs13105983 | ADAMTS3 | rs13010737 | ARHGAP15 |
| rs1250229 | FN1 | rs13118820 | MTND1P22 | rs13035055 | UBBP1, HNRNPKP2 |
| rs13232179 | WDR86-AS1, CRYGN | rs13131930 | GUCY1A1 | rs1307145 | VPS11 |
| rs13255004 | NCALD | rs13134800 | MAD2L1, LINC02502 | rs13079221 | FGD5 |
| rs13279522 | DNAJC5B | rs13139571 | GUCY1A1 | rs13082914 | KCNAB1 |
| rs1332844 | PHACTR1 | rs1317507 | MCF2L | rs13088645 | HMGB3P13, ANAPC13 |
| rs1333042 | CDKN2B-AS1 | rs13200993 | CASC15 | rs13092876 | IGF2BP2 |
| rs1333048 | CDKN2B-AS1 | rs1321309 | Y_RNA, RNU1-88P | rs1344228 | - |
| rs1333049 | CDKN2B-AS1 | rs1322052 | TEK | rs13702 | LPL |
| rs13354746 | ANKRD31, HMGCR | rs1561198 | VAMP8, VAMP5 | rs13723 | CORO6, ABHD15-AS1 |
| rs13383201 | LINC01808 | rs157595 | APOC1P1, APOC1 | rs1395821 | - |
| rs13396935 | TMEM18, LINC01875 | rs1591805 | CENPW, MIR588 | rs1412444 | LIPA |
| rs1429141 | MIR548G, PRMT5P1 | rs1623003 | NPC1 | rs1412445 | LIPA |
| rs1434538 | BMPR1B | rs1629862 | EDN1 | rs16986953 | CISD1P1, LINC01808 |
| rs143537377 | LINC02212 | rs16880442 | ITGA1, B3GNTL1P1 | rs16998073 | PRDM8, FGF5 |
| rs143723948 | PAX5 | rs16893526 | LINC01526, SNORA70 | rs170041 | SMG6 |
| rs1476098 | BCAS3 | rs1692580 | SKI, FAAP20 | rs17037390 | MTHFR |
| rs1481345 | - | rs16945184 | LINC00379, LINC00380 | rs17080091 | PLEKHG1 |
| rs1508798 | SNHG18, TAS2R1 | rs16948048 | PHB, ZNF652 | rs17081933 | REST |
| rs15285 | LPL | rs17465637 | MIA3 | rs17087335 | NOA1 |
| rs1550115 | CENPO | rs17465982 | MIA3 | rs17091891 | RPL30P9, LPL |
| rs1555321 | NRP1 | rs175040 | PGF, EIF2B2 | rs1727949 | ARHGEF26 |
| rs1800449 | LOX | rs17514846 | FURIN | rs1728918 | NRBP1, PPM1G |
| rs1800775 | CETP, HERPUD1 | rs17581137 | LINC00924 | rs17398575 | LINC02577, CCDC71L |
| rs1801251 | GIGYF2, KCNJ13 | rs17612693 | PRMT5P1, MIR548G | rs1746048 | LINC02881 |
| rs1806524 | PHF2 | rs17672135 | FMN2 | rs2011767 | FLJ40194 |
| rs1807214 | CARMAL, KRT18P47 | rs17691129 | - | rs2028900 | MAT2A |
| rs1831733 | CDKN2B-AS1 | rs1882758 | DSCAM | rs204993 | PBX2 |
| rs1842896 | GUCY1A1 | rs1888657 | KIAA1217 | rs2052923 | LINC02580, ZFP36L2 |
| rs1847331 | LINC02502, MAD2L1 | rs1892094 | ATP1B1 | rs2071382 | FES |
| rs1848599 | ARHGAP20, FDX1 | rs1893261 | TLCD5 | rs2072633 | CFB |
| rs185244 | MRAS | rs1894400 | FES | rs2073533 | ETV1 |
| rs1867624 | PECAM1, RPL31P57 | rs1903542 | RNA5SP173 | rs2074158 | DHX58 |
| rs1870634 | LINC00841 | rs1912483 | EFCAB13 | rs2074164 | KAT2A |
| rs1879454 | CFAP161 | rs1924981 | FLT1 | rs2075650 | TOMM40 |
| rs1964272 | GIPR, SNRPD2 | rs2076438 | TMEM204, IFT140 | rs2111485 | IFIH1, FAP |
| rs1966248 | LINC01312, TARID | rs2081914 | CEP120 | rs2123536 | CISD1P1 |
| rs1967604 | RNU6-1064P | rs2083458 | KRT18P47, CARMAL | rs2128739 | DYNC2H1, MIR4693 |
| rs1970112 | CDKN2B-AS1 | rs2083460 | CARMAL, KRT18P47 | rs2133189 | MIA3 |
| rs1982072 | B9D2, TMEM91 | rs2083636 | LPL | rs2140480 | RNU6-1064P |
| rs1994016 | ADAMTS7 | rs2098297 | TMED10 | rs2145598 | ARID4A |
| rs2000813 | LIPG | rs2107595 | TWIST1, HDAC9 | rs2153219 | ZC3H12D, TAB2 |
| rs2000999 | TXNL4B, HPR | rs2107713 | BCAP29, DUS4L-BCAP29 | rs216158 | MYH11 |
| rs2288874 | TGFB1 | rs2161967 | TNS1 | rs216172 | SMG6 |
| rs2184103 | LINC01767, PLPP3 | rs2281719 | GALNT2 | rs2296285 | FLT1 |
| rs2189839 | DUS4L-BCAP29, BCAP29 | rs2281727 | SMG6 | rs2306556 | GUCY1A1 |
| rs2219939 | ADAMTS7, CHRNB4 | rs2286200 | TBXAS1 | rs2327429 | TARID |
| rs222826 | RNU7-2P | rs2286526 | TBX2-AS1 | rs2337157 | DYM, SMAD7 |
| rs2229238 | IL6R | rs2452600 | PDLIM5 | rs2342781 | ZFPM2 |
| rs2229357 | INHBC | rs2454899 | LINC01950 | rs236710 | MRPS16P2, ZNF831 |
| rs2246942 | LIPA | rs246600 | ARHGAP26 | rs2380472 | C8orf34 |
| rs2250644 | LIPA | rs2472299 | CYP1A1, CYP1A2 | rs2421028 | HTRA1 |
| rs2252641 | TEX41 | rs2268344 | HTRA1 | rs2439557 | TBC1D7, GFOD1 |
| rs282546 | PARP8 | rs2269997 | PARP12, TBXAS1 | rs2166529 | RPSAP22, PARTICL |
| rs2259816 | HNF1A | rs2715408 | RNA5SP173 | rs223290 | RAB23, PRIM2 |
| rs247616 | CETP, HERPUD1 | rs2722224 | RNU6-148P, BRWD1P2 | rs2235642 | IFT140, TMEM204 |
| rs247617 | CETP, HERPUD1 | rs2727020 | TRIM77BP, UBTFL7 | rs2241714 | B9D2, TMEM91 |
| rs2487928 | JCAD | rs2738447 | LDLR | rs2244608 | HNF1A, HNF1A-AS1 |
| rs2493135 | AGT | rs2738464 | LDLR, SPC24 | rs2744427 | TAB2 |
| rs2507 | STOML1 | rs2812 | PECAM1 | rs2811686 | SLC2A12 |
| rs2546343 | PRDM6 | rs2820315 | LMOD1 | rs256277 | EPB41L4A, NREP-AS1 |
| rs2571445 | TNS1 | rs2257129 | RPL19P16, LINC01153 | rs261332 | LIPC, LIPC-AS1, ALDH1A2 |
| rs259990 | ZNF831 | rs2832227 | MAP3K7CL | rs264 | LPL |
| rs260020 | ZNF831 | rs2832232 | MAP3K7CL | rs2641438 | SMG6 |
| rs2607903 | LINC02366, YBX3 | rs2836633 | LINC00114, ERG | rs2671540 | RPL7AP50, DBF4P1 |
| rs2839812 | DYNC2H1, MIR4693 | rs2838344 | RRP1B | rs2679053 | MIR129-2, HSD17B12 |
| rs28416651 | ZFPM2-AS1, ZFPM2 | rs2681472 | ATP2B1 | rs2843152 | SKI, MORN1 |
| rs2681492 | ATP2B1 | rs28455815 | MORF4L1 | rs2880765 | AKAP13 |
| rs2848859 | - | rs2952286 | PRKAR1A, WIPI1 | rs2891168 | CDKN2B-AS1 |
| rs28522673 | LOXL1 | rs2954026 | TRIB1 | rs2895811 | HHIPL1 |
| rs28524990 | ENAM, AMBN | rs2954029 | TRIB1, LINC00861 | rs2908290 | GCK |
| rs28601761 | LINC00861, TRIB1 | rs2972146 | MIR5702, NYAP2 | rs2916260 | LRFN2 |
| rs2866611 | ADI1P1, ZHX3 | rs2980853 | TRIB1 | rs293914 | FGD5 |
| rs28709375 | MIA3 | rs3008629 | MIA3 | rs3130342 | TNXB |
| rs2876301 | PHACTR1 | rs302953 | ZFPM2 | rs3130683 | C2 |
| rs2876643 | CASC15 | rs3127599 | - | rs3184504 | ATXN2, SH2B3 |
| rs2880099 | GUCY1A1, MTND1P22 | rs2943634 | - | rs318719 | RGL3 |
| rs35146811 | CNPY4 | rs2943652 | MIR5702, NYAP2 | rs328 | LPL |
| rs35158675 | MIA3 | rs294883 | FNDC1 | rs337366 | DOK3 |
| rs35237252 | RPL30P9, LPL | rs34633411 | RNU4-17P | rs33998987 | KCNH7 |
| rs35462537 | FGD5 | rs34880628 | GIGYF2 | rs34042070 | TXNL4B, HPR |
| rs35489971 | CD300LF, RAB37 | rs34914832 | GUCY1A1 | rs34156428 | SOX6 |
| rs35885398 | SOX6 | rs34991912 | FGD5 | rs34232196 | PCSK9, BSND |
| rs360153 | SWAP70 | rs3825807 | ADAMTS7 | rs3758911 | CWF19L2 |
| rs36069016 | DOCK5 | rs3851738 | CFDP1 | rs3775058 | UNC5C |
| rs365302 | FNDC1 | rs3853444 | LINC01438, MIR297 | rs3782774 | TSPAN9 |
| rs3739998 | JCAD | rs3858704 | CUX2 | rs3786723 | SMARCA4 |
| rs3741380 | EHBP1L1 | rs3869109 | HLA-C, HCG27 | rs378825 | SWAP70 |
| rs3745348 | MYO9B | rs3895874 | GIP, IGF2BP1 | rs3796587 | GUCY1A1 |
| rs3752958 | CYP46A1 | rs3936510 | C5orf67 | rs3803802 | DNAH2 |
| rs375800732 | PTGES3P5 | rs3936511 | C5orf67 | rs3813127 | ATP7BP1 |
| rs4072980 | SF3A3, RNU6-510P | rs3993105 | ARNTL | rs4496284 | COL6A3 |
| rs41269133 | LPA, PLG | rs3995091 | HTR4 | rs4533264 | FES |
| rs4148005 | ABCA8 | rs4468572 | MORF4L1 | rs4535004 | ALK |
| rs4149309 | ABCA1 | rs4395384 | FGD5 | rs4561781 | LINC00881 |
| rs4237540 | HTRA1 | rs4403732 | PSAP, CHST3 | rs4566357 | COL4A4 |
| rs425105 | PRKD2 | rs4411878 | ADAMTS9-AS2 | rs459193 | C5orf67, RPL26P19 |
| rs4266144 | LINC00881 | rs4420638 | APOC1, APOC1P1 | rs4593108 | PRMT5P1, MIR548G |
| rs4299376 | ABCG8 | rs4458205 | GIGYF2, EFHD1 | rs460452 | PCNX4, LRRC9 |
| rs430918 | PRIM2, RAB23 | rs46522 | UBE2Z | rs460976 | MX1 |
| rs4345206 | ZNF827 | rs4704221 | CERT1 | rs4613862 | LINC02542 |
| rs4380028 | MORF4L1 | rs4719608 | MACC1, ITGB8-AS1 | rs4644046 | SLC17A2, PKHD1 |
| rs4389656 | TENT4A | rs472109 | SWAP70 | rs4646248 | NAT2, PSD3 |
| rs4394694 | DPYD | rs4723406 | TBX20 | rs518594 | LINC02881 |
| rs4743150 | ANP32B, HEMGN | rs4888378 | CFDP1 | rs539958 | SLC22A3 |
| rs4752700 | HTRA1 | rs4894803 | FNDC3B | rs55762216 | SPINK2, REST |
| rs4753788 | ASS1P13 | rs4896604 | HIVEP2 | rs55902013 | SAYSD1, KCNK5 |
| rs4754698 | ARHGAP42 | rs4906332 | MARK3, CKB | rs55940034 | COL4A2 |
| rs4757137 | ARNTL | rs4918072 | STN1, SLK | rs56015508 | KCNK5, SAYSD1 |
| rs475957 | TTC39B | rs4932371 | RN7SL363P, FURIN | rs56079296 | FTMT, SRFBP1 |
| rs4762479 | FGD6 | rs4932373 | FES | rs560810 | PHACTR1 |
| rs4766566 | CUX2 | rs4937126 | ST3GAL4 | rs56131196 | APOC1, APOC1P1 |
| rs4766578 | ATXN2 | rs4970837 | PSRC1, CELSR2 | rs56215311 | PLCG2 |
| rs4773140 | COL4A1 | rs4970926 | HORMAD1 | rs56228609 | HERPUD1, CETP |
| rs4773141 | COL4A1 | rs4977574 | CDKN2B-AS1 | rs562338 | APOB, TDRD15 |
| rs4773144 | COL4A2 | rs4977575 | CDKN2B-AS1 | rs56393506 | LPA, PLG |
| rs478442 | TDRD15 | rs501120 | LINC02881 | rs566818 | GDPD5 |
| rs4800401 | ATP7BP1 | rs512535 | APOB, TDRD15 | rs57265257 | NOA1 |
| rs4803455 | TGFB1 | rs587648 | SPATA2, SLC9A8 | rs57301765 | TWIST1, HDAC9 |
| rs4803458 | B9D2, TMEM91 | rs590121 | SERPINH1 | rs5744680 | POLK |
| rs4803459 | TMEM91 | rs5993586 | CLTCL1, HIRA | rs57481061 | ATP2B1 |
| rs4803750 | BCL3, CEACAM16-AS1 | rs59950280 | HGFAC, DOK7 | rs57677839 | PLEKHA7 |
| rs4842662 | POC1B-AS1 | rs599839 | PSRC1, CELSR2 | rs578627 | MMP7, TMEM123 |
| rs4845570 | TDRKH | rs6001960 | MRTFA | rs582384 | PRKCE |
| rs4845625 | IL6R | rs6006426 | OSM, CASTOR1 | rs585967 | TDRD15, APOB |
| rs61904693 | DYNC2H1, MIR4693 | rs602633 | PSRC1, CELSR2 | rs651821 | APOA5 |
| rs62010554 | CHRNB4 | rs603424 | PKD2L1 | rs6533530 | MIR297, LINC01438 |
| rs62139085 | SPRED2, ACTR2 | rs604723 | ARHGAP42 | rs6541297 | GALNT2 |
| rs62190384 | PID1 | rs6055069 | DEFB127, DEFB126 | rs6544713 | ABCG8 |
| rs629301 | CELSR2 | rs6060235 | EDEM2, TRPC4AP | rs6552940 | SORBS2 |
| rs633185 | ARHGAP42 | rs606452 | SERPINH1 | rs6557122 | RNU6-300P, AKAP12 |
| rs638634 | COL4A1 | rs60724295 | SLC25A21 | rs655720 | STAG1 |
| rs6426551 | PARP1, YBX1P9 | rs6088590 | NCOA6 | rs6587520 | CTSS |
| rs644045 | C2 | rs6102322 | ZHX3 | rs6593297 | CCT6A |
| rs6456496 | CASC15 | rs6102343 | ZHX3 | rs659418 | MAP6, SERPINH1 |
| rs6458545 | PHACTR1 | rs6129767 | PLCG1, ZHX3 | rs662799 | APOA5, APOA4 |
| rs646776 | CELSR2, PSRC1 | rs61403071 | FOXL1, LINC02188 | rs663129 | RNU4-17P, MC4R |
| rs6494488 | OAZ2, RBPMS2 | rs61776719 | RNU6-510P, FHL3 | rs667920 | STAG1 |
| rs6495335 | MORF4L1 | rs6504218 | PECAM1 | rs6689306 | IL6R |
| rs61848342 | CDC123, RN7SL198P | rs734780 | KRT18P47, CARMAL | rs668948 | TDRD15, APOB |
| rs6842241 | EDNRA, PRMT5P1 | rs73562014 | LINC02732 | rs6700559 | DDX59-AS1 |
| rs685031 | C2 | rs73855810 | EDNRA, PRMT5P1 | rs6704 | SERPINH1 |
| rs6876942 | LINC01377 | rs7420881 | EHBP1, RSL24D1P2 | rs67180937 | MIA3 |
| rs688359 | IGF2R | rs742115 | NEDD9 | rs67213764 | LINC01683 |
| rs6905288 | VEGFA, LINC02537 | rs745386 | KRT18P47, CARMAL | rs6736093 | MERTK, EEF1E1P1 |
| rs6905628 | LYRM2 | rs748431 | FGD5 | rs6740731 | ZEB2 |
| rs6909752 | CASC15 | rs7485656 | SCARB1 | rs67414899 | USP43 |
| rs6919211 | TARID | rs7500448 | CDH13 | rs75585236 | MYO9B |
| rs6922162 | FAM83B, RNU6-1023P | rs6743030 | RPSAP22, PARTICL | rs6810798 | MIR548G, PRMT5P1 |
| rs6930083 | RNU1-88P, Y_RNA | rs7566501 | PID1 | rs6841473 | EDNRA |
| rs6982502 | LINC00861, TRIB1 | rs7568458 | GGCX | rs6841581 | EDNRA, PRMT5P1 |
| rs699 | AGT | rs7580831 | COL6A3, COPS8 | rs7795996 | TBXAS1 |
| rs6997340 | PSD3, NAT2 | rs7586970 | TFPI | rs780094 | GCKR |
| rs701145 | GPR149 | rs7591175 | PARTICL, RPSAP22 | rs7808424 | ASZ1, CFTR |
| rs7033354 | BNC2 | rs7604403 | MERTK | rs7809950 | DUS4L-BCAP29, BCAP29 |
| rs7088704 | TSPAN14 | rs760762 | PLCG1 | rs781658 | REST |
| rs7090277 | CDC123 | rs7617773 | RN7SL664P, CDC25A | rs7842194 | GYG1P1 |
| rs7115190 | WT1 | rs7632505 | SEMA5B | rs78490412 | ARHGAP15 |
| rs7115242 | SIK3 | rs7633770 | FAM240A, ALS2CL | rs78517437 | - |
| rs7116641 | HSD17B12, MIR129-2 | rs7651039 | BTD | rs7857345 | CDKN2B-AS1 |
| rs7132794 | EMP1, LINC01559 | rs765547 | - | rs7865618 | CDKN2B-AS1 |
| rs7136259 | ATP2B1 | rs7673759 | SORBS2, RNU4-64P | rs789294 | GPR149 |
| rs7137828 | ATXN2 | rs7678555 | LINC02502, MAD2L1 | rs7910601 | WBP1L |
| rs7139170 | RPH3A | rs7680806 | SORBS2 | rs7924772 | ARHGEF12 |
| rs7145159 | ZC2HC1C, NEK9, HIF1AP1 | rs7692387 | GUCY1A1 | rs7926712 | ARNTL |
| rs7160041 | PPM1A, RPL17P2 | rs7696431 | PALLD | rs7928523 | HSD17B12 |
| rs7164479 | MORF4L1 | rs72627509 | NOA1 | rs7941600 | DENND5A |
| rs7173743 | MORF4L1 | rs7703260 | SPRY4-AS1 | rs7998440 | N4BP2L2, PDS5B |
| rs7177338 | FES | rs77620124 | C1S | rs8003602 | HHIPL1, CYP46A1 |
| rs7190256 | ZFHX3 | rs7766436 | CASC15 | rs8006867 | U3 |
| rs7199941 | PLCG2 | rs7783857 | H4P1, KLF14 | rs8027011 | CHRNB4, ADAMTS7 |
| rs7209400 | ZNF652, PHB | rs7703051 | ANKRD31, HMGCR | rs8042271 | CARMAL, KRT18P47 |
| rs7209460 | SMG6 | rs8046696 | CFDP1 | rs877549 | CASTOR1, OSM |
| rs7220401 | ABHD15-AS1, ANKRD13B | rs8046697 | CFDP1 | rs886126 | CUX2 |
| rs7229520 | SMAD7, DYM | rs8050136 | FTO | rs895953 | SETD1B |
| rs72675151 | SLC25A3P1, LINC02812 | rs8053891 | PKD1L3 | rs896655 | RN7SL151P, MTAP |
| rs7277800 | LINC00310, KCNE2 | rs8055236 | CDH13 | rs899997 | CHRNB4 |
| rs7279974 | KCNE2 | rs8057203 | CFDP1 | rs9268402 | TSBP1-AS1 |
| rs72822411 | SPRED2, ACTR2 | rs8068952 | BCAS3 | rs9283706 | MAST4 |
| rs72862334 | METAP2P1 | rs8069437 | WNT3 | rs9319428 | FLT1 |
| rs7306455 | NDUFA12 | rs8072532 | SMG6 | rs9326246 | BUD13, LINC02702 |
| rs7311541 | NDUFA12 | rs8079951 | BCAS3 | rs9351209 | LYRM2, ANKRD6 |
| rs73193808 | MAP3K7CL | rs8103278 | FOXA3 | rs9351814 | LINC01626 |
| rs73222236 | MSL2 | rs8108474 | RSPH6A | rs9367716 | RAB23, PRIM2 |
| rs7342694 | PLCG2 | rs8108632 | TGFB1 | rs9369640 | PHACTR1 |
| rs9508025 | FLT1 | rs8124182 | FTLP1, MMP9 | rs9389642 | ABRACL, REPS1 |
| rs9513112 | FLT1 | rs8141797 | SUSD2 | rs940274 | ABCA12 |
| rs9513116 | FLT1 | rs832540 | MAP3K1, SETD9 | rs944172 | RNU6-1064P, RNU6-996P |
| rs9515203 | COL4A2 | rs833509 | DYM, SMAD7 | rs944797 | CDKN2B-AS1 |
| rs9521678 | COL4A1 | rs840611 | CALCRL, RNU6-989P | rs948386 | ATP7BP1 |
| rs9546711 | MTND4P1 | rs840616 | RNU6-989P, CALCRL | rs9486719 | FHL5 |
| rs9547692 | SMAD9 | rs867186 | PROCR | rs9486729 | SEC63, SCML4 |
| rs9549328 | MCF2L | rs869396 | PALLD | rs950037 | PKN2 |
| rs9556903 | FARP1 | rs9693598 | DOCK5 | rs9939609 | FTO |
| rs9583531 | ING1, RPL21P107 | rs9720071 | RN7SKP280, INSIG1-DT | rs9951447 | ATP7BP1 |
| rs9591012 | N4BP2L2 | rs974819 | MIR4693, DYNC2H1 | rs9964304 | SMUG1P1, ACAA2 |
| rs964184 | ZPR1 | rs9751370 | RPSAP22, PARTICL | rs9969220 | CADPS2 |
| rs965652 | SLC2A12 | rs975722 | CFTR | rs9972711 | MYH11 |
| rs9902260 | PECAM1 | rs9892152 | PECAM1 | rs998584 | LINC02537, VEGFA |
| rs9912587 | VAT1 | rs9897366 | RNU6-1249P | rs9995627 | BMPR1B |
|  |  | rs9897596 | RAI1 | rs999675 | RNU6-758P, BCAR1 |
| **c. GWAS Risk variants of High SBP unique to and shared among populations** | | | | | |
| **Intersection of populations** | **RSID** | **Gene Mapped to** | **Intersection of populations** | **RSID** | **Gene Mapped to** |
| AFR:EAS | rs73324844 | BCAS3 | AFR:EAS:EUR:AMR | rs12665245 | ENPP3 |
| AFR:EAS:SAS | rs2282143 | SLC22A1 |  | rs1550576 | ALDH1A2 |
|  | rs3774427 | CACNA1D | AFR:EAS:SAS:AMR | rs77163128 | ZNF619P1, TTC4P1 |
|  | rs58318008 | SORCS2 |  | rs9814480 | CACNA1D |
|  | rs59110359 | TAB2 | EAS:AMR | rs17589290 | CCDC34P1 |
| AFR:SAS | rs77932157 | CTNNA3 | EAS:SAS:AMR | rs17004869 | FGF5 |
| EUR:AMR | rs72934583 | NBEAL1 |  | rs58244401 | FGD5 |
| AFR:EAS:SAS:EUR | rs12231049 | MYL2 | AFR | rs10026364 | LINC02462 |
|  | rs3782889 | MYL2 |  | rs76987554 | TARID |
|  | rs471678 | MRPL23 |  | rs78192203 | GPR20 |
| SAS:EUR:AMR | rs10067451 | LINC00461 |  | rs115795127 | FRMD3 |
|  | rs10490923 | ARMS2 |  | rs143255889 | LINC01519 |
|  | rs11039149 | NR1H3 |  | rs185169399 | LINC02241 |
|  | rs112963849 | HIC1 |  | rs7016717 | - |
|  | rs12423664 | FBRSL1 |  | rs61503757 | ZNF107, ZNF138 |
|  | rs1799945 | HFE |  | rs6750839 | SEPHS1P7 |
|  | rs34328549 | INSR |  | rs7006531 | CDH17 |
|  | rs3762515 | EFEMP1 |  | rs76702537 | LINC02468 |
|  | rs62172472 | RN7SKP42 |  | rs7153038 | NUMB |
|  | rs62286033 | LINC01839, LINC01840 |  | rs190531342 | UGCG, RNU6-710P |
| EUR | rs11537751 | PTPMT1, NDUFS3 |  | rs115476423 | ZNF767P, ZNF746 |
|  | rs13107325 | SLC39A8 |  | rs76967376 | MYO5B |
|  | rs34592089 | BANK1 |  | rs10279895 | HNRNPA1P73, RPL35P4 |
| EAS:SAS | rs200741614 | SLC6A15, RPL6P25 | AFR:EUR:AMR | rs7801190 | SLC12A9 |
|  | rs2305013 | ARHGEF12 |  | rs16982520 | ZNF831 |
|  | rs2515920 | UQCRHP1, NCR3 |  | rs71427857 | DNAJC10 |
|  | rs28495639 | USP8 |  | rs73007615 | - |
|  | rs769177 | LTB, TNF |  | rs11563582 | HNRNPA1P73, RPL35P4 |
| AFR:SAS:EUR:AMR | rs10096908 | ANK1 | EAS | rs11066280 | HECTD4 |
|  | rs11657730 | BAHCC1 |  | rs11066453 | OAS1 |
|  | rs11685652 | CHORDC1P1 |  | rs12229654 | CUX2, LINC01405 |
|  | rs11914354 | TMEM212, TMEM212-AS1 |  | rs2021783 | TNXB |
|  | rs12258967 | CACNB2 |  | rs2072134 | OAS3 |
|  | rs12941507 | AMZ2P1, GNA13 |  | rs2074356 | HECTD4 |
|  | rs2493292 | PRDM16 |  | rs671 | ALDH2 |
|  | rs56012466 | PRKAG2 |  | rs77768175 | HECTD4 |
|  | rs56352102 | SBF2 |  | rs78813487 | GAB4 |
|  | rs6015450 | ZNF831 |  | rs79105258 | CUX2 |
|  | rs7174250 | ABHD17C |  | rs79549409 | GBX1 |
|  | rs991316 | ADH7, ADH1B | SAS | rs79455834 | - |
| EAS:SAS:EUR:AMR | | | | | |
| rs10184428 | KNOP1P3, PRPS1P1 | rs12129649 | MOV10 | rs62525059 | GML |
| rs10776752 | WNT2B | rs12219304 | NT5C2 | rs7120548 | MTCH2 |
| rs10833346 | HMGB1P40, PRMT3 | rs12413409 | CNNM2 | rs72914247 | DCC |
| rs10895001 | RN7SL222P, PPIAP43 | rs12752401 | - | rs732998 | NT5C2 |
| rs10980694 | LPAR1 | rs1378942 | CSK | rs78423498 | - |
| rs11002605 | LINC00856, ZMIZ1-AS1 | rs1458038 | PRDM8, FGF5 | rs8184986 | CHEK2 |
| rs11099098 | PRDM8, FGF5 | rs1530440 | CABCOCO1 | rs820430 | SLC4A7, RNU1-96P |
| rs11191548 | CNNM2 | rs2306899 | DDHD2 | rs9349379 | PHACTR1 |
| rs11191580 | NT5C2 | rs2398162 | NR2F2-AS1 | rs9462492 | KCNK5 |
| rs11191593 | NT5C2 | rs253447 | SPRY4-AS1 | rs3858414 | ARHGAP42 |
| rs112913898 | NT5C2 | rs35085068 | PRMT5, HAUS4 | rs4846567 | ZC3H11B, LYPLAL1-AS1 |
| rs11619475 | ITM2B, POLR2KP2 | rs35473622 | ARHGAP42 | rs55823223 | TRIM65 |
| rs11674660 | ARNILA, DNMT3A | rs3790604 | WNT2B |  |  |
| AFR:EAS:SAS:EUR:AMR | | | | | |
| rs10021303 | BMPR1B | rs6590811 | ARHGAP42 | rs7554672 | - |
| rs10033366 | ENPEP | rs6590812 | ARHGAP42 | rs7584120 | PNPT1 |
| rs1004467 | CYP17A1 | rs6590816 | ARHGAP42 | rs7625237 | TKT |
| rs10069554 | LINC02160 | rs6596140 | FSTL4 | rs7651190 | ULK4 |
| rs10103692 | RNU105C, RN7SL250P | rs661348 | LSP1 | rs7654819 | - |
| rs1027989 | LYN | rs66978877 | PGPEP1 | rs7715779 | NPR3 |
| rs10495809 | LINC01320 | rs6712094 | PRPS1P1 | rs7756992 | CDKAL1 |
| rs10745332 | CAPZA1 | rs675026 | OPRM1 | rs7763581 | GMDS, FOXC1 |
| rs10756197 | - | rs6822892 | PDGFC | rs782513 | AKR1B10, AKR1B1 |
| rs10774625 | ATXN2 | rs6825911 | ENPEP | rs7893462 | ODAD2 |
| rs10786772 | SH3PXD2A | rs6871246 | MAP3K1 | rs7914095 | FANK1 |
| rs10828266 | DNAJC1 | rs6940540 | PLEKHG1 | rs7924431 | FOLH1 |
| rs10838835 | OR4B2P, OR4B1 | rs6969780 | HOXA-AS2, HOXA3 | rs79787332 | EIF4EP4 |
| rs10849937 | PHETA1, CUX2 | rs6987059 | LINC00529, XKR6 | rs8036030 | SEMA7A |
| rs10857147 | FGF5, PRDM8 | rs7004825 | XKR6 | rs8039305 | FURIN |
| rs10859915 | PGAM1P5 | rs704 | VTN, SARM1 | rs805293 | LY6G6C, MPIG6B |
| rs10948071 | ZNF318 | rs7115856 | HSD17B12 | rs805303 | BAG6 |
| rs11014166 | CACNB2 | rs7136259 | ATP2B1 | rs8068318 | TBX2-AS1, TBX2 |
| rs1105956 | RSPO3, RPS4XP9 | rs7199751 | FOXC2, FOXL1 | rs8098380 | ENOSF1, YES1 |
| rs11067763 | LINC02463, RN7SL865P | rs72680374 | LINC02319 | rs8101673 | MYO9B |
| rs1110183 | ARMC8P1, TCEA1P3 | rs72806698 | EML6 | rs830179 | KLHL24 |
| rs11105364 | ATP2B1 | rs72850439 | SBF2 | rs880315 | CASZ1 |
| rs11105368 | ATP2B1 | rs7297206 | ATP2B1 | rs891511 | NOS3 |
| rs11204676 | MCL1, ENSA | rs7302981 | CERS5 | rs932764 | PLCE1 |
| rs11229457 | OR5B12 | rs7372217 | ULK4 | rs936226 | CSK, CYP1A2 |
| rs11230728 | LRRC10B | rs7406910 | HOXB7 | rs9373768 | - |
| rs11249906 | PPP1R3B | rs742047 | RSL24D1P1, GPR89P | rs9375459 | - |
| rs11249945 | TNKS | rs7497304 | FES | rs9385284 | CLVS2 |
| rs112587279 | RIBC2 | rs751984 | LRRC10B | rs9390459 | STXBP5 |
| rs1126464 | DPEP1 | rs4910498 | SWAP70 | rs9603420 | RXFP2 |
| rs1133400 | INPP5A | rs4937126 | ST3GAL4 | rs9683944 | RPS23P2, PCDH18 |
| rs11646213 | CDH13 | rs4980389 | LSP1 | rs972283 | KLF14, H4P1 |
| rs1173727 | LINC02120, NPR3 | rs5006548 | FGD4 | rs9787901 | CHST1, MIR7154 |
| rs1173771 | LINC02120, NPR3 | rs55794313 | ZNF385D | rs9810888 | CACNA1D |
| rs11749255 | MSX2, HIGD1AP3 | rs55897749 | DCAKD, NMT1 | rs9907379 | LINC02875 |
| rs11754682 | - | rs55940034 | COL4A2 | rs9932220 | RN7SKP142, LINC01571 |
| rs11775334 | MSRA | rs56373378 | MTCO1P57 | rs3732103 | SLC66A3 |
| rs11857726 | CHP1 | rs569550 | LSP1 | rs3744760 | PLCD3 |
| rs11977526 | FTLP15, IGFBP3 | rs5762197 | - | rs3766160 | CELA2B |
| rs11998678 | DEFB136, OR7E161P | rs6021247 | NFATC2 | rs3800688 | PODXL |
| rs12046278 | CASZ1 | rs604723 | ARHGAP42 | rs3808869 | ARID3C |
| rs12144175 | RIT1 | rs62434120 | PLEKHG1 | rs3942852 | PTPRJ |
| rs1214776 | MSH2 | rs62524579 | CYP11B2, LY6E-DT | rs403814 | L3MBTL4 |
| rs12149202 | GSE1 | rs633185 | ARHGAP42 | rs4247284 | ESRP1 |
| rs12228810 | AACS, BRI3BP | rs6418 | CYP11B2, GML | rs433750 | - |
| rs1242765 | UNC79 | rs642961 | IRF6, UTP25 | rs4373814 | SLC39A12, CACNB2 |
| rs12443113 | RASL12 | rs6461992 | HOXA11 | rs4387287 | STN1 |
| rs12579302 | ATP2B1 | rs6487504 | LINC00592, LMNTD1 | rs4409766 | BORCS7-ASMT, BORCS7 |
| rs12579720 | LINC02398 | rs6495127 | MPI, FAM219B | rs4462906 | Y_RNA, RPL32P3 |
| rs12630213 | FGD5 | rs6506537 | YES1 | rs459193 | C5orf67, RPL26P19 |
| rs12679242 | CYP11B2 | rs6536088 | GUCY1B1, GUCY1A1 | rs4630220 | SH3PXD2A |
| rs12715461 | CACNA1D | rs6586390 | TARBP1, LINC01354 | rs4665630 | KLHL29 |
| rs1275923 | KCNK3 | rs2675149 | GACAT3 | rs4678408 | MRAS, NME9 |
| rs1275988 | RPL37P11, KCNK3 | rs2681472 | ATP2B1 | rs4684242 | FGD5 |
| rs1285841 | CCDC88C | rs2681492 | ATP2B1 | rs4690974 | MTND1P22 |
| rs12902197 | MIR4713HG | rs2725371 | PURG | rs4691380 | PDGFC |
| rs13001925 | RPL7P13, RNU6-439P | rs27434 | ERAP1 | rs4705752 | MCC |
| rs13002573 | - | rs277152 | B3GLCT, RXFP2 | rs4722675 | HOTTIP |
| rs13062241 | FGD5 | rs2782643 | SZT2 | rs4728142 | IRF5, KCP |
| rs13112725 | NPNT | rs2814949 | ILRUN | rs4757391 | SOX6 |
| rs13139571 | GUCY1A1 | rs2820037 | - | rs4785340 | ZNF423 |
| rs13143871 | GUCY1A1 | rs28379706 | PLXNB2 | rs4843553 | C16orf95 |
| rs13144136 | CLNK | rs284844 | WBP1L | rs4888372 | RNU6-758P, BCAR1 |
| rs13149993 | PRDM8, FGF5 | rs2856830 | HLA-DPA1 | rs2217560 | CBLN2 |
| rs13266183 | RN7SL250P, RNU105C | rs2867695 | ANTXR2, RPSAP39 | rs2229840 | NCOR2 |
| rs1327235 | LINC02871, FAT1P1 | rs28724242 | HLA-DQB1 | rs2240736 | TBX2, TBX2-AS1 |
| rs1333047 | CDKN2B-AS1 | rs28792763 | LINC00928, TICRR | rs2251473 | MTMR9 |
| rs13333226 | UMOD | rs2932538 | MOV10 | rs2254613 | RN7SKP21, MBIP |
| rs1401982 | ATP2B1 | rs2972146 | MIR5702, NYAP2 | rs2257205 | TSPOAP1-AS1, RNF43 |
| rs1442386 | DLGAP1 | rs3130482 | SLC44A4 | rs2270860 | CRIP3, SLC22A7 |
| rs1446477 | FIGN, PRPS1P1 | rs31864 | EBF1 | rs2274224 | PLCE1, PLCE1-AS1 |
| rs1446802 | EXTL2P1, RPS15AP14 | rs324498 | PTPRD | rs2293579 | PSMC3 |
| rs147428270 | ULK4 | rs35021474 | KCNK3 | rs2303655 | ZNF474-AS1 |
| rs1475591 | UBE3AP2, TIAM1 | rs35434 | Y_RNA | rs2304615 | REXO1 |
| rs1563788 | ZNF318 | rs35443 | UBA52P7, TBX3 | rs2320299 | LCORL |
| rs1580004 | AMPD3 | rs35444 | UBA52P7, TBX3 | rs2326187 | LINC02523 |
| rs16849211 | - | rs36034102 | FGF5 | rs2341599 | MTND1P22 |
| rs16849225 | FIGN, PRPS1P1 | rs366178 | RPL10P19, RNU6-682P | rs2342883 | FGD5 |
| rs16849273 | PRPS1P1 | rs1902859 | FGF5, PRDM8 | rs2392929 | CCDC71L, LINC02577 |
| rs16896398 | SLC22A7, TTBK1 | rs1908127 | - | rs2517521 | HCG22 |
| rs16905753 | OR4X1 | rs1948948 | HNRNPA1P48, UNGP1 | rs2521501 | FES |
| rs16934621 | BNC2 | rs1973765 | LSP1 | rs2524095 | LINC02571 |
| rs16998073 | PRDM8, FGF5 | rs2044693 | PNO1 | rs2586886 | KCNK3 |
| rs17030613 | CAPZA1 | rs2046301 | OR4C4P, OR4C3 | rs2596473 | LINC01149, HCP5 |
| rs17046380 | EML6 | rs2067087 | HOTTIP | rs2643826 | RNU1-96P, Y_RNA |
| rs17249754 | ATP2B1 | rs2075571 | THBS3, THBS3-AS1 | rs2649044 | SWAP70 |
| rs1731243 | KCNK3 | rs2125067 | GDF10 | rs26653 | ERAP1 |
| rs1731249 | KCNK3 | rs217727 | H19 | rs1872167 | NUP160 |
| rs1735151 | IGSF5 | rs2206734 | CDKAL1 | rs1887320 | FAT1P1, LINC02871 |
| rs17367504 | MTHFR | rs2212606 | ERG, LINC00114 | rs1898841 | PRPS1P1 |
| rs17514846 | FURIN | rs1859168 | HOTTIP | rs1799998 | LY6E-DT, CYP11B2 |
| rs180912 | NHLRC2, ADRB1 |  |  |  |  |
| **d. GWAS Risk variants of T2D unique to and shared among populations** | | | | | |
| **Intersection of populations** | **RSID** | **Gene Mapped to** | **Intersection of populations** | **RSID** | **Gene Mapped to** |
| AFR:EAS:EUR | rs3769873 | COBLL1 | AFR:AMR | rs116549635 | ASCL2 |
| AFR | rs11466334 | TGFB1 |  | rs16858462 | TNS1 |
|  | rs11873305 | MC4R | AFR:EAS | rs16955379 | CMIP |
|  | rs12277475 | MIR4686, ASCL2 |  | rs4632135 | GP2 |
|  | rs13424957 | RNA5SP111, RNA5SP110 |  | rs66477705 | Y_RNA, LINC01871 |
|  | rs16881572 | PKHD1 |  | rs7560163 | - |
|  | rs17054480 | PALLD |  | rs77402029 | KCNQ1, KCNQ1OT1 |
|  | rs1819564 | PKHD1 | AFR:EAS:AMR | rs11199826 | LINC01153, RN7SKP167 |
|  | rs202178099 | PARP8, LINC02106 |  | rs13342232 | SLC16A11 |
|  | rs3842770 | INS-IGF2 |  | rs138997002 | KRT18P9, CYCSP55 |
|  | rs4072825 | TH |  | rs186568031 | SLC16A11, CLEC10A |
|  | rs57261374 | - |  | rs2106463 | KCNQ1, KCNQ1OT1 |
|  | rs60461843 | SLC30A8 |  | rs2237897 | KCNQ1 |
|  | rs6506284 | LINC01892, PPIAP14 |  | rs2288176 | TNS1 |
|  | rs73284431 | AGMO |  | rs2780215 | NUDT3, RPL35P2 |
|  | rs7364276 | - |  | rs3792615 | MARCHF1 |
|  | rs73872717 | ZBTB38 |  | rs7071036 | RPL19P16, LINC01153 |
|  | rs73954691 | LIMS2 |  | rs73239895 | CLEC10A, SLC16A11 |
|  | rs74073568 | FYB2 |  | rs8181588 | KCNQ1 |
|  | rs74452128 | MC4R | AFR:EAS:EUR:AMR | rs2237892 | KCNQ1 |
|  | rs74927455 | ST18 |  | rs2283228 | KCNQ1 |
|  | rs76197067 | CCDC68 | AFR:EAS:SAS | rs11033115 | SLC1A2 |
|  | rs76859863 | KATNAL1, LINC00385 |  | rs2421897 | SLC1A2 |
|  | rs7747641 | RPL12P23 |  | rs3773506 | PLS1 |
|  | rs80240198 | RPL32P14 |  | rs58090211 | SLC1A2 |
|  | rs9478961 | MTHFD1L, RNU6-300P |  | rs7304270 | CCND2-AS1, CCND2 |
|  | rs9842137 | SUMF1 |  | rs9505085 | RREB1 |
| AFR:EAS:SAS:AMR | rs11114650 | LIN7A | AFR:EUR | rs111973643 | FCGR2A, RNU6-481P |
|  | rs112915006 | MOV10L1, PANX2 |  | rs16988333 | HORMAD2 |
|  | rs1374910 | IGF2BP2 |  | rs28265 | ASCC2 |
|  | rs17797882 | - |  | rs3845281 | OTULINL |
|  | rs28599782 | MOB1B |  | rs41278853 | HORMAD2-AS1, MTMR3 |
|  | rs3864095 | MASP1 |  | rs56392746 | HORMAD2-AS1 |
|  | rs3869115 | HLA-C, HCG27 | AFR:EUR:AMR | rs10173251 | THADA |
|  | rs60089934 | TRIB1 |  | rs113135335 | MIR4435-2HG, BCL2L11 |
|  | rs6815464 | MAEA |  | rs11655898 | ERN1 |
|  | rs6919504 | H3P28, SNORD28B |  | rs13414381 | THADA |
|  | rs72501964 | CTBP1-DT |  | rs13426680 | CYTIP |
|  | rs73541251 | GP2 |  | rs150017767 | SLC13A3 |
|  | rs7674402 | MOB1B |  | rs35720761 | THADA |
|  | rs79407053 | CTBP1-DT |  | rs57767539 | ERN1 |
| AFR:EAS:SAS:EUR | rs28792187 | STAU2 |  | rs60276348 | ERN1 |
|  | rs7046845 | AOPEP |  | rs62080313 | - |
| AFR:SAS | rs11820019 | CCND1 |  | rs6426514 | RHOU |
|  | rs12885036 | SPTB |  | rs6743071 | THADA |
|  | rs1531583 | PCGF3 |  | rs7568172 | CYTIP |
|  | rs1575972 | - |  | rs7636 | ACHE |
|  | rs17053082 | SGCD |  | rs7957197 | OASL |
|  | rs2031847 | MED20, USP49 | AFR:SAS:EUR | rs34340810 | ZFPM2 |
| AFR:SAS:AMR | rs11034036 |  | AMR | rs117483894 | TCF12 |
|  | rs3918298 | CCND1 |  | rs137893140 | CDKN2B-AS1 |
|  | rs55816909 | CCND1 |  | rs149265787 | JPH1 |
| EAS | rs10305745 | ARNT |  | rs149483638 | IGF2, INS-IGF2 |
|  | rs11066453 | OAS1 |  | rs17737404 | ASCL2, MIR4686 |
|  | rs117267808 | GP2 |  | rs1800961 | HNF4A |
|  | rs117414485 | C10orf67 |  | rs2283159 | KCNQ1 |
|  | rs117601636 | KCNQ1, KCNQ1OT1 |  | rs75307421 | DEPDC5 |
|  | rs117737118 | GRM8 |  | rs76920233 | LINC01413, LINC00926 |
|  | rs12229654 | CUX2, LINC01405 | EAS:AMR | rs111765639 | JMJD1C |
|  | rs16907058 | NELL1 |  | rs16884025 | PARP8 |
|  | rs17866443 | ZNF800 |  | rs16902871 | RANBP3L |
|  | rs2074356 | HECTD4 |  | rs17013314 | UBE2E2 |
|  | rs2233580 | PAX4 |  | rs2237896 | KCNQ1 |
|  | rs3735567 | JAZF1 |  | rs34642578 | ASAH1 |
|  | rs61342118 | SND1, MIR129-1 |  | rs4711389 | SMIM29 |
|  | rs74334916 | PARP8 |  | rs59050225 | JMJD1C |
|  | rs77065181 | C10orf67, PTF1A |  | rs74577409 | CLEC10A, SLC16A11 |
|  | rs77768175 | HECTD4 |  | rs75418188 | SLC16A11 |
|  | rs77978149 | FAIM2 |  | rs75493593 | SLC16A11 |
|  | rs79826452 | IGF1R | EAS:EUR1 | rs113036477 | RMST |
|  | rs9349064 | BTBD9 |  | rs146716733 | ARID5B |
| EAS:EUR:AMR | rs11181613 | LINC02451 |  | rs75872811 | ARID5B |
|  | rs113154802 | PTCH1 |  | rs77864822 | RMST |
|  | rs113547729 | ZZEF1 | EAS:SAS | rs10993738 | SYK |
|  | rs11763876 | C1GALT1 |  | rs12236906 | AOPEP |
|  | rs12027542 | PCNX2 |  | rs12437434 | NYNRIN |
|  | rs17513135 | PABPC4, PABPC4-AS1 |  | rs147538848 | SINHCAF |
|  | rs17815608 | E2F7 |  | rs17045328 | CR2 |
|  | rs3768321 | PABPC4-AS1, PABPC4 |  | rs28638142 | SNORA70, OASL2P |
|  | rs72802342 | CTRB2, ZFP1 |  | rs55783344 | HNF1A |
|  | rs870992 | ITGA1 |  | rs59020573 | KIAA1522, SYNC |
| EAS:SAS:AMR | rs144052331 | CEP120 |  | rs61967710 | HS6ST3 |
|  | rs2303720 | CEP120 |  | rs72751723 | EMB |
|  | rs3765467 | GLP1R |  | rs76541615 | HCG22 |
|  | rs60054445 | ADCY5, HACD2 |  | rs80234489 | SINHCAF |
|  | rs74790763 | PRDM6, CEP120 |  | rs9552911 | SGCG, SACS |
|  | rs77741372 | AP3B1 | EAS:SAS:EUR | rs10101067 | EYA1 |
|  | rs79223353 | ADCY5 |  | rs13059382 | CASR |
|  | rs79490558 | GTF2F2, TPT1 |  | rs16993330 | LARGE1, LINC01643 |
|  | rs9380826 | GLP1R |  | rs60251368 | BRAF |
|  | rs9394574 | GLP1R |  | rs6633421 | CNKSR2 |
| EUR | rs111824905 | STX12 |  | rs75990271 | FAM185A, RASA4DP |
|  | rs112324411 | MEG3 | EUR:AMR | rs11716527 | U3 |
|  | rs113414093 | CRYBA2, MIR375 |  | rs12427353 | HNF1A |
|  | rs117001013 | YWHAH |  | rs13085136 | SHQ1 |
|  | rs11848361 | UNC79 |  | rs13166103 | PLK2 |
|  | rs12140153 | PATJ |  | rs17265513 | ZHX3 |
|  | rs13107325 | SLC39A8 |  | rs17334919 | THADA |
|  | rs149027146 | OR4C9P, OR4R1P |  | rs17772814 | CASC11 |
|  | rs17662402 | RNU6-526P |  | rs302864 | IGBP1P2, TEX14 |
|  | rs35164294 | L3MBTL3, TMEM244 |  | rs362307 | HTT |
|  | rs35169799 | PLCB3 |  | rs62262091 | SMARCC1 |
|  | rs3900856 | C5orf67 |  | rs62370480 | FST |
|  | rs41277236 | NEUROG3 |  | rs71372253 | RNF135, MIR4733HG |
|  | rs62059712 | KDM6B |  | rs72802365 | CTRB2, CTRB1 |
|  | rs62271373 | TSC22D2, LINC01214 |  | rs72836348 | BCL2L11, MIR4435-2HG |
|  | rs72692804 | SV2A, SF3B4 |  | rs75332279 | ONECUT1, RPSAP55 |
|  | rs72692805 | SF3B4, SV2A |  | rs76675804 | THADA |
|  | rs72926932 | TCF4 |  | rs80147536 | THADA |
|  | rs72928978 | TPCN2 | SAS:AMR | rs12494424 | SAMMSON |
|  | rs73146095 | CGGBP1, ZNF654 | SAS | rs13081389 | GSTM5P1, SYN2 |
|  | rs75401573 | AP1B1, RFPL4AP6 |  | rs144245804 | CCND1 |
|  | rs75423501 | KIF9-AS1 |  | rs17036101 | GSTM5P1, SYN2 |
|  | rs75686861 | HHIP |  | rs1800574 | HNF1A-AS1, HNF1A |
|  | rs79687284 | PROX1-AS1 |  | rs55911137 | CCND1 |
|  | rs9844972 | TSC22D2, LINC01214 |  | rs73221116 | PCGF3 |
| SAS:EUR | | | | | |
| rs11065299 | SPPL3 | rs17405722 | STAT3, CAVIN1 | rs112515915 | ZFPM2 |
| rs11108094 | USP44 | rs56981400 | EIF4E3 | rs13257283 | ZFPM2 |
| rs78020297 | FTO |  |  |  |  |
| SAS:EUR:AMR | | | | | |
| rs10512488 | BECN1 | rs17122772 | SLC7A7 | rs2727301 | GH2, CSH1 |
| rs10848958 | PARP11-AS1 | rs17496664 | FILIP1 | rs34715063 | RASGRP1, LINC02694 |
| rs10973627 | SHB | rs17608340 | LMO3, SKP1P2 | rs35011184 | TCF7L2 |
| rs114447556 | - | rs17746147 | VTI1A | rs35264875 | TPCN2 |
| rs11639470 | C15orf54 | rs17791513 |  | rs55691245 | ACSL1 |
| rs116913033 | CAMK2B | rs17802463 | DTNB | rs58730668 | ACSL1 |
| rs11709077 | PPARG | rs17810376 | GLP2R | rs61881115 | MYEOV, SMIM38 |
| rs12128213 | LINC00862, CCNQP1 | rs1801282 | PPARG | rs62482405 | PSMC2 |
| rs12185519 | UHRF1, KDM4B | rs2012444 | PPARG | rs71304101 | PPARG |
| rs12736701 | AK5 | rs2065703 | CDK5RAP1 | rs72695645 | ACSL1 |
| rs12912777 | RASGRP1 | rs2189301 | HNF1B | rs735949 | ACSL1 |
| rs12933120 | SLX4 | rs2283164 | KCNQ1 | rs78761021 | GLP2R |
| rs17036160 | PPARG | rs2408252 | ANO6 | rs9852406 | NDUFS6P1 |
| EAS:SAS:EUR:AMR | | | | | |
| rs10048404 | WDR7 | rs17072370 | SPRY2, LINC01080 | rs4826580 | ZXDB, FAAH2 |
| rs10305420 | GLP1R | rs17105012 | RN7SKP17, LINC01629 | rs4833687 | PRDM5 |
| rs1043246 | ATP2A3 | rs17294565 | SGCZ | rs4834232 | LARP1B |
| rs10460009 | LPIN2 | rs174541 | FADS2 | rs4846569 | ZC3H11B |
| rs10771372 | RN7SKP15 | rs17584499 | PTPRD | rs4922793 | BDNF |
| rs10811660 | CDKN2B-AS1, DMRTA1 | rs17791483 |  | rs4923864 | CCDC9B, PHGR1 |
| rs10811661 | DMRTA1, CDKN2B-AS1 | rs1802295 | VPS26A | rs4924455 | CCDC9B |
| rs10821311 | MIRLET7A1HG, LINC02603 | rs1872635 | SMUG1 | rs4996963 | WFS1 |
| rs10830963 | MTNR1B | rs1917717 | RPL3P8, IMMP2L | rs56218782 | MAML3 |
| rs10841868 | LDHB, GYS2 | rs1966265 | FGFR4 | rs56281442 | SHROOM3 |
| rs10842994 | PTHLH, RN7SKP15 | rs201018682 | EIF5A2, KLF7P1 | rs56365443 | MARK3 |
| rs10965247 | CDKN2B-AS1 | rs2063640 | RNU6-461P, ZPLD1 | rs564398 | CDKN2B-AS1 |
| rs10965250 | DMRTA1, CDKN2B-AS1 | rs2129869 | ITPR2, SSPN | rs56687477 | KCNU1 |
| rs10993072 | MIRLET7A1HG, PTPDC1 | rs2290402 | TMEM175 | rs58432198 | FAF1 |
| rs11046164 | LDHB | rs2294120 | ZNF34 | rs58542926 | TM6SF2 |
| rs11150745 | RPTOR | rs2296172 | MACF1 | rs59457600 | LINC02030 |
| rs11199755 | RPL19P16 | rs2296173 | MACF1 | rs601945 | HLA-DQA1, HLA-DRB1 |
| rs11258422 | BEND7 | rs2382249 | EPC2 | rs61779275 | MACF1 |
| rs1127787 | MRPS35 | rs243513 | CUL1 | rs61779284 | MACF1 |
| rs113810779 | HNF4A | rs2526678 | FADS2 | rs61910828 | CCND2-AS1, HSPA8P5 |
| rs11545861 | GRIPAP1 | rs2608953 | MED23 | rs62007683 | MARK3 |
| rs11561066 | PHKG1P3, GTF2IP11 | rs2722769 | MTND5P21, LINC02752 | rs62075585 | CYTH1 |
| rs11602873 | ARAP1 | rs279744 | ARL15 | rs62255926 | RFT1 |
| rs11842871 | HMGB1 | rs2820443 | ZC3H11B, LYPLAL1-AS1 | rs62405419 | TFAP2B |
| rs12441261 | RGMA, SEPHS1P2 | rs2820446 | ZC3H11B | rs62481355 | GCC1, CACNA2D1 |
| rs12496230 | - | rs2862954 | ERLIN1 | rs6714523 | BMPR2, RN7SL40P |
| rs12505942 | MAML3 | rs303760 | RMC1 | rs6976111 | CTTNBP2 |
| rs12527410 | UNC5CL, LRFN2 | rs34329895 | PLEKHM3 | rs7041847 | GLIS3 |
| rs12613372 | GALNT14, CAPN13 | rs34454109 | LINC01524 | rs7109575 | ARAP1 |
| rs12669521 | GSAP, GCNT1P5 | rs34855406 | RETREG3, PSMC3IP | rs71495046 | LINC00838 |
| rs12688220 | EEF1A1P40, MORC4 | rs35004890 | STK11 | rs72631105 | LINC01153, RPL19P16 |
| rs12932337 | RPL13 | rs35668226 | RPL19P16 | rs72951506 | NUS1 |
| rs12945601 | RAI1 | rs35753840 | SGCZ | rs72951548 | NUS1, SLC35F1 |
| rs13005841 | ERBB4 | rs36051838 | LINC00838 | rs7313668 | TSPAN8, PTPRR |
| rs13007524 | - | rs36062478 | PTEN | rs73226048 | MYCLP2, KRT8P17 |
| rs13036344 | COMMD7 | rs3742305 | HMGB1 | rs747360 | ZZEF1 |
| rs1316776 | DMGDH | rs3751236 | RN7SKP15 | rs75080135 | GGNBP1 |
| rs13239186 | CTTNBP2 | rs3751237 | RN7SKP15 | rs755249 | BMP8A, PPIEL |
| rs13262861 | ANK1, NKX6-3 | rs3751239 | GRIP1, RN7SKP15 | rs7566955 | RN7SL283P, LINC01853 |
| rs13268508 | HSF1 | rs3751297 | FBRSL1 | rs7572857 | CEP68 |
| rs13406624 | LINC01931, MMADHC-DT | rs3757969 | SCRT1, DGAT1 | rs75772194 | SMARCE1P4, KCNU1 |
| rs1408579 | ERLIN1 | rs3764002 | WSCD2 | rs76789970 | SBF2 |
| rs1412830 | CDKN2B-AS1 | rs3862791 | STARD10, ARAP1 | rs77106637 | ARAP1, STARD10 |
| rs142385484 | FCGRT | rs3916765 | MTCO3P1, HLA-DQB3 | rs77464186 | ARAP1 |
| rs1426371 | WSCD2 | rs4273712 | RNU6-200P, PRELID1P1 | rs7923866 | Y_RNA |
| rs152839 | PARP8 | rs4397977 | MIR3168, RN7SL597P | rs7988007 | - |
| rs1552224 | ARAP1 | rs4709746 | RN7SL366P, QKI | rs79976124 | ADH5P4 |
| rs16826069 | MACF1 | rs4729854 | RASA4DP, FAM185A | rs80005921 | GTF3AP5, RAPGEF5 |
| rs16829174 | EPC2 | rs4771648 | IRS2 | rs80196932 | NEPNP, NUS1 |
| rs16861329 | ST6GAL1 | rs4817543 | IFNAR2, LINC01548 | rs816750 | RIT2, SYT4 |
| rs9379084 | RREB1 | rs9358912 | H4C5, H2BC8 | rs9273401 | HLA-DQB1, HLA-DQA1 |
| rs9568868 | LINC00558, ZNF646P1 | rs9370243 | LRRC1, MLIP | rs9275184 | MTCO3P1, HLA-DQB1 |
| rs9569864 | - |  |  |  |  |
| AFR:SAS:EUR:AMR | | | | | |
| rs10145154 | NRXN3 | rs17689007 | MSRA | rs663640 | RNU4-17P |
| rs10146997 | NRXN3 | rs17818197 | EBF2 | rs66502159 | CTRB1, CTRB2 |
| rs10159026 | LINC02790 | rs17836088 | NRXN3 | rs67013744 | CHD4 |
| rs10203174 | THADA | rs1899951 | PPARG | rs67269808 | CHCHD2P9 |
| rs1060105 | SBNO1 | rs2410767 | TMEM161B-AS1, LINC02060 | rs6795197 | PXYLP1, ZBTB38 |
| rs10923931 | NOTCH2 | rs2453051 | NOTCH2 | rs6870983 | TMEM161B-AS1 |
| rs11063069 | CCND2-AS1 | rs2493394 | NOTCH2 | rs7144011 | NRXN3 |
| rs112667817 | ETF1, RPL7P19 | rs2613499 | RPL31P12, NEGR1 | rs7156625 | NRXN3 |
| rs11496066 | FBXL13 | rs2732469 | OR10AD1, H1-7 | rs7219033 | GLP2R |
| rs11657492 | PITPNC1 | rs2732480 | ZNF641 | rs7222481 | GLP2R |
| rs11708067 | ADCY5 | rs275856 | OSBPL1A | rs7249758 | UHRF1 |
| rs11717195 | ADCY5 | rs2793829 | NOTCH2 | rs72846863 | PI16, C6orf89 |
| rs11720108 | ADCY5 | rs28719468 | LINC01416, LINC01415 | rs72964564 | ADCY5 |
| rs11830241 | EP400 | rs3217792 | CCND2-AS1, CCND2 | rs7315028 | Y_RNA, ALG10B |
| rs11870735 | VPS53 | rs34589210 | TMEM87B, MERTK | rs7316626 | CHD4 |
| rs11922794 | SHQ1 | rs34773007 | EXOC6 | rs73167517 | LINC02144, CHD1 |
| rs11940813 | SLIT2 | rs34845373 | DTNB | rs73184014 | LHFPL3 |
| rs11998023 | EBF2 | rs34855922 | TCF7L2 | rs73689877 | LINC01450, SUGCT |
| rs12192275 | RSPO3 | rs34872471 | TCF7L2 | rs74677818 | TMEM87B |
| rs12194820 | RSPO3 | rs34990153 | MSRA | rs75253922 | INSR |
| rs12263348 | REEP3 | rs35742417 | RREB1 | rs7578597 | THADA |
| rs12746673 | PLEKHM2 | rs35895680 | IGF2BP1, GIP | rs7607980 | - |
| rs12977104 | UHRF1 | rs36111056 | FSD2 | rs7619708 | LINC00885, TFRC |
| rs13099581 | EGFEM1P | rs3963364 | PPARG | rs7629630 | EGFEM1P |
| rs1328412 | CHCHD2P9 | rs41463147 | NOTCH2 | rs7645517 | ST6GAL1 |
| rs13292262 | CHCHD2P9 | rs4506565 | TCF7L2 | rs77258096 | CTRB2, CTRB1 |
| rs13405158 | THADA | rs4673712 | IKZF2, MIR4776-1 | rs7786095 | UBE3C |
| rs13414140 | THADA | rs4684848 | PPARG | rs78025551 | TCF7L2 |
| rs139722172 | FAM185A | rs4902833 | TTC9, RN7SL77P | rs7841082 | FAM86B3P, PRAG1 |
| rs1449348 | EGFEM1P | rs4918796 | TCF7L2 | rs7901695 | TCF7L2 |
| rs1493694 | NOTCH2 | rs55812705 | MFSD4B, REV3L | rs7903146 | TCF7L2 |
| rs1513272 | JAZF1 | rs56348580 | HNF1A | rs844215 | FOXP1, EIF4E3 |
| rs1708302 | JAZF1 | rs60384372 | MSRA | rs849135 | JAZF1 |
| rs17175860 | INSR | rs6063048 | EYA2 | rs853866 | FOXP1, EIF4E3 |
| rs17354348 | MIR4776-1, IKZF2 | rs6066138 | EYA2 | rs9814945 | SHQ1 |
| rs17439448 | SUGCT | rs60780116 | ACSL1 | rs9828772 | TMCC1, PLXND1 |
| rs17631783 | MAP3K3, TACO1 | rs60980157 | GPSM1 | rs9900074 | NFE2L1-DT |
| rs17684074 | WDR7 | rs61953351 | OASL |  |  |
| AFR:EAS:SAS:EUR:AMR | | | | | |
| rs11114650 | LIN7A | rs2056857 | LRRC74A | rs6021276 | NFATC2 |
| rs112915006 | MOV10L1, PANX2 | rs2066827 | CDKN1B | rs6026382 | APCDD1L-DT |
| rs1374910 | IGF2BP2 | rs2073721 | TCF19 | rs602652 | CCND1 |
| rs17797882 | - | rs2074120 | CALCR | rs6031563 | HNF4A-AS1, HNF4A |
| rs28599782 | MOB1B | rs2074314 | ABCC8, KCNJ11 | rs60519666 | BEND3 |
| rs3864095 | MASP1 | rs2075423 | PROX1-AS1 | rs60573766 | LINC01141 |
| rs3869115 | HLA-C, HCG27 | rs2080090 | BPTF | rs6059662 | RALY |
| rs60089934 | TRIB1 | rs2088315 | - | rs6065725 | HNF4A, HNF4A-AS1 |
| rs6815464 | MAEA | rs2102278 | DCUN1D4, LRRC66 | rs6070625 | GNAS-AS1 |
| rs6919504 | H3P28, SNORD28B | rs2103132 | AUTS2 | rs6073143 | IFT52 |
| rs72501964 | CTBP1-DT | rs210618 | DCBLD1 | rs61021634 | - |
| rs73541251 | GP2 | rs2107167 | EIF3IP1 | rs6103716 | HNF4A |
| rs7674402 | MOB1B | rs2114824 | GRID1 | rs610930 | AUTS2 |
| rs79407053 | CTBP1-DT | rs2115107 | MAP2K7, LRRC8E | rs6122888 | CEBPB, PELATON |
| rs28792187 | STAU2 | rs2125799 | GUCY1B1 | rs6134031 | LINC01752 |
| rs7046845 | AOPEP | rs2126736 | ANKRD31, HMGCR | rs6137042 | STK35 |
| rs10011174 | RPS3AP18, TMEM154 | rs2134223 | CASR | rs61579137 | EHHADH-AS1 |
| rs10011838 | TMEM154, RPS3AP18 | rs2138157 | NYAP2, MIR5702 | rs616279 | FILNC1 |
| rs1002061 | GTF3AP5 | rs2147479 | DAOA-AS1 | rs61676547 | BPTF |
| rs1002389 | UBE3C | rs2150999 | FOCAD | rs61736066 | SLC39A11 |
| rs10050304 | HSP90AA4P, LINC02508 | rs2153827 | RN7SL644P, EIF2S2P3 | rs61850200 | NEUROG3, TMEM256P1 |
| rs1005752 | LINGO1, HMG20A | rs2169033 | LCORL | rs61975988 | CLEC14A |
| rs10077431 | YTHDC2 | rs217256 | NME5, WNT8A | rs620191 | LBX1-AS1, RNU2-43P |
| rs10087241 | PURG | rs2188848 | STK31 | rs62024526 | IGF1R |
| rs1009358 | LINC02245, LINC02576 | rs2191348 | GTF3AP5 | rs62033400 | FTO |
| rs10096633 | RPL30P9, LPL | rs2191349 | GTF3AP5 | rs62034975 | PDILT |
| rs10097617 | NDUFAF6 | rs2197973 | USP44 | rs62069176 | RTN4RL1 |
| rs10100265 | PINX1 | rs2206734 | CDKAL1 | rs62182438 | SLC25A12, HAT1 |
| rs10114341 | MIRLET7A1HG, PTPDC1 | rs2216063 | IRX3, LINC02140 | rs622217 | SLC22A2, SLC22A3 |
| rs10119430 | EPB41L4B | rs2233632 | SPDEF, IFITM3P3 | rs62310934 | OCIAD1, OCIAD2 |
| rs10137475 | ARID4A | rs2235856 | NFATC2 | rs62319060 | UNC5C |
| rs1016565 | LINC01230, H3P29 | rs2236033 | SFI1 | rs623323 | NXN, MRM3 |
| rs10169613 | MIR4435-2HG | rs2237895 | KCNQ1 | rs62366901 | EMB, PARP8 |
| rs10172939 | SAG | rs2238689 | GIPR | rs62450857 | FOXK1 |
| rs10184004 | GRB14, COBLL1 | rs2240716 | ARVCF | rs62452060 | FOXK1 |
| rs10188334 | TMEM18 | rs2240885 | SLX4 | rs62486442 | LINC00681, LONRF1 |
| rs10190052 | TMEM18 | rs2243102 | GP1BA, SLC25A11 | rs62490267 | ZNF746, ZNF767P |
| rs1019029 | ETV1 | rs2244020 | ZDHHC20P2, FGFR3P1 | rs62492368 | AOC1 |
| rs10193538 | LINC01122 | rs2246012 | ARG1, MED23 | rs62508166 | ANK1 |
| rs10195252 | COBLL1 | rs2249105 | CEP68 | rs62515938 | LINC00968, RPL37P6 |
| rs10201401 | CAMKMT | rs2250301 | WBP1L | rs62530366 | HSF1 |
| rs10228066 | GTF3AP5 | rs2252115 | TSHZ2 | rs633715 | LINC01741, SEC16B |
| rs10228456 | GTF3AP5 | rs2252221 | TSHZ2 | rs6416749 | ZFHX3 |
| rs10228796 | RBM33, GTF3AP5 | rs2255703 | PLXND1 | rs642858 | FILNC1 |
| rs10229583 | PAX4, FSCN3 | rs2257883 | RPSAP52 | rs6432613 | RBMS1 |
| rs10231021 | GTF3AP5, AGMO | rs2258238 | HMGA2 | rs6438234 | EIF4E2P2, ZBTB20 |
| rs10231619 | HECW1 | rs2261181 | RPSAP52 | rs6438247 | EIF4E2P2 |
| rs10240790 | CFAP69 | rs2268078 | RALY | rs6446298 | UBA7, TRAIP |
| rs10258962 | AUTS2 | rs2268382 | CPA1 | rs6453133 | CERT1 |
| rs10260148 | H4P1, KLF14 | rs2269245 | PGM1 | rs6458354 | LINC02537 |
| rs10261386 | CPED1 | rs2269247 | PGM1 | rs6459733 | UBE3C |
| rs10262104 | CALCR | rs2272163 | ROBO2 | rs646123 | MLX |
| rs10268294 | MAD1L1 | rs2276853 | KIF9-AS1, KIF9 | rs6467136 | GCC1, ZNF800 |
| rs10278336 | YKT6 | rs2277339 | PRIM1, HSD17B6 | rs6467314 | KLF14, H4P1 |
| rs10281892 | DGKB | rs2278524 | ANKFY1, CYB5D2 | rs6479591 | AOPEP |
| rs10401969 | SUGP1 | rs2280141 | PLEKHA1 | rs6495168 | COMMD4 |
| rs10404333 | RN7SL836P, GIPR | rs2282456 | TENT5C | rs6495182 | PTPN9 |
| rs10404726 | CRTC1 | rs2283220 | KCNQ1 | rs6496742 | PRC1-AS1, PRC1 |
| rs10406327 | PEPD | rs2284219 | CRHR2 | rs649891 | PTPRD |
| rs10406431 | RN7SL836P, GIPR | rs2287711 | POLK, ANKDD1B | rs649961 | SLC12A8 |
| rs10407429 | GIPR, RN7SL836P | rs2289739 | LTK | rs6515236 | LINC00261 |
| rs10408163 | ZC3H4 | rs2290203 | PRC1-AS1, PRC1 | rs6518681 | HORMAD2, LIF-AS1 |
| rs10408179 | GIPR, RN7SL836P | rs2292626 | PLEKHA1 | rs6537855 | TSPAN2 |
| rs10420309 | RN7SL836P, EML2 | rs2292662 | SCAANT1, ATXN7 | rs6538804 | RMST |
| rs10422861 | PEPD | rs2292893 | LINC01991 | rs6538805 | RMST |
| rs1042725 | HMGA2 | rs2297508 | SREBF1 | rs6545714 | LINC01122 |
| rs10440833 | CDKAL1 | rs2299383 | RELN | rs654629 | RPS6P12 |
| rs10461617 | MAP3K1 | rs2303700 | MAP2K7 | rs6547692 | GCKR |
| rs1046316 | WFS1 | rs2307111 | POC5 | rs6549112 | - |
| rs10469140 | PHLPP1 | rs2313211 | EEF1A1P8, ABCC5 | rs6554060 | DCUN1D4 |
| rs10469860 | LINC01102 | rs231349 | KCNQ1OT1, KCNQ1 | rs6556925 | CAST, PCSK1 |
| rs10471048 | SCD5 | rs231356 | KCNQ1, KCNQ1OT1 | rs6557473 | - |
| rs1048886 | SDHAF4 | rs231361 | KCNQ1, KCNQ1OT1 | rs6558173 | BIN3 |
| rs10490871 | ARPP21 | rs231362 | KCNQ1OT1, KCNQ1 | rs6561273 | ZC3H13, SIAH3 |
| rs10498828 | EYS | rs2327777 | RPL35AP3, Y_RNA | rs6565922 | ZNF236 |
| rs10499183 | MED23 | rs233449 | KCNQ1 | rs6567160 | RPS3AP49, RNU4-17P |
| rs1050226 | SSR1 | rs233450 | KCNQ1 | rs6583826 | KIF11 |
| rs10507349 | RNF6 | rs234853 | KCNQ1 | rs6584116 | FRAT1, FRAT2 |
| rs10510110 | PLEKHA1 | rs234866 | KCNQ1 | rs6597649 | FIBCD1 |
| rs1058018 | UBE2Z | rs2351707 | AP3S2, ARPIN-AP3S2 | rs6598475 | PCSK6 |
| rs1059592 | CCAR2 | rs2358954 | MIR6074, HMGA2 | rs6600191 | FAM234A |
| rs1061810 | HSD17B12 | rs2361478 | MOB1B, RNU6-891P | rs663129 | RNU4-17P, MC4R |
| rs1061813 | ANKH | rs2383208 | CDKN2B-AS1 | rs665268 | MLX |
| rs1063192 | CDKN2B, CDKN2B-AS1 | rs238763 | NUP133 | rs667920 | STAG1 |
| rs1063355 | HLA-DQB1-AS1, HLA-DQB1 | rs2390725 | CERS6 | rs66815886 | ADAMTS9-AS2 |
| rs10734252 | KCNJ11 | rs2394186 | HLA-G, HCP5B | rs6685701 | RPS6KA1 |
| rs10736116 | ARHGAP19, Metazoa_SRP | rs2396083 | LINC02537, VEGFA | rs6691335 | PKN2-AS1 |
| rs10737818 | TBCE | rs2403221 | SBF2 | rs6708643 | ZFP36L2, LINC02580 |
| rs10741243 | TCERG1L | rs2409742 | XKR6, LINC00529 | rs6710938 | RBMS1 |
| rs10745460 | RPS4XP15 | rs2421016 | PLEKHA1 | rs6712905 | DNAH7, STK17B |
| rs10748694 | ARHGAP19, Metazoa_SRP | rs242105 | ACTN1-AS1, RPS29P1 | rs6712932 | GPR45, LINC01918 |
| rs10750397 | ETS1 | rs2426439 | LINC01524 | rs67156297 | ATP8B2, RPSAP17 |
| rs10750840 | TPCN2 | rs243018 | MIR4432HG | rs6715901 | TTN |
| rs10757282 | CDKN2B-AS1 | rs243019 | MIR4432HG | rs6716394 | - |
| rs10757283 | CDKN2B-AS1 | rs243021 | MIR4432HG | rs6723108 | TMEM163, CCNT2-AS1 |
| rs1075855 | ZFHX3 | rs243024 | MIR4432HG | rs67232546 | ETS1-AS1, ETS1 |
| rs10758593 | GLIS3 | rs243088 | MIR4432HG | rs67361341 | TSHZ3-AS1 |
| rs10758950 | PTPRD | rs2431115 | C5orf67 | rs6741676 | SCHLAP1 |
| rs10761745 | JMJD1C | rs2435907 | TCF12 | rs6759355 | DGKD |
| rs10762670 | LRMDA | rs244415 | NFAT5 | rs6762208 | SENP2 |
| rs10766076 | ARNTL | rs244420 | NFAT5 | rs676387 | HSD17B1-AS1, HSD17B1 |
| rs10767659 | BDNF-AS, BDNF | rs2447198 | STRC, CKMT1B | rs6766859 | MRAS, NME9 |
| rs10769936 | TRIM66 | rs2456530 | ONECUT1, RPSAP55 | rs6769511 | IGF2BP2 |
| rs10771367 | KLHL42, RN7SKP15 | rs2465043 | - | rs6777684 | LINC01991, LPP-AS2 |
| rs10771813 | MREGP1 | rs247975 | NLGN1 | rs6780013 | PTPN23 |
| rs10773000 | C12orf65, CDK2AP1 | rs2482506 | WBP1L, RNU6-1231P | rs6780171 | IGF2BP2 |
| rs10787287 | PDCD4 | rs2488597 | STRBP | rs6780569 | RPL24P7, UBE2E2-AS1 |
| rs10787518 | ADRB1, RNU6-709P | rs2495637 | IL13RA1, DOCK11 | rs67839313 | CCDC9B, INAFM2 |
| rs10788575 | MED6P1, PTEN | rs2497306 | Y_RNA, EXOC6 | rs67924081 | FAM89B, EHBP1L1 |
| rs10795945 | CDC123, RN7SL198P | rs2506125 | TLE1 | rs6792892 | RBM6 |
| rs10806906 | LNC-LBCS | rs2510078 | ZNRD2-AS1, LTBP3 | rs6795735 | ADAMTS9-AS2 |
| rs10808671 | NDUFAF6 | rs2513505 | EMSY | rs679992 | RGS7 |
| rs10811662 | CDKN2B-AS1, DMRTA1 | rs253412 | ANKDD1B | rs6806156 | EIF4E2P2 |
| rs10814916 | GLIS3 | rs2540949 | CEP68 | rs6808574 | LPP-AS2, LINC01991 |
| rs10818763 | ZBTB26 | rs2548498 | ITGA1 | rs6813195 | RPS3AP18, TMEM154 |
| rs10823559 | PALD1 | rs2548724 | SLCO4C1 | rs6819331 | TMEM154, RPS3AP18 |
| rs256904 | C5orf67 | rs256903 | C5orf67 | rs6821438 | HMGB3P15 |
| rs10830962 | SNRPGP16, MTNR1B | rs10824307 | LRMDA, ZNF503-AS2 | rs6831006 | TACC3, FGFR3 |
| rs10835690 | MPPED2-AS1 | rs2578011 | PSMD6 | rs6835992 | CDKL2, ODAPH |
| rs10840102 | TRIM66 | rs2581685 | LINC01910, LARP7P3 | rs684214 | NAGLU, HSD17B1P1 |
| rs10841855 | GYS2 | rs2581787 | RFT1 | rs6853272 | RNU6-891P, MOB1B |
| rs10844518 | - | rs2583921 | RPSAP52 | rs6857 | NECTIN2 |
| rs10844519 | - | rs2583934 | HMGA2 | rs686998 | NLGN1 |
| rs10852123 | AP3S2, ARPIN-AP3S2 | rs2583938 | RPSAP52 | rs6878122 | ZBED3-AS1 |
| rs10860209 | RMST | rs2591392 | JMY | rs6879147 | PELO, ITGA1 |
| rs10879261 | TSPAN8 | rs2604566 | C1QTNF7-AS1 | rs6884702 | LINC02224, RN7SL383P |
| rs10882101 | Y_RNA, HHEX | rs2605281 | CWH43 | rs6885132 | ANKH |
| rs10882891 | Metazoa_SRP, RPL12P27 | rs2613503 | RPL31P12 | rs6885157 | MGAT1 |
| rs10886471 | GRK5 | rs2616132 | RPL5P26, LINC02651 | rs6897117 | SLC38A9 |
| rs10886863 | RPL19P16, LINC01153 | rs261967 | CAST, PCSK1 | rs6918311 | RPL35AP3, Y_RNA |
| rs10886864 | RPL19P16, LINC01153 | rs2633310 | CAMK2G | rs6931514 | CDKAL1 |
| rs10889560 | LEPR | rs2633311 | CAMK2G | rs6937795 | RPL35AP3, Y_RNA |
| rs10893829 | LINC02725 | rs2642587 | LINC02651, RPL5P26 | rs6946660 | UBE3C |
| rs10899283 | TSKU | rs2642588 | LINC02651, RPL5P26 | rs6947395 | AUTS2 |
| rs10906115 | RN7SL198P, CDC123 | rs2648731 | MFSD4BP1, ITGA1 | rs6950739 | UBE3C |
| rs10915188 | GMEB1 | rs2658746 | ZNF236 | rs6951280 | DOCK4 |
| rs10916780 | LINC01141 | rs2662390 | RAB3C | rs6956980 | STEAP2-AS1, STEAP2 |
| rs10916784 | LINC01141 | rs2675662 | CAMK2G | rs6960043 | GTF3AP5 |
| rs10937208 | EHHADH-AS1, C3orf70 | rs2679745 | AZIN1 | rs6972291 | SUMO2P3, CICP11 |
| rs10937721 | PPP2R2C, WFS1 | rs2688419 | RPL24P7, SALL4P5 | rs697239 | ZMIZ1 |
| rs10938398 | PRDX4P1, THAP12P9 | rs2706710 | PITPNC1 | rs6975279 | AUTS2 |
| rs10946398 | CDKAL1 | rs2714337 | RREB1 | rs6978327 | AOAH |
| rs10947804 | KCNK17 | rs2723065 | LINC02245, LINC02576 | rs6985028 | TEX15, PURG |
| rs10954772 | PURG | rs2725370 | PURG, TEX15 | rs6986080 | GPAT4, NKX6-3 |
| rs10962 | HNF1B | rs2725371 | PURG | rs7003257 | CPA6 |
| rs10963942 | HAUS6 | rs2730827 | PDZRN4 | rs7018475 | CDKN2B-AS1 |
| rs10965248 | CDKN2B-AS1, DMRTA1 | rs2733289 | PDZRN4 | rs7020996 | DMRTA1, CDKN2B-AS1 |
| rs10974438 | GLIS3 | rs2737226 | TRPS1 | rs7022807 | HAUS6 |
| rs10998304 | TET1 | rs2767036 | PDHX | rs702634 | ARL15 |
| rs10998338 | TET1 | rs2796441 | TLE1, RNU6-1035P | rs7026688 | STRBP |
| rs11001398 | LRMDA | rs2800733 | RSPO3 | rs7029718 | - |
| rs11038672 | CRY2, SLC35C1 | rs2812533 | RPL5P26, LINC02651 | rs7034200 | GLIS3 |
| rs11042596 | H19, IGF2 | rs2816177 | COX5BP8, SOAT1 | rs703972 | ZMIZ1 |
| rs11043003 | MIR4686, ASCL2 | rs2820426 | LYPLAL1-AS1 | rs703977 | ZMIZ1 |
| rs11043007 | ASCL2, MIR4686 | rs2820444 | ZC3H11B, LYPLAL1-AS1 | rs703978 | ZMIZ1 |
| rs11048456 | SSPN, ITPR2 | rs2833610 | HUNK | rs703980 | ZMIZ1 |
| rs11048457 | SSPN, ITPR2 | rs28362333 | HCG27, POU5F1 | rs703981 | ZMIZ1 |
| rs11048458 | ITPR2, SSPN | rs28375915 | PIM3, CRELD2 | rs703983 | ZMIZ1 |
| rs11060464 | TMEM132D | rs2838820 | ADARB1 | rs705145 | LINC02641 |
| rs11063028 | HSPA8P5, CCND2-AS1 | rs28403309 | TNS1 | rs7071943 | CPEB3 |
| rs11070332 | RPAP1, LTK | rs28408270 | HMGB3P15 | rs7072204 | PLEKHA1 |
| rs11073147 | LINC02853 | rs2844623 | HCG27, HLA-C | rs7078559 | CPEB3 |
| rs11073333 | RASGRP1 | rs28488636 | DLGAP1-AS4, DLGAP1 | rs7087591 | Y_RNA |
| rs11078916 | NEUROD2, CDK12 | rs28490139 | TCF12 | rs7099048 | LRMDA |
| rs11095909 | MAGEC3 | rs2851447 | MPHOSPH9 | rs7107217 | RPS27P20, LINC01395 |
| rs11096542 | RDH14 | rs28525376 | LINC01819 | rs7107784 | MIR4686, ASCL2 |
| rs1109754 | LINC00315, LINC00316 | rs2854313 | NF1 | rs7111341 | MIR4686, ASCL2 |
| rs11098676 | NUDT6 | rs2856721 | HLA-DQB1, MTCO3P1 | rs7113297 | SNRPGP16, MTNR1B |
| rs11099942 | DCHS2 | rs2857605 | NFKBIL1 | rs7115753 | MAPK8IP1 |
| rs11107116 | SOCS2, CRADD | rs28637892 | ATXN10, WNT7B | rs7119 | HMG20A |
| rs11113776 | WSCD2 | rs28637955 | ZBTB46, ZBTB46-AS1 | rs712315 | SRP54-AS1 |
| rs11117364 | BANP, LINC02182 | rs28642213 | GPSM1 | rs7123361 | EMSY |
| rs1111875 | HHEX, Y_RNA | rs2867125 | LINC01875, TMEM18 | rs7124681 | CELF1 |
| rs11123406 | MIR4435-2HG | rs2867570 | TSHZ3-AS1 | rs7126460 | LINC02687, CHST1 |
| rs111246699 | KSR2 | rs28690107 | MTND3P22 | rs7127212 | HMBS, VPS11 |
| rs1112718 | Y_RNA, EXOC6 | rs28691713 | PIM3 | rs7130522 | MAML2 |
| rs111283203 | AMFR, NUDT21 | rs286925 | EHF | rs71320321 | IGF2BP2 |
| rs11129735 | TRANK1 | rs28712435 | TM4SF4, WWTR1 | rs7132351 | KSR2, RFC5 |
| rs11131794 | RN7SKP188 | rs2872246 | ABCC5, EEF1A1P8 | rs7132908 | FAIM2 |
| rs11137820 | MTND2P8 | rs28758542 | GACAT3 | rs7134150 | PDE3A |
| rs11155073 | LINC01625, ATP5PBP6 | rs2876354 | RPL35AP3 | rs7138300 | TSPAN8 |
| rs11159086 | ISCA2 | rs2876826 | DDC, FIGNL1 | rs7147483 | CLEC14A |
| rs11159347 | OR7K1P, LINC02306 | rs2877716 | ADCY5 | rs7161785 | C2CD4B, NPM1P47 |
| rs1116357 | EIF2S2P7 | rs28790585 | LINC02915, THBS1 | rs716316 | MACROD2-AS1, MACROD2 |
| rs111640200 | CFAP44 | rs28819812 | PDGFC, LINC02272 | rs7163629 | AP3S2, ARPIN-AP3S2 |
| rs11165354 | TGFBR3 | rs2908274 | GCK | rs7163757 | NPM1P47, C2CD4B |
| rs111669836 | KCNK7, MAP3K11 | rs2908279 | RNA5SP230, MYL7 | rs7166281 | IMP3, SNUPN |
| rs11172254 | KIF5A | rs2908282 | YKT6 | rs7167878 | NPM1P47, C2CD4B |
| rs11173646 | - | rs2908286 | GCK | rs7169799 | FAM227B |
| rs11199116 | SEC23IP | rs2908334 | AGMO | rs7172432 | C2CD4B, NPM1P47 |
| rs11201992 | GRID1 | rs2913873 | RANBP17 | rs7174878 | ARPIN-AP3S2, AP3S2 |
| rs11201999 | GRID1 | rs2925979 | CMIP | rs7177055 | LINGO1, HMG20A |
| rs11202589 | KLLN, CFL1P1 | rs2933211 | MDGA2 | rs7178572 | HMG20A |
| rs11202627 | MED6P1, PTEN | rs2943640 | NYAP2, MIR5702 | rs7178762 | USP3, USP3-AS1 |
| rs11205766 | FAF1 | rs2943641 | - | rs718314 | ITPR2, SSPN |
| rs1122518 | ETV1 | rs2943650 | MIR5702, NYAP2 | rs7185735 | FTO |
| rs11236524 | RN7SL786P, DGAT2 | rs2943656 | - | rs7188071 | RABEP2 |
| rs11237460 | GAB2 | rs2943657 | - | rs7202877 | CTRB2, CTRB1 |
| rs11240074 | BCL9, LINC00624 | rs2946504 | TRMT9B | rs7213402 | MYH10, CCDC42 |
| rs11240351 | CNTN2 | rs2950835 | GCKR, C2orf16 | rs7220340 | TWF1P1, CRYBA1 |
| rs112498319 | RREB1, RN7SL554P | rs2952858 | LINC02465 | rs7224685 | ZZEF1 |
| rs11257600 | CDC123 | rs2956092 | APIP | rs7224711 | CYTH1 |
| rs11257655 | RN7SL198P, CDC123 | rs29582 | LINC02152 | rs7227272 | MIR924HG |
| rs11257657 | CDC123, RN7SL198P | rs2972143 | MIR5702, NYAP2 | rs723355 | MDGA2 |
| rs112674299 | ATXN7 | rs2972144 | NYAP2, MIR5702 | rs7240767 | LAMA1 |
| rs1127215 | PTGFRN | rs2972145 | - | rs7246440 | ZNF799 |
| rs1127655 | PTGFRN | rs2980766 | PRAG1, RN7SL178P | rs72501962 | CTBP1-DT |
| rs1128249 | COBLL1 | rs2982521 | LINC01625, ATP5PBP6 | rs7250869 | PEPD |
| rs11514706 | AGMO, GTF3AP5 | rs3012060 | LRMDA | rs7261425 | CFAP61 |
| rs1153188 | VDAC1P5, DCD | rs3020781 | PKLR | rs72640313 | MIR17HG, PPIAP23 |
| rs11558471 | SLC30A8 | rs302395 | LINC01933, GLRA1 | rs7274134 | - |
| rs11583755 | KLHL21 | rs305686 | KCNH7 | rs7274168 | CHMP4B |
| rs11591741 | CHUK | rs3094682 | LINC02571 | rs727734 | JARID2 |
| rs11608419 | PCED1B | rs3111316 | FARSA | rs72892910 | RPS17P5, TFAP2B |
| rs11614914 | FBRSL1 | rs3115960 | CHST6 | rs72906810 | MIR4421, CDKN2C |
| rs11616380 | SPRY2, LINC01080 | rs3122231 | ZNF239, ZNF487 | rs72982988 | RPS3AP49, RNU6-567P |
| rs11634397 | FAH, ZFAND6 | rs312457 | SLC16A13 | rs730497 | GCK |
| rs11635657 | TRPM1 | rs3130283 | AGPAT1, PPT2-EGFL8 | rs7305618 | HNF1A-AS1 |
| rs11642015 | FTO | rs3130501 | POU5F1 | rs730570 | DLK1, LINC00523 |
| rs11642430 | TLCD3B | rs3130931 | POU5F1, TCF19 | rs73069940 | CTBP1 |
| rs11642841 | FTO | rs3132524 | POU5F1 | rs7307263 | LINC01234 |
| rs11646052 | GINS2 | rs3135911 | ZNF346, FGFR4 | rs730831 | CTBP1 |
| rs11656775 | RAI1 | rs3217860 | CCND2 | rs73085586 | LINC00261 |
| rs11659939 | NOL4, ASXL3 | rs3176466 | CDKN2C | rs73121277 | DDC, FIGNL1 |
| rs11662800 | LDLRAD4 | rs319598 | PCBD2, TXNDC15 | rs73230612 | ZBTB20 |
| rs11666603 | RPS29P23, ZNF799 | rs314879 | DDX39AP1, SNORD36 | rs7330796 | TBC1D4 |
| rs11667244 | ZC3H4 | rs328 | LPL | rs73347525 | MEG3 |
| rs11671664 | GIPR | rs328301 | FGFR1, C8orf86 | rs73541184 | ARAP1 |
| rs11677370 | DCDC2C | rs328506 | HMGB1P1, RBM38 | rs73642097 | NFIB |
| rs11680058 | GACAT3, CYRIA | rs329118 | JADE2 | rs73708054 | EFR3A |
| rs11688682 | Y_RNA, LINC01101 | rs329122 | JADE2 | rs738408 | PNPLA3 |
| rs11688931 | LINC01101, Y_RNA | rs335810 | FOXB1 | rs738409 | PNPLA3 |
| rs1169288 | HNF1A-AS1, HNF1A | rs3383 | HIVEP2 | rs7403531 | RASGRP1 |
| rs1169299 | HNF1A | rs33932777 | CDC123, RN7SL198P | rs7412314 | FMO4 |
| rs11699802 | CEBPB, PELATON | rs340841 | PROX1-AS1 | rs742762 | GLP1R |
| rs11717959 | IGF2BP2 | rs340874 | PROX1, PROX1-AS1 | rs74322397 | ATP5MGP7, TUBG1 |
| rs11723275 | SHROOM3 | rs34143602 | HERC1 | rs7432739 | PXK |
| rs11759026 | MIR588, CENPW | rs34247110 | KCNK16, KCNK17 | rs746673 | CCDC9B, PHGR1 |
| rs11774915 | RNU6-526P, RNU6-1151P | rs34298980 | LRFN2 | rs7482891 | ASCL2, MIR4686 |
| rs11786992 | ESRP1 | rs343092 | HMGA2-AS1, HMGA2 | rs7483027 | OR5B1P, OR5B17 |
| rs11787792 | GPSM1 | rs34341 | ANKDD1B | rs7502556 | NF1 |
| rs11793035 | LINC01451 | rs345367 | ARHGAP24 | rs7507912 | GIPR, RN7SL836P |
| rs11793831 | - | rs34573045 | TM4SF4 | rs7531962 | CHD1L, LINC00624 |
| rs11802899 | LDLRAD2, USP48 | rs34584161 | RNF6 | rs753270 | ZMIZ1 |
| rs11819995 | ETS1 | rs34617913 | KCTD8 | rs7534008 | DEPDC1-AS1 |
| rs1182389 | UBE3C | rs34620785 | HSPE1P14, RNU6ATAC8P | rs7538321 | SLC41A1, PM20D1 |
| rs1182394 | UBE3C | rs346240 | LINC00907 | rs7542900 | SLC44A3-AS1, F3 |
| rs1182395 | UBE3C | rs34669198 | SRBD1 | rs7546252 | DNM3, PIGC |
| rs1182436 | UBE3C | rs34744311 | HHEX, Y_RNA | rs7554251 | MTOR |
| rs1182444 | UBE3C | rs34759301 | USP48, LDLRAD2 | rs7558413 | KCNS3, RDH14 |
| rs11830243 | EP400 | rs348330 | ABCB10 | rs7558502 | EPC2 |
| rs11858759 | ALDH1A2 | rs34885433 | SLCO4A1 | rs7559658 | - |
| rs11913442 | L3MBTL2, EP300-AS1 | rs34907385 | LRMDA | rs756145 | LINC01230, H3P29 |
| rs11925227 | TNIK, SLC2A2 | rs349359 | KCNB2 | rs7561798 | SPHKAP |
| rs11926494 | UBE2E2 | rs34965774 | RFC5, KSR2 | rs75619936 | RFX3 |
| rs11926707 | PTH1R | rs35072907 | FAF1 | rs7564708 | LINC01875, TMEM18 |
| rs11927381 | IGF2BP2 | rs35251247 | ALKBH3, HSD17B12 | rs757110 | ABCC8 |
| rs11929640 | SIDT1 | rs35261542 | CDKAL1 | rs7572970 | RBMS1 |
| rs1194592 | ATP8B2, RPSAP17 | rs35352848 | UBE2E2 | rs75756987 | PCNT |
| rs1194606 | AQP10 | rs35452727 | CAMK2B | rs7578326 | - |
| rs11960799 | MARCHF3 | rs35497231 | ATP5MGP5 | rs7585737 | CAPN14 |
| rs11967262 | VEGFA, LINC02537 | rs35612982 | CDKAL1 | rs7589501 | MIR4432HG |
| rs11980500 | DGKB | rs35654957 | FGFRL1 | rs7593730 | RBMS1 |
| rs11994255 | ELP3, RPL5P22 | rs35678078 | PPP2R2C | rs7609422 | - |
| rs12001437 | RN7SKP114, DCAF12 | rs35859536 | MED30, SLC30A8 | rs7612463 | UBE2E2 |
| rs12010175 | CCNQ | rs358806 | LINC02030 | rs7626079 | SLC25A26 |
| rs12031188 | FAF1 | rs35901985 | SORBS2 | rs7629 | TRIM59 |
| rs12041243 | MDM4 | rs35906730 | EIF2S2P3, HHEX | rs7630877 | PEX5L |
| rs12048743 | DSTYK | rs35913461 | TMEM18 | rs7633673 | MBNL1 |
| rs12052648 | RNA5SP87, RN7SL117P | rs35999103 | - | rs7633675 | IGF2BP2 |
| rs12056338 | LINC00681, LONRF1 | rs36027443 | RASGRP1 | rs7640539 | IGF2BP2 |
| rs12088739 | CDKN2C, MIR4421 | rs366577 | ENO3 | rs7644981 | CASR |
| rs12109081 | - | rs367943 | MCC | rs7645613 | EIF4E2P2 |
| rs12116935 | SH3D21, EVA1B | rs372558 | UBE2O | rs7646242 | TOMM22P6 |
| rs12151653 | TEX41 | rs3735491 | CCM2, NACAD | rs7651090 | IGF2BP2 |
| rs1215468 | SPRY2, LINC01080 | rs3743140 | INAFM2 | rs765142292 | ZCCHC12, IL13RA1 |
| rs12187734 | - | rs3744347 | CBX1 | rs7653569 | EIF4E2P2, ZBTB20 |
| rs12219514 | Y_RNA, HHEX | rs3747207 | PNPLA3 | rs76550717 | ARAP1 |
| rs12231031 | - | rs3751837 | CLUAP1 | rs7656001 | CCSER1 |
| rs1225052 | CPNE4 | rs3753693 | GMEB1, YTHDF2 | rs7656416 | CTBP1-DT |
| rs12299509 | CCND2 | rs3755879 | UNC5C | rs7659468 | SLC9B1 |
| rs12304921 | HIGD1C | rs3755934 | CTBP1 | rs7659604 | SMIM43, ANXA5 |
| rs12305809 | ZNF268 | rs376993806 | GPSM1 | rs7660000 | FAM13A |
| rs123378 | DLEU1 | rs3772071 | RBMS1 | rs7660590 | FBXW7 |
| rs1236816 | PTEN | rs377457 | C16orf74 | rs7664347 | SLIT2 |
| rs12380322 | HAUS6 | rs3774723 | ATXN7 | rs7669833 | TMEM154, RPS3AP18 |
| rs12403994 | PROX1-AS1 | rs3779272 | MAGI2 | rs76704029 | HERC2 |
| rs12419690 | CRY2, SLC35C1 | rs3786897 | PEPD | rs7674212 | SLC9B2 |
| rs12422600 | HOXC4 | rs379417 | BRD3OS | rs7685296 | FBXW7 |
| rs12433335 | PRKD1 | rs3794205 | CAMKK2 | rs7695096 | SLC9B1 |
| rs12453376 | ZZEF1 | rs3794991 | GATAD2A | rs7719891 | RASA1 |
| rs12453394 | UBE2Z | rs3798519 | TFAP2B | rs7721099 | LINC00461 |
| rs12454712 | BCL2 | rs3802177 | SLC30A8 | rs77300780 | GIPR, RN7SL836P |
| rs12463719 | MTCO1P17, BMPR2 | rs3802219 | TRPS1 | rs7732130 | ZBED3-AS1 |
| rs12497133 | SLC12A8 | rs3810291 | ZC3H4 | rs7739842 | ENPP3 |
| rs12507026 | - | rs3811978 | ITGA1 | rs7742292 | NHSL1 |
| rs12509379 | LARP1B, PGRMC2 | rs3816157 | PSMD6, PSMD6-AS1 | rs77424687 | HEATR5B |
| rs12518099 | LINC01339 | rs3816605 | NUP160 | rs7752666 | BEND3, RNU6-1299P |
| rs12519500 | DMGDH | rs38221 | - | rs7754840 | CDKAL1 |
| rs12539264 | LINC02838 | rs3826482 | ATP2A3 | rs7756992 | CDKAL1 |
| rs12546365 | NDUFAF6 | rs3828242 | ERBB4 | rs7758002 | RGS17 |
| rs12549902 | ANK1, NKX6-3 | rs3843467 | C5orf67 | rs7758115 | LINC02523, HEY2 |
| rs12550613 | ANK1, NKX6-3 | rs3845843 | NMI, TNFAIP6 | rs7765207 | NHSL1 |
| rs12569857 | CAMK1D | rs3872707 | SETD5 | rs7766070 | CDKAL1 |
| rs12571751 | ZMIZ1 | rs3887059 | RPL37P6, LINC00968 | rs7781440 | GRB10 |
| rs391300 | SRR | rs3887925 | ST6GAL1 | rs7787720 | ETV1, RBMX2P4 |
| rs12578639 | WSB2 | rs3897727 | ANO4 | rs7795991 | ETV1 |
| rs12586772 | SYNDIG1L, NPC2 | rs12578595 | RN7SKP15, PTHLH | rs77989445 | RNU11-5P, PRKD1 |
| rs12600132 | PKD1L3 | rs39204 | STEAP2-AS1 | rs780093 | GCKR |
| rs12602834 | NF1, EVI2B | rs39218 | STEAP2-AS1 | rs780094 | GCKR |
| rs1260326 | GCKR | rs39223 | STEAP2-AS1 | rs781831 | ZZEF1 |
| rs12603589 | BPTF | rs3923113 | COBLL1, GRB14 | rs781852 | ZZEF1 |
| rs12617659 | Y_RNA, LINC01101 | rs392794 | C5orf67 | rs78403475 | GPSM1 |
| rs12625671 | HNF4A | rs39328 | RELN | rs7845219 | NDUFAF6 |
| rs12630883 | UBE2E2 | rs3934712 | LINC02227 | rs7847880 | FOCAD |
| rs12633613 | UBE2E2 | rs3996350 | H4P1, KLF14 | rs7858727 | EPB41L4B |
| rs12640250 | DCAF16, FAM184B | rs4077129 | PIM3 | rs7867635 | - |
| rs12642790 | SCD5 | rs410150 | LINC00907 | rs7895872 | BBIP1 |
| rs12649012 | MOB1B, RNU6-891P | rs41276588 | VPS25P1, TENT5C | rs7900112 | LRMDA |
| rs12662968 | RPL35AP3, Y_RNA | rs4129858 | LAMC1 | rs79090772 | FAF1 |
| rs12667919 | CAPZA1P4 | rs41304257 | IPO9 | rs7912336 | HSPA12A |
| rs12680028 | TRHR | rs4132228 | ADAMTS9-AS2 | rs791595 | LEP, MIR129-1 |
| rs12680217 | LINC01605 | rs4148646 | ABCC8 | rs7918400 | TCF7L2 |
| rs12681990 | SMARCE1P4, KCNU1 | rs4148856 | ABCB9 | rs7922045 | LINC01153 |
| rs12692738 | COBLL1 | rs419842 | MYBL2 | rs7929543 | - |
| rs12698877 | AUTS2 | rs4234733 | WFS1, PPP2R2C | rs79310463 | KSR2 |
| rs12698897 | AUTS2 | rs4237150 | GLIS3 | rs7931302 | ETS1 |
| rs12712928 | KRTCAP2P1, SIX3 | rs4238013 | CCND2-AS1 | rs7941510 | TRIM66 |
| rs12714314 | MYT1L | rs4258054 | GLIS3 | rs7943101 | WT1-AS |
| rs12719778 | Metazoa_SRP, ARHGAP39 | rs4267949 | CASC15, RNU6-1060P | rs79548680 | RCCD1 |
| rs12741141 | KLHL21 | rs4275659 | ABCB9 | rs7955901 | TSPAN8 |
| rs12743974 | IL23R | rs4279506 | IGF2BP3, RPS2P32 | rs7961581 | TSPAN8 |
| rs12779790 | RN7SL198P, CDC123 | rs4281707 | RBL2 | rs7968902 | HMGA2, MIR6074 |
| rs12780155 | CDC123 | rs4287436 | LINC00370 | rs7970687 | ZNF10 |
| rs12782078 | CPN1 | rs429358 | APOE | rs7978610 | RFLNA, ZNF664 |
| rs12789028 | LTBP3 | rs4294149 | MIR30B, ZFAT | rs7978895 | LINC02461 |
| rs12802972 | FAM99B | rs4325 | ACE | rs797973 | RPL35AP35, NUFIP2 |
| rs12811407 | FBRSL1 | rs4335 | ACE | rs7983505 | TOMM22P3, KL |
| rs12820906 | PITPNM2 | rs4343858 | NSD1 | rs7985179 | MIR17HG, PPIAP23 |
| rs12823740 | ZNF664, RFLNA | rs4358140 | DNAH7 | rs798549 | AMZ1 |
| rs12825669 | - | rs4382480 | MFHAS1 | rs7987740 | LINC00370 |
| rs12849637 | IL13RA1, DOCK11 | rs4384608 | KLHDC4, SLC7A5 | rs7991679 | LINC02338 |
| rs12854077 | MTCL1P1, RNU6-54P | rs4391603 | DUSP9, KRT18P48 | rs799661 | SRP54-AS1 |
| rs12856169 | HMGB1, UBE2L5 | rs4402960 | IGF2BP2 | rs8005994 | - |
| rs12883788 | AKAP6 | rs4414887 | IGF2BP2 | rs8008540 | NPC2 |
| rs12890750 | MARK3 | rs4420638 | APOC1, APOC1P1 | rs8010382 | PPP4R3A |
| rs12892257 | PSMA3-AS1, ARMH4, PSMA3 | rs4440243 | ART3 | rs8017808 | KRT8P1, CLEC14A |
| rs12899811 | VPS33B | rs445084 | SLC22A18AS | rs8018512 | CLEC14A |
| rs12907887 | INAFM2, PLCB2 | rs4454866 | - | rs8026714 | PRC1-AS1, PRC1 |
| rs12910361 | LINGO1, HMG20A | rs4457053 | ZBED3-AS1 | rs8031576 | ARPIN-AP3S2, AP3S2 |
| rs12910825 | PRC1-AS1, PRC1 | rs4458523 | WFS1 | rs8032416 | NPM1P47, C2CD4B |
| rs12912009 | AP3S2, ARPIN-AP3S2 | rs4463416 | - | rs8032939 | RASGRP1 |
| rs12917449 | PML | rs4465929 | BTD, ANKRD28 | rs8033580 | BBS4 |
| rs12918782 | LMF1 | rs4472028 | MBNL1 | rs8033609 | RORA |
| rs12920022 | SPG7 | rs4479849 | LINC02224 | rs8035957 | RASGRP1 |
| rs12943365 | NF1 | rs4482463 | KCNS3, PARD3B | rs8037894 | C2CD4B, NPM1P47 |
| rs1296328 | RNU1-89P, TERF1P3 | rs448918 | BRD3OS, VAV2 | rs8038760 | SIN3A |
| rs12967878 | RPS3AP49, RNU4-17P | rs4502156 | C2CD4B, NPM1P47 | rs8042680 | PRC1, PRC1-AS1 |
| rs12970134 | RNU4-17P | rs4517619 | SOX5 | rs8043085 | RASGRP1 |
| rs12986742 | LINC01122 | rs4527850 | CCN4, TG | rs8046545 | ATP2A1 |
| rs12992995 | SP9, LINC01305 | rs4532315 | CSPG4BP | rs8050136 | FTO |
| rs13020443 | NMI, TNFAIP6 | rs459193 | C5orf67, RPL26P19 | rs8054556 | TMEM219 |
| rs13039863 | HNF4A | rs4607103 | ADAMTS9-AS2 | rs8056890 | ATP2A1 |
| rs13040225 | CEBPB, PELATON | rs4607517 | GCK | rs8061528 | SLX4 |
| rs13041756 | GSTM3P1, RN7SKP140 | rs4611812 | ADAMTS9-AS2 | rs806215 | FSCN3 |
| rs13043901 | ZNF217, Y_RNA | rs463924 | KCNQ1, KCNQ1OT1 | rs8067056 | MAPT |
| rs13065698 | MBNL1 | rs464605 | RPL26P19, C5orf67 | rs8068804 | ZZEF1 |
| rs13073970 | KLF7P1, EIF5A2 | rs465002 | C5orf67 | rs8071043 | ZZEF1 |
| rs13086331 | LINC01991 | rs4655617 | SGIP1 | rs8089364 | RNU4-17P |
| rs13092876 | IGF2BP2 | rs4658234 | HSP90B3P, WDR82P2 | rs8090011 | LAMA1 |
| rs13094957 | UBE2E2 | rs4671799 | LINC01829 | rs8100204 | SUGP1 |
| rs13129612 | MOB1B, RNU6-891P | rs4678059 | CASR | rs8101064 | INSR |
| rs13155752 | RN7SL383P, LINC02224 | rs4678408 | MRAS, NME9 | rs8101149 | ZC3H4 |
| rs13162708 | LINC00461 | rs4686392 | - | rs8107527 | GIPR, RN7SL836P |
| rs13163173 | LINC00461 | rs4686471 | LINC01991 | rs8107974 | SUGP1 |
| rs1317617 | ZMIZ1, PPIF | rs4688760 | RBM6 | rs8108269 | RN7SL836P, GIPR |
| rs1320164 | TP53INP1, NDUFAF6 | rs4689388 | WFS1, LINC02495 | rs8188241 | - |
| rs13234269 | KLF14, H4P1 | rs4703911 | RPL5P16 | rs8192675 | SLC2A2 |
| rs13237518 | TMEM106B | rs4711750 | LINC02537, VEGFA | rs824248 | MIR876 |
| rs13266634 | SLC30A8 | rs4712523 | CDKAL1 | rs825452 | RFLNA |
| rs1327123 | TSEN15, COLGALT2 | rs4712524 | CDKAL1 | rs825476 | RFLNA |
| rs1327315 | - | rs4714422 | OARD1 | rs827237 | LINC02622, PCBD1 |
| rs1327796 | PALM2AKAP2 | rs4715207 | TFAP2B | rs831571 | PSMD6, PRICKLE2-AS1 |
| rs13288108 | - | rs4721089 | MAD1L1 | rs838720 | DGKD |
| rs13292136 | CHCHD2P9 | rs472265 | ACP7 | rs838735 | DGKD |
| rs133015 | PLA2G6 | rs4731420 | MIR129-1, LEP | rs849133 | JAZF1 |
| rs13330163 | IL34 | rs4731701 | KLF14, H4P1 | rs849134 | JAZF1 |
| rs1333051 | DMRTA1, CDKN2B-AS1 | rs4734193 | TRHR | rs853974 | RPS4XP9, RSPO3 |
| rs1333052 | CDKN2B-AS1 | rs4736819 | NKX6-3, ANK1 | rs858519 | SHBG |
| rs13365225 | KCNU1, SMARCE1P4 | rs4736999 | NKX6-3 | rs860262 | JAZF1 |
| rs13389219 | COBLL1 | rs4739515 | LINC01605 | rs862016 | NELFCD, GNAS |
| rs13406280 | GALNT3 | rs474513 | SLC22A3 | rs862320 | NFAT5 |
| rs13415288 | SPHKAP | rs4747971 | CDC123, RN7SL198P | rs864745 | JAZF1 |
| rs13417036 | LINC01122 | rs475002 | SNAPC2 | rs867489 | PELATON, CEBPB |
| rs13428870 | LINC01122 | rs4760790 | TSPAN8 | rs878017 | BEND7 |
| rs13434089 | ATXN7 | rs476828 | - | rs878521 | CAMK2B, YKT6 |
| rs1359790 | SPRY2, LINC01080 | rs4776970 | MAP2K5 | rs884847 | NOS1 |
| rs136867 | DEPDC5 | rs4777857 | SEPHS1P2 | rs887400 | MIR633, PRELID3BP3 |
| rs13737 | IMP3 | rs4782568 | MLYCD, OSGIN1 | rs890940 | EBF1 |
| rs1377807 | ZZEF1 | rs4788815 | MARVELD3, TAT-AS1 | rs893617 | ARPIN-AP3S2, AP3S2 |
| rs1378254 | DLC1 | rs4804181 | ZNF799 | rs896852 | NDUFAF6, TP53INP1 |
| rs137848 | IL17REL | rs4804833 | MAP2K7 | rs896854 | NDUFAF6, TP53INP1 |
| rs1379871 | DMD | rs4805681 | TSHZ3 | rs896911 | - |
| rs1387153 | MTNR1B, SNRPGP16 | rs4805881 | PEPD | rs904372 | OCA2 |
| rs138771 | TOM1 | rs4807125 | TCF3 | rs911300 | PIEZO1P2, MIR296 |
| rs1412234 | LINGO2 | rs480840 | XRRA1 | rs917195 | CRHR2 |
| rs1421085 | FTO | rs4809369 | C20orf181, ZBTB46 | rs924753 | BTD |
| rs1430780 | LINC02831 | rs4809627 | EYA2 | rs9257408 | LINC01556, KRT18P1 |
| rs1431819 | COL27A1 | rs4809906 | LINC01524 | rs9271774 | HLA-DRB1, HLA-DQA1 |
| rs1436953 | NPM1P47, C2CD4B | rs4810145 | GNAS-AS1 | rs9273368 | HLA-DQA1, HLA-DQB1 |
| rs1436955 | NPM1P47, C2CD4B | rs4810426 | HNF4A-AS1, HNF4A | rs9295474 | CDKAL1 |
| rs1437055 | VGLL3 | rs4812829 | HNF4A | rs9296095 | BAK1, GGNBP1 |
| rs1451506 | SNRPGP17, YPEL2 | rs4820323 | PLA2G6, MAFF | rs9300039 | LINC02745, LINC01499 |
| rs1468308 | OR7A11P, OR7A3P | rs4823182 | SAMM50 | rs9308614 | Y_RNA |
| rs1470560 | ARPP21, RNU6-243P | rs4832290 | KDM3A | rs9309245 | - |
| rs1470579 | IGF2BP2 | rs4845987 | MTOR | rs9316500 | DLEU1, DLEU7 |
| rs1475655 | PPIAP23, MIR17HG | rs484943 | BMF | rs9316706 | LINC00540 |
| rs1480474 | HMGA2 | rs4854343 | TMEM18, LINC01875 | rs9319382 | POLR1D |
| rs148462165 | UBE2E2 | rs4865473 | TACC3, FGFR3 | rs9319943 | GRP, SEC11C |
| rs1495377 | TSPAN8 | rs4865796 | ARL15 | rs9348440 | CDKAL1 |
| rs1496653 | UBE2E2 | rs488166 | TOMM22P3, KL | rs9348441 | CDKAL1 |
| rs150111048 | IGF2BP2 | rs488321 | TOMM22P3, KL | rs9350271 | CDKAL1 |
| rs1505210 | OR51C1P, OR51E2 | rs4897182 | MIR588, CENPW | rs9368112 | LNC-LBCS |
| rs1515104 | MIR5702, NYAP2 | rs4899280 | DCAF5 | rs9368222 | CDKAL1 |
| rs1516728 | DGKG | rs4901812 | CLEC14A, KRT8P1 | rs9369425 | LINC02537, VEGFA |
| rs1517037 | SEC11C, GRP | rs4902002 | MNAT1 | rs9376382 | ECT2L |
| rs1528287 | PTPRQ | rs4906272 | TRAF3 | rs9383649 | RGS17 |
| rs1531343 | RPSAP52 | rs4916253 | PIGC, DNM3 | rs9384 | GCDH, SYCE2 |
| rs1535500 | KCNK16 | rs4923543 | METTL15 | rs9384193 | OPRM1, IPCEF1 |
| rs1561927 | LINC00824 | rs4925109 | RAI1 | rs9390022 | HIVEP2 |
| rs1562396 | H4P1, KLF14 | rs4929965 | MIR4686, ASCL2 | rs940904 | PITPNM2 |
| rs1562398 | H4P1, KLF14 | rs4930045 | ASCL2, MIR4686 | rs9410573 | RNU6-1035P, TLE1 |
| rs1563575 | RBMS1 | rs4930091 | CD81-AS1 | rs9411425 | MED27 |
| rs944801 | CDKN2B-AS1 | rs4930726 | CCDC92 | rs9438610 | PDIK1L, TRIM63 |
| rs1570247 | GBA2 | rs4932143 | AP3S2, ANPEP | rs1567353 | H3P29, LINC01230 |
| rs1573090 | RPL35AP3, NHEG1 | rs4932265 | AP3S2, ARPIN-AP3S2 | rs9449295 | EEF1B2P5 |
| rs1573219 | NTRK2 | rs4942883 | KPNA3, RNY4P30 | rs945187 | FRAT1, FRAT2 |
| rs1574285 | GLIS3 | rs4946812 | BEND3 | rs9460550 | CDKAL1 |
| rs1580278 | CENPE, LINC02428 | rs4964665 | WSCD2 | rs9465871 | CDKAL1 |
| rs1625526 | MIR466 | rs4975241 | LINC02615 | rs9470794 | ZFAND3 |
| rs1631619 | MTND5P17 | rs4976033 | VWA8P1, PIK3R1 | rs9472138 | LINC02537, VEGFA |
| rs163177 | KCNQ1 | rs4977213 | BOP1 | rs9472139 | LINC02537 |
| rs163182 | KCNQ1 | rs4984980 | LMF1 | rs9479 | PML |
| rs163184 |  | rs5010712 | EML6 | rs9494624 | RPL35AP3, NHEG1 |
| rs1635852 | JAZF1 | rs501470 | SLC22A3 | rs9502478 | LY86, LY86-AS1 |
| rs1641523 | ATP1B2, SHBG | rs5015480 | HHEX, Y_RNA | rs9502570 | SSR1, RREB1 |
| rs1650505 | EBF1 | rs5017300 | LINC01875, TMEM18 | rs9505097 | RREB1, SSR1 |
| rs1656794 | SEPTIN9 | rs5017301 | TMEM18, LINC01875 | rs9505118 | SSR1 |
| rs1662185 | CCDC88B, PRDX5 | rs506597 | POP7, EPO | rs9515905 | MIR17HG, PPIAP23 |
| rs1670754 | LINC02506, LINC02353 | rs508419 | ANK1 | rs952471 | HMG20A |
| rs1694068 | ARL15 | rs510062 | GDF6, GAPDHP30 | rs952472 | HMG20A |
| rs16988991 | HNF4A | rs510807 | GLIS3 | rs9537803 | PCDH17 |
| rs17035289 | TET2 | rs513320 | TOMM22P3, KL | rs9555581 | LINC00370 |
| rs1705263 | TSPAN8 | rs515071 | ANK1 | rs9560114 | LINC02337 |
| rs1707498 | PTPRR, TSPAN8 | rs516946 | ANK1 | rs9562987 | DLEU7, DLEU1 |
| rs17086692 | - | rs5213 | KCNJ11 | rs9563574 | LINC02338, RNA5SP30 |
| rs17091891 | RPL30P9, LPL | rs5215 | KCNJ11 | rs9563615 | RNY4P29 |
| rs17106184 | FAF1 | rs5219 | KCNJ11 | rs9564268 | - |
| rs17168486 | DGKB | rs523288 | RNU4-17P, MC4R | rs9568861 | ZNF646P1, LINC00558 |
| rs1724557 | - | rs524903 | HPSE2 | rs9581587 | RNF6 |
| rs17261179 | MFSD4BP1 | rs529623 | FXYD6-FXYD2, FXYD2 | rs9581591 | RNF6 |
| rs1727313 | MPHOSPH9 | rs531676 | CRTAC1 | rs9583907 | PPIAP23, MIR17HG |
| rs17359493 | INTS8 | rs532504 | SEC16B | rs9587811 | LINC00370 |
| rs173964 | C5orf67 | rs534043 | EPO, POP7 | rs963740 | DLEU7, DLEU1 |
| rs17411031 | RPL30P9, LPL | rs538801 | SUPT3H | rs9648716 | BRAF |
| rs17447640 | ATP8A1 | rs539515 | LINC01741, SEC16B | rs965480 | LINGO1, HMG20A |
| rs1752169 | DENND1A | rs543159 | SLC22A3 | rs96844 | SETD9, MAP3K1 |
| rs17522122 | AKAP6 | rs545608 | CRYZL2P-SEC16B, SEC16B | rs9686661 | C5orf67 |
| rs1754680 | - | rs555784 | RPS6P12 | rs9687832 | C5orf67 |
| rs1758632 | UBAP2 | rs55653563 | LINC02603 | rs9687833 | C5orf67 |
| rs17624303 | LINC01102 | rs55700915 | RCOR1, TRAF3 | rs972119 | HIVEP2 |
| rs177045 | NEUROG3, TMEM256P1 | rs55775214 | RN7SL366P, QKI | rs972283 | KLF14, H4P1 |
| rs17747955 | NOL4 | rs55857387 | FAM234A | rs9784137 | Y_RNA |
| rs17819328 | TSEN2, PPARG | rs55858476 | ATP2A3 | rs978444 | - |
| rs1783541 | SCYL1 | rs56008953 | RNU2-9P, EPC2 | rs980183 | LINC01793, LINC01122 |
| rs1794138 | TRIM51DP, OR4A1P | rs56094641 | FTO | rs982077 | USP3 |
| rs1796330 | TSPAN8 | rs56200889 | ARAP1 | rs9828933 | PSMD6 |
| rs1799884 | GCK | rs56337234 | FGFR3, TACC3 | rs9847133 | LMCD1-AS1 |
| rs1800437 | GIPR | rs56394279 | TRIM59, B3GAT3P1 | rs9854769 | IGF2BP2 |
| rs1801212 | WFS1 | rs565491 | ANK1 | rs9857204 | CPNE4 |
| rs1801214 | WFS1 | rs567185 | NAV1, IPO9-AS1 | rs9859381 | CASR |
| rs1801645 | PIM3 | rs56799554 | LINC00910 | rs9859406 | IGF2BP2 |
| rs1812707 | ADAMTSL3 | rs56805921 | ETV1 | rs9860730 | ADAMTS9-AS2 |
| rs1815591 | SLCO4A1 | rs56823429 | CMIP | rs9866168 | TFRC, LINC00885 |
| rs1825307 | USP48 | rs57235767 | DEUP1, SLC36A4 | rs9869477 | RBMS3 |
| rs182719694 | PMP2 | rs57286125 | TOMM22P3, KL | rs9872347 | TFRC, LINC00885 |
| rs1845900 | UBE2E2 | rs57327348 | XKR6 | rs9873519 | SLC12A8 |
| rs1850421 | - | rs5744598 | POLK | rs9873618 | SLC2A2 |
| rs1861410 | LINC01122 | rs5751061 | EP300-AS1, L3MBTL2 | rs987949 | NKX6-1 |
| rs1861612 | DNER | rs5753043 | HORMAD2, LIF-AS1 | rs9891146 | C17orf58 |
| rs1868139 | MIR514A3, SLIRPP1 | rs5758223 | EP300 | rs989128 | CACNA1G-AS1 |
| rs1873747 | ROR2 | rs5762925 | ZNRF3 | rs9892728 | RAMP2 |
| rs1874832 | LINC02206, HMGN2P47 | rs576674 | TOMM22P3, KL | rs9894220 | UBE2Z |
| rs1877712 | PNKD | rs5771069 | IL17REL | rs9911983 | OSBPL7 |
| rs1885234 | ASTN2 | rs58202132 | SNTB1 | rs9912236 | LINC01978, LINC01979 |
| rs1894299 | DUSP9 | rs58304657 | GIPR | rs9913225 | TWF1P1, CRYBA1 |
| rs1903002 | FAM13A | rs583769 | HDAC9 | rs9927842 | PDXDC1 |
| rs1904096 | - | rs58383906 | RBMS3-AS3, RBMS3 | rs9931702 | AKTIP |
| rs1914711 | AGTR2, API5P1 | rs5848 | GRN | rs993380 | SCD5 |
| rs1922879 | CNTNAP2 | rs58524310 | LINC01629, RN7SKP17 | rs9936385 | FTO |
| rs1929883 | MTND2P8 | rs5945326 | KRT18P48, DUSP9 | rs9937296 | ZFPM1 |
| rs193457 | FGFR4, NSD1 | rs5946791 | MYCLP2, KRT8P17 | rs9939609 | FTO |
| rs1955163 | TMEM256P1, TSPAN15 | rs59489841 | NLGN1 | rs9940149 | FAM234A |
| rs1968204 | DPY19L2P2, RPL23AP95 | rs59646751 | IGF1R | rs9948462 | LAMA1 |
| rs197374 | INKA2, INKA2-AS1 | rs59944054 | TCEA2 | rs9957145 | GRP, SEC11C |
| rs197379 | INKA2 | rs6005338 | LINC01638 | rs9958640 | - |
| rs197482 | HIVEP2, LINC02828 | rs6011155 | ZBTB46 | rs9967620 | NANOS2, MYPOP |
| rs1996617 | LRRC66, DCUN1D4 | rs6017317 | R3HDML, FITM2 | rs9991328 | FAM13A |
| rs2008027 | HEY2, LINC02523 | rs2049477 | SDK1-AS1 | rs2055901 | HMG20A |
| rs2010825 | GCK | rs2050188 | TSBP1-AS1 | rs2055942 | GABRA4 |
| rs2011603 | LCORL | rs2052261 | RAB1A | rs2055997 | CDKL2 |
| rs2028150 | SPRED2 | rs2032844 | TTLL6 | rs2033159 | ZEB2 |
| rs2028299 | AP3S2 | rs2032912 | NFAT5, CYB5B | rs2040792 | NF1 |
| rs2032217 | CDH7 |  |  |  |  |
| **e. GWAS Risk variants of T1D unique to and shared among populations** | | | | | |
| **Intersection of populations** | **RSID** | **Gene Mapped to** | **Intersection of populations** | **RSID** | **Gene Mapped to** |
| AFR:EAS:SAS | rs60587303 | STK39 | AFR:EAS:AMR | rs2237897 | KCNQ1 |
| AFR | rs2128344 | LINC00320 | AFR:EUR:AMR | rs10933559 | FARP2 |
|  | rs7524087 | LINC02771 |  | rs13018977 | SEPTIN2 |
|  | rs79538630 | SLC15A3, CD6 |  | rs74537115 | LINC02341 |
| AFR:SAS | rs10258381 | - | EAS | rs3757787 | SND1, PAX4 |
| EAS:EUR | rs35122968 | MTCO3P1, HLA-DQB3 | AFR:SAS:EUR:AMR | rs10517086 | LINC02357 |
| EAS:EUR:AMR | rs72802342 | CTRB2, ZFP1 |  | rs11203203 | UBASH3A |
| EAS:SAS:EUR | rs74629033 | PLGRKT |  | rs11258747 | PRKCQ |
| EAS:SAS:EUR:AMR | rs1050979 | IRF4 |  | rs12444268 | GP2, UMOD |
|  | rs10961433 | NFIB |  | rs12464462 | MIR4432HG |
|  | rs17388568 | ADAD1 |  | rs16956936 | DNAH2 |
|  | rs1893217 | PTPN2 |  | rs17143056 | EEF1A1P27 |
|  | rs2069762 | IL2 |  | rs1805731 | M6PR |
|  | rs34740712 | TLL1 |  | rs193778 | RMI2 |
|  | rs35766765 | PHF20L1 |  | rs2641348 | ADAM30 |
|  | rs402072 | PRKD2 |  | rs3093664 | TNF |
|  | rs478222 | EFR3B |  | rs34090353 | RPAP2 |
|  | rs57209021 | ARHGAP27P2, RDM1P3 |  | rs516246 | FUT2 |
|  | rs61759532 | ACAP1 |  | rs55846421 | NEMF |
|  | rs72853903 | MIR4686, ASCL2 |  | rs6547853 | FOSL2, PLB1 |
|  | rs9405661 | EXOC2, IRF4 |  | rs7171171 | RASGRP1, LINC02694 |
| EUR:AMR | rs968567 | FADS2, FADS1 |  | rs7805218 | ITGB8 |
| EUR | rs113010081 | CCRL2, LINC02009 |  | rs7903146 | TCF7L2 |
|  | rs114378220 | CAMK4 | SAS:EUR:AMR | rs10213692 | ANKRD55 |
|  | rs12720356 | TYK2 |  | rs1052553 | MAPT |
|  | rs45626136 | TRIM39-RPP21, RPP21 |  | rs1265564 | CUX2 |
|  | rs61839660 | IL2RA |  | rs151234 | CLN3 |
|  | rs6679677 | RSBN1, PHTF1 |  | rs17696736 | NAA25 |
| SAS | rs41295121 | RPL32P23, RBM17 |  | rs17711850 | BACH2 |
|  | rs77155259 | SLC4A10 |  | rs1983853 | LPAR3 |
|  | rs78824139 | EYS |  | rs2647044 | MTCO3P1, HLA-DQB1 |
| SAS:EUR | rs12722495 | IL2RA |  | rs3024505 | IL10, Y_RNA |
|  | rs78037977 | SLC25A38P1, FASLG |  | rs72928038 | BACH2 |
| AFR:EAS:SAS:EUR:AMR | | | | | |
| rs1004446 | INS-IGF2 | rs2229238 | IL6R | rs56337234 | FGFR3, TACC3 |
| rs10137082 | EFS, IL25 | rs2269241 | PGM1 | rs56380902 | GSDMB |
| rs10169963 | MIR3681HG | rs2269242 | - | rs56994090 | MEG3 |
| rs10245867 | JAZF1 | rs2269247 | PGM1 | rs574384 | PSMB2 |
| rs10255565 | HECW1 | rs2281808 | SIRPG | rs5753037 | HORMAD2, LIF-AS1 |
| rs10277986 | COBL | rs2290400 | GSDMB | rs6043409 | SIRPG, SIRPG-AS1 |
| rs10509540 | RNLS | rs2292239 | ERBB3 | rs605093 | FLI1 |
| rs10751776 | MIR4425, RUNX3 | rs229533 | C1QTNF6 | rs607703 | FLI1 |
| rs10758593 | GLIS3 | rs229541 | C1QTNF6 | rs62447205 | IKZF1 |
| rs10786436 | HPSE2 | rs2304256 | TYK2 | rs645078 | RPS6KA4 |
| rs10788599 | RNLS | rs238265 | LINC02341 | rs6461299 | AHR |
| rs10795791 | RPL32P23, IL2RA | rs241427 | TAP2 | rs6476839 | GLIS3 |
| rs10811662 | CDKN2B-AS1, DMRTA1 | rs2471863 | HLA-K, HLA-U | rs6518350 | GATD3A |
| rs10876866 | RPS26, ERBB3 | rs2476601 | PTPN22 | rs653178 | ATXN2 |
| rs10919543 | FCGR3A, RPS23P10 | rs2542151 | PTPN2, LINC01882 | rs6534347 | KIAA1109 |
| rs10974438 | GLIS3 | rs2597169 | PRR15L | rs6691977 | CAMSAP2 |
| rs11033048 | SLC1A2 | rs2611215 | LINC01179 | rs689 | INS, INS-IGF2 |
| rs11052552 | LINC02390, CLEC2D | rs2664170 | GAB3 | rs6897932 | IL7R |
| rs1110458 | USP31, HS3ST2 | rs2816313 | RGS1 | rs7020673 | GLIS3 |
| rs11120029 | TATDN3 | rs2847278 | LINC01882, PTPN2 | rs705699 | RAB5B |
| rs11171739 | ERBB3 | rs2872507 | ZPBP2, GSDMB | rs705705 | RPS26 |
| rs11203202 | UBASH3A | rs2903692 | CLEC16A | rs706779 | IL2RA |
| rs113881148 | DGKQ | rs2941522 | GRB7, IKZF3 | rs7111341 | MIR4686, ASCL2 |
| rs11651753 | PNPO, PRR15L | rs3087243 | ICOS, CTLA4 | rs7167984 | SEPHS1P2, RGMA |
| rs11755527 | BACH2 | rs3128930 | COL11A2P1, HLA-DPB2 | rs7202877 | CTRB2, CTRB1 |
| rs11954020 | IL7R, CAPSL | rs3134938 | TSBP1-AS1, NOTCH4 | rs7221109 | CCR7, SMARCE1 |
| rs12128789 | LINC02773, BATF3 | rs3135365 | TSBP1-AS1, HLA-DRA | rs722988 | IATPR, NRP1 |
| rs12251307 | RBM17, RPL32P23 | rs3184504 | ATXN2, SH2B3 | rs72727394 | RASGRP1 |
| rs12416116 | RNLS | rs337637 | KLF3-AS1 | rs7301381 | KLRG1 |
| rs12453507 | GSDMB, ZPBP2 | rs34593439 | CTSH | rs7313065 | SPRYD3 |
| rs12644686 | LINC02362 | rs34941730 | HLA-A, HLA-W | rs743777 | IL2RB |
| rs1265057 | C6orf15, RNU6-1133P | rs3741208 | IGF2-AS, INS-IGF2 | rs7567242 | MIR4432HG, RNA5SP94 |
| rs12665429 | LINC02539 | rs3757247 | BACH2 | rs757411 | CCR7, SMARCE1 |
| rs12679857 | RNU6-12P, TNFRSF11B | rs3764021 | CLEC2D | rs7582694 | STAT4 |
| rs12708716 | CLEC16A | rs3788013 | UBASH3A | rs763361 | CD226 |
| rs12712067 | AFF3 | rs3802214 | AGO2 | rs7760731 | HLA-DQA1, HLA-DRB1 |
| rs12742756 | INPP5B | rs3802604 | GATA3 | rs7795896 | CFTR |
| rs12927355 | CLEC16A | rs3825932 | CTSH | rs7804356 | SKAP2 |
| rs12971201 | PTPN2 | rs3842727 | TH, INS | rs78896587 | IGF2, H19 |
| rs13147255 | LINC02516, ANKRD50 | rs3842753 | INS-IGF2, INS | rs7936434 | EMSY, LINC02757 |
| rs13259300 | COLEC10 | rs3997848 | HLA-DQB3, MTCO3P1 | rs8035957 | RASGRP1 |
| rs13415583 | AFF3 | rs4084127 | FGF4, FGF3 | rs8046043 | - |
| rs138748427 | HLA-DRB9, HLA-DRB5 | rs4148870 | TAP2 | rs8056814 | CTRB2, CTRB1 |
| rs1405209 | STX17-AS1, NR4A3 | rs416603 | RMI2 | rs855330 | PGM1 |
| rs1445898 | CAPSL | rs4238595 | GP2, UMOD | rs857786 | OR10AA1P, MNDA |
| rs1456988 | LINC01550 | rs425105 | PRKD2 | rs911263 | RAD51B |
| rs1465788 | ZFP36L1, MAGOH3P | rs4263714 | COBL | rs917911 | CD69 |
| rs1493696 | NOTCH2 | rs4320356 | BTN2A3P | rs924043 | - |
| rs1534422 | MIR3681HG | rs4366464 | INS-IGF2, IGF2-AS | rs926169 | CTLA4, RNU6-474P |
| rs1538171 | MIR588, CENPW | rs4505848 | KIAA1109 | rs9264758 | RPL3P2 |
| rs1578060 | CENPW, MIR588 | rs4513644 | LINC02225 | rs926592 | TSBP1, TSBP1-AS1 |
| rs1615504 | CD226 | rs4763879 | CD69 | rs9268576 | HLA-DRA, TSBP1-AS1 |
| rs1701704 | IKZF4 | rs4788084 | IL27, NUPR1 | rs9268645 | HLA-DRA |
| rs17166496 | FSTL4 | rs4820830 | HORMAD2 | rs9272346 | HLA-DQA1 |
| rs17407280 | KIF17 | rs4837404 | PTGES | rs9273363 | HLA-DQB1, HLA-DQA1 |
| rs1770 | HLA-DQB1-AS1, HLA-DQB1 | rs4849135 | ACOXL | rs9273367 | HLA-DQA1, HLA-DQB1 |
| rs1876142 | TTC33 | rs4900384 | LINC01550 | rs9275358 | HLA-DQB1, MTCO3P1 |
| rs1881146 | MIR3681HG | rs4929965 | MIR4686, ASCL2 | rs9388489 | CENPW |
| rs1947178 | TOX | rs4948088 | COBL | rs941576 | MEG3 |
| rs1990760 | IFIH1 | rs537544 | GATA3 | rs947474 | LINC02649 |
| rs2111485 | IFIH1, FAP | rs539514 | LMO7 | rs9501130 | LINC01149, HCP5 |
| rs212408 | TAGAP-AS1 | rs550448 | JAZF1-AS1 | rs9585056 | TM9SF2, CCR12P |
| rs2165738 | NCOA1, ITSN2 | rs56178904 | GATD3A | rs9653442 | LINC01104 |
|  |  |  |  | rs9976767 | UBASH3A |
| **f. GWAS Risk variants of CKD unique to and shared among populations** | | | | | |
| **Intersection of populations** | **RSID** | **Gene Mapped to** | **Intersection of populations** | **RSID** | **Gene Mapped to** |
| AFR:SAS:EUR:AMR | rs11645800 | - | EAS:SAS:EUR:AMR | rs10941694 | HCN1 |
|  | rs12207180 | SLC22A2 |  | rs2411192 | MYO19 |
|  | rs13538 | ALMS1P1, NAT8 |  | rs6066043 | SLC13A3 |
|  | rs6546869 | ALMS1P1 |  | rs6497441 | GRIN2A |
|  | rs73728279 | PRKAG2 |  | rs77924615 | PDILT |
|  | rs7805747 | PRKAG2 |  | rs9535020 | RB1 |
| AMR | rs10783124 | AGL, SLC35A3 | EUR | rs1044261 | IDI2, GTPBP4 |
| EAS | rs13070584 | COL8A1 | SAS:EUR | rs12736457 | PPM1J, PPM1J-DT |
|  | rs183951714 | CDON |  | rs16864170 | SOX11, LINC01810 |
| SAS:EUR:AMR | rs11039182 | MADD |  |  |  |
|  | rs12917707 | PDILT, UMOD |  |  |  |
|  | rs267734 | ANXA9, CERS2 |  |  |  |
| AFR:EAS:SAS:EUR:AMR | | | | | |
| rs10109414 | STC1 | rs17319721 | SHROOM3 | rs4976646 | RGS14 |
| rs1032843 | RNU6-627P | rs17730281 | WDR72 | rs61897431 | SLC39A13, MIR4487 |
| rs1047891 | CPS1 | rs690346 | WDR72 | rs626277 | DACH1 |
| rs10774021 | SLC6A13 | rs1933182 | SYPL2, PSMA5 | rs6420094 | SLC34A1 |
| rs1078442 | TFCP2L1 | rs2039424 | PIP5K1B | rs6431731 | LINC01804 |
| rs10794720 | WDR37 | rs2279463 | SLC22A2 | rs6465825 | RSBN1L, TMEM60 |
| rs10846157 | RERG | rs2440165 | SLC47A1 | rs653178 | ATXN2 |
| rs10906850 | FAM171A1, NMT2 | rs2453533 | RNU6-953P, SLC28A2 | rs1813937 | OR4C50P, LINC02750 |
| rs11123169 | PSD4, PAX8-AS1 | rs2467853 | SPATA5L1 | rs7123489 | AP5B1, KRT8P26 |
| rs1145077 | GATM | rs2490391 | SDCCAG8 | rs7127946 | OR4X2, OR4B2P |
| rs11959928 | C9, DAB2 | rs2727040 | TRIM49B | rs7129190 | HTR3B |
| rs12460876 | SLC7A9 | rs347685 | TFDP2 | rs7475348 | MYPN |
| rs12509595 | FGF5, PRDM8 | rs2823139 | CYCSP42, NRIP1 | rs759341 | RRM2, RN7SL66P |
| rs1260326 | GCKR | rs2837554 | DSCAM | rs7740107 | L3MBTL3 |
| rs12913015 | LINC02694 | rs28817415 | SHROOM3 | rs80282103 | LARP4B |
| rs13072153 | HACD2, ADCY5 | rs2971880 | SPTBN1 | rs8096658 | NFATC1 |
| rs13157326 | RAI14, C1QTNF3-AMACR | rs2749153 | ZNF436-AS1, TCEA3 | rs8101667 | CEP89 |
| rs13230625 | UNCX | rs34861762 | RNU1-148P, STC1 | rs868822 | LINC01006 |
| rs13329952 | UMOD | rs3812036 | SLC34A1 | rs875860 | CDH23 |
| rs1362800 | C9, DAB2 | rs4408777 | RGS19 | rs881858 | LINC02537, VEGFA |
| rs1394125 | UBE2Q2 | rs4014195 | AP5B1, KRT8P26 | rs911119 | CST3 |
| rs1505141 | - | rs4293393 | UMOD | rs9310709 | SALL4P5, RPL24P7 |
| rs1548945 | FABP5P14, RNA5SP120 | rs3925584 | DCDC1, MPPED2-AS1 | rs963837 | DCDC1, MPPED2-AS1 |
| rs16942751 | AQP4-AS1 | rs4744712 | PIP5K1B | rs9375818 | ARG1 |
| rs1719934 | EPB41L3 | rs491567 | WDR72 | rs9895661 | BCAS3 |
| **g. GWAS Risk variants of High BMI unique to and shared among populations** | | | | | |
| **Intersection of populations** | **RSID** | **Gene Mapped to** | **Intersection of populations** | **RSID** | **Gene Mapped to** |
| AFR:SAS:EUR:AMR | rs115474151 | SLC7A14, SLC7A14-AS1 | EAS:SAS:EUR:AMR | rs13104545 | - |
|  | rs1161397 | TFAP2D | EUR | rs75398113 | SNRPC |
|  | rs12735657 | SEC16B | EUR:AMR | rs13135092 | SLC39A8 |
| AFR:EAS:SAS:EUR:AMR | | | | | |
| rs11209947 | FTO | rs2384060 | ADCY3 | rs6748821 | TMEM18, LINC01875 |
| rs112446794 | CEP120 | rs3760091 | SULT1A1 | rs7132908 | FAIM2 |
| rs12101726 | RPL31P12 | rs4447506 | PIK3C3 | rs7927262 | FAIM2 |
| rs12507026 | - | rs506589 | SEC16B | rs8096590 | KIAA1549L, HIPK3 |
| rs1475010 | LINC02306 | rs57988840 | TFAP2B, RPS17P5 | rs9928094 | LINC01541 |
| rs1993709 | RPL31P12 | rs66906321 | LINC01875, TMEM18 | rs9930333 | FTO |
| rs2168711 | MC4R, RNU4-17P | rs6738433 | DNAJC27, ADCY3 |  |  |
| **h. GWAS Risk variants of High LDL unique to and shared among populations** | | | | | |
| **Intersection of populations** | **RSID** | **Gene Mapped to** | **Intersection of populations** | **RSID** | **Gene Mapped to** |
| AFR:EAS | rs74257940 | LDLR | AFR:EAS:AMR | rs35443082 | CEACAM20 |
| AFR | rs1081105 | APOE, APOC1 |  | rs919213 | NR1H4 |
|  | rs11206517 | PCSK9 | AFR:EAS:EUR | rs1065853 | APOE, APOC1 |
|  | rs113190300 | LDLR, SMARCA4 |  | rs7412 | APOE |
|  | rs11568318 | UGT1A6, UGT1A7, UGT1A9, UGT1A5, UGT1A8, UGT1A10, UGT1A3, UGT1A4 | AFR:EAS:EUR:AMR | rs17029617 | CMTM6 |
|  | rs11669133 | SMARCA4 |  | rs17047429 | RNU7-190P, DDX18 |
|  | rs11669576 | LDLR |  | rs3773777 | CMTM6 |
|  | rs11672431 | QTRT1 |  | rs7640978 | CMTM6 |
|  | rs12106385 | - |  | rs9312517 | SPOCK3 |
|  | rs12721054 | APOC1 |  | rs9834932 | CMTM6 |
|  | rs140480140 | APOC1 | AFR:EAS:SAS | rs141622900 | APOC1P1, APOC1 |
|  | rs140584594 | GSTM1, GSTM2 |  | rs2282143 | SLC22A1 |
|  | rs145859971 | APOA4, APOA5 |  | rs56903760 | ALDH1A2 |
|  | rs157599 | APOC1P1, APOC1 |  | rs7108486 | TRIM6-TRIM34, TRIM5 |
|  | rs17111684 | USP24 |  | rs7254892 | NECTIN2 |
|  | rs28362263 | PCSK9 |  | rs76898656 | ALDH1A2 |
|  | rs28399613 | BCAM |  | rs928911 | PPP1R13L |
|  | rs35804417 | GATB | AFR:EAS:SAS:AMR | rs11810371 | PCSK9, BSND |
|  | rs389261 | APOC1 |  | rs16980741 | HNRNPMP2, SLC1A5 |
|  | rs394819 | TOMM40 |  | rs2278426 | DOCK6, ANGPTL8 |
|  | rs490243 | TOMM40 |  | rs4971516 | LDAH |
|  | rs5471 | TXNL4B, HP |  | rs57618243 | OTX2-AS1 |
|  | rs56315738 | SMARCA4 |  | rs8101801 | DOCK6 |
|  | rs62219001 | - | AFR:EAS:SAS:EUR | rs11671653 | DNM2 |
|  | rs6693893 | CELSR2 |  | rs17145738 | TBL2 |
|  | rs6752026 | APOB |  | rs77287598 | SMARCA4 |
|  | rs7200153 | PMFBP1, LINC01572 |  | rs8176746 | ABO |
|  | rs72902579 | TDRD15, APOB | AFR:EUR | rs55660224 | MYBPHL |
|  | rs72911441 | USP24 |  | rs61988556 | ZFYVE1 |
|  | rs73219351 | PSMC1P10, ZFYVE9P2 |  | rs67038483 | SCAF4 |
|  | rs73339979 | GLTPD2, PSMB6 |  | rs7249753 | LDLR, SMARCA4 |
|  | rs73667470 | LPL | AFR:SAS | rs112170101 | RELB |
|  | rs73729083 | CREB3L2 |  | rs1499279 | ITGA1 |
|  | rs73936968 | TOMM40 |  | rs66900043 | ABCG8 |
|  | rs74017329 | RNF111 |  | rs72798837 | ABCG8 |
|  | rs7569328 | LINC02850, APOB |  | rs73921514 | GTF3C2 |
|  | rs76366838 | TOMM40 | AFR:SAS:AMR | rs11920719 | TNIK |
|  | rs7775698 | HBS1L |  | rs73025516 | IGF2R |
|  | rs79783247 | GCNT4, RPL27AP5 | AMR | rs117859111 | PNLDC1, MAS1 |
|  | rs9784624 | TIMD4, HAVCR1 |  | rs140570886 | LPA |
| AFR:EUR:AMR | rs11206510 | BSND, PCSK9 |  | rs145730801 | SERPINA6, SERPINA10 |
|  | rs12246352 | C10orf88 |  | rs147032017 | ZFPM1-AS1, ZFPM1 |
|  | rs12272004 | BUD13, LINC02702 |  | rs1800777 | CETP |
|  | rs147539187 | ADTRP, AMD1P4 |  | rs1800961 | HNF4A |
|  | rs2498323 | HGFAC |  | rs7110984 | IFT46 |
|  | rs3135506 | APOA5 | EAS | rs11066015 | ACAD10 |
|  | rs41360247 | ABCG8 |  | rs13306194 | APOB |
|  | rs4953023 | ABCG8 |  | rs139262716 | Y_RNA |
|  | rs6756629 | ABCG8, ABCG5 |  | rs141965732 | IFT81 |
|  | rs679224 | ELAPOR1 |  | rs34480710 | MIR4422HG |
|  | rs684138 | CELSR2 |  | rs57825321 | APOB |
|  | rs72875462 | ABCG8 |  | rs79105258 | CUX2 |
|  | rs72905574 | TMEM61, DHCR24-DT |  | rs79106033 | LINC02850, APOB |
|  | rs75331444 | ABCG8 | EAS:EUR:AMR | rs12920967 | LINC01572, PMFBP1 |
|  | rs76866386 | ABCG8 |  | rs2066717 | ABCA1 |
|  | rs9470298 | AMD1P4 |  | rs2303857 | DYNC1LI1 |
| EAS:EUR | rs28695210 | RHCE |  | rs74629722 | LINC02850, APOB |
|  | rs2937738 | SV2C |  | rs870992 | ITGA1 |
| EAS:AMR | rs117733303 | LPAL2 | EAS:SAS | rs10462509 | RPL27AP5, FAM169A |
|  | rs17036085 | PSRC1, MYBPHL |  | rs11555891 | IRGC |
|  | rs204906 | CLPTM1 |  | rs12469758 | NUTF2P8 |
|  | rs3798220 | LPA |  | rs41280378 | OIT3 |
|  | rs75061399 | NR1H4 |  | rs57176252 | OIT3 |
|  | rs79073069 | NR1H4 |  | rs75214121 | CYP7A1, UBXN2B |
|  | rs926664 | RNA5SP484, MAFB |  | rs77348447 | VN1R18P |
|  | rs9357121 | USP8P1, HLA-C |  | rs79018068 | MARK4, PPP1R37 |
| EAS:SAS:AMR | rs11220477 | ST3GAL4 | SAS | rs10205003 | NUTF2P8, TDRD15 |
|  | rs5763662 | MTMR3 |  | rs114523982 | RPL27AP5, FAM169A |
| EAS:SAS:EUR | rs112771035 | ST3GAL4 |  | rs116958755 | BLOC1S3, MARK4 |
|  | rs117692263 | CD163L1, CD163 |  | rs13408439 | RN7SL117P, LINC01822 |
|  | rs2401 | GSEC, DCPS |  | rs73013139 | SMARCA4 |
|  | rs769446 | APOE, TOMM40 |  | rs73121275 | RNU2-52P, RNA5SP484 |
|  | rs76970536 | ST3GAL4 |  | rs73596816 | LPA |
| SAS:AMR | rs8102280 | MAU2 |  | rs77987064 | FAM110B, UBXN2B |
| SAS:EUR:AMR | | | | | |
| rs1043879 | RSRP1 | rs13076933 | PPARG, GSTM5P1 | rs41280463 | TRIM2 |
| rs10490625 | INSIG2 | rs13098031 | SYN2, GSTM5P1 | rs4722551 | NFE2L3, MIR148A |
| rs10928512 | TMEM163 | rs13288021 | DENND4C, PLIN2 | rs56369308 | USP3, LINC02568 |
| rs11550029 | NUDCD3 | rs17696736 | NAA25 | rs62011285 | USP3, LINC02568 |
| rs11066301 | PTPN11 | rs17789218 | SIM1, PRDX2P4 | rs62132803 | FGF21, RNU6-317P |
| rs115030251 | MYBPHL | rs1874124 | MTARC1 | rs662138 | SLC22A1 |
| rs115478735 | ABO | rs2228671 | LDLR | rs72729610 | TRIM2 |
| rs11065987 | ATXN2-AS, BRAP | rs2504959 | SLC22A2, SLC22A3 | rs72823013 | NHLRC2, ADRB1 |
| rs11652957 | EFCAB13 | rs34150222 | DENND4C | rs72823020 | ADRB1, NHLRC2 |
| rs11668327 | TOMM40 | rs34503352 | ZNF329 | rs73107473 | NPC1L1 |
| rs117492019 | ZNF329, ZNF274 | rs35081008 | ZNF329 | rs76177059 | NPHP3-ACAD11, NPHP3 |
| rs11753995 | SLC22A1 | rs35106244 | FUT2 | rs76240114 | UQCRHL, RPL12P14 |
| rs12751807 | CNIH4, NVL | rs35866622 | MAMSTR | rs76702063 | ATXN1L, IST1 |
| rs12983728 | ZNF329, ZNF274 | rs9884390 | UGT2B29P, TMPRSS11E | rs7735249 | ARL15 |
| rs13015955 | LINC01884 | rs41279633 | NPC1L1 | rs80051818 | EIF2B4 |
| rs373579 | FCGR2A |  |  |  |  |
| SAS:EUR | | | | | |
| rs113177823 | DNAJC13 | rs34243815 | DOCK6 | rs73001065 | MAU2 |
| rs1160984 | TOMM40 | rs35271870 | SARS1 | rs73011369 | SMARCA4 |
| rs11800243 | PCSK9 | rs369298568 | ATP8B1 | rs73496517 | ATG4D |
| rs12718462 | APOA1, APOC3 | rs4560319 | PXK | rs76967117 | ILRUN |
| rs13315871 | PXK | rs55784804 | SHBG | rs77241309 | BCAM, NECTIN2 |
| rs17561351 | NECTIN2 | rs6907508 | ILRUN | rs78946096 | DNAJC13 |
| rs17657025 | ZNF226 | rs71311871 | PDHB, KCTD6 | rs79355265 | NUTF2P8, TDRD15 |
| rs1800027 | FUT2 | rs72731954 | TCF12 | rs9825431 | PXK |
| rs28709068 | PXK | rs72733928 | TCF12 | rs9844432 | PXK |
| EUR:AMR | | | | | |
| rs111174163 | HDGFL2 | rs35315097 | LINC01460 | rs71367412 | RAI1 |
| rs11244035 | OBP2B | rs412334 | TMEM258 | rs72750932 | USP3 |
| rs11670384 | SLC44A2 | rs41302673 | STKLD1 | rs72938315 | CYP20A1 |
| rs12143028 | USP24 | rs4253772 | PPARA | rs73009507 | C19orf38 |
| rs13135092 | SLC39A8 | rs61797139 | SYPL2, PSMA5 | rs73013159 | SMARCA4 |
| rs17476364 | HK1 | rs61871243 | NAP1L4 | rs73050293 | NECTIN2 |
| rs17664900 | PKD1L3 | rs61876729 | GATD1, CEND1 | rs75109222 | HAVCR2 |
| rs17580 | SERPINA1 | rs62022693 | CSNK1G1, PCLAF | rs75120785 | RNU6-1151P, RNU6-526P |
| rs17565182 | SAP18P1, BIN2P2 | rs62064572 | HOXB-AS4, HOXB7 | rs76692773 | TOMM40 |
| rs1801701 | APOB | rs62118464 | MARK4, PPP1R37 | rs76995491 | TDRD15, NUTF2P8 |
| rs267733 | ANXA9 | rs62210638 | LINC01370, MAFB | rs77680021 | ACADVL |
| rs35127135 | TDRD15, NUTF2P8 | rs62259757 | DUSP7, RPL29 | rs78073763 | PPP1R37, MARK4 |
| rs79588679 | CTAGE1 | rs8111071 | RSPH6A |  |  |
| EUR1 | | | | | |
| rs10455872 | LPA | rs145090930 | HYDIN | rs72768400 | SLC25A5P9, POC5 |
| rs10490626 | INSIG2, RN7SL111P | rs150474434 | RN7SL111P, INSIG2 | rs72774870 | LINC00570 |
| rs10957055 | UBXN2B, CYP7A1 | rs17242346 | SMARCA4, LDLR | rs72828723 | CMAHP |
| rs111548358 | TDRD15, APOB | rs17660708 | SCAF4 | rs72838078 | INSIG2, RN7SL111P |
| rs112639655 | R3HDM2 | rs17698734 | LINC01497, LINC01483 | rs72839644 | THORLNC |
| rs114067101 | HLA-DRB9, HLA-DRB5 | rs191555775 | LPA | rs72961013 | RSPO3, RNF146 |
| rs114863007 | SNRPC | rs34095326 | TOMM40 | rs73070437 | DNAH11 |
| rs11542916 | AP1M2 | rs34333163 | SLC39A8 | rs73201545 | SCAF4 |
| rs11693526 | - | rs34592089 | BANK1 | rs73230015 | STAG1 |
| rs117210556 | SMARCA4 | rs35814746 | ZSCAN31 | rs74617384 | LPA |
| rs118039278 | LPA | rs36005514 | SMARCA4, LDLR | rs75211012 | NUTF2P8, LINC01822 |
| rs12208357 | SLC22A1 | rs4970829 | SARS1 | rs75262919 | PPARG |
| rs12464355 | INSIG2 | rs55809639 | INSIG2, RN7SL111P | rs76079263 | ZSCAN16-AS1 |
| rs12691202 | APOB | rs56208677 | CETP | rs76095103 | AP1G1 |
| rs12748152 | RN7SL165P, PIGV | rs56212732 | HYDIN | rs77907512 | ITSN2 |
| rs13016086 | BABAM2 | rs62123888 | LINC02850, LDAH | rs78425119 | IGF2R |
| rs13107325 | SLC39A8 | rs62366598 | CERT1 | rs79953286 | DNAJC13 |
| rs141783576 | RSPO3 | rs71559014 | RPL10P2, MIR3143 | rs80000866 | CEACAM16-AS1 |
| EAS:SAS:EUR:AMR | | | | | |
| rs10272002 | C7orf50 | rs16923637 | NSMAF | rs3812594 | SEC16A |
| rs10417602 | MARK4, NKPD1 | rs17050272 | LINC01101, Y_RNA | rs3812945 | SCAMP5 |
| rs10490120 | FSHR, CTBP2P5 | rs17216525 | CILP2, PBX4 | rs3865427 | NECTIN2 |
| rs10851478 | FAM227B | rs17345563 | DNAJC13 | rs4135275 | PPARG |
| rs10876168 | GALNT6 | rs17404153 | DNAJC13 | rs4871601 | LINC00861, TRIB1 |
| rs10877955 | SLC2A13 | rs174533 | MYRF, TMEM258 | rs55649657 | DNAH11 |
| rs11216126 | LINC02702, BUD13 | rs174541 | FADS2 | rs55696093 | DNAH11 |
| rs11220462 | ST3GAL4 | rs174546 | FADS2, FADS1 | rs55714927 | ASGR1 |
| rs11220463 | ST3GAL4 | rs174547 | FADS2, FADS1 | rs56236159 | RPS7, COLEC11 |
| rs1122608 | SMARCA4 | rs174549 | FADS2, FADS1 | rs566865466 | FAM227B |
| rs112595563 | DUS2 | rs174550 | FADS1, FADS2 | rs5754102 | UBE2L3 |
| rs11571696 | BRCA2 | rs174551 | FADS1, FADS2 | rs58542926 | TM6SF2 |
| rs11583680 | PCSK9 | rs174553 | FADS1, FADS2 | rs60239918 | GEMIN7-AS1 |
| rs11601507 | TRIM5 | rs174554 | FADS2, FADS1 | rs6129744 | TOP1 |
| rs11648003 | DHODH | rs174570 | FADS2 | rs61962260 | DLEU1 |
| rs11784833 | PARP10 | rs1800978 | ABCA1 | rs62074057 | TBKBP1, TBX21 |
| rs11855284 | ALDH1A2 | rs1907631 | LRRK2, LRRK2-DT | rs62130341 | SEC1P |
| rs12052058 | SMARCA4 | rs1981405 | GAB2 | rs624612 | PCSK9 |
| rs12052201 | SMARCA4 | rs217180 | PMFBP1 | rs6453139 | ANKDD1B |
| rs12055996 | JAZF1 | rs2172241 | LRRK2, LRRK2-DT | rs688 | LDLR |
| rs12145455 | KANK4, USP1 | rs2238675 | NCAN | rs703212 | N4BP2L2 |
| rs12445804 | LITAF | rs2256814 | SLC2A4RG | rs71630059 | NCK1 |
| rs12588415 | YLPM1 | rs2291609 | MECR | rs7188 | KANK2 |
| rs12686004 | ABCA1 | rs2350809 | BMPR2 | rs72480795 | ZNF224, ZNF284 |
| rs12909598 | FAM227B | rs257377 | PRKAR2B | rs72631343 | ABCA10 |
| rs12917376 | LMAN1L | rs2738446 | LDLR | rs72861370 | MAP2K6 |
| rs12931964 | PKD1L3 | rs2738452 | LDLR | rs73066485 | DNAH11 |
| rs1293259 | RHCE | rs28399637 | BCAM | rs740516 | ABCA6 |
| rs12983082 | LDLR | rs287227 | USP24 | rs7544735 | RNU6ATAC35P, MTARC1 |
| rs13161895 | RNF130 | rs34180575 | HLX | rs769449 | APOE |
| rs13284665 | ZER1 | rs35203651 | UGT1A4, UGT1A8, UGT1A6, UGT1A1, UGT1A5, UGT1A9, UGT1A10, UGT1A3, UGT1A7 | rs79316815 | - |
| rs140244541 | WDR12, CARF | rs35786788 | HBS1L | rs7950543 | ATP5MGP1, SPINDOC |
| rs1458038 | PRDM8, FGF5 | rs3750329 | ZER1 | rs79828839 | SGMS1 |
| rs16839065 | RN7SL753P, NOP58 | rs3761081 | MEF2B, BORCS8-MEF2B, BORCS8 | rs869412 | C7orf50 |
| rs16844401 | HGFAC | rs3786722 | SMARCA4 | rs9376090 | HBS1L |
|  |  |  |  | rs960596 | Y_RNA, RBX1 |
| AFR:SAS:EUR:AMR | | | | | |
| rs1042031 | APOB | rs1799898 | LDLR | rs602662 | FUT2 |
| rs10423733 | SMARCA4 | rs1805738 | PHC1 | rs60309576 | B4GALNT3 |
| rs10443196 | PLPPR5 | rs2021092 | XRCC1 | rs611917 | CELSR2 |
| rs10842724 | KLRG1, HNRNPA1P34 | rs217381 | DDX56 | rs62072497 | ALKBH5, LLGL1 |
| rs11014204 | CACNB2 | rs217386 | NPC1L1, DDX56 | rs62522646 | C8orf37-AS1, LINC01298 |
| rs11048036 | PHC1 | rs2434885 | ZNF19 | rs6511720 | LDLR |
| rs11102002 | GSTM5, EPS8L3 | rs2452170 | MAMSTR, FUT2 | rs6657811 | CELSR2 |
| rs111338114 | EPO, ZAN | rs2718717 | LIMS1 | rs6675858 | CNIH4 |
| rs112374545 | SMARCA4 | rs314311 | EPHB4 | rs6709904 | ABCG8 |
| rs11407 | PINLYP | rs314316 | SLC12A9 | rs67648651 | RTL9 |
| rs11544715 | ABCA5 | rs33997707 | HLA-DRB5, RNU1-61P | rs67710536 | RPS6 |
| rs11636087 | LINC02568, USP3 | rs34019521 | B4GALNT3 | rs6785233 | TNIK, SLC2A2 |
| rs11709868 | PCOLCE2, PAQR9 | rs34041362 | TMPRSS11E, MT2P1 | rs679574 | FUT2 |
| rs11806638 | PCSK9 | rs35882350 | B4GALNT3 | rs6909746 | FRK |
| rs12130333 | ATG4C, DOCK7 | rs36178701 | DENND4C | rs6917747 | IGF2R |
| rs12151108 | SMARCA4, LDLR | rs430096 | NUTF2P8, TDRD15 | rs7124487 | NAP1L4 |
| rs12543764 | DMTN | rs4646283 | SLC22A1, SLC22A2 | rs72701021 | ELAPOR1 |
| rs12713956 | APOB | rs4711589 | KCNK17, KCNK5 | rs72951954 | FRK, NT5DC1 |
| rs12740374 | CELSR2 | rs4860987 | UGT2B17, UGT2B15 | rs73015011 | SMARCA4 |
| rs12984266 | CEACAM16-AS1 | rs492602 | FUT2 | rs73015020 | SMARCA4, LDLR |
| rs13179 | EIF1AX | rs4970834 | CELSR2 | rs73447108 | EIF1AX |
| rs139306531 | LDLR, SMARCA4 | rs507766 | FUT2 | rs73545584 | DMTN |
| rs143020224 | SMARCA4 | rs516246 | FUT2 | rs7528419 | CELSR2 |
| rs1521516 | DIP2B | rs516316 | FUT2 | rs7715739 | ANKDD1B |
| rs1529711 | CARM1 | rs518076 | GNAI3 | rs8017377 | NYNRIN |
| rs1564348 | SLC22A1 | rs55804343 | KCNK17, KCNK5 | rs826681 | LIMS1 |
| rs16926246 | HK1 | rs59328596 | DMTN | rs826682 | LIMS1 |
| rs17031488 | RN7SKP66, DYNC2LI1 | rs5942937 | TDGF1P3, RTL9 | rs942922 | RN7SL554P |
| rs17185536 | PRDX2P4, SIM1 | rs5942956 | TDGF1P3 | rs9496567 | SIM1, PRDX2P4 |
| rs17564079 | POC5, SLC25A5P9 | rs5985471 | TDGF1P3, RTL9 |  |  |
| AFR:EAS:SAS:EUR:AMR | | | | | |
| rs1000778 | FADS3 | rs2183573 | BRWD1 | rs55831924 | PLEC |
| rs10038095 | HMGCR | rs221057 | ARHGEF19 | rs55892601 | SP4, RNU1-15P |
| rs10045497 | HMGCR | rs221797 | GIGYF1 | rs55921103 | MITF |
| rs1007938 | ITPR2 | rs221798 | GIGYF1 | rs56000661 | TRIM65 |
| rs10087499 | CYP7A1, UBXN2B | rs2218260 | ALDH1A2 | rs56047090 | PRPF3 |
| rs10096633 | RPL30P9, LPL | rs2235215 | MYLIP | rs56113850 | CYP2A6 |
| rs10102164 | RP1, TRMT112P7 | rs2238162 | BRCA2, IFIT1P1 | rs56130071 | DNAH11 |
| rs10107182 | CYP7A1, UBXN2B | rs2239619 | TRAM2-AS1 | rs56131196 | APOC1, APOC1P1 |
| rs1010759 | FER1L4 | rs2239620 | TRAM2-AS1 | rs56156922 | CETP, HERPUD1 |
| rs10128711 | SPTY2D1 | rs2242391 | SRPRA | rs56204645 | TRMT112P7, RP1 |
| rs1014283 | ABCB4 | rs2244608 | HNF1A, HNF1A-AS1 | rs562338 | APOB, TDRD15 |
| rs10151517 | RPL12P6, CFL2 | rs2247056 | LINC02571 | rs56267346 | CYP2A6 |
| rs10158897 | USP1 | rs2249742 | USP8P1, HLA-C | rs56299595 | PLXND1 |
| rs10164853 | ACVR1C | rs2250802 | GPAM | rs56325564 | KPNB1, TBKBP1 |
| rs10166144 | NUTF2P8, TDRD15 | rs2253736 | SLC2A13, LINC02555 | rs563290 | TDRD15, APOB |
| rs1016988 | IRF1-AS1, SLC22A5 | rs2254287 | COL11A2 | rs56394279 | TRIM59, B3GAT3P1 |
| rs10172650 | APOB | rs2254819 | ABCA1 | rs56402930 | CRTC3, IQGAP1 |
| rs10175646 | LINC02850, APOB | rs2255141 | GPAM | rs56668103 | ZHX3 |
| rs10183198 | - | rs2264750 | C12orf43 | rs568938 | APOB, TDRD15 |
| rs10184004 | GRB14, COBLL1 | rs2266788 | APOA5 | rs57579470 | PPM1N, VASP |
| rs10184376 | ACVR1C | rs2274136 | - | rs5758128 | XPNPEP3 |
| rs10199768 | APOB | rs2281636 | CUTC | rs576573069 | SORCS2 |
| rs10201242 | ACVR1C | rs2285947 | DNAH11 | rs577584 | APOB, TDRD15 |
| rs102275 | TMEM258 | rs2287621 | ABCB11 | rs57940795 | FUT2 |
| rs10260606 | DDX56, NPC1L1 | rs2287622 | ABCB11 | rs581411 | APOB, TDRD15 |
| rs10275712 | C7orf50 | rs2287623 | ABCB11 | rs58198139 | HAVCR1, TIMD4 |
| rs1030431 | UBXN2B | rs2288002 | DHODH | rs584007 | APOE, APOC1 |
| rs1038026 | TOMM40 | rs2291217 | TMEM70 | rs58458113 | IGF2R |
| rs10401969 | SUGP1 | rs2294261 | MDH1P2, MYLIP | rs58466006 | CSNK1G3 |
| rs10402271 | BCAM, NECTIN2 | rs2294915 | PNPLA3 | rs585362 | SARS1, CELSR2 |
| rs10402457 | SPC24 | rs2297372 | IGF2R | rs586178 | RHCE |
| rs10415983 | MARK4, BLOC1S3 | rs2297374 | SLC22A1 | rs5882 | CETP |
| rs1041968 | APOB | rs2297991 | GPAM | rs5904726 | MIR514B, MIR508 |
| rs1042034 | APOB | rs2302429 | POR | rs59325138 | APOC1, APOE |
| rs10420888 | BLOC1S3, EXOC3L2, MARK4 | rs2304088 | CARM1 | rs59379014 | ST3GAL4 |
| rs10421247 | NKPD1, MARK4 | rs2304128 | GMIP | rs59509802 | DNAH11 |
| rs10421404 | CLPTM1, APOC4-APOC2 | rs2304130 | ZNF101 | rs59613675 | MIR4422HG |
| rs10422256 | LDLR | rs2306486 | TMEM70 | rs597808 | ATXN2 |
| rs1042522 | TP53 | rs2328223 | RNU6-192P | rs59916403 | FARP2 |
| rs10445374 | NPEPPS | rs2332328 | NYNRIN | rs59950280 | HGFAC, DOK7 |
| rs1044573 | ENTPD6 | rs6016373 | - | rs599839 | PSRC1, CELSR2 |
| rs10448340 | PMPCA, INPP5E | rs233716 | RPH3A | rs60049679 | APOC1, APOC1P1 |
| rs10460181 | CEACAM16-AS1 | rs233721 | RPH3A | rs2335418 | HMGCR, ANKRD31 |
| rs1047743 | UNK | rs2347699 | BPGM, TUBB3P2 | rs6016381 | - |
| rs10500834 | SPTY2D1 | rs2358956 | TAF13 | rs6016505 | TOP1 |
| rs10504255 | CYP7A1, UBXN2B | rs2389606 | ABCB11 | rs6022850 | SUMO1P1, BCAS1 |
| rs10513551 | IFT80 | rs2395943 | PEX6 | rs6029526 | TOP1 |
| rs10761756 | JMJD1C | rs2435386 | NUTF2P8, TDRD15 | rs6029626 | ZHX3, ADI1P1 |
| rs10764055 | RPL7P37, PCAT5 | rs246179 | Y_RNA, MIR193BHG | rs603424 | PKD2L1 |
| rs10786068 | NIP7P1, CYP26A1 | rs247616 | CETP, HERPUD1 | rs604500 | ELAPOR1, SARS1 |
| rs10791660 | PDGFD | rs247617 | HERPUD1, CETP | rs6050463 | PYGB, ENTPD6 |
| rs10792318 | MYRF | rs2479394 | PCSK9, BSND | rs6062343 | TCEA2 |
| rs10794579 | C10orf88, FAM24A | rs2479409 | PCSK9, BSND | rs6065311 | TOP1 |
| rs10795464 | TRDMT1, VIM-AS1 | rs2495477 | PCSK9 | rs6065904 | PLTP |
| rs10808546 | LINC00861, TRIB1 | rs2495504 | BSND, PCSK9 | rs6073958 | PCIF1, PLTP |
| rs10810657 | BNC2 | rs249756 | SPRY4-AS1 | rs6088882 | FER1L4 |
| rs10832963 | SPTY2D1, SRSF3P1 | rs2500340 | BSND | rs6090040 | TCEA2 |
| rs10843391 | ERGIC2 | rs2510344 | NPC1 | rs6090101 | PCMTD2 |
| rs10846744 | SCARB1 | rs2522061 | IRF1, IRF1-AS1 | rs6093446 | PLCG1 |
| rs10885997 | PNLIPRP2 | rs2548957 | RNU6-317P, FGF21 | rs6102059 | LINC01370, MAFB |
| rs10888897 | PCSK9 | rs2569537 | LDLR | rs6129772 | ZHX3 |
| rs10888908 | MIR4422HG | rs2571077 | ZNF285, ZNF112 | rs61303167 | STYXL1 |
| rs10889348 | DOCK7 | rs2575876 | ABCA1 | rs6139104 | RBCK1 |
| rs10889353 | DOCK7 | rs2587534 | LINC01132, LINC00184 | rs61433703 | BANF2, RNU6-192P |
| rs10893499 | ST3GAL4 | rs261290 | ALDH1A2 | rs61602173 | HMGCR, ANKRD31 |
| rs10895547 | PDGFD | rs261334 | ALDH1A2, LIPC | rs61775180 | MACO1 |
| rs10903129 | MACO1 | rs261342 | ALDH1A2, LIPC-AS1, LIPC | rs61776818 | LDLRAP1, MACO1 |
| rs10910476 | U8, IRF2BP2 | rs2618566 | RNU6-192P, BANF2 | rs61778883 | ASAP3 |
| rs10950865 | DNAH11 | rs2618567 | BANF2, RNU6-192P | rs61946985 | N4BP2L1 |
| rs10953298 | TFR2, MOSPD3 | rs2618568 | RNU6-192P | rs62033400 | FTO |
| rs10957054 | UBXN2B | rs2642438 | MTARC1 | rs62116303 | ZNF229, ZNF285B |
| rs11024735 | SPTY2D1 | rs2642442 | MTARC1 | rs62116889 | CEACAM20 |
| rs11057353 | DNAH10 | rs2645492 | VMP1 | rs62120394 | PDE4C |
| rs11057397 | CCDC92, DNAH10 | rs2650000 | HNF1A-AS1 | rs62128801 | KRI1 |
| rs11057830 | SCARB1 | rs2678379 | APOB | rs62252794 | DYNC1LI1 |
| rs11065961 | ATXN2 | rs2706383 | IRF1, IRF1-AS1 | rs62275880 | DOK7 |
| rs11066320 | PTPN11 | rs2706770 | TIA1, C2orf42 | rs624249 | SLC22A2 |
| rs11080150 | NF1 | rs2710642 | EHBP1 | rs629301 | CELSR2 |
| rs11099097 | PRDM8, FGF5 | rs2710644 | EHBP1 | rs632057 | LINC01625, ATP5PBP6 |
| rs11127048 | GCKR, C2orf16 | rs2712199 | COG5 | rs636523 | DOCK7 |
| rs11136341 | PLEC | rs2716769 | - | rs6457976 | RPL12P2, COX6A1P2 |
| rs11136343 | PARP10 | rs2722733 | ZNF112 | rs6459450 | MDH1P2, MYLIP |
| rs11149612 | OSGIN1, MLYCD | rs2737245 | TRPS1 | rs6461564 | SP4 |
| rs111571253 | B3GNTL1 | rs2737252 | TRPS1 | rs646776 | CELSR2, PSRC1 |
| rs11164654 | EVI5, GFI1 | rs2737265 | TRPS1 | rs6475606 | CDKN2B-AS1 |
| rs11206788 | LINC01767 | rs2738445 | LDLR | rs648324 | PEX14 |
| rs11207995 | DOCK7 | rs2738447 | LDLR | rs6496691 | CRTC3 |
| rs11218721 | GLULP3, UBASH3B | rs2738459 | LDLR | rs6505081 | SLC46A1, H3P41 |
| rs11227247 | RELA | rs2738464 | LDLR, SPC24 | rs6511721 | LDLR |
| rs112403212 | SCARB1 | rs2740488 | ABCA1 | rs651821 | APOA5 |
| rs112521149 | CLCN6 | rs2741991 | THOP1 | rs653178 | ATXN2 |
| rs112758337 | BAIAP2L1 | rs2745865 | RNU6-192P, BANF2 | rs6542680 | RPS7, COLEC11 |
| rs112875651 | LINC00861, TRIB1 | rs2792703 | GPAM | rs6544713 | ABCG8 |
| rs112927854 | MTCO3P1, HLA-DQB1 | rs2792735 | GPAM | rs6557781 | DMTN |
| rs1129555 | GPAM | rs2803621 | GPAM | rs6560499 | PCSK5 |
| rs112987086 | LMAN1L | rs2807834 | MTARC1 | rs6573778 | NYNRIN |
| rs113270900 | ZIC4, RPL21P71 | rs2807835 | MTARC1 | rs6573971 | ADAM20, MED6 |
| rs1134027 | PARP10 | rs2807854 | HLX-AS1 | rs6579245 | C20orf173, ERGIC3 |
| rs1135062 | BCAM | rs2814982 | IFITM3P3, ILRUN | rs6587971 | KANK4, USP1 |
| rs11485618 | DOCK7 | rs2839619 | PKNOX1 | rs6587980 | DOCK7 |
| rs6589566 | ZPR1 | rs11563251 | UGT1A6, UGT1A1, UGT1A9, UGT1A7, UGT1A4, UGT1A3, UGT1A5, UGT1A8, UGT1A10 | rs28406917 | SP4, RNU1-15P |
| rs11571717 | BRCA2 | rs28456 | FADS2, FADS1 | rs6589939 | UBASH3B, GLULP3 |
| rs115739682 | PDGFD | rs28478252 | STAG1 | rs6595460 | CSNK1G3 |
| rs1160985 | TOMM40 | rs28503706 | RP1 | rs6601299 | RNU6-526P, RNU6-1151P |
| rs11620731 | SYNJ2BP-COX16, COX16 | rs2853579 | ABCA1 | rs660240 | CELSR2 |
| rs11621792 | NYNRIN | rs28537499 | MIR148A | rs6602909 | GAS6 |
| rs116403919 | RPL7AP64, ASGR1 | rs2854322 | NF1 | rs6602911 | GAS6-AS1, GAS6 |
| rs11665829 | NECTIN2 | rs2854332 | NF1, RAB11FIP4 | rs6602912 | GAS6-AS1, GAS6 |
| rs11668313 | KANK2 | rs28555129 | MLYCD, OSGIN1 | rs6603997 | EVI5 |
| rs11668477 | SMARCA4, LDLR | rs28601761 | LINC00861, TRIB1 | rs6667939 | LINC01221 |
| rs1167998 | DOCK7 | rs28631087 | RN7SL668P, LINC01348 | rs668948 | TDRD15, APOB |
| rs1168013 | DOCK7 | rs2866745 | CHD6 | rs6698843 | CELSR2 |
| rs1168127 | DOCK7 | rs28727898 | STAT5A | rs67050321 | EEPD1, MARK2P13 |
| rs11684202 | DTNB | rs287621 | KLF14, H4P1 | rs67143178 | SMIM12P1, SULT1C4 |
| rs1169288 | HNF1A-AS1, HNF1A | rs28768427 | PTK2 | rs6714750 | DARS1-AS1, CXCR4 |
| rs1169294 | HNF1A | rs2877660 | COX16, SYNJ2BP, SYNJ2BP-COX16 | rs67337506 | LDLR |
| rs1169314 | C12orf43 | rs28849176 | HLA-H, HCP5B | rs6734002 | EHBP1, RSL24D1P2 |
| rs11755689 | HLA-DRB1, HLA-DQA1 | rs2902940 | - | rs673548 | APOB |
| rs11781667 | PLEC | rs2911987 | MCPH1-AS1 | rs6756778 | ABCG8 |
| rs11783655 | PLEC | rs2912054 | AGPAT5 | rs676210 | APOB |
| rs11786083 | PLEC | rs2920503 | GSTM5P1, PPARG | rs67890964 | OSGIN1, MLYCD |
| rs11789603 | ABCA1 | rs2925677 | LINC02056 | rs6818397 | RGS12 |
| rs11802413 | MACO1 | rs2927439 | BCL3, CEACAM16-AS1 | rs682178 | CASZ1 |
| rs11807418 | Y_RNA, EFNA1 | rs2954021 | TRIB1, LINC00861 | rs6831256 | DOK7 |
| rs11812460 | CYP26C1, CYP26A1 | rs2954022 | TRIB1 | rs685001 | ATP6V0E1P4 |
| rs11846704 | CFL2, RPL12P6 | rs2954029 | TRIB1, LINC00861 | rs6857 | NECTIN2 |
| rs1186380 | RPL12P33, HNF1A-AS1 | rs2954031 | LINC00861, TRIB1 | rs6859 | NECTIN2 |
| rs11870935 | KPNB1 | rs2954038 | TRIB1, LINC00861 | rs6871667 | HMGCR, ANKRD31 |
| rs11881101 | RELB | rs2965164 | CEACAM16-AS1 | rs6871748 | IL7R, CAPSL |
| rs11902417 | APOB, LINC02850 | rs2972557 | NECTIN2 | rs6873053 | TIMD4 |
| rs11911615 | DOP1B | rs2980869 | LINC00861, TRIB1 | rs6874202 | HAVCR1, TIMD4 |
| rs11997161 | PTK2 | rs2980874 | TRIB1 | rs6882076 | HAVCR1, TIMD4 |
| rs12027135 | MACO1 | rs2980875 | TRIB1 | rs6882345 | HAVCR1, TIMD4 |
| rs12042319 | DOCK7 | rs2986164 | RSRP1, RHD | rs6882842 | HMGCR |
| rs12047226 | DOCK7 | rs2992753 | KLHDC7A | rs6894249 | IRF1-AS1, IRF1 |
| rs12078100 | ARHGEF19-AS1, EPHA2 | rs3010239 | LINC02056, RPL7P22 | rs6896579 | HAVCR2 |
| rs12117661 | BSND, PCSK9 | rs3010276 | RPL7P22, LINC02056 | rs6920309 | MYLIP |
| rs12124310 | DOCK7, ATG4C | rs306890 | SPRY3 | rs6920323 | MICA, HLA-S |
| rs12136600 | PCSK9 | rs3093679 | TNFAIP1 | rs6926458 | LPA |
| rs12162782 | PPP6R2 | rs3096644 | LINC02086 | rs693 | APOB |
| rs12171249 | PPP6R2 | rs3105749 | SLC22A3 | rs6932852 | FRK, NT5DC1 |
| rs12191504 | - | rs3108680 | Y_RNA, PRMT6 | rs693668 | PCSK9 |
| rs12197047 | L3MBTL3 | rs3110609 | LINC02086 | rs6938647 | LPA |
| rs12239737 | DOCK7 | rs3127573 | SLC22A2 | rs6944635 | ZNF619P1 |
| rs1229984 | ADH1B | rs312949 | TDRD15, APOB | rs6968865 | AHR |
| rs12318598 | KRAS, RNU4-67P | rs312976 | TDRD15 | rs6987702 | TRIB1 |
| rs1242229 | SIDT2 | rs314253 | DLG4 | rs7012637 | RNU6-1151P, RNU6-526P |
| rs12471768 | SERTAD2 | rs31674 | ABCB4 | rs7016880 | RPL30P9, LPL |
| rs12473839 | ACMSD, CCNT2-AS1 | rs3177928 | HLA-DRA | rs704 | VTN, SARM1 |
| rs1250229 | FN1 | rs3179865 | HLA-B | rs7090758 | REEP3 |
| rs1250258 | FN1 | rs3184504 | ATXN2, SH2B3 | rs7096937 | GPAM |
| rs12588332 | SYNJ2BP-COX16, COX16 | rs3213282 | XRCC1 | rs71352239 | APOC1, APOC1P1 |
| rs12595292 | PLEKHO2 | rs328996 | RALBP1 | rs7140110 | GAS6-AS1, GAS6 |
| rs12597024 | ZFPM1 | rs329007 | RALBP1 | rs7164309 | SECISBP2L |
| rs1260326 | GCKR | rs334558 | GSK3B | rs7185272 | PKD1L3 |
| rs12603885 | NF1 | rs34042070 | TXNL4B, HPR | rs719148 | PUM2 |
| rs12614487 | ACVR1C | rs34071855 | CASZ1 | rs7192750 | PKD1L3 |
| rs12620918 | EHBP1-AS1, EHBP1 | rs34318965 | LITAF | rs7202323 | LINC01572, PMFBP1 |
| rs12654264 | HMGCR | rs34361 | POC5, ANKDD1B | rs7203193 | LITAF |
| rs12657266 | TIMD4, HAVCR1 | rs34392107 | MTCO3P1, HLA-DQB1 | rs7205804 | CETP |
| rs12665537 | LINC01564, KLHL31 | rs34783010 | GIPR | rs7206971 | EFCAB13 |
| rs12670798 | DNAH11 | rs34791230 | MACO1 | rs7207466 | DPH1 |
| rs12701824 | C7orf50 | rs35136575 | APOC4, APOC1P1 | rs7222397 | LINC01483 |
| rs12705932 | PPP1R3A | rs35143646 | ARSL | rs7225700 | EFCAB13-DT |
| rs12709893 | SYMPK | rs351855 | FGFR4 | rs7241918 | SMUG1P1, LIPG |
| rs12712955 | PRKCE | rs35624969 | TBL2 | rs7258950 | SPC24 |
| rs12826964 | FBRSL1 | rs35891370 | RELB | rs7259004 | APOC1P1 |
| rs12891477 | LINC00637, ATP5MJ | rs36043200 | NPEPPS | rs7259278 | LDLR |
| rs12916 | HMGCR, CERT1 | rs360789 | EHBP1 | rs7259567 | CLASRP |
| rs12920772 | STX1B | rs360801 | EHBP1 | rs72626214 | CEACAM16-AS1 |
| rs12925859 | CASC22, LINC02180 | rs360804 | EHBP1 | rs72626215 | DMWD |
| rs12927205 | PKD1L3, RPL39P31 | rs364585 | LINC01723 | rs72694391 | NYNRIN |
| rs1293261 | RHCE | rs365653 | NECTIN2 | rs72749770 | C2CD4A, NPM1P47 |
| rs12936113 | ARSG | rs367881 | ADGRL2 | rs72784625 | LINC01814 |
| rs12945299 | ASGR1 | rs3729931 | RAF1, MKRN2 | rs72843670 | HLA-G, HCP5B |
| rs12948394 | PGS1 | rs3732359 | NR1I2 | rs72848251 | HLA-DQA1 |
| rs12960731 | TAF4B | rs3736206 | FOXK2 | rs7315593 | RPH3A |
| rs12976464 | GIPR, RN7SL836P | rs3745150 | NECTIN2 | rs7333748 | COL4A2 |
| rs12979813 | DOCK6 | rs3746778 | NTSR1 | rs73352129 | H3-3B |
| rs12986015 | CYP4F12 | rs3747910 | GPCPD1 | rs73461870 | CREB3L1 |
| rs12989783 | ABCB11 | rs3756772 | FRK | rs73553519 | MYMK, SLC2A6 |
| rs12990177 | THADA | rs3757354 | MDH1P2, MYLIP | rs73572039 | BCAM, NECTIN2 |
| rs13008369 | APOB, LINC02850 | rs3758413 | VIM-AS1 | rs737337 | DOCK6 |
| rs13108218 | HGFAC | rs3762637 | WDR5B-DT, KPNA1 | rs7386762 | DMTN |
| rs13146272 | CYP4V2 | rs3764261 | HERPUD1, CETP | rs738703 | FER1L4 |
| rs13205804 | HLA-DQB1, MTCO3P1 | rs3775228 | AFF1 | rs74410908 | APOB |
| rs13263353 | SLC7A13 | rs377584195 | PPIAP82, BAIAP2L1 | rs74435866 | MAFB, LINC01370 |
| rs13273592 | TRIB1, LINC00861 | rs3777411 | IGF2R | rs744487 | U8, LINC01354 |
| rs13277568 | TRPS1 | rs3780181 | VLDLR | rs74551598 | WHRN |
| rs13277801 | UBXN2B | rs3790136 | ERGIC3 | rs7481842 | FADS2, FADS3 |
| rs13283282 | PKN3 | rs3794695 | HPR, TXNL4B | rs7485656 | SCARB1 |
| rs13289095 | PKN3 | rs3798164 | SLC22A1 | rs74869459 | ZDHHC24, BBS1 |
| rs13301006 | ABCA1 | rs3798167 | SLC22A1 | rs7515577 | EVI5 |
| rs1331698 | HMGB1, UBE2L5 | rs3811055 | IL1F10 | rs7519530 | MACO1 |
| rs13379043 | MIDEAS | rs3820897 | COLEC11, RPS7 | rs7523141 | BSND, PCSK9 |
| rs13389219 | COBLL1 | rs3822855 | FRK | rs7523242 | BSND, PCSK9 |
| rs13392272 | APOB | rs3824667 | GATA3 | rs7525649 | BSND, PCSK9 |
| rs1350559 | TNKS | rs3826804 | DNM2 | rs753381 | PLCG1 |
| rs1365041 | MCPH1-AS1 | rs3843482 | HMGCR | rs7538216 | EPHA2, ARHGEF19-AS1 |
| rs1367117 | APOB | rs3846661 | HMGCR | rs754340 | PVR, CEACAM16-AS1 |
| rs13702 | LPL | rs3846662 | HMGCR | rs754523 | TDRD15, APOB |
| rs1377870 | TMPRSS11E | rs3846663 | CERT1, HMGCR | rs7562734 | EHBP1 |
| rs138352 | XPNPEP3 | rs3850634 | DOCK7 | rs7567229 | UGT1A8, UGT1A7, UGT1A6, UGT1A9, UGT1A10 |
| rs138730 | TOM1 | rs3852789 | LINC01572, PMFBP1 | rs7569317 | FAM117B |
| rs1389742 | MTARC1 | rs3852856 | NECTIN2 | rs7570971 | RAB3GAP1 |
| rs140145058 | HLA-DQA1 | rs3865453 | CYP2F2P, CYP2A6 | rs7575840 | TDRD15, APOB |
| rs14234 | FAM136A | rs390082 | APOC1, APOE | rs75816352 | COL4A2 |
| rs1423527 | HMGCR, ANKRD31 | rs3902354 | CELSR2, PSRC1 | rs7601153 | ACVR1C |
| rs143375141 | APOB, LINC02850 | rs3932048 | STAG1 | rs7602171 | ABCB11 |
| rs145184406 | PDGFD | rs3936511 | C5orf67 | rs7603427 | RAPH1 |
| rs1461729 | RNU6-526P, RNU6-1151P | rs395908 | NECTIN2 | rs7610507 | SLC35G2 |
| rs1476442 | MTERF1 | rs3999089 | JMJD1C | rs76162994 | TPM1 |
| rs147841808 | HLA-DQA1 | rs4021 | FUT1 | rs7623486 | MITF |
| rs1489502 | - | rs406456 | NECTIN2 | rs7645585 | RPL32P8, TM4SF1-AS1 |
| rs1495741 | NAT2, PSD3 | rs4082919 | PGS1 | rs76530229 | ZNF14, ZNF101 |
| rs1495743 | PSD3, NAT2 | rs409331 | NUTF2P8, TDRD15 | rs76828422 | STAT5B |
| rs1500188 | CSNK1G3, HMGB3P17 | rs4129767 | PGS1 | rs769450 | APOE |
| rs1501908 | HAVCR1, TIMD4 | rs4148218 | ABCG8 | rs7703051 | ANKRD31, HMGCR |
| rs4149291 | ABCA1 | rs4148826 | ABCB4 | rs7703282 | POLK, ANKDD1B |
| rs1532085 | ALDH1A2 | rs1531517 | CEACAM16-AS1, BCL3 | rs7704651 | CSNK1G3, HMGB3P17 |
| rs1535 | FADS2 | rs416041 | NECTIN2 | rs7709540 | CSNK1G3 |
| rs1551891 | CEACAM16-AS1, BCL3 | rs41785 | CAPZA2 | rs77257036 | TARS2 |
| rs1553318 | HAVCR1 | rs4240624 | RNU6-1151P, RNU6-526P | rs77303550 | TXNL4B |
| rs1556562 | EVI5 | rs4244029 | SPRY4-AS1 | rs7734476 | CSNK1G3 |
| rs1558861 | BUD13, LINC02702 | rs4245786 | ABCG5 | rs7741091 | MICA, HLA-S |
| rs1571790 | PCSK5 | rs4245791 | ABCG8 | rs7746081 | MYLIP, MDH1P2 |
| rs157580 | TOMM40 | rs429358 | APOE | rs7756550 | MAP3K4-AS1, PLG |
| rs157588 | TOMM40 | rs4297946 | PLCG1, ZHX3 | rs7776054 | HBS1L |
| rs157595 | APOC1P1, APOC1 | rs4299376 | ABCG8 | rs7780562 | MIR148A, NFE2L3 |
| rs1584063 | GSTM5P1, SYN2 | rs4307732 | ST3GAL4 | rs780093 | GCKR |
| rs1610095 | DNM2 | rs4360309 | TRIB1 | rs780094 | GCKR |
| rs1640273 | THOP1 | rs4374942 | RN7SKP280, INSIG1-DT | rs7821092 | RP1L1 |
| rs165722 | COMT | rs438568 | LINC01723 | rs7832643 | PLEC |
| rs1663564 | WASHC4 | rs438811 | APOC1, APOE | rs7873387 | ABCA1 |
| rs16971384 | ZFHX3 | rs4418728 | NIP7P1, CYP26A1 | rs7902274 | C10orf88B |
| rs16973585 | PKD1L3 | rs4420638 | APOC1, APOC1P1 | rs7902343 | JMJD1C |
| rs16975985 | RPL32P31 | rs4452060 | BCAM, NECTIN2 | rs7904973 | C10orf88 |
| rs16979595 | CLPTM1 | rs445925 | APOC1, APOE | rs7908745 | MARCHF8 |
| rs16988410 | LIF-AS1, HORMAD2 | rs4495740 | DOCK7 | rs7928577 | ST3GAL4 |
| rs16996148 | CILP2, PBX4 | rs4530754 | CSNK1G3 | rs7941030 | UBASH3B, GLULP3 |
| rs17001095 | SMARCA4 | rs45537841 | ABCB4, ABCB1 | rs7955221 | NR1H4, SLC17A8 |
| rs17091905 | LPL, RPL30P9 | rs4564803 | APOB, LINC02850 | rs7968419 | Y_RNA, SSH1 |
| rs17111479 | BSND | rs4587594 | DOCK7 | rs799157 | MLXIPL |
| rs17111503 | PCSK9, BSND | rs4588831 | - | rs79953563 | THEM7P |
| rs17118739 | CYSTM1 | rs4604177 | POLK | rs8005362 | ADAM21P1, COX16 |
| rs1713222 | TDRD15, APOB | rs4616688 | IFT80 | rs8008068 | SYNE2 |
| rs17135399 | GSEC, DCPS | rs4620259 | - | rs8022288 | SYNJ2BP, COX16, SYNJ2BP-COX16 |
| rs17205170 | HLA-DQA1 | rs4651135 | LAMC1, RNU6-41P | rs8051431 | PKD1L3 |
| rs17231506 | CETP, HERPUD1 | rs4665710 | LINC02850, APOB | rs8062895 | DHODH |
| rs1730859 | NTNG1, PRMT6 | rs4665972 | SNX17 | rs8078686 | KPNB1 |
| rs1731243 | KCNK3 | rs4671050 | EHBP1 | rs8105074 | SLC44A2, AP1M2 |
| rs17321515 | LINC00861, TRIB1 | rs4689653 | SORCS2 | rs8106922 | TOMM40 |
| rs17336182 | PLEKHH2 | rs4703676 | POC5 | rs8107974 | SUGP1 |
| rs173539 | CETP, HERPUD1 | rs4704834 | TIMD4, HAVCR1 | rs8112975 | PDE4C |
| rs17405319 | TRIB1 | rs4714638 | RPL24P4, CNPY3 | rs8126001 | OPRL1 |
| rs174529 | TMEM258, MYRF | rs4719841 | MIR148A | rs8176720 | ABO |
| rs174548 | FADS1, FADS2 | rs472495 | PCSK9 | rs8191754 | IGF2R |
| rs174564 | FADS2, FADS1 | rs4738679 | CYP7A1, UBXN2B | rs821840 | HERPUD1, CETP |
| rs174566 | FADS1, FADS2 | rs4738684 | UBXN2B, CYP7A1 | rs836550 | RAC1 |
| rs174577 | FADS2 | rs4745 | EFNA1 | rs855791 | TMPRSS6 |
| rs174583 | FADS2 | rs4751996 | PNLIPRP2 | rs871841 | ARHGEF15 |
| rs174601 | FADS2 | rs4756996 | CSRP3-AS1, NAV2 | rs880315 | CASZ1 |
| rs1748195 | DOCK7 | rs4766578 | ATXN2 | rs881376 | LINC00612, VDAC2P2 |
| rs17489373 | LPL, RPL30P9 | rs4767631 | KSR2 | rs892161 | HDGFL2 |
| rs17561950 | PGS1 | rs4773173 | COL4A2 | rs907866 | RN7SL140P, RPS16P2 |
| rs1757216 | CACNB2 | rs4782568 | MLYCD, OSGIN1 | rs913499 | COX6A1P2, RPL12P2 |
| rs17663874 | LDAH | rs4788609 | PMFBP1 | rs9266229 | HLA-B, DHFRP2 |
| rs17725246 | NPC1L1, DDX56 | rs4791641 | PFAS | rs9275052 | HLA-DQB1, MTCO3P1 |
| rs1797912 | PPARG | rs4794048 | TBKBP1, KPNB1 | rs9293641 | GCNT4 |
| rs1799955 | BRCA2 | rs4803750 | BCL3, CEACAM16-AS1 | rs9297994 | CYP7A1, UBXN2B |
| rs1807675 | C22orf34, RN7SKP252 | rs4804146 | SPC24 | rs9298506 | RP1 |
| rs1808192 | TBX21, TBKBP1 | rs4804149 | KANK2 | rs9327296 | HMGB3P17, CSNK1G3 |
| rs1809165 | LINC00861, TRIB1 | rs4804155 | DOCK6 | rs9330459 | SLC2A6, MYMK |
| rs182472492 | FAM117B, MTCO1P54 | rs4804564 | SMARCA4 | rs934197 | APOB, TDRD15 |
| rs183130 | HERPUD1, CETP | rs4808360 | CYP4F12 | rs934198 | TDRD15, APOB |
| rs1856066 | LINC00402, KLF12 | rs4808762 | PDE4C | rs9371220 | CCDC170 |
| rs1861398 | SERTAD2 | rs4808765 | PDE4C | rs9378220 | TRIM31-AS1, TRIM31 |
| rs1862719 | - | rs4818025 | HMGN1, BRWD1-AS1 | rs9389268 | HBS1L |
| rs1883025 | ABCA1 | rs483082 | APOE, APOC1 | rs9391803 | HCG17 |
| rs188437955 | GDI1, FAM50A | rs1891110 | FAM24B | rs941408 | THOP1 |
| rs484084 | LINC00184, LINC01132 | rs4841132 | RNU6-526P, RNU6-1151P | rs9396646 | MRPL42P2, RNU6-1114P |
| rs1895233 | CSNK1G3 | rs4844614 | CR1L | rs9438901 | RSRP1 |
| rs1919903 | TMPRSS11E | rs4846922 | GALNT2 | rs9438905 | MACO1 |
| rs1936800 | RSPO3, RPS4XP9 | rs4850047 | RPS7, COLEC11 | rs9438909 | MACO1 |
| rs1963676 | BRWD1-AS1, HMGN1 | rs486142 | LINC00184, LINC01132 | rs9471975 | RPL24P4, CNPY3 |
| rs2000999 | TXNL4B, HPR | rs4870470 | H3P28 | rs9488822 | FRK |
| rs2001846 | LINC00861, TRIB1 | rs4870941 | LINC00861, TRIB1 | rs9491699 | RSPO3 |
| rs2001945 | TRIB1 | rs4870949 | TRIB1, LINC00861 | rs9577874 | TMEM255B, GAS6-AS1 |
| rs2002574 | SH3TC1 | rs4876611 | TRPS1 | rs9577924 | GAS6, GAS6-AS1 |
| rs201796 | DLEU1 | rs4897364 | L3MBTL3 | rs9616822 | PPP6R2 |
| rs2027061 | EVI5 | rs4925325 | CDH4 | rs9616847 | PPP6R2 |
| rs2030746 | Y_RNA, LINC01101 | rs4931005 | RESF1 | rs964184 | ZPR1 |
| rs203273 | RBM47, RNU7-74P | rs4937122 | ST3GAL4 | rs9646133 | RN7SL77P, TTC9 |
| rs204469 | CLPTM1 | rs4938362 | PCSK7 | rs9673065 | SCAMP2, MPI |
| rs2047596 | ZFPM1 | rs4942486 | BRCA2 | rs9686661 | C5orf67 |
| rs2050058 | ZHX3 | rs4946713 | PRDM1, RN7SKP211 | rs9687846 | C5orf67 |
| rs206319 | N4BP2L1 | rs2066714 | ABCA1 | rs976058 | TMPRSS11E |
| rs4954192 | CCNT2-AS1, ACMSD | rs4954559 | CXCR4, DARS1-AS1 | rs9832727 | PCOLCE2, PAQR9 |
| rs2068888 | CYP26A1, NIP7P1 | rs4959063 | HLA-B, DHFRP2 | rs983309 | RNU6-526P, RNU6-1151P |
| rs2072183 | NPC1L1 | rs4962050 | ADAMTS13 | rs9841897 | PARP9 |
| rs2073547 | DDX56, NPC1L1 | rs496800 | LINC00184, LINC01132 | rs984976 | ANKDD1B |
| rs2075290 | ZPR1 | rs4969183 | PGS1 | rs9894946 | TP53 |
| rs2075650 | TOMM40 | rs9918079 | CC2D2A | rs9897063 | NPEPPS, MRPL45P2 |
| rs2076674 | SLC25A17 | rs505151 | PCSK9 | rs990619 | GUCY1A1 |
| rs2080401 | LINC01124, MYO3B | rs2126259 | RNU6-526P, RNU6-1151P | rs503662 | TDRD15, NUTF2P8 |
| rs2081687 | UBXN2B, CYP7A1 | rs514230 | LINC00184, LINC01132 | rs9929977 | MARVELD3, TAT-AS1 |
| rs211642 | ARSL | rs515135 | APOB, TDRD15 | rs9939224 | CETP |
| rs2125345 | UNK | rs519380 | RBM14-RBM4, RBM4 | rs995000 | DOCK7 |
| rs506234 | TREH | rs525028 | APOC3, APOA1 | rs9956878 | - |
| rs2131925 | DOCK7 | rs551473284 | COL4A2 | rs99780 | FADS2, FADS1 |
| rs9988450 | DOCK7 | rs541041 | TDRD15, APOB | rs9982111 | BRWD1 |
| rs2147324 | CIART | rs550619 | APOB | rs998584 | LINC02537, VEGFA |
| rs2159324 | MARK4, BLOC1S3 | rs526936 | LINC00184, LINC01132 | rs9987289 | RNU6-1151P, RNU6-526P |
| rs2160994 | RNU6-1093P, LIMA1 | rs553427 | LINC01132, LINC00184 | rs2142672 | MRPL42P2, RNU6-1114P |
| rs2168608 | IL34 | rs556107 | LINC01132, LINC00184 | rs9989419 | HERPUD1, CETP |
| rs2169387 | RNU6-526P, RNU6-1151P | rs557933 | LINC01132, LINC00184 | rs998974 | OTX2P1, RNU6-1228P |
| rs2179050 | HMGXB4 |  |  | rs55747707 | MLXIPL |
