## Supplementary file 5 for "Ethnic and region-specific genetic risk variants of stroke and its comorbid conditions can define the variations in the burden of stroke and its phenotypic traits"

**Table S7. Continent Regions.**

The individual countries included in each ethnogeographic region.

| **Continent** | **Countries included in Continent** |
| --- | --- |
| **Central & South Asia** | Afghanistan, Bangladesh, Bhutan, India, Kazakhstan, Kyrgyzstan, Maldives, Nepal, Pakistan, Sri Lanka, Tajikistan, Turkmenistan, Uzbekistan |
| **East Asia** | Brunei Darussalam, Cambodia, China, Democratic People's Republic of Korea, Indonesia, Japan, Lao People's Democratic Republic, Malaysia, Mongolia, Myanmar, Philippines, Republic of Korea, Singapore, Taiwan (Province of China), Thailand, Timor-Leste, Viet Nam |
| **Middle East** | Bahrain, Iran (Islamic Republic of), Iraq, Israel, Jordan, Kuwait, Lebanon, Oman, Palestine, Qatar, Saudi Arabia, Syrian Arab Republic, United Arab Emirates, Yemen, Armenia, Azerbaijan, Georgia, Turkey |
| **Africa** | Algeria, Angola, Benin, Botswana, Burkina Faso, Burundi, Cabo Verde, Cameroon, Central African Republic, Chad, Comoros, Congo, Côte d'Ivoire, Democratic Republic of the Congo, Djibouti, Egypt, Equatorial Guinea, Eritrea, Eswatini, Ethiopia, Gabon, Gambia, Ghana, Guinea, Guinea-Bissau, Kenya, Lesotho, Liberia, Libya, Madagascar, Malawi, Mali, Mauritania, Mauritius, Morocco, Mozambique, Namibia, Niger, Nigeria, Rwanda, Sao Tome and Principe, Senegal, Seychelles, Sierra Leone, Somalia, South Africa, Sudan, Togo, Tunisia, Uganda, United Republic of Tanzania, Zambia, Zimbabwe |
| **Europe** | Albania, Andorra, Austria, Belarus, Belgium, Bosnia and Herzegovina, Bulgaria, Croatia, Cyprus, Czechia, Denmark, Estonia, Finland, France, Germany, Greece, Greenland, Hungary, Iceland, Ireland, Italy, Latvia, Lithuania, Luxembourg, Malta, Montenegro, Netherlands, North Macedonia, Norway, Poland, Portugal, Republic of Moldova, Romania, Russian Federation, Serbia, Slovakia, Slovenia, Spain, Sweden, Switzerland, Ukraine, United Kingdom |
| **America** | Antigua and Barbuda, Argentina, Bahamas, Barbados, Belize, Bermuda, Bolivia (Plurinational State of), Brazil, Canada, Chile, Colombia, Costa Rica, Cuba, Dominica, Dominican Republic, Ecuador, El Salvador, Grenada, Guatemala, Guyana, Haiti, Honduras, Jamaica, Mexico, Nicaragua, Panama, Paraguay, Peru, Puerto Rico, Saint Kitts and Nevis, Saint Lucia, Saint Vincent and the Grenadines, Suriname, Trinidad and Tobago, United States of America, Uruguay, Venezuela (Bolivarian Republic of) |
| **Oceania** | American Samoa, Australia, Cook Islands, Fiji, Kiribati, Marshall Islands, Micronesia (Federated States of), Nauru, New Zealand, Niue, Palau, Papua New Guinea, Samoa, Solomon Islands, Tokelau, Tonga, Tuvalu, Vanuatu |
