## Supplementary file 6 for "Ethnic and region-specific genetic risk variants of stroke and its comorbid conditions can define the variations in the burden of stroke and its phenotypic traits"

**Table S8. Number of final risk variants for each disease.** The final number of GWAS risk variants for each disease (i) after the removal of loci which are less than 10000 bp apart and (ii) which have alternate allele frequency above or equal to 0.05.

| **Disease** | **# of risk variants (biallelic SNPS) at least 10000 bp apart** | **# of risk variants (biallelic SNPS)**  **With alternate allele frequency >= 0.05** |
| --- | --- | --- |
| Stroke | 326 | 291 |
| Ischemic heart disease | 866 | 1080 |
| Hypertension | 378 | 392 |
| Type 1 diabetes | 227 | 256 |
| Type 2 diabetes | 1728 | 2072 |
| Chronic Kidney Disease | 117 | 96 |
| High BMI | 24 | 26 |
| High LDL | 1107 | 1458 |
