## Supplementary file 7 for "Ethnic and region-specific genetic risk variants of stroke and its comorbid conditions can define the variations in the burden of stroke and its phenotypic traits"

SUPPLEMENTARY FIGURES

**Table of Contents**

| **Title** | **Page** |
| --- | --- |
| Figure 1-figure supplement 1. Global incidence, mortality and prevalence rates for Stroke, its subtypes and comorbid factors | 2 |
| Figure 1-figure supplement 2. Boxplots of point estimates and uncertainty intervals of age-standardised mortality rates of (A) stroke, its subtypes and (B) comorbid conditions across regions from 2009-2019 | 3-4 |
| Figure 1-figure supplement 3. Boxplots of point estimates and uncertainty intervals of age-standardised prevalence rates of (A) stroke, its subtypes and (B) comorbid conditions across regions from 2009-2019 | 5-6 |
| Figure 4-figure supplement 1. Population structure of risk variants for Strokes, High SBP and Metabolic disorders using a model-based clustering | 7 |
| Figure 8-figure supplement 1. Linkage disequilibrium of common variants proxy to risk variants of stroke unique in South Asian population. | 8 |
| Appendix Figure 1. Population across different continents in 2009, 2014 and 2019 | 9 |

**Figure 1-figure supplement 1. Global incidence, mortality and prevalence rates for S**
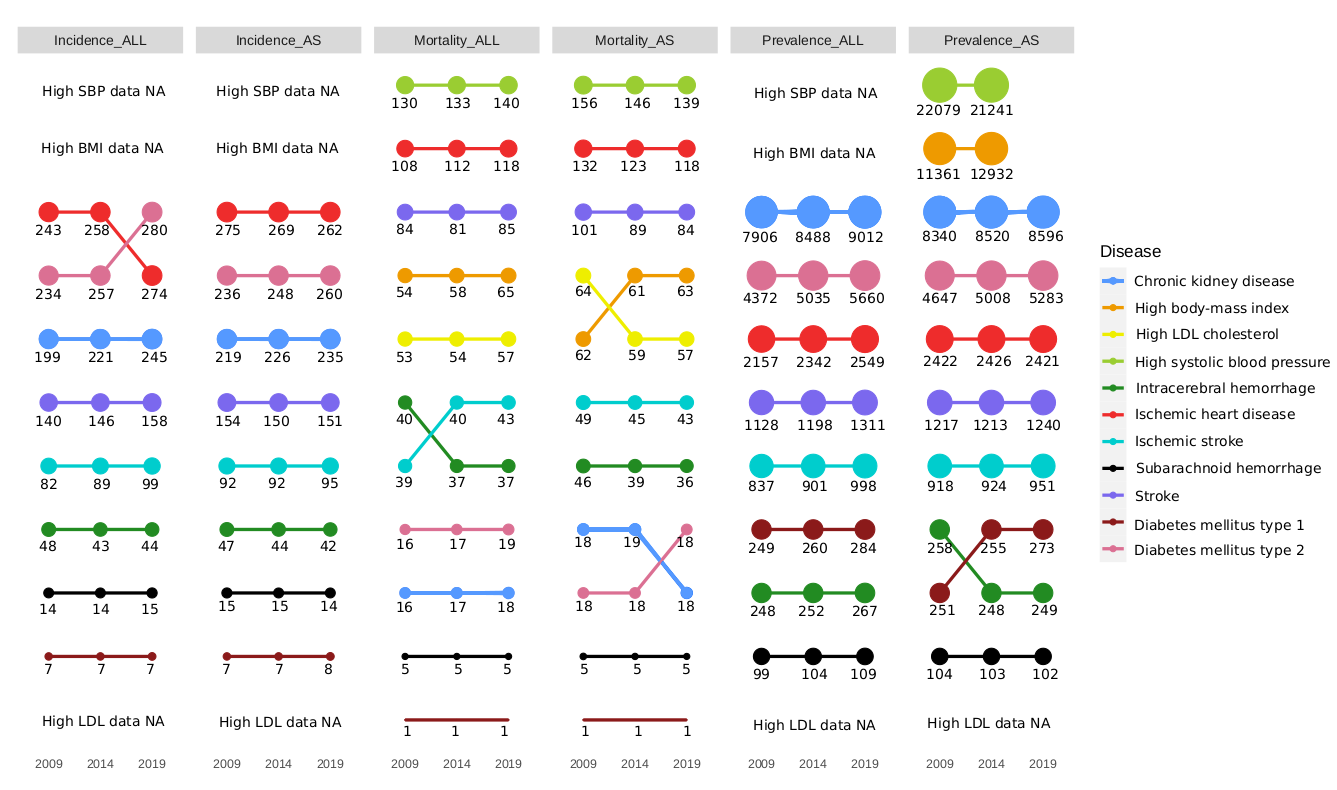
**troke, its subtypes and comorbid factors.** The figure shows the global crude and age-standardized incidence, mortality and prevalence rates per 100,000 people for Stroke, its subtypes and its comorbid factors in 2009, 2014 and 2019. The size of the points indicates the rate and position indicates rank. *_ALL indicates crude rates and *_AS indicates age-standardized rates

**Figure 1-figure supplement 2. Boxplots of point estimates and uncertainty intervals of age-standardised mortality rates of (A) stroke, its subtypes and (B) its comorbid conditions across regions from 2009-2019.** X-axis shows the different regions, y-axis shows the range of ASMRs per 100,000 for each disease.

(A)

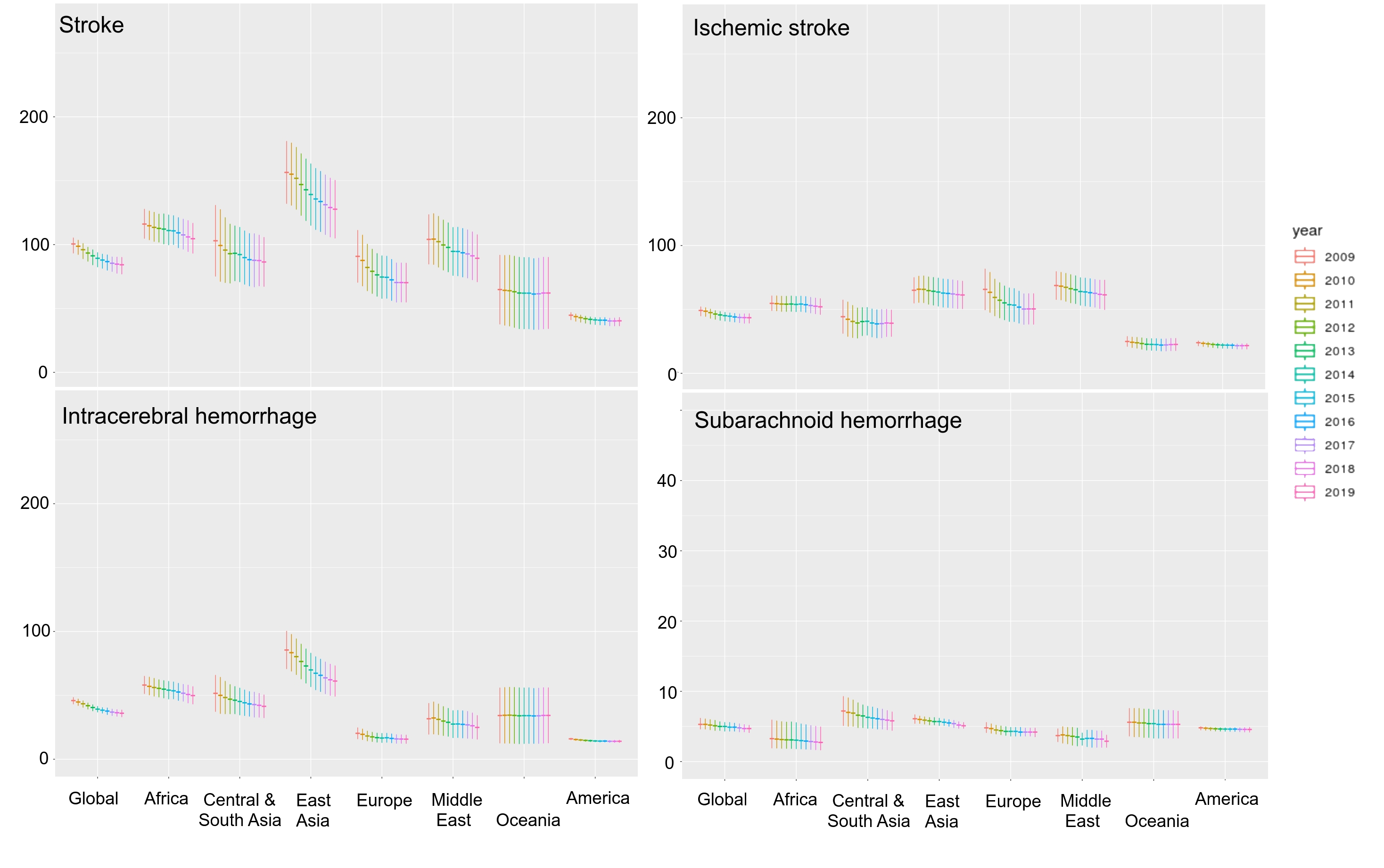

**(B)**
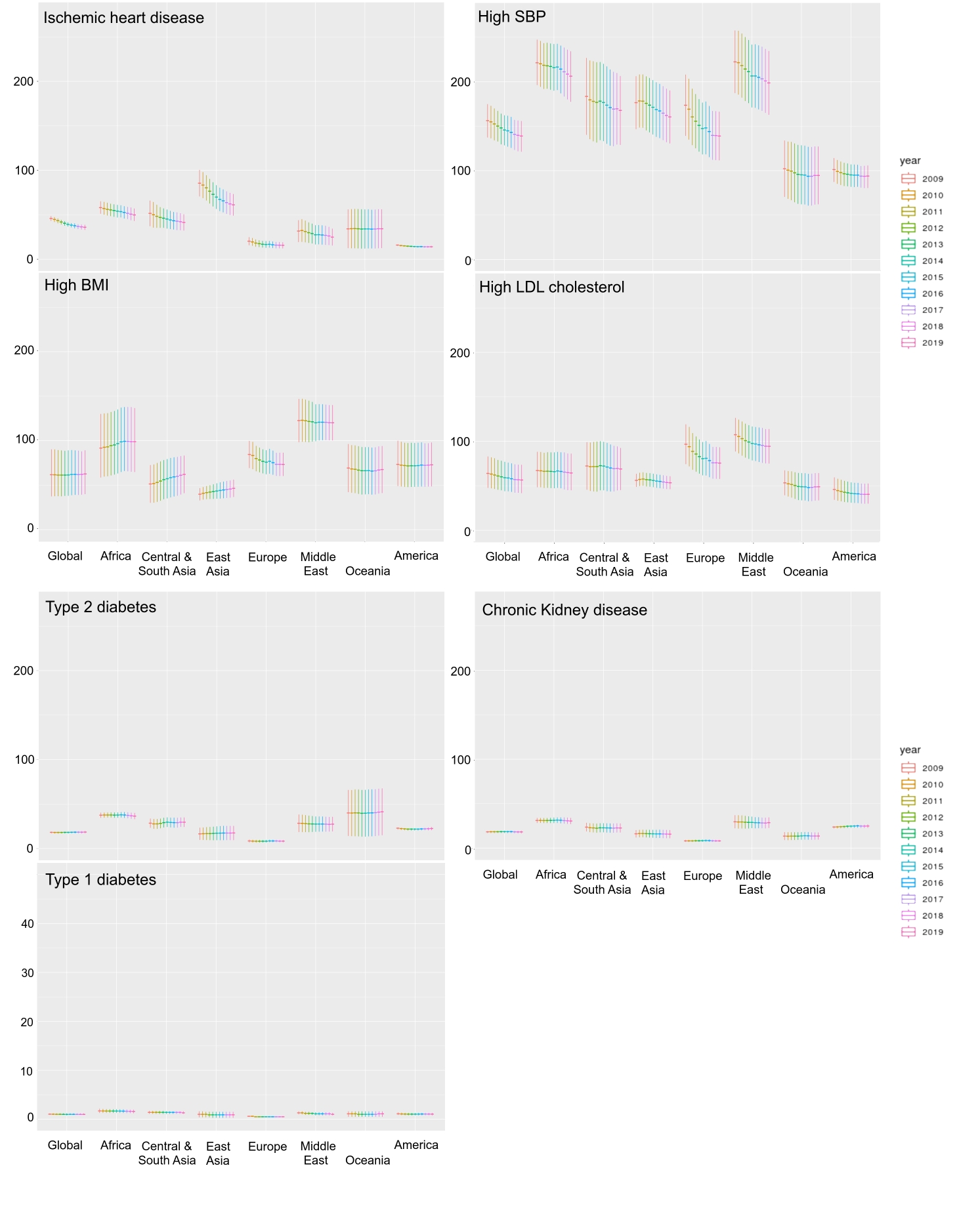

**Figure 1-figure supplement 3. Boxplots of point estimates and** **uncertainty intervals of age-standardised prevalence rates of (A) stroke, its subtypes and (B) its comorbid conditions across regions from 2009-2019.** X-axis shows the different regions, y-axis shows the range of ASPRs per 100,000 for each disease.

(A)

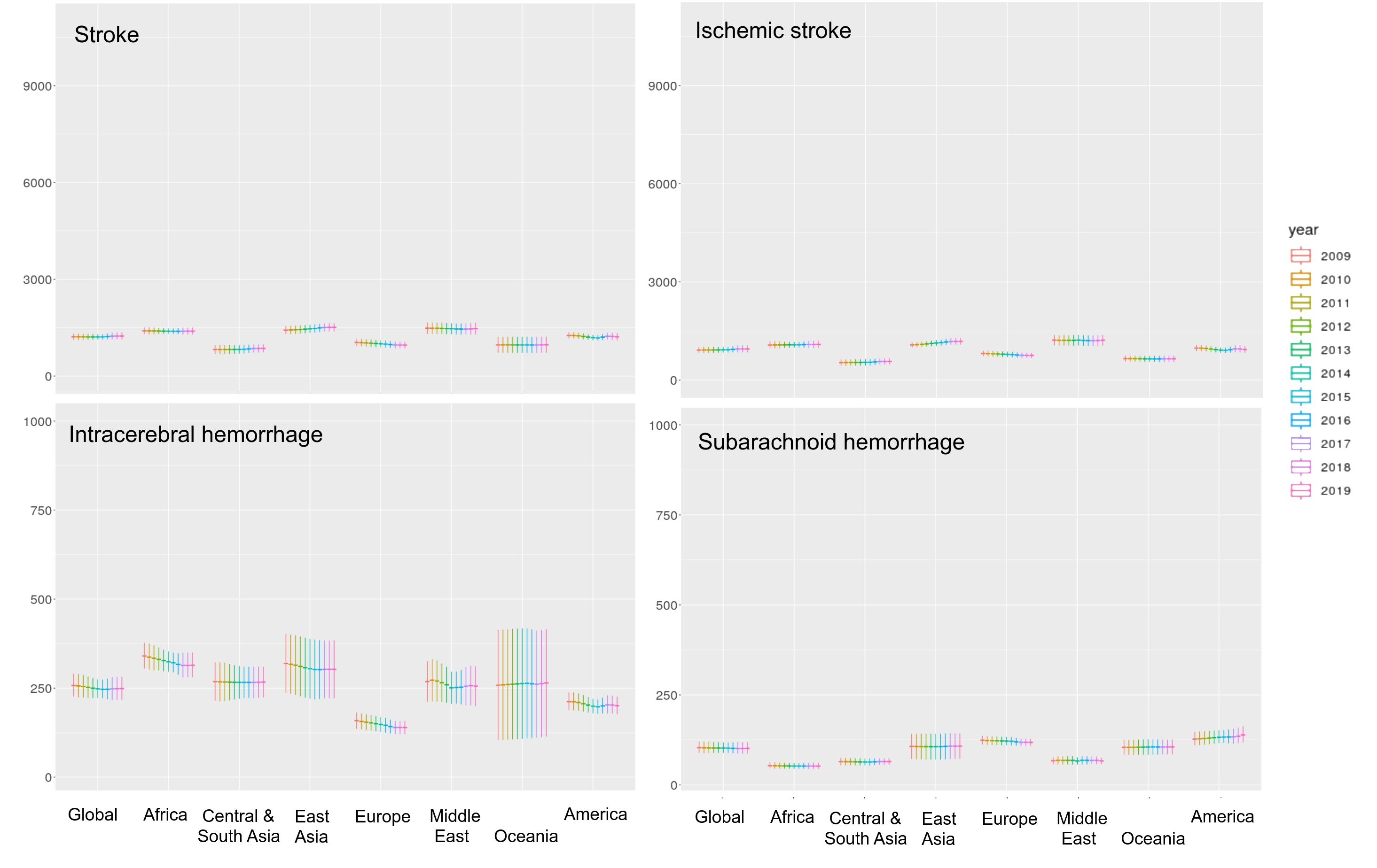

(B)

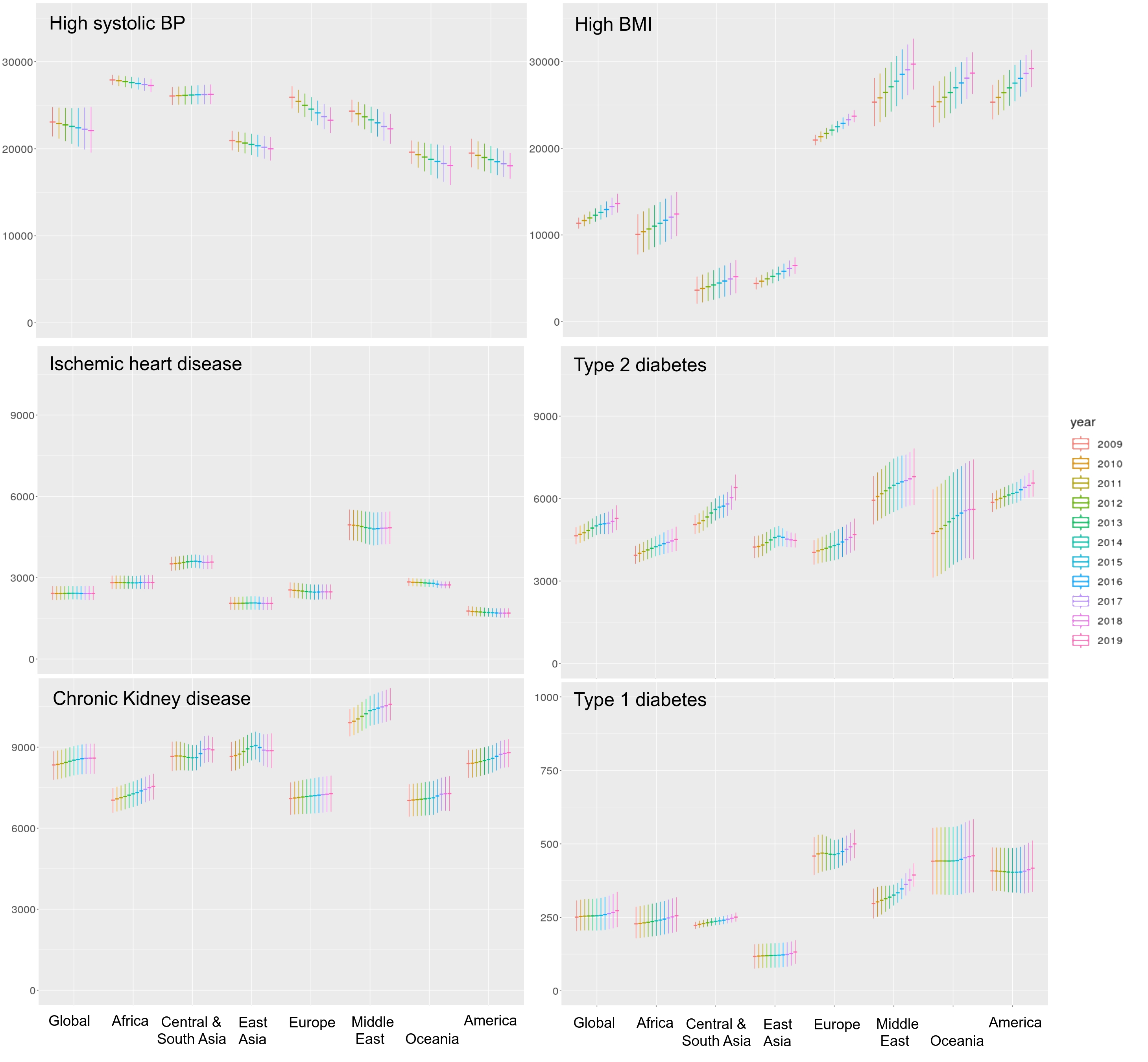

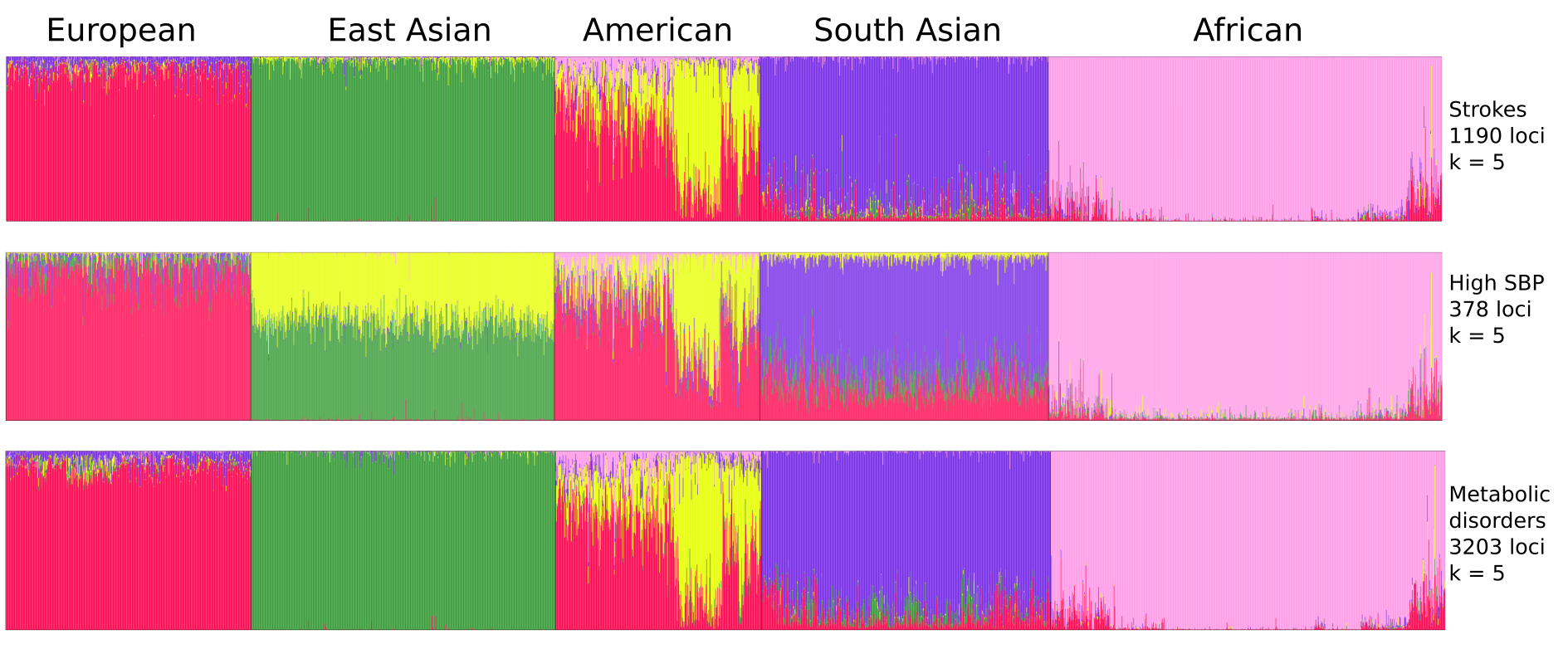
**Figure 4-figure supplement 1. Population structure of risk variants for Strokes, High SBP and Metabolic disorders using a model-based clustering.** The proportion of ancestral populations was estimated from the genotype of risk variants of each disease in unrelated individuals from the 1000 Genomes project using a model-based clustering approach. The individuals were represented by their super-population in 1000 Genomes (African, East Asian, South Asian, European and American). The estimated ancestral populations clustered into either 5 clusters represented by the different colors.

**Figure 8-figure supplement 1.** **Linkage disequilibrium of common variants proxy to risk variants of stroke unique in South Asian population.** LD plots of common proxy variants in different 1000 Genome super-populations (AFR- Africa, EAS- East Asia, SAS – South Asia, AMR – America, EUR – Europe). Proxy variants with frequency greater than 0.1 in 1MB region flanking the stroke risk variants unique in South Asia (a) rs528002287 and (b) rs148010464. The common variants in (a) also includes a risk variant present in all five super-populations for the cormorbid condition of T2D, bounded in a blue box.

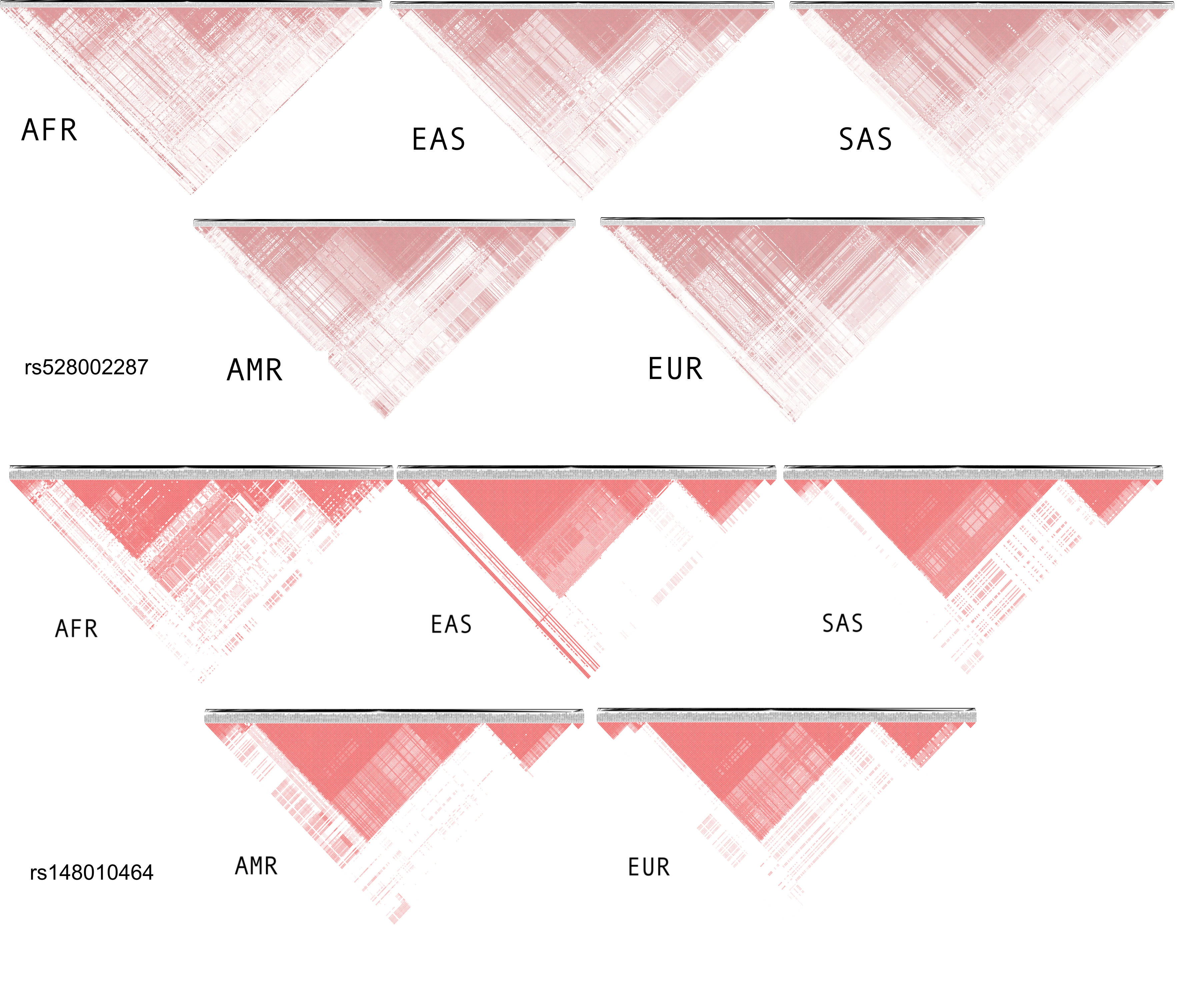

**Appendix Figure 1.**
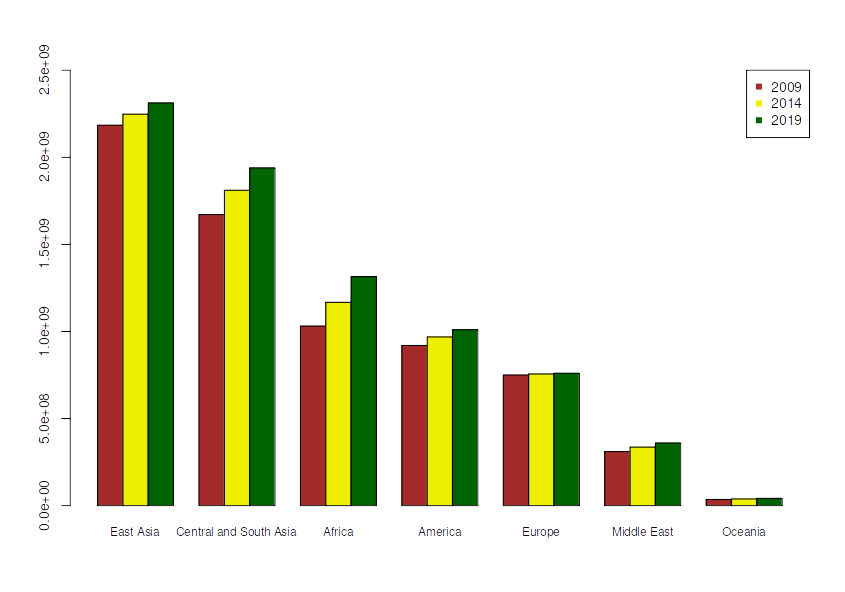
Population across different continents in 2009, 2014 and 2019.
